# Impact of Comorbidity and Drug Exposure Patterns on Recurrence and Mortality Risks in Patients with Primary Hepatocellular Carcinoma: A Chinese Cohort Network Analysis Study

**DOI:** 10.1101/2025.07.05.25330803

**Authors:** Bingqing Yang, Jiang Liu, Oscar Hou-In Chou, Qingze Gu, Bernard Man Yung Cheung, Gary Tse, Wing Tak Jack Wong, Tingting Zhu, Kevin Tak-Pan Ng, William Chi Wai Wong, Nancy Kwan Man, David Ka Ki Wong, Jiandong Zhou

**Affiliations:** Department of Family Medicine and Primary Care, Li Ka Shing Faculty of Medicine, The University of Hong Kong, Hong Kong SAR, China; Department of Surgery, School of Clinical Medicine, Li Ka Shing Faculty of Medicine, The University of Hong Kong, Hong Kong SAR, China; Division of Clinical Pharmacology and Therapeutics, Department of Medicine, Li Ka Shing Faculty of Medicine, The University of Hong Kong, Hong Kong SAR, China; Nuffield Department of Orthopaedics, Rheumatology and Musculoskeletal Sciences, University of Oxford, Oxford, United Kingdom; Tianjin Key Laboratory of Ionic-Molecular Function of Cardiovascular Disease, Department of Cardiology, Tianjin Institute of Cardiology, Second Hospital of Tianjin Medical University, Tianjin 300211, China; Department of Health Sciences, School of Nursing and Health Studies, Hong Kong Metropolitan University, Hong Kong, China; School of Life Sciences, Faculty of Science, The Chinese University of Hong Kong, Hong Kong, China; Institute of Biomedical Engineering, Department of Engineering Science, University of Oxford, Oxford, United Kingdom; School of Public Health, Li Ka Shing Faculty of Medicine, The University of Hong Kong, Hong Kong SAR, China; Department of Pharmacology and Pharmacy, The University of Hong Kong, Hong Kong SAR, China

**Keywords:** Hepatocellular carcinoma, population cohort study, comorbidity, multi-drug exposure, early recurrence

## Abstract

**Background & Aims:** Comorbidities in hepatocellular carcinoma (HCC) impair liver function, limit treatment choices, disrupt drug metabolism, and adversely impact survival. This study identifies comorbidity and multidrug combination patterns to evaluate their relationships with HCC outcomes.

**Methods:** This cohort study comprised 15,998 patients (62.7% male, age at HCC diagnosis: 69.2 [IQR: 58.7-78.7] years old) from the Hong Kong Hospital Authority, enrolled between January 2008 and December 2019 with follow-up until May 2024. Primary endpoints were HCC recurrence and liver cancer-related mortality, while secondary endpoints included cancer-related mortality, non-cancer-related mortality, and all-cause mortality. We identified the most frequent double, triple, quadruple, quintuple patterns of both comorbidities and multidrug combinations using UpSet plots, then examined their associations with clinical outcomes using network analyses. Risk factors were evaluated through Cox proportional hazards regression models, with robustness confirmed via sensitivity analyses and subgroup assessments.

**Results:** In this cohort, 93.0% of patients had at least one pre-existing comorbidity (mean 3.0 per patient), of whom 87.8% (N=13,084) had 1-5 comorbidities. Diabetes mellitus, hypertension, and existing chronic liver diseases were the predominant comorbidities across all combinations, followed by colorectal cancer and renal diseases. Males demonstrated greater comorbidity burden, particularly involving metabolic disorders and advanced liver diseases. Notably, males with moderate-to-severe liver diseases had elevated HCC recurrence risk compared to females. Multidrug use rate among those with HCC recurrence was 79.5% (N=9999), of whom 25.7% with double-drug exposure, 13.8% with triple-drug exposure. Anti-diabetic drugs, ACEI/ARB, and diuretics for heart failure, anti-cancer drugs, and NSAIDs were the most common combination use. Both adjusted Cox regression models and subgroup analyses confirmed multi-comorbidity as a robust predictor of clinical outcomes.

**Conclusion:** Our gender and age-specific characterization of comorbidity and multidrug patterns in HCC provides clinicians and patients with actionable insights to guide proactive management and patient-centered care, enabling earlier comorbidity detection and prevention strategies.

**Highlights:**

- A Chinese population-level characterization of comorbidity profiles, multidrug combinations, and their interactions in HCC.
- Diabetes mellitus, hypertension, and existing chronic liver diseases were significant risk factors of HCC recurrence and mortality.
- Network analyses revealed sex/age/CCI-stratified associations between comorbidity-drug clusters and clinical outcomes.
- Identified comorbidity and multi-drug patterns showed prognostic utility for HCC recurrence and mortality risk prevention.

**Research in context:** *Evidence before this study:* According to our PubMed search (without language restriction) on March 30, 2025, using (“hepatocellular carcinoma” and “comorbidity” and “network”) or (“hepatocellular carcinoma” and “drug” and “network”) as search criteria, we found only two preliminary studies. However, they do not consider the heterogeneity of the network, nor the relationship between comorbidities or drugs and outcomes such as HCC recurrence or all-cause mortality. Therefore, to the best of our knowledge, there is no research study investigating age, sex, and CCI stratified network of multimorbidity and multi-drugs exposure, nor exploring the associations between co-comorbidities or co-drugs exposure and follow-up HCC recurrence and mortality risk.

*Added value of this study:* We proposed a dual perspective (comorbidity and drug) on primary and secondary outcomes by exploring the most frequent comorbidity and multidrug patterns in terms of patients initially diagnosed with HCC. In addition, we explored heterogeneity of comorbidities and drugs with outcome based on sex, age, and CCI. Followed by identifying risk factors associated with outcomes.

*Impact and implications:* Using Chinese population-based cohort data, we implement visualization and network analysis to identify comorbidity and multidrug combinations in patients with initial HCC. It can encourage health-care servers and people to provide proactive search for the presence of comorbidities and supply patient-centered treatment. Clinicians can consider a patients’ co-morbidities and medication history when evaluating patients with HCC, leading to safer and more effective treatment regimens.

**Graphical illustration abstracts:** 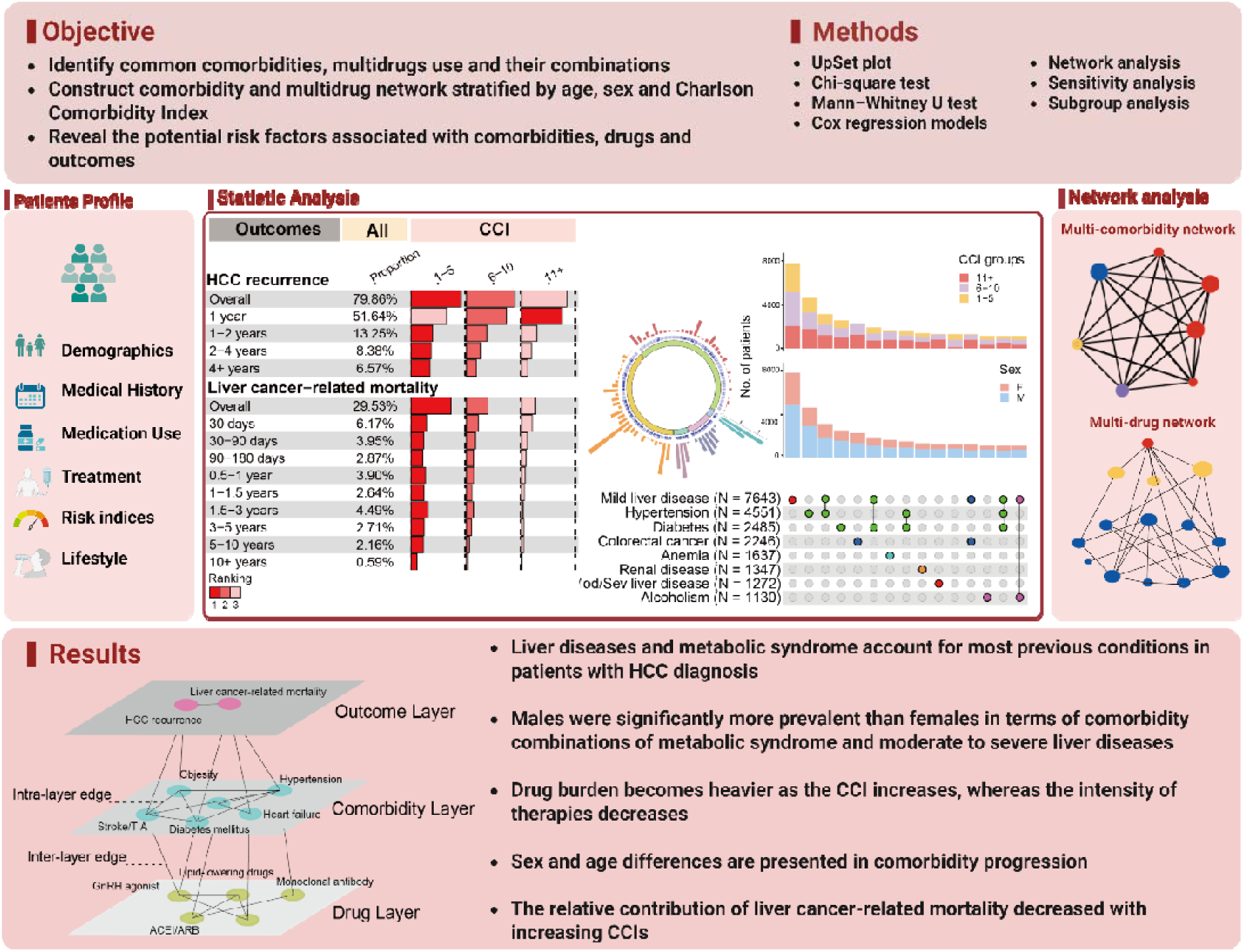

## Introduction

Liver cancer poses a significant global health threat, with an estimated 920 thousands individuals will experience death duo to liver cancer in 2030.^1^ HCC is the dominant kind of liver cancer, representing approximately 90% of all cases.^2^ About two-thirds of patients experienced HCC recurrence within 5 years after receiving curative therapy with resection or radiofrequency ablation. ^3^ Comorbidity is extremely common in patients with HCC, with 86.8% patients experiencing comorbidity combinations.^4^ Many studies has shown that diabetes is an independent risk factors for HCC recurrence.^5–7^ It is well-established that HCC recurrence is associated with chronic liver diseases, typically cirrhosis from nonalcoholic fatty liver disease,^8^ chronic hepatitis B^4,9,10^, chronic hepatitis C^10^. At the same time, the complex polypharmacy associated with presence of comorbidities, which may compromise therapeutic efficacy and exacerbate treatment-related toxicities.^11^

Contemporary healthcare systems remain predominantly anchored to single-disease paradigms, rendering them increasingly inadequate for addressing the complex challenges of multimorbidity management and prevention.^12^ Ignoring the impact of comorbidities is a loss, as studies have shown that comorbidities are associated with specific cancers^13,14^ and its recurrence^15^. Within HCC, comprehensive characterization of comorbidity patterns is essential for accurate prognostic stratification and therapeutic optimization. Comorbidities fundamentally reshape risk trajectories; notably, synergistic interactions between diabetes and obesity demonstrably elevate HCC recurrence risk and compromise survival following hepatic resection.^16^ Limited comorbidity awareness frequently excludes eligible patients from potentially curative therapies,^17^ and contributes to treatment-related toxicities (grade>3 adverse events occurring in 63.0% of combination therapy recipients).^18^ Systematic mapping of prevalent comorbidity clusters provides clinicians and patients with a proactive framework for early detection of associated conditions and personalized risk mitigation.

Despite the significance of comorbidity becoming more widely acknowledged, few studies had comprehensive explorations on it in HCC. For example, one study considered only 11 comorbidities and employed a simple frequency count approach, limiting both the applicability and comprehensiveness of its findings.^19^ Another study focused on liver-related comorbidity,^4^ neglecting the significant impact of extrahepatic comorbidities, such as metabolic syndrome^5,20,21^, cardiovascular diseases^22^, renal diseases^23^, anaemia,^20,21^ and gastrointestinal bleeding^24^, on overall survival in HCC patients. There were existing studies on network heterogeneity analysis considering sex^25^ or age^26,27^. However, limited studies capture the comprehensive associations between comorbidities, multidrug use, and clinical outcomes at the same time.

We addressed these gaps by leveraging real data from patients across 43 public hospitals in Hong Kong, capturing 31 comorbidities (e.g., metabolic syndrome, cardiovascular diseases, renal diseases, anaemia, gastrointestinal bleeding) among those with primary diagnosis of HCC. The Upset plot, an extension and visualization tool, was implemented to identify prevalent comorbidity patterns and multidrug combinations stratified by sex, age, and CCI. Our analysis included both primary and secondary outcomes. Network analysis and Cox regression models were performed to explore association between comorbidities, drugs and outcomes.

## Patients and methods

### Study population

This study was approved by the Institutional Review Board of The University of Hong Kong/Hospital Authority Hong Kong West Cluster Institutional Review Board (HKU/HA HKWC IRB) (UW-20-250) and complied with the Declaration of Helsinki. The requirement for informed consent was waived by the Institutional Review Board because the study utilized de-identified data. This was a retrospective population-based study of prospectively collected electronic health records using the Clinical Data Analysis and Reporting System (CDARS) by the Hospital Authority of Hong Kong. The records cover hospitalization in public hospitals in Hong Kong. This system has been used extensively by our teams and other research teams in Hong Kong, including diabetes and medication research.^28–30^

Patients who attended the Hospital Authority with a primary diagnosis of HCC between Jan 1, 2008, and Dec 31, 2019, were included, and were followed up until May 31, 2024. The following information was extracted: demographics, admission histories, past comorbidities, medication prescriptions, and laboratory examinations within 5 years before the initial diagnosis of HCC and from diagnosis until mortality or study end (May 31, 2024) were extracted. The following patient groups were excluded: (1) without complete demographics; (2) without mortality data records or missing follow-up during the study period; (3) non-adults under 18 years old at diagnosis. Finally, 15,998 patients with an initial primary diagnosis of HCC were included in this study for the analysis.

### Predictors and variables

Patients’ demographics, include sex and age at initial drug use (baseline), as well as clinical and biochemical data, were extracted for the present study. Prior comorbidities were identified using the *International Classification of Diseases Ninth Edition (ICD-9)* codes (**Table S1**). The Charlson Comorbidity Index was calculated.^31^ Additionally, histories of drug prescriptions were extracted, including angiotensin-converting enzyme inhibitors/angiotensin receptor blockers (ACEI/ARB), calcium channel blockers (CCBs), alpha blockers (AlphaBLKs), beta blockers (BetaBLKs), diuretics for hypertension, diuretics for heart failure, lipid-lowering drugs, statins and fibrates, nitrates, antibiotics, anticoagulants, eye drugs, antiestrogen, antiplatelets, anti-diabetic drugs, hormonal drugs, steroids, nonsteroidal anti-inflammatory drugs (NSAIDs), anti-cancer drugs (tyrosine kinase inhibitors, endocrine therapy, aromatase inhibitors, endocrine therapy excluding aromatase inhibitors, cytotoxic therapy, androgen deprivation therapy, targeted therapy, monoclonal antibodies, immunomodulatory drugs, tamoxifen, anti-HER2 monoclonal antibody, anthracycline, antimetabolite fluoropyrimidines, microtubule inhibitors, platinum, GnRH agonist, and immune checkpoint inhibitors).(**Table S2**)

Patient baseline laboratory examinations around HCC diagnosis, including complete blood counts, liver and renal functional tests, lipid and glucose profiles, were extracted.(**Table S3**) We also calculated the following clinical biomarkers: the albumin-bilirubin (ALBI) score, which is an indicator for liver function, calculated using linear model.^32^ The Fibrosis-4 score, which estimates the amount of scarring in the liver, was calculated.^33^ Aspartate aminotransferase-to-platelet ratio index (APRI) serves as a measure of liver health in patients with liver disease when aspartate aminotransferase (AST) levels are above the upper limit of normal.^34^ The serum aspartate (AST; U/L or IU/L) to alanine aminotransferase (ALT; U/L or IU/L) ratio (AST/ALT ratio) is a clinically validated biomarker for inferring the etiology of liver disease. For instance, clinically, elevated AST/ALT ratio demonstrates diagnostic utility: values >2.0 strongly correlate with alcoholic liver disease, whereas ratios <1.0 are associated with chronic viral hepatitis or cholestatic pathologies including primary biliary cholangitis.^35^ The alanine aminotransferase to platelet count ratio (ALT/PLT ratio) is a noninvasive biomarker primarily used to assess the risk and severity of liver fibrosis or cirrhosis. Here, ALT activity (expressed in U/L) is normalized to platelet counts (reported as ×10^9^/L or cells/μL, with 1 ×10^9^/L equal to 1,000 cells/μL). Urea-to-creatinine ratio (UCR) can assess kidney function and fluid status by distinguishing kidney problems from other types of dehydration or kidney injury.^36^ More details can be referred to ***Supplementary Methods***.

### Study outcomes

Comorbidity has consistently demonstrated a detrimental effect on cancer survival.^17,37^ An RCT study concluded that the degree of comorbidity may affect the recurrence of cancer.^15^ Therefore, our primary outcomes were HCC recurrence and liver cancer-related mortality after the primary diagnosis of HCC, defined using ICD*-9* codes 155.0, 155.2, 197.7) (**Table S1**). The secondary outcomes were cancer-related mortality, non-cancer-related mortality, and all-cause mortality. Mortality data were obtained from the Hong Kong Death Registry, a population-based official government registry containing the registered death records of all Hong Kong citizens linked to CDARS. Mortality was recorded using the *International Classification of Diseases Tenth Edition (ICD-10)* coding (**Table S1**). The endpoint date of interest for eligible patients was the date of the event. The endpoint for patients without a primary outcome was the date of death or the end of the study period (May 31^st^, 2024).

### Statistical analysis

Descriptive statistics were used to summarize baseline clinical and biochemical characteristics. Continuous variables were presented as medians (95% confidence interval [CI] or interquartile range [IQR]), and categorical variables were presented as frequency (percent). The Mann–Whitney U test was used to compare continuous variables. The χ2 test with Yates’ correction was applied to categorical variables in a 2□×□2 contingency table.

We define degree of comorbidity by the number of coexisting conditions: double, triple, quadruple, quintuple comorbidity. Similarly, we examined tuple and triple multidrug use in our study. Prevalent comorbidity patterns were identified in patients with recurrence risk (**Fig. S1**) and liver cancer-related mortality (**Fig. S2**). We also identified the 20 most common combinations of tuple, triple, quadruple, quintuple comorbidities (**Table S4-S7**) as well as the most prevalent tuple and triple multidrug combinations (**Table S8-S9**), in relation to primary and secondary outcomes. The common combination of comorbidities and multidrug use was presented in **Table S10-S13**.

### Cox regression models

The cumulative incidence curves for the primary and secondary outcomes were constructed (**Fig. S3, S4**, and **S5**). Cox proportional hazards regression models were employed to identify significant risk predictors of adverse study outcomes, with adjustments for demographic characteristics, comorbidities, medication use, renal function, lipid profiles, and glycaemic tests. Results are reported as HRs, 95% CIs, and P-values (significance threshold: two-sided P<0.05). Landmark analysis for CCI, drug combinations, calculated biomarkers,liver and renal functions, and lipid and glocuse profiles were conducted for sensitivity analysis, after excluding mortality within 30, 60, 90, 180 days, and 1 and 2 years post-HCC diagnosis. Interaction P-value (P*_interaction_*) was applied to examine the differences in the associations between the subgroups. All statistical analyses were performed with RStudio (Version: 1.1.456) and Python (Version: 3.6).

### UpSet plot analysis

UpSet plots provide an efficient visualization of comorbidity and multidrug combination frequencies through intersecting set representations. The intersection matrix (right) uses filled circles to denote set membership within specific combinations, with connecting lines illustrating combinatorial relationships^38^ Set cardinalities are shown as horizontal bars (left), while vertical bars (bottom) quantify intersection frequencies. All visualizations were generated using the ‘ComplexUpset’ package (Version 1.3.6) in R (Version 4.3.2). In addition, we identify the most frequent double, triple, quadruple, and quintuple patterns with higher incidence of outcomes among those with primary HCC.

### Network analysis

In comorbidity and multi-drug exposure network analysis, nodes represent diseases or drugs, and edges mean the co-occurrence pattern. Wider edges represent stronger associations and higher co-occurrence frequencies for that pattern. We normalized nodes and edge size across different sample sizes to make networks comparable. Degree centrality is a measure of the importance of a node in network structure based on the number of direct edges connected to it. It has generally been extended by weighted degree centrality,^39^ which considers the weights of the edges connected to a node and is the focus of our analysis. Network analyses were performed using the package of ‘igraph’ (Version 1.5.1) in R (Version 4.3.2).

### Subgroup analysis

The subgroup analysis evaluated the consistency of CCI as a predictor for primary and secondary outcomes according to sex, age group (<65 or ≥65 at HCC diagnosis), previous drug classes (0,1 or ≥2), ACEI/ARB, CCBs, BetaBLKs, diuretics for hypertension, diuretics for heat failure, anti-diabetic drugs, NSAIDs, anti-cancer drugs, eGFR (Glomerular Filtration Rate) (<30 or ≥30) calculated by CKD-EPI (Chronic Kidney Disease Epidemiology Collaboration)^40^. The results of analysis were expressed as count, percentage, hazard ratios with 95% confidence intervals, P-value, and P*_interaction_*. P-value determines whether the association between the CCI and outcomes within an individual subgroup is statistically significant. P*_interaction_* assesses if there are significant differences across CCI subgroups.

## Results

### Patient characteristics and outcome analysis

No significant differences were observed in characteristics between patients with HCC recurrence (**Fig. 1A**) and liver cancer-related mortality (**Fig. 1B**). In the overall cohort, HCC recurrence rates demonstrated an inverse relationship with CCI severity, declining progressively from 85.8% (CCI 1-5) to 79.8% (CCI 6-10) and 69.3% (CCI ≥11). (**Fig. 1C**). This pattern persisted across all recurrence intervals except early recurrence (<1 year post-diagnosis). Paradoxically, while higher CCI typically indicates poorer health status, recurrence rates decreased with increasing comorbidity burden. This apparent contradiction likely reflects survival bias, evidenced by the CCI ≥11 subgroup’s substantially higher all-cause mortality (97.3%). Thus, patients in CCI groups (≥11) may not survive long enough to experience HCC recurrence. We also explored this by sensitivity analyses afterwards. Furthermore, patients with CCI ≥11 were at elevated risk of non-liver-cancer-related mortality, as indicated by a high cumulative hazard of cancer-related mortality but a low cumulative hazard of liver cancer-related mortality. (**Fig. S3**)

**Fig. 1.**
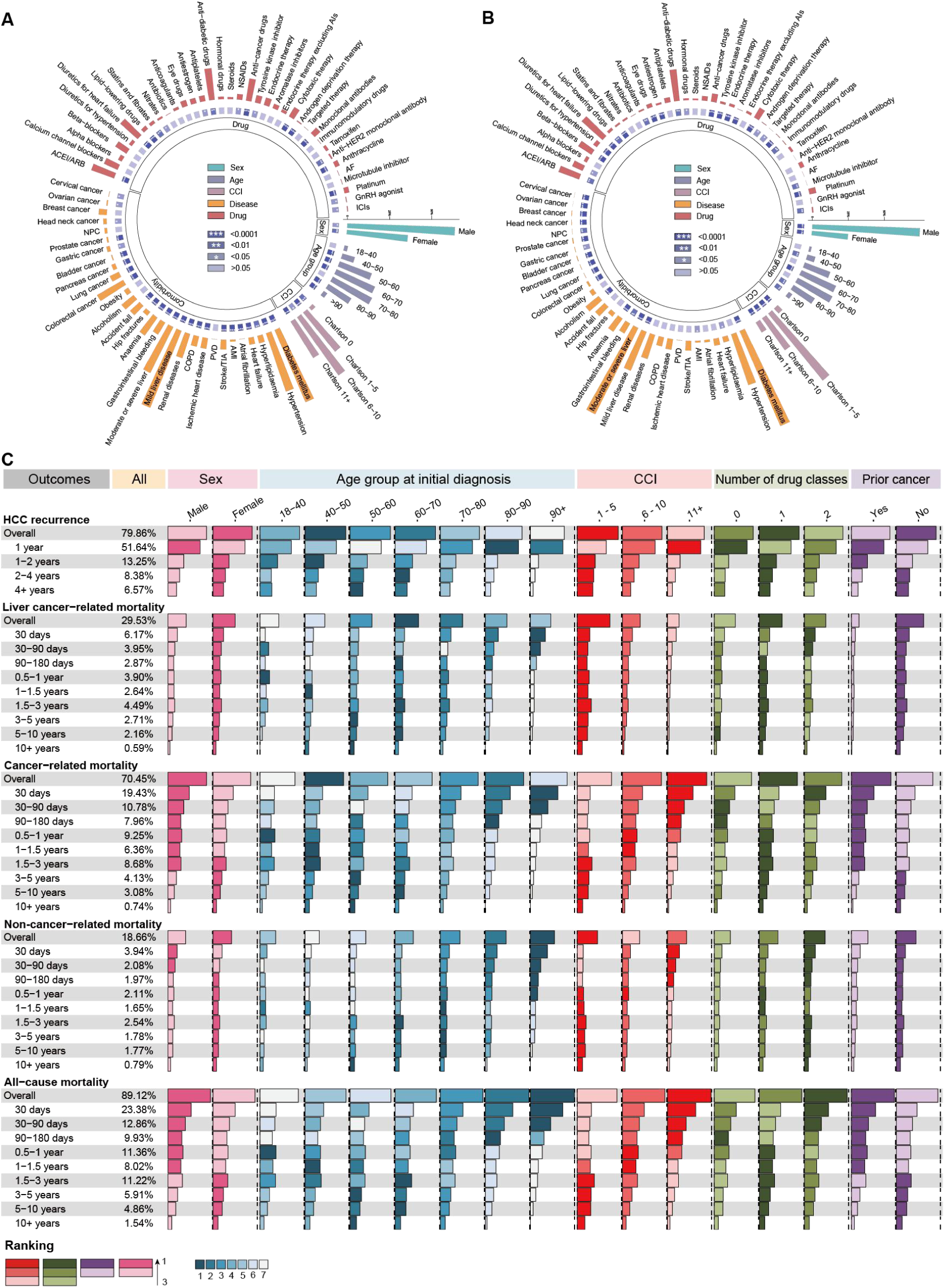
Descriptive statistics for different outcomes. (A) The distribution of characteristics in patients with HCC recurrence. The length of each bar represents the percentage of characteristic values accounting for the whole patients with HCC recurrence. The number of asterisks with colored cubes indicates the significance level in characteristic values derived by using χ2 test with Yates’ correction. Significance levels were calculated to determine whether the difference in characteristics is significant between HCC recurrence patients and non-HCC recurrence patients. (B) The distribution of characteristics in patients with liver cancer-related mortality. (C) Ranking results of patients’ characteristic values in primary and secondary outcomes. Characteristics include sex, age group at initial HCC diagnosis, CCI, number of drug classes, and prior cancer. The result should be viewed horizontally. Different colors represent the ranking of the characteristic. The length of the horizon bar depicts the percentage of the feature.

### Comorbidity and multidrug exposure patterns

Common comorbidities and comorbidity combinations derived from patients with HCC recurrence were identified, demonstrating that metabolic syndrome and liver diseases accounted for majority of previous conditions. Many HCC recurrence events were observed among patients with previous mild liver disease (6189[49.2%] of 12571 individuals with HCC recurrence), diabetes mellites (5864[46.6%]), hypertension (4589[36.5%]), and moderate/severe liver disease (3834[30.4%]), as shown in **Fig. 2A**. The comorbidity rate (at least one previous disease diagnosis) among patients with HCC recurrence is 93.1% (N=11705), of whom 80.3% (N=9409) had 1-4 diseases. The single-comorbidity rate is 17.3% (N=2176), double comorbidity rate is 24.5% (N=3083), and triple comorbidity rate is 18.7% (N=2351).

**Fig. 2.**
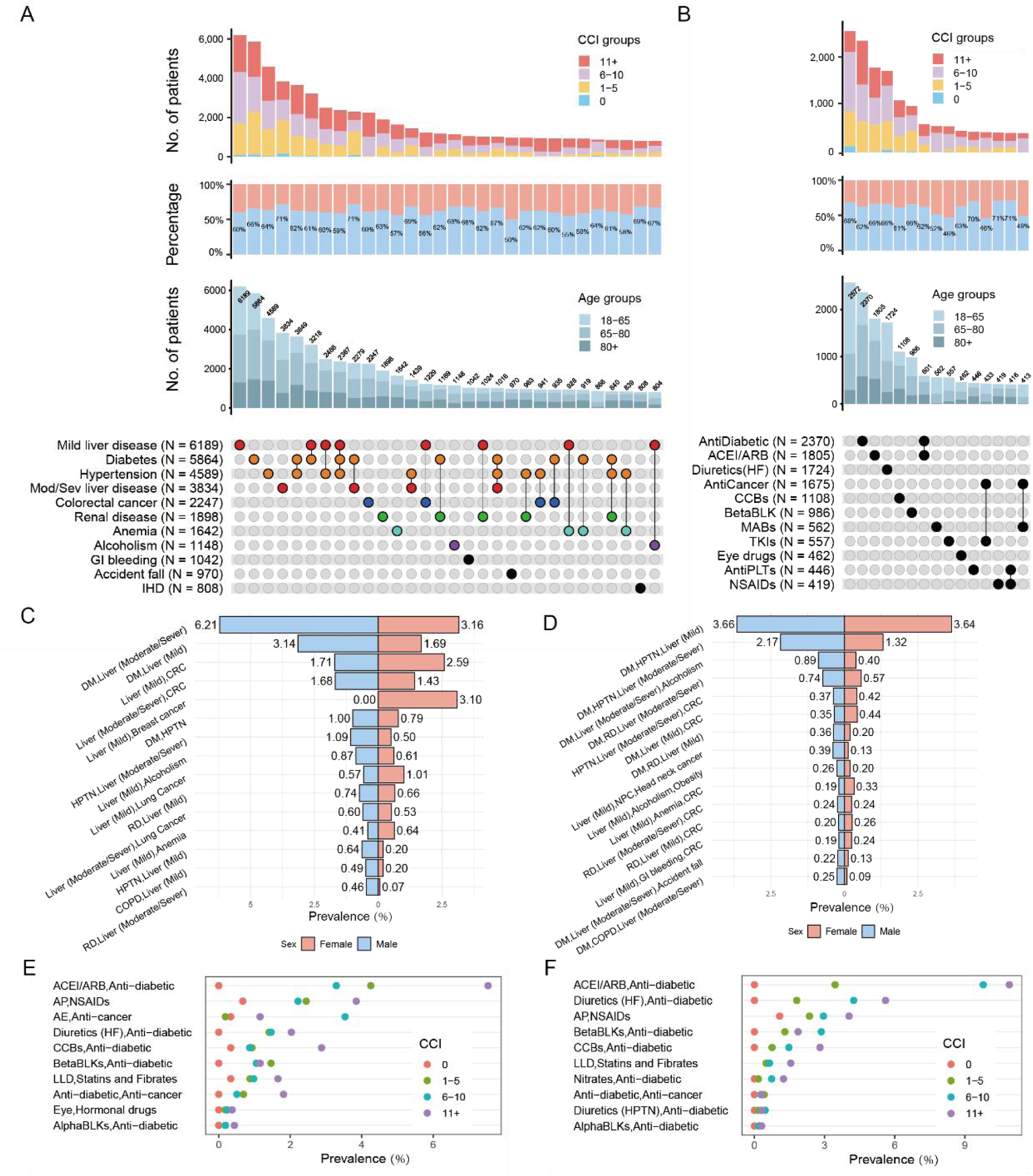
Comorbidity and co-drug exposure patterns in HCC patients with subsequent recurrence. (A) The UpSet plot of comorbidity intersections in patients with HCC recurrence. Each row indicates a disease, with the number of patients diagnosed with that disease in parentheses. Each column represents the intersection (or lack thereof) of one or more historical disease, shown through the colored, filled-in, and connected circles. The stacked bar chart illustrates the number of patients with comorbidities, categorized by CCI and sex within the current intersection sets. (B) The UpSet plot of co-drugs intersections in patients with HCC recurrence. (C) Top 15 common double comorbidity combinations among males and females. (D) Top 15 common triple comorbidity combinations among males and females. (E) Top 10 common double multidrug combinations stratified by CCIs in patients with HCC recurrence. (D) Top 10 common double multidrug combinations stratified by CCIs in patients with liver cancer-related mortality.

**Fig. 3.**
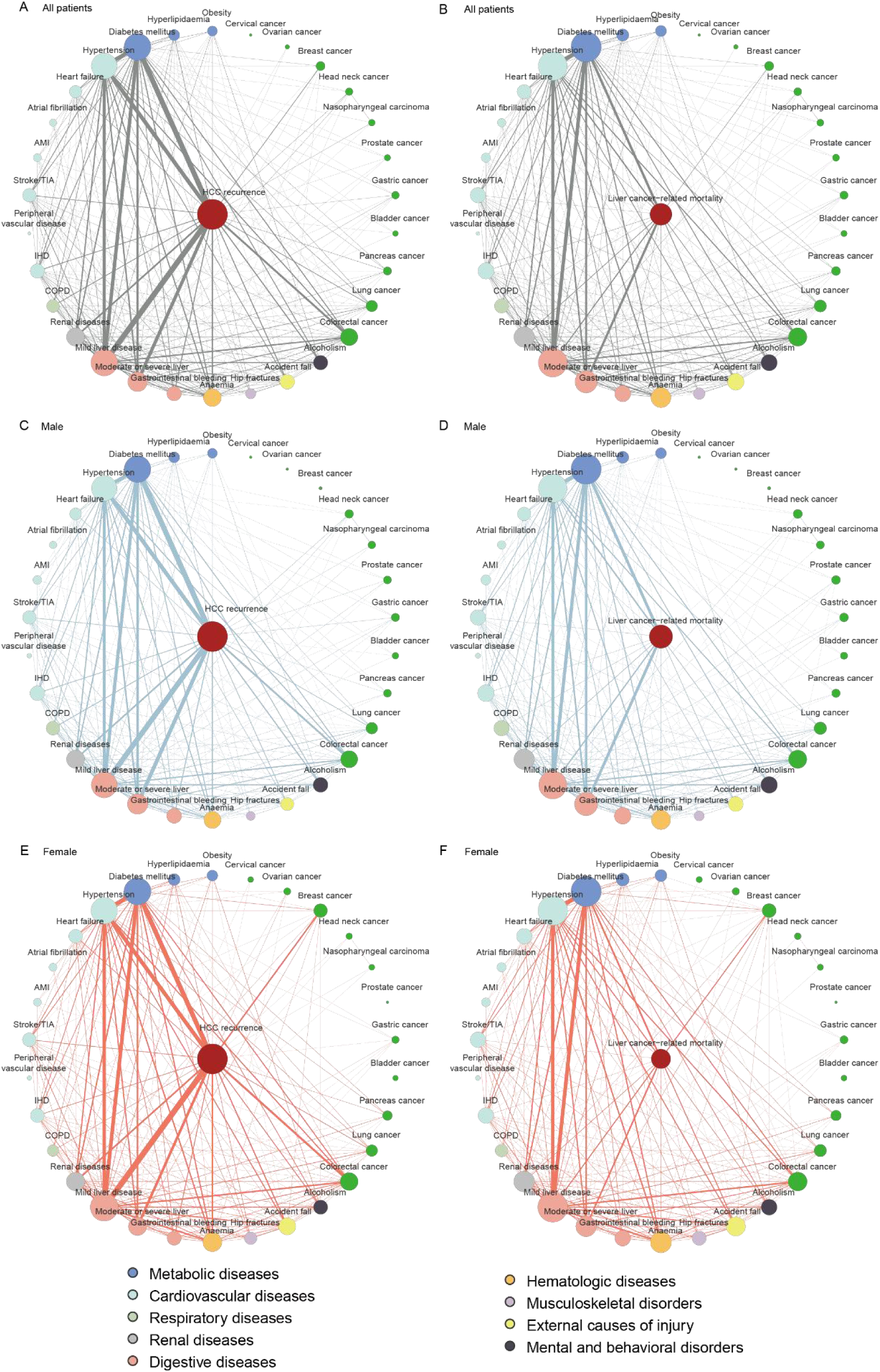
Comorbidity networks in all patients and sex subgroups. (A) comorbidity network centered on HCC recurrence in all patients; (B) comorbidity network centred on liver cancer-related mortality in all patients; (C) comorbidity network centred on HCC recurrence in male patients; (D) comorbidity network centred on liver cancer-related mortality in male patients; (E) comorbidity network centred on HCC recurrence in female patients; (F) comorbidity network centred on liver cancer-related mortality in female patients.

**Fig. 4.**
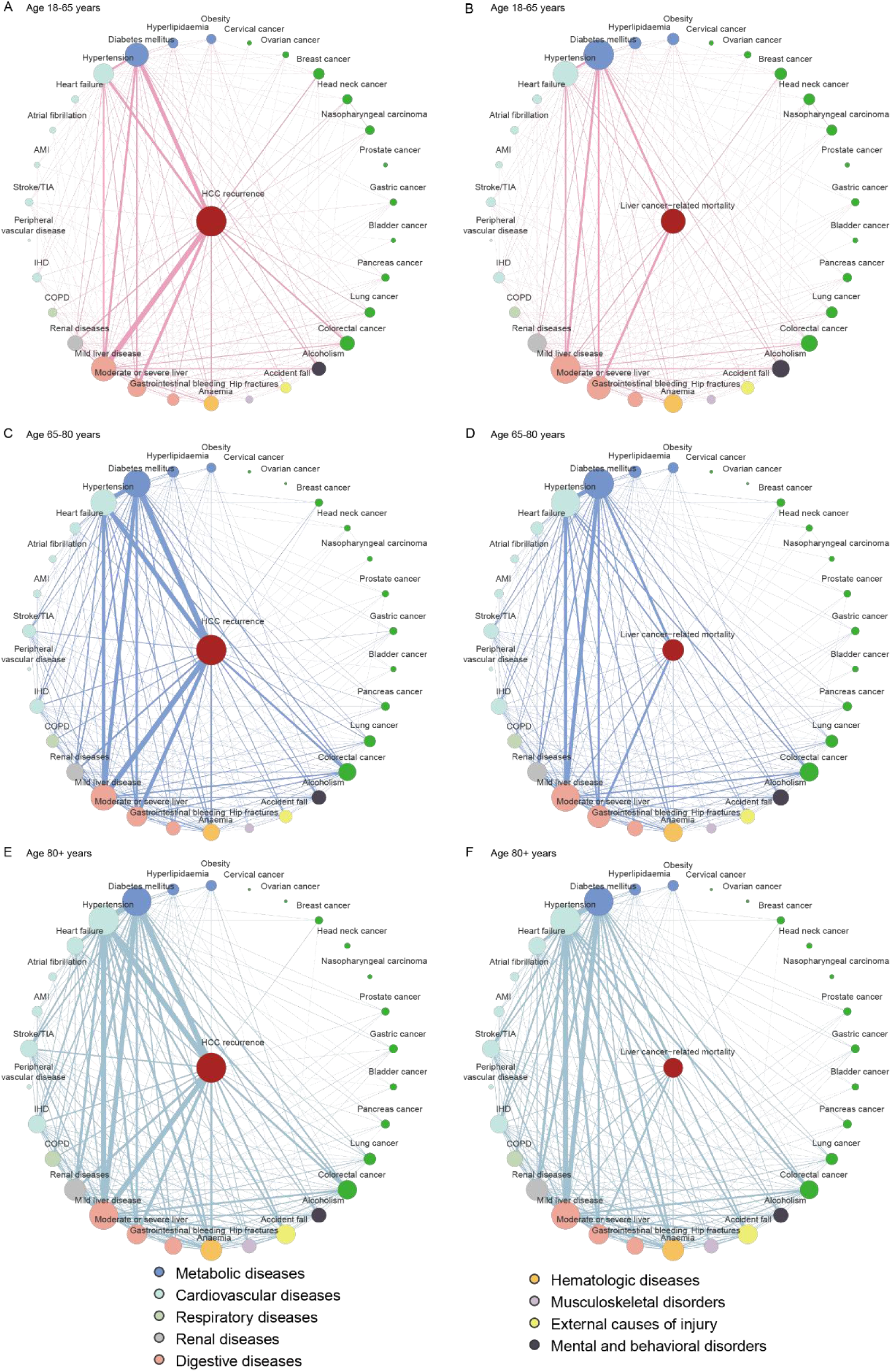
Comorbidity networks in age subgroups. (A) comorbidity network centered on HCC recurrence in patients aged 18-65; (B) comorbidity network centred on liver cancer-related mortality in patients aged 18-65; (C) comorbidity network centred on HCC recurrence in patients aged 65-80; (D) comorbidity network centred on liver cancer-related mortality in patients aged 65-80; (E) comorbidity network centred on HCC recurrence in patients aged over 80; (F) comorbidity network centred on liver cancer-related mortality in patients aged over 80.

Most prevalent comorbidity pairs observed were combinations of diabetes mellitus (DM) with moderate or severe liver diseases (N=642, 20.8% of the total 3083 patients with double comorbidity), diabetes mellitus with mild liver disease (N=329, 10.6%), followed by mild liver disease with colorectal cancer (N=255, 8.2%) as shown in **Table S4**. Majority triplet comorbidity combinations in patients with HCC recurrence were diabetes mellitus, hypertension with mild liver disease (N=459, accounting for 19.5% of total 2351 patients with triple comorbidity), diabetes mellitus, hypertension with moderate to severe liver diseases (N=234, 9.9%), followed by diabetes mellitus, moderate to severe liver diseases with alcoholism (N=89, 3.7%) as illustrated in **Table S5**. We performed common comorbidity and multidrug patterns in patients with liver cancer-related mortality by UpSet plot (**Fig. S1 and S2**). Cumulative incidence curves for primary and secondary outcomes were presented, with stratification analysis of CCI (**Fig. S3**), age (**Fig. S4**), and sex (**Fig. S5**) at HCC diagnosis.

We identify the top 20 most common double, triple, quadruple, quintuple comorbidity patterns among patients with HCC diagnosis across primary and secondary outcomes. (**Table S4-S7**). From the common comorbidity pattern we listed, diabetes mellitus and liver diseases emerged as predominant comorbidity combinations, consistently associated with all outcomes except non-cancer-related mortality. Among patients with pre-existing DM and HPTN, mild liver diseases demonstrated stronger association with HCC recurrence, evidenced by the incidence rate of 3,651 per 100,000 persons. This exceeds the second comorbidity patterns moderate to severe liver diseases with the incidence rate of 1,861 per 100,000 persons. Conversely, moderate to severe liver diseases dominate liver cancer-related mortality. The comorbidity pattern of DM-HPTN-moderate to severe liver diseases exhibited an incidence of 5,801 per 100,000, dramatically higher than the DM-HPTN-mild liver diseases pattern (2.598 per 100,000). This finding is consistent with prior cohort study indicating that moderate to severe liver diseases (OR: 3.19, CI: 2.833.58, P < 0.05) shows higher odds of mortality compared to mild liver diseases (OR: 1.38, CI: 1.23-1.57, P < 0.05).^41^

Sex-specific analyses reveal significant differences in comorbidity patterns. Males exhibited a higher burden of pre-existing comorbidity combinations of metabolic syndrome and moderate to severe liver diseases compared to females, with male-to-female prevalence ratios (PR) exceeding 2 for specific combinations: hypertension (HPTN) with moderate to severe liver diseases:1.09% in males vs 0.5% in females (PR = 2.1), HPTN with mild liver diseases: 0.64% in males vs 0.2% in females (PR = 3.2), Chronic obstructive pulmonary disease (COPD) with mild liver diseases:0.49% in males vs 0.2% in females (PR = 2.4), renal diseases (RD) with moderate to severe liver diseases:0.46% in males vs 0.07% in females (PR = 7.0), as illustrated in **Fig. 2C**. Comorbidity burden among males remained significant in triple comorbidity combinations, for example: DM, moderate to severe liver diseases, alcoholism: 2.17% in males vs 1.32% in females (PR = 2.2), mild liver diseases, nasopharyngeal carcinoma, head neck cancer: 0.39% in males vs 0.13% in females (PR = 2.9), DM, COPD, moderate to severe diseases: 0.25% in males vs 0.09% in females (PR = 2.8) (**Fig. 2D**).

The multidrug use rate among patients with HCC recurrence is 79.5% (N=9999), of whom 46.2% (N=4617) were exposed with at least double-drug therapy, 25.7% (N=2574) were administered double-drug exposure, 13.8% (N=1380) were given triple-drug exposure. Anti-diabetic drugs, ACEI/ARB, and diuretics for heart failure were the most common drug use in patients with HCC recurrence (**Fig. 2B**). Additionally, the most prevalent double-drug therapy used were anti-diabetic drugs with ACEI/ARB (N=576, accounting for 26.6% of total 2165 patients with double-drug use), and anti-cancer drugs with NSAIDs (N=335, 15.4%) showed in **Table S9**. Most common triple-drug combination was eye drugs, hormonal drugs, and steroids (N=134, representing 27.1% of the total 493 patients with triple-drug use) and more detailed are provided in **Table S10**. The prevalence of drug use increased with rising CCI in multidrug combinations. Patients in high CCI subgroups consistently demonstrate significantly greater rates of multidrug use compared to those in lower CCI subgroups, with CCI 11+ subgroups exhibiting the highest drug prevalence across nearly all drug combinations (**Fig. 2E and 2F**).

Common double drug-and triple-drug combinations in patients diagnosed with HCC diagnosis for primary and secondary outcomes can be found in **Table S8 and S9**. Several high-risk multidrug uses were be found, such as antiplatelets with NSAIDs, ACEI/ARB with anti-diabetic drugs. Such combinations are very problematic, as previous studies have demonstrated a significantly increased risk of serious adverse events, including gastrointestinal bleeding and renal dysfunction.^42,43^ We also identified the top 20 double, triple, quadruple, and quintuple comorbidity-drug patterns among patients with different outcomes (**Table S10-S13**). It illustrates that patients with severe liver diseases were exposed to multidrug use. The management of comorbidities should be prioritized in patients with moderate to severe liver diseases, whereas in those with mild liver diseases, concurrent management of cancer may be considered.

### Characteristics stratified by CCI

Patients’ characteristics stratified by CCI categories were presented in **Table 1**. It suggests that the comorbidity burden actively guided the reduction in therapeutic intensity. Specifically, platinum therapy utilization declined across CCIs. (CCI 1-5: 5.96%; CCI 6-10: 1.65%; CCI 11+: 0.17%). Similarly, cytotoxic therapy depicts a decline trend (CCI 1-5: 9.22%; CCI 6-10: 6.10%; CCI 11+: 1.27%). These patterns indicate increasingly conservative selection of high-intensity regimens for patients with elevated comorbidity burdens in clinical practice. Descriptive statistics were also conducted to analyze patients’ characteristics based on the presence or absence of HCC recurrence (**Table S14**), baseline exposure of drug classes number (**Table S15**), mortality outcome among patients after HCC diagnosis (**Table S16**), age at initial diagnosis of HCC (**Table S17**). Univariable Cox regression models identify significant risk predictors of secondary outcomes (**Table S18**).

**Table 1.**
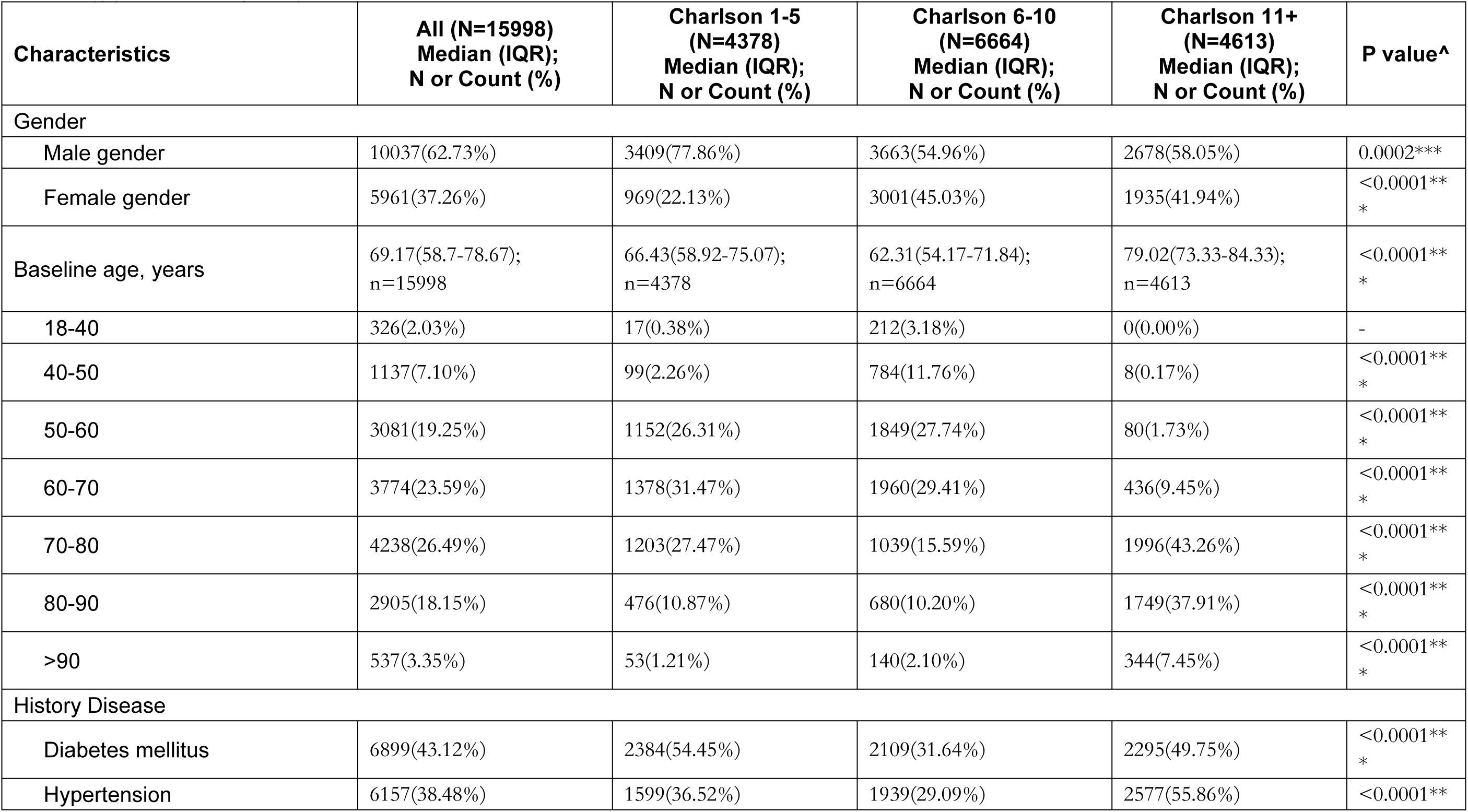
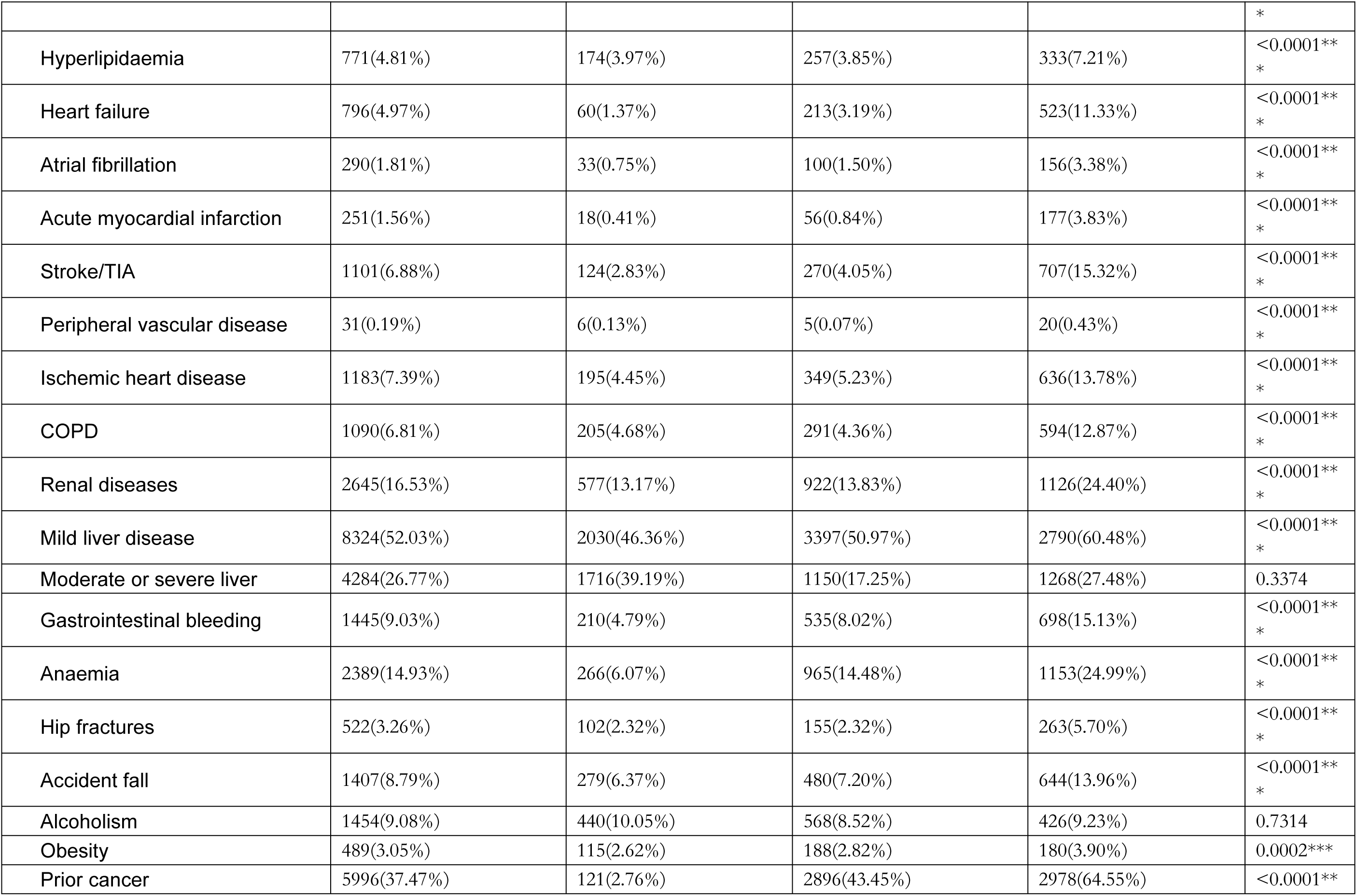
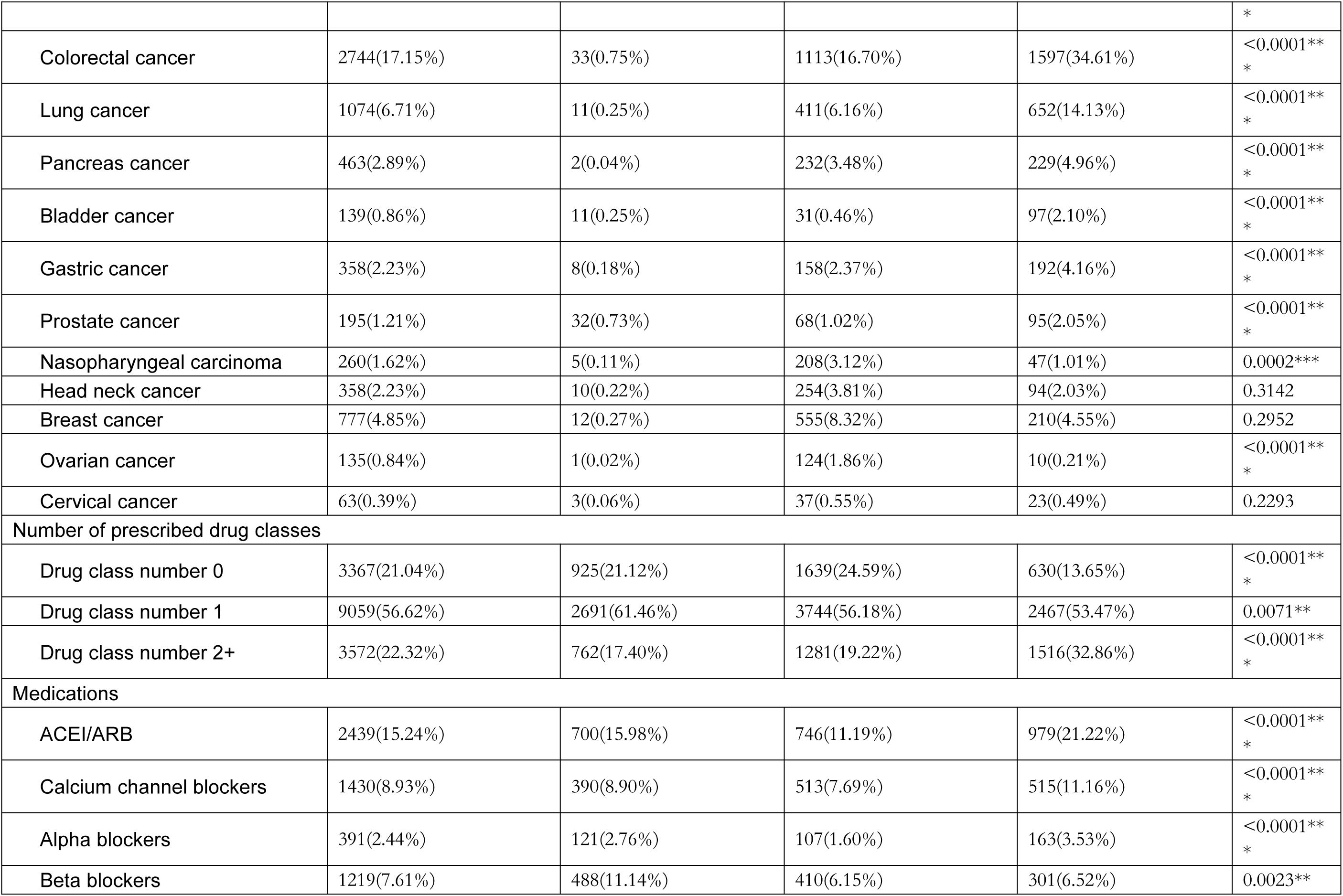
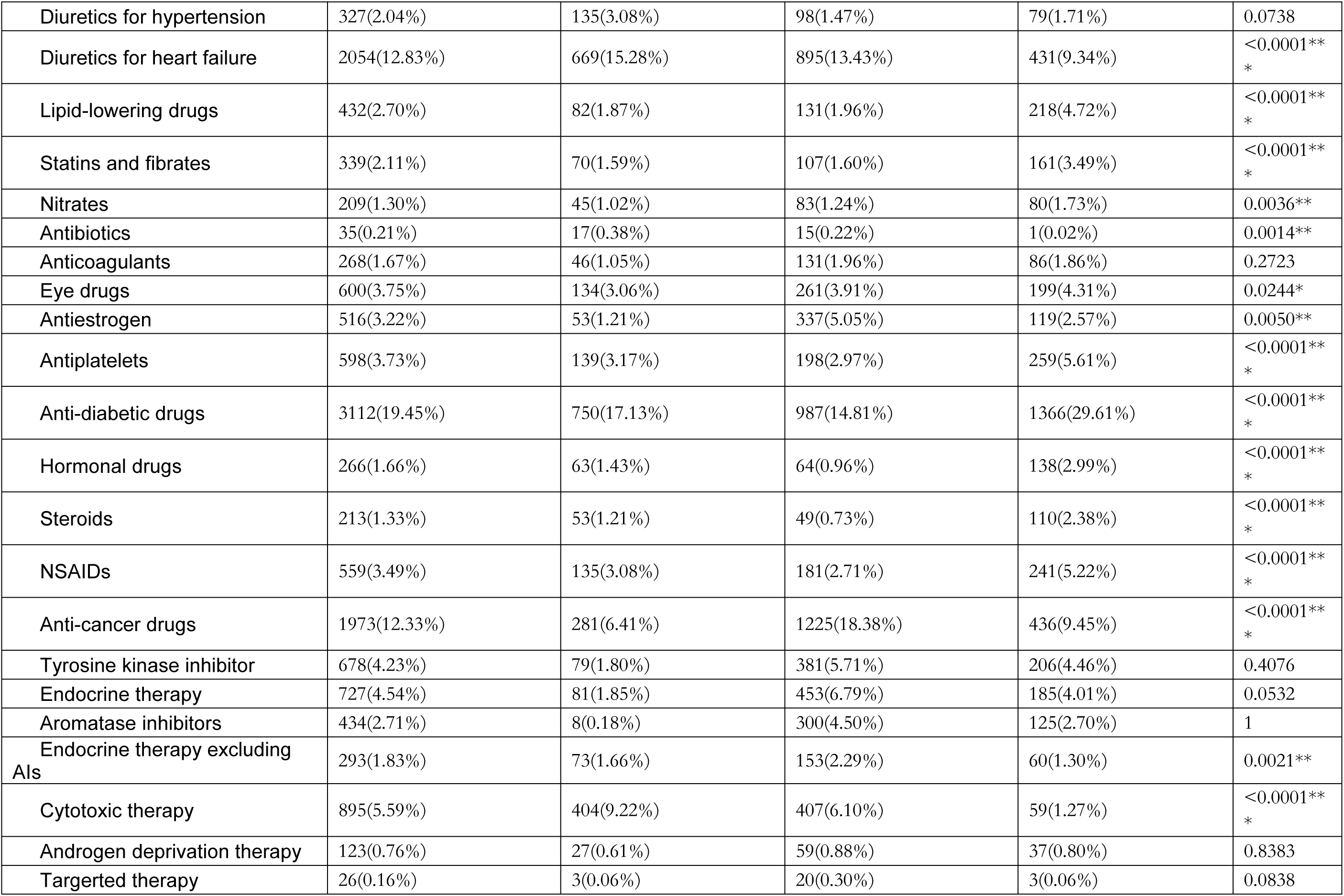
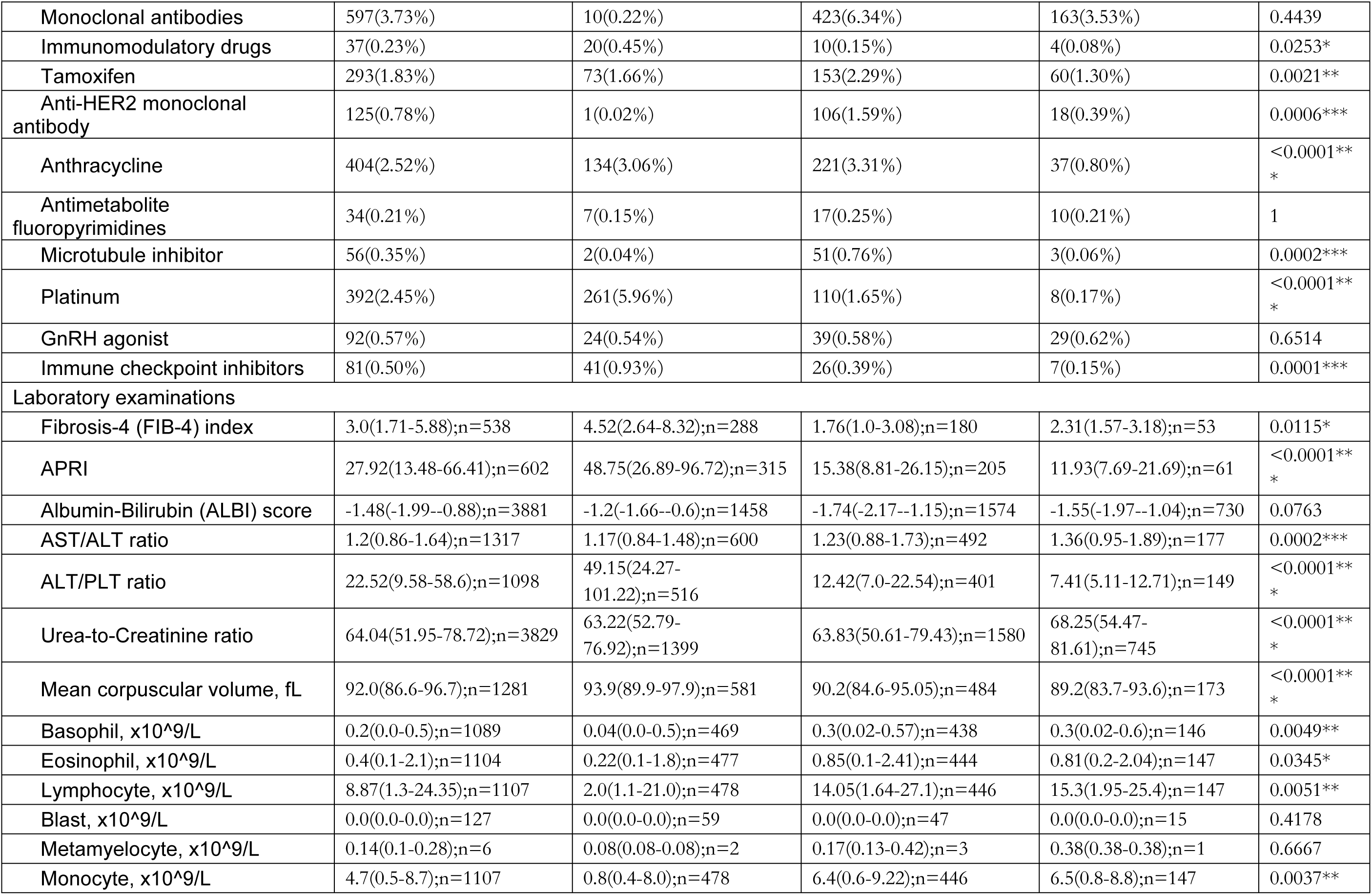
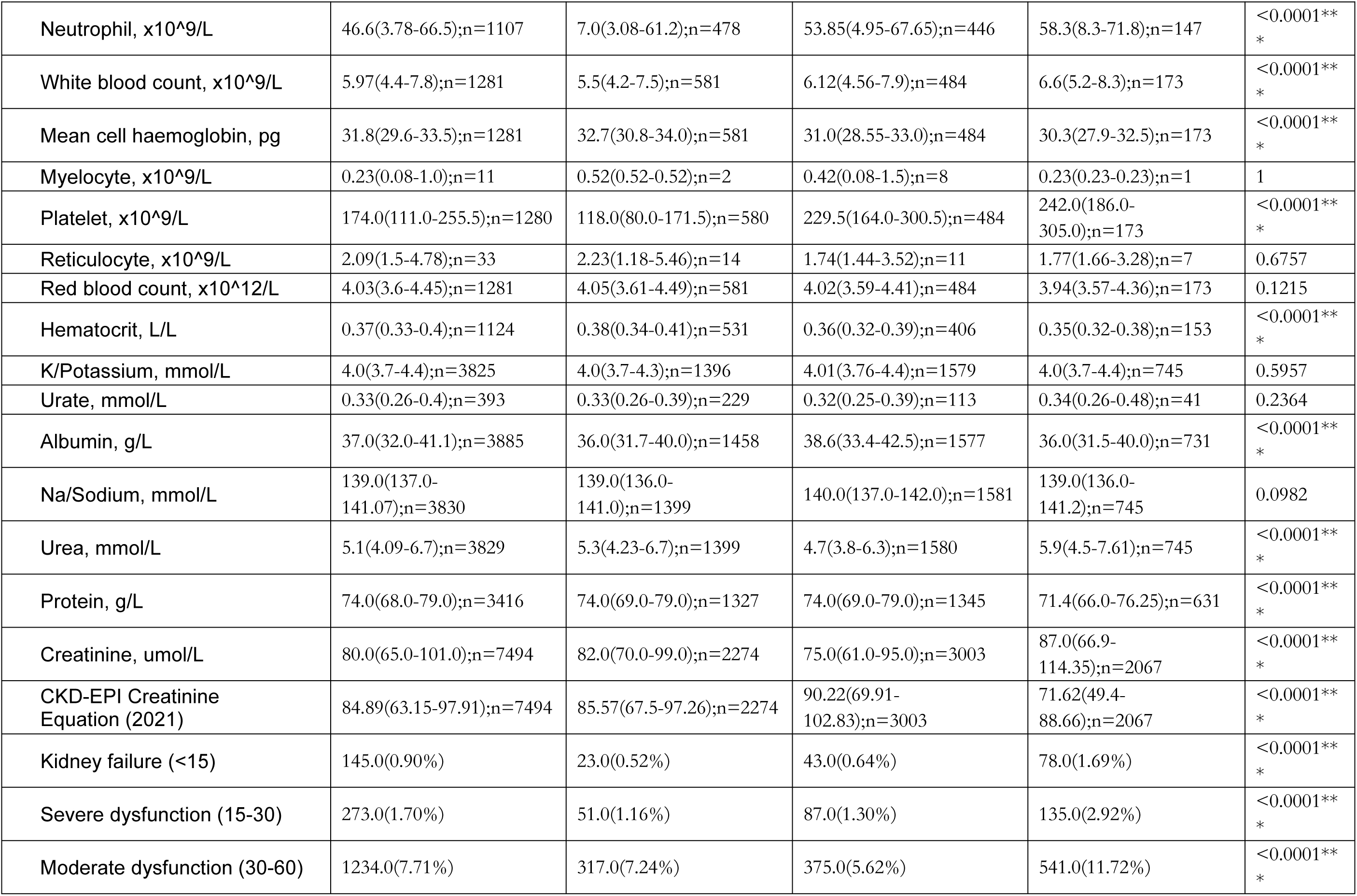
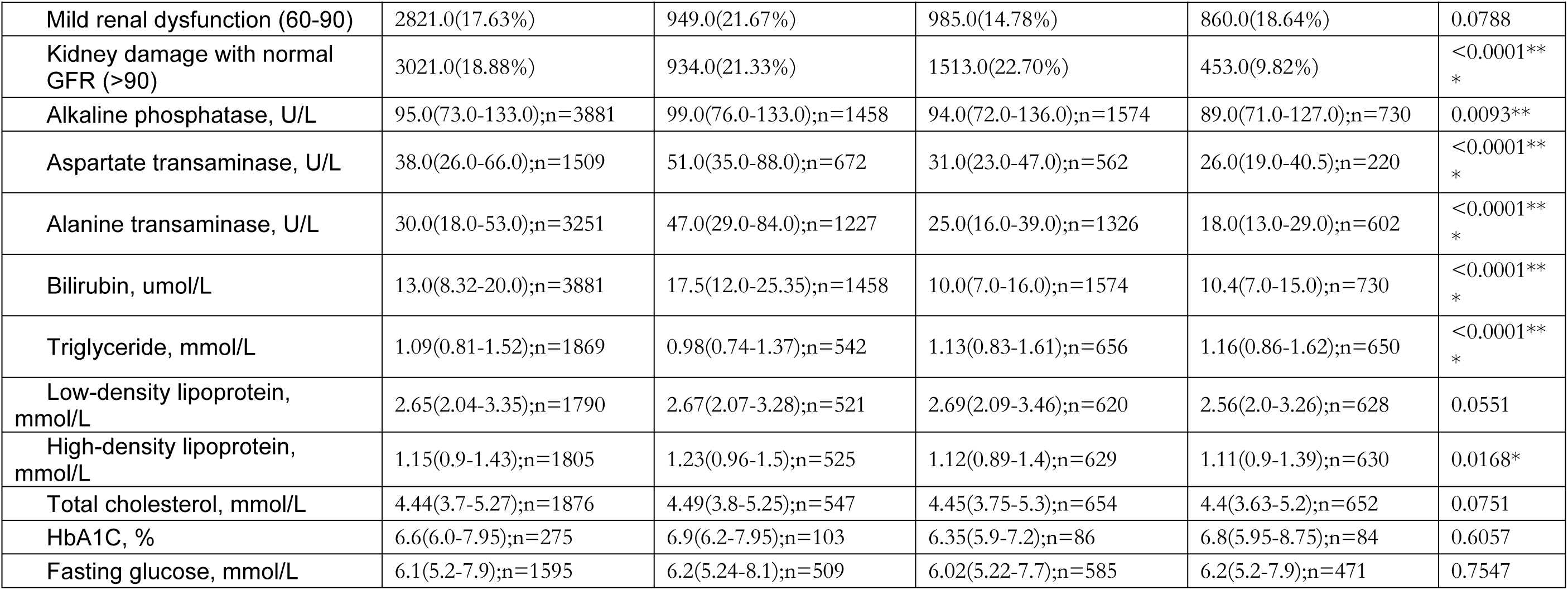
Descriptive summary of patient characteristics stratified by CCI. ^ P value indicates statistical difference among patients of three CCI subgroups. * for p≤ 0.05, ** for p ≤ 0.01, *** for p ≤ 0.001. Abbreviations: IQR (interquartile range); TIA (transient ischemic attack); COPD (chronic obstructive pulmonary disease); ACEI (angiotensin-converting enzyme inhibitors); ARB (Angiotensin Receptor Blockers); NSAIDs (nonsteroidal anti-inflammatory drugs); AST (aspartate transaminase); APRI (AST to platelet ratio index); ALT (alanine transaminase); PLT (platelet); CKD-EPI (Chronic Kidney Disease Epidemiology Collaboration); GFR (Glomerular Filtration Rate); HbA1C (glycated hemoglobin).

### Sex-stratified comorbidity networks

Significant gender-specific differences in comorbidities and comorbidity patterns were observed (**Fig. 3**). Patients with previous accident fall and hip fractures were more popular in females. Hip fractures had a higher prevalence in female (305/5961, 5.1%) than in males (194/10037; 1.9%) (**Fig. 3E)**. Breast cancer was more frequent in females (4/10037,0.04% for males; 773/5961, 13.0% for females) in **Fig. 3E**. Conversely, males were more likely to have head neck cancer (290/10037, 2.9% for males; 68/5961, 1.1% for females) and nasopharyngeal carcinoma (207/10037, 2.1% for males; 53/5961, 0.9% for females). (**Fig. 3C**). Males with moderate/severe liver diseases were at a higher risk of experiencing HCC recurrence than females (2738; 2738/10037, 27.3% in males, 1096; 1096/5961, 18.4% in females) as shown in **Fig. 3C and 3E**. Patients with a previous of mild liver disease (6189/15998, 38.7%), diabetes (5864/15998, 36.7%), hypertension (4589/15998, 28.7%) or moderate/severe liver disease (3834/15998, 24.0%) were more likely to experience HCC recurrence, represented by wider edges between those with HCC recurrence node. (**Fig. 3A**) Patients with liver cancer-related mortality (3965/15998, 24.7%) were less than patients with HCC recurrence (12571/15998, 78.5%) (**Fig. 3A and 3B)**. Liver cancer-related mortality was more likely observed in patients with a history of DM and moderate or severe liver diseases (**Fig. 3B)**.

### Age-stratified comorbidity networks

Network density increases with age, demonstrating more complex co-occurrence patterns of medical conditions with increasing age. Prior nasopharyngeal carcinoma (203/6466, 3.1%), cervical cancer (33/6466, 0.5%), ovarian cancer (106/6466, 1.6%) and breast cancer (525/6466, 8.1%) were more prevalent in patients before being diagnosed with HCC at 18-65 years old, compared to those over 65 years (NPC: 57/9532, 0.6%, cervical cancer: 30/9532, 0.3%, ovarian cancer: 29/9532, 0.3%, breast cancer: 252/9532, 2.6%) (**Fig. 4A)**. Patients with HCC who already have liver diseases, DM, HPTN were at a high risk of encountering HCC recurrence (**Fig. 4A,4C and 4E**). Patients with existing DM and moderate/severe liver disease were more likely to pass away from liver cancer causes (**Fig. 4B, 4D, and 4F**).

### CCI-stratified drug networks

To illustrate the differences in drug networks by CCI subgroup (**Fig. 5**), we summarize the findings below. The node of patients aged over 80 (2093/4613, 45.4% for CCI:11+, 820/6664, 12.3% for CCI:6-10; 529/4378,12.1% for CCI:1-5) increases in size with higher CCI, while the node of patients between 18 and 65 decreases as CCI increases. The prevalence of cytotoxic and platinum therapies decreased as CCI increased (261/4378, 6.0% for CCI:1-5; 110/6664, 1.7% for CCI:6-10; 8/4613, 0.17% for CCI:11+).(**Fig. 5A,5C and 5E**) However, the multidrug patterns of antiestrogen with ant-cancer drugs (337/6664=5.0% vs 53/4378=1.2%), antiestrogen with aromatase inhibitors (237/6664=3.6% vs 7/4378=0.16%), antiestrogen with endocrine therapy (336/6664=5.0% vs 53/4378=1.2%) were more prevalent in patients with CCI 6-10 than CCI 1-5. HCC patients who have ever taken anti-cancer drugs (1675/1973, 84.8%), and diuretics for heart failure (1724/2054, 83.9%), anti-diabetic drugs (2370/3112, 76.1%), ACEI/ARB (1805/2439, 74.0%) were more likely to experience HCC recurrence. We discovered that the size of node representing liver cancer-related mortality declined with increasing CCI, suggesting that there was a decrease in contribution of death due to liver cancer with increasing CCI (**Fig. 5B, 5D, and 5F**). This finding is plausible, as patients with a high CCI usually had multiple comorbidities and were likely to die from other serious illnesses, thereby reducing the proportion of deaths from liver cancer.

**Fig. 5.**
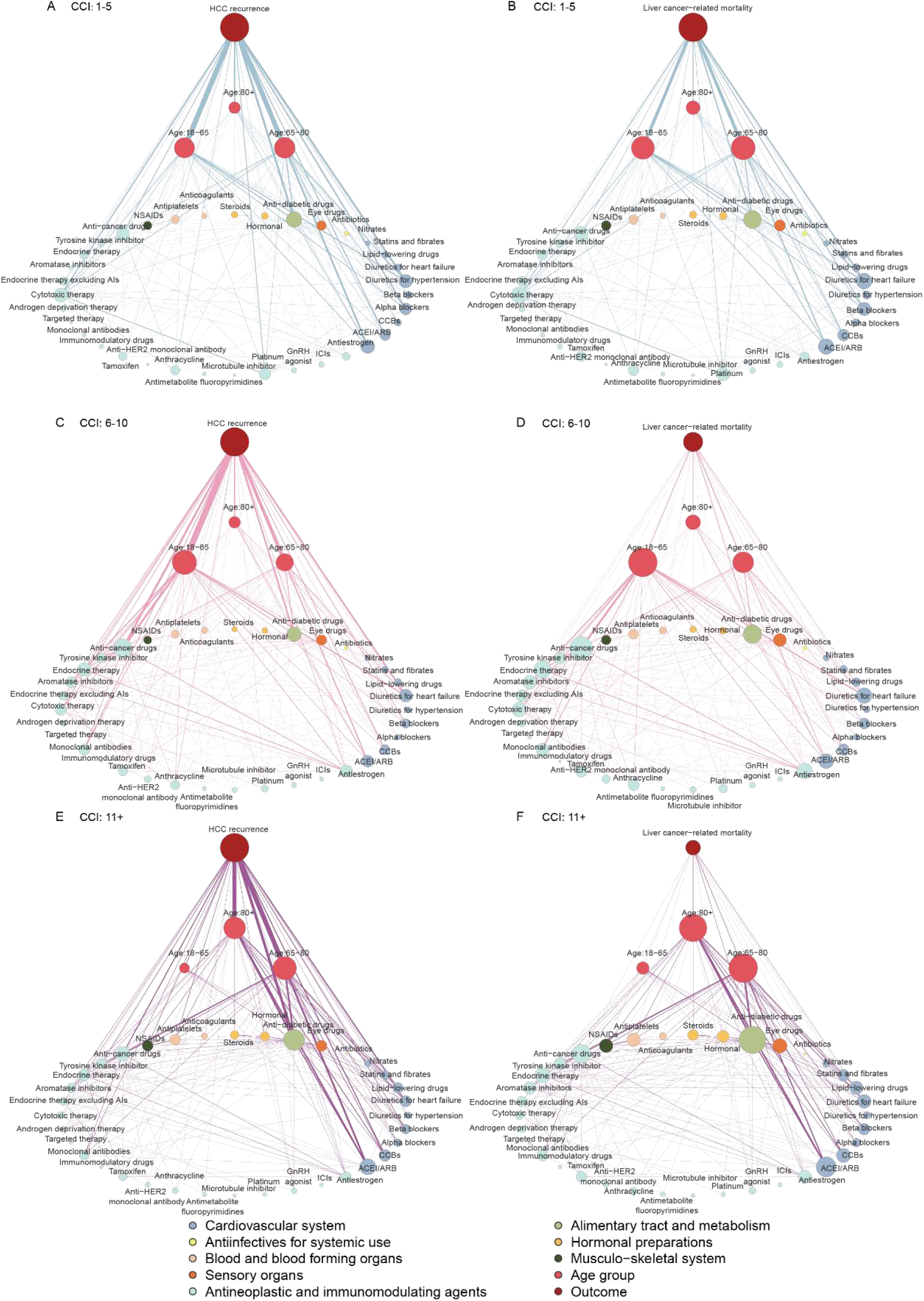
Drug network in CCI and sex subgroups. (A) drug network centered on HCC recurrence in patients with CCI 1-5; (B) drug network centred on liver cancer-related mortality in patients with CCI 1-5; (C) drug network centred on HCC recurrence in patients with CCI 6-10; (D) drug network centred on liver cancer-related mortality in patients with CCI 6-10; (E) drug network centred on HCC recurrence in patients with CCI over 11; (F) drug network centred on liver cancer-related mortality in patients with CCI over 11.

### Cox regression analysis

In the univariate analysis cox regression analysis for liver cancer-related mortality, males (HR [95%CI]: 1.51[1.40-1.63]; p < 0.001) was associated with increased risk compared to females. Among comorbidities, moderate to severe liver disease (9.36[8.67-10.10]; p<0.001) and diabetes mellitus (3.56[3.30-3.83]; p<0.0001) were the strongest comorbidity risk factors. Notably, CCI (0.90[0.89-0.90]; p<0.0001) demonstrated a negative association with liver cancer-related mortality. (**Fig. S6**) Moderate to severe liver diseases (1.41[1.36-1.47]; p<0.000) and Diabetes mellitus (1.30[1.25-1.34]; p<0.0001) were the two risk factors with highest HR among comorbidities in patients with HCC recurrence. (**Fig. S7**) In the multivariate cox regression analysis, CCI was a robust prognosis factor in cancer-related mortality and all-cause mortality, even after accounting for multiple confounders in multivariate models adjusting for demographics, comorbidities, drugs use, eGFR, and glucose tests. A correlation between glucose test results and the CCI can be observed when interpreting the association between CCI and outcomes (HCC recurrence, liver cancer-related mortality, and non-cancer-related mortality), suggesting that the CCI plays an important role in HCC risk prediction. (**Fig. S8**)

### Subgroup analysis

The descriptive statistics of patients’ outcomes and characteristics by prior cancer status upon new diagnosis of HCC are shown in **Table S19**. The significant effect of CCI on both primary and secondary outcomes remained consistent across all patient subgroups including sex, age at HCC diagnosis, previous multi-drug exposure, and eGRF. (**Fig. S10-S13**).

### Sensitivity analysis

A series of landmark sensitivity analysis were conducted impact of CCI (**Table S20A**), the number of drug classes (**Table S20B**), biomarkers (**Table S20C**), liver tests (**Table S20D**), as well as lipid and glucose test (**Table S20E**) on subsequent outcomes after initial HCC diagnosis, after excluding mortality within 30, 60, 90, 180 days, and 1 and 2 years were conducted for sensitivity analysis. We observed CCI score showed a constant positive association with the risk of HCC recurrence (HR>1, and P<0.05), indicating that the heavier the burden of comorbidities, the higher the risk of recurrence. (**Table S20**). In contrast, the relationship between CCI and liver cancer-related mortality was non-linear (U-shaped), with elevated risk at low CCI scores and attenuated risk at high CCI scores (Table S20). This attenuation likely reflects competing mortality risks; patients with numerous comorbidities (high CCI) are more susceptible to death from other causes, potentially precluding them from living long enough to succumb to liver cancer itself. Regarding the number of previous drug classes, a greater number of prescribed drug classes predicted higher risk of HCC prognosis outcomes, particularly non-cancer-related mortality and all-cause mortality. Patients prescribed no drug use (N=0) exhibited lower risks (HR < 1) across all outcomes. (**Table S20B**) ALBI score and CKD-EPI reveal the strongest and most robust performance as predictors of HCC prognosis outcomes compared to other biomarkers. (**Table S20C**) All liver function tests demonstrated significant associations with liver cancer-related mortality. Among these, alkaline phosphatase exhibited the most robust and significant prognostic value across all examined clinical endpoints. (**Table S20D**) Sensitivity analyses showed that the protective association of LDL with liver cancer mortality became stronger as longer early death exclusion windows were applied: the effect was significant when excluding deaths within 2 years (HR=0.80), weaker at 60 days (HR=0.89), and disappeared at 30 days (HR=0.93). (**Table S20E**)

## Discussion

To our knowledge, this is the first research study exploring comorbidity and multidrug combinations stratified by sex, age and CCI in patients with primary HCC. Over 15,000 patients with primary HCC diagnosis were included in this study, with common comorbidities, multidrug use, and combinations of comorbidities with drug well recorded and identified. We assessed clinical characteristics, biochemical value, prior comorbidities, drug use, and laboratory examinations for primary outcomes (HCC recurrence and liver-related mortality) and secondary outcomes (all-cause, cancer-related, and non-cancer-related mortality). We also explored the association between comorbidity, drug and primary outcomes using networks stratified by sex, age, and CCI.

Our main findings could be organized as follows: (1) liver diseases and metabolic syndrome (DM or HPTN) account for most previous conditions in patients with HCC, regardless of whether they presented as a single comorbidity, a double comorbidity, or a triple comorbidity. (2) Rising drug burden was observed with higher CCI, while treatment intensity, especially the use of cytotoxic and platinum therapies, tends to decline in high CCI group. (3) Sex- and age-difference were presented in comorbidities. (4) patients with previous diabetes and moderate to severe liver diseases were more likely to pass away from liver cancer-related mortality.

This study extends the field in five important ways. In terms of methodology, this is the first study that simultaneously explores comorbidities, multidrug use, and combinations of comorbidities with drug. Many studies explore the association by single direction, such as multidrug pattern ^44,45^, and comorbidity pattern^4,19,46^. All these studies have only assessed individual elements in isolation, either a single comorbidity or a single drug. Whereas we considered multiple comorbidities and drugs, as well as its heterogeneity in sex and CCI. The cohort size of one study^46^ included 6475 patients. However, our study utilized a cohort more than twice the size (>15000). Existing comorbidity research^44,45^ relies on frequency analysis, failing to capture interconnected pathways. In contrast, our study expands this by exploring comorbidity and multidrug combinations using UpSet plot and stratified networks.

Second, our study confirms that critical prognostic significance of pre-existing liver diseases and metabolic syndrome in HCC, evidenced by their high prevalence within multiple comorbidity combinations. High-fructose diet promotes HCC progression via microbiota-derived acetate enhancing O-GlcNAcylation of proliferative drivers, as mechanistically validated in spontaneous and chemically-induced HCC mouse models.^47^ One study about how cirrhosis remodels hepatic architecture suggests that >90% of human HCC arises in the context of underlying cirrhosis or chronic hepatitis, driven by viral infection (HBV/HCV), metabolic dysfunction (NAFLD/NASH), or toxic insults (alcohol/aflatoxin).^48^ Similar findings are prevalent in epidemiological and clinical studies regarding liver diseases^4,19^ and DM^5,23,49^. Beyond these established factors, we implicates colorectal cancer, renal disease, anaemia^20,21^, alcoholism, and GI bleeding^24^ as potential additional prognostic indicators in modulating HCC prognosis. Crucially, our work continues previous research highlighting the importance of comorbidities and drug use on cancer,^17^ consolidating evidence previously fragmented across multiple studies into a single study.

Third, this study reveals a therapeutic paradox in patients with the highest comorbidity burden (CCI ≥11): while polypharmacy prevalence increased, anti-cancer treatment intensity decreased with rising CCI. This pattern reflects critical trade-offs in multimorbidity management—limited life expectancy, heightened drug-interaction risks, and prioritization of palliative goals frequently drive therapeutic de-escalation. It is consistent with the research stating that older patients with HCC are less likely to receive curative treatment and have worse survival than their younger counterparts.^50^ These findings corroborate prior studies^51–53^ but extend it by demonstrating how comorbidity burden interacts with age to constrain therapeutic decision-making, thereby filling in research gaps in drug management for individuals with comorbidity.

Fourth, sex and age difference have been explored in disease prognosis. In females, hormonal signaling (e.g., antiestrogen pathways) and metabolic dysfunction may play a significant role in tumor recurrence. This is consistent with clinical studies suggesting that estrogen receptor signaling may influence the pathogenesis of HCC in women.^54,55^ A retrospective 10-year follow-up cohort study based on 10 million people revealed that the incidence of head neck cancers varied significantly by sex.^56^ Males were more likely to experience moderate to severe liver diseases such as cirrhosis compared to females, which was consistent with previous studies.^57,58^ The same situation also happened on DM, with more males diagnosed with it in the word^59^.

Individuals aged 18 to 65 face a higher risk of developing nasopharyngeal carcinoma, cervical cancer, and ovarian cancer compared to those in the 65 to 85 age group. There is a single peak in the incidence of NPC between the age of 45 and 59, which has been repeatedly observed in studies.^60,61^ Therefore, our study provides novel granularity in investigating the heterogeneity and progression of comorbidities.

Finally, we found that patients with moderate to severe liver disease faced high risk of liver cancer-related mortality. This aligns with cause-specific mortality studies in metabolic dysfunction-associated steatotic liver disease (MASLD), which concluded that MASLD is strongly associated with liver-related and HCC-related mortality.^62^ We address a critical gap^17^ in clinical resources by providing clinicians with an intuitive visualization method for identifying complex patient comorbidity and polypharmacy patterns. This bridges advanced statistical analysis with evidence-based clinical decision-making, ultimately optimizing personalized patient-centred precision medication through data-driven anticancer drug selection.

This study had several limitations. The characteristics we used did not incorporate socio-economic and lifestyle factors, which are essential determinants of health and may influence the development and progression of comorbidities. We acquired common comorbidity combinations but did not infer comorbidity trajectories. These limitations will be addressed in our future study by leveraging alternative methods and other larger comprehensive databases.

This study reveals a complex web of relationships between comorbidities, medications, and outcomes in HCC patients. It highlights the predominant role of liver diseases and metabolic syndrome in prognosis, underscores the impact of disease burden on treatment patterns, and emphasizes sex and age differences in disease characteristics. These results establish the foundation for more individualized management approaches and for further research on the impact of comorbidities.

## Data Availability

Data are not available, as the data custodians (the Hospital Authority and the Department of Health of Hong Kong SAR) have not given permission for sharing due to patient confidentiality and privacy concerns. Local academic institutions, government departments, or nongovernmental organizations may apply for the access to data through the Hospital Authority’s data sharing portal (https://www3.ha.org.hk/data).

## Authors’ contributions

Data analysis: BQY, JDZ

Data review: BQY, JL, OHIC, GT, JDZ

Data acquisition: OHIC, BMYC, GT

Data interpretation: BQY, OHIC, JL, QZG, GT, JL, JDZ

Critical revision of manuscript: BQY, OHIC, JL, QZG, BMYC, GT, JW, KNG, WCWW, TTZ, DKKW, NKM, JDZ

Supervision: DKKW, JDZ Manuscript writing: BQY, JDZ

Manuscript revision: BQY, OHIC, QZG, GT, WCWW, NKM, DKKW, JDZ

## Acknowledgements

None.

## Availability of codes

Codes are available at https://github.com/jadonzhou/ComorbidityNetworks-HCC, which will be disclosed upon formal publication.

## References

1. Cancer Tomorrow, https://gco.iarc.who.int/today/ (accessed 20 June 2025).

2. Llovet JM, Kelley RK, Villanueva A, et al. Hepatocellular carcinoma. Nat Rev Dis Primers 2021; 7: 6.

3. Kim J, Kang W, Sinn DH, et al. Substantial risk of recurrence even after 5 recurrence-free years in early-stage hepatocellular carcinoma patients. Clin Mol Hepatol 2020; 26: 516–528.

4. Mu X-M, Wang W, Jiang Y-Y, et al. Patterns of Comorbidity in Hepatocellular Carcinoma: A Network Perspective. IJERPH 2020; 17: 3108.

5. Kim HY, Lee HA, Radu P, et al. Association of modifiable metabolic risk factors and lifestyle with all-cause mortality in patients with hepatocellular carcinoma. Sci Rep 2024; 14: 15405.

6. Satriano L, Lewinska M, Rodrigues PM, et al. Metabolic rearrangements in primary liver cancers: cause and consequences. Nat Rev Gastroenterol Hepatol 2019; 16: 748–766.

7. Starley BQ, Calcagno CJ, Harrison SA. Nonalcoholic fatty liver disease and hepatocellular carcinoma: A weighty connection. Hepatology 2010; 51: 1820–1832.

8. Caldwell SH, Oelsner DH, Iezzoni JC, et al. Cryptogenic Cirrhosis: Clinical Characterization and Risk Factors for Underlying Disease. Hepatology 1999; 29: 664–669.

9. Tsukuma H, Hiyama T, Tanaka S, et al. Risk Factors for Hepatocellular Carcinoma among Patients with Chronic Liver Disease. N Engl J Med 1993; 328: 1797–1801.

10. Sasaki Y, Yamada T, Tanaka H, et al. Risk of Recurrence in a Long-term Follow-up After Surgery in 417 Patients With Hepatitis B- or Hepatitis C-Related Hepatocellular Carcinoma: Annals of Surgery 2006; 244: 771–780.

11. Hu JX, Thomas CE, Brunak S. Network biology concepts in complex disease comorbidities. Nat Rev Genet 2016; 17: 615–629.

12. Russell CD, Lone NI, Baillie JK. Comorbidities, multimorbidity and COVID-19. Nat Med 2023; 29: 334–343.

13. Lavery JA, Boutros PC, Moskowitz CS, et al. Comorbidity in Midlife and Cancer Outcomes. JAMA Netw Open 2025; 8: e253469.

14. Koroukian SM, Kim U, Rose J. Moving Closer to Personalized Cancer Prevention Strategies by Assessing Comorbidity and Multimorbidity. JAMA Netw Open 2025; 8: e253476.

15. Meyerhardt JA, Catalano PJ, Haller DG, et al. Impact of Diabetes Mellitus on Outcomes in Patients With Colon Cancer. JCO 2003; 21: 433–440.

16. Shinkawa H, Kaibori M, Ueno M, et al. Impact of Diabetes Mellitus and Obesity Comorbidities on Survival Outcomes after Hepatocellular Carcinoma Resection: A Multicenter Retrospective Study. Liver Cancer 2024; 14: 80–91.

17. Sarfati D, Koczwara B, Jackson C. The impact of comorbidity on cancer and its treatment. CA: A Cancer Journal for Clinicians 2016; 66: 337–350.

18. Wang Z, Song S, Zhang L, et al. Hepatic arterial infusion chemotherapy combined with immune checkpoint inhibitors and molecular targeted therapies for advanced infiltrative hepatocellular carcinoma: a single-center experience. Front Immunol 2025; 15: 1474442.

19. Cammarota S, Citarella A, Guida A, et al. The inpatient hospital burden of comorbidities in HCV-infected patients: A population-based study in two Italian regions with high HCV endemicity (The BaCH study). PLoS ONE 2019; 14: e0219396.

20. Meischl T, Balcar L, Park Y-R, et al. Anaemia is independently associated with mortality in patients with hepatocellular carcinoma. ESMO Open 2024; 9: 103593.

21. Rashidi-Alavijeh J, Nuruzade N, Frey A, et al. Implications of anaemia and response to anaemia treatment on outcomes in patients with cirrhosis. JHEP Reports 2023; 5: 100688.

22. Fortuny M, García-Calonge M, Arrabal Ó, et al. Cardiological Adverse Events in Hepatocellular Carcinoma Patients Receiving Immunotherapy: Influence of Comorbidities and Clinical Outcomes. Epub ahead of print 2025. DOI: 10.2139/ssrn.5122563.

23. Glass LM, Hunt CM, Fuchs M, et al. Comorbidities and Nonalcoholic Fatty Liver Disease: The Chicken, the Egg, or Both?

24. Biecker E. Gastrointestinal Bleeding in Cirrhotic Patients with Portal Hypertension. ISRN Hepatology 2013; 2013: 1–20.

25. Zhang Z, He P, Yao H, et al. A network-based study reveals multimorbidity patterns in people with type 2 diabetes. iScience 2023; 26: 107979.

26. Cruz-Ávila HA, Vallejo M, Martínez-García M, et al. Comorbidity Networks in Cardiovascular Diseases. Front Physiol 2020; 11: 1009.

27. Dervić E, Ledebur K, Thurner S, et al. Comorbidity Networks From Population-Wide Health Data: Aggregated Data of 8.9M Hospital Patients (1997–2014). Sci Data 2025; 12: 215.

28. Chou OHI, Ning J, Chan RNC, et al. Lower Risks of New-Onset Hepatocellular Carcinoma in Patients With Type 2 Diabetes Mellitus Treated With SGLT2 Inhibitors Versus DPP4 Inhibitors. Journal of the National Comprehensive Cancer Network 2024; 22: e237118.

29. Chou OHI, Zhou J, V Mui J, et al. Lower risks of new-onset acute pancreatitis and pancreatic cancer in sodium glucose cotransporter 2 (SGLT2) inhibitors compared to dipeptidyl peptidase-4 (DPP4) inhibitors: A propensity score-matched study with competing risk analysis. Diabetes Epidemiology and Management 2023; 9: 100115.

30. Zhou J, Zhang G, Chang C, et al. Metformin versus sulphonylureas for new onset atrial fibrillation and stroke in type 2 diabetes mellitus: a population-based study. Acta Diabetol 2022; 59: 697–709.

31. Charlson ME, Carrozzino D, Guidi J, et al. Charlson Comorbidity Index: A Critical Review of Clinimetric Properties. Psychother Psychosom 2022; 91: 8–35.

32. Johnson PJ, Berhane S, Kagebayashi C, et al. Assessment of Liver Function in Patients With Hepatocellular Carcinoma: A New Evidence-Based Approach—The ALBI Grade. JCO 2015; 33: 550–558.

33. Sterling RK, Lissen E, Clumeck N, et al. Development of a simple noninvasive index to predict significant fibrosis in patients with HIV/HCV coinfection†‡. Hepatology 2006; 43: 1317–1325.

34. Wai C-T, Greenson JK, Fontana RJ, et al. A Simple Noninvasive Index Can Predict Both Significant Fibrosis and Cirrhosis in Patients With Chronic Hepatitis C. Hepatology 2003; 38: 518–526.

35. Amernia B, Moosavy SH, Banookh F, et al. FIB-4, APRI, and AST/ALT ratio compared to FibroScan for the assessment of hepatic fibrosis in patients with non-alcoholic fatty liver disease in Bandar Abbas, Iran. BMC Gastroenterol 2021; 21: 453.

36. Van Der Slikke EC, Star BS, De Jager VD, et al. A high urea-to-creatinine ratio predicts long-term mortality independent of acute kidney injury among patients hospitalized with an infection. Sci Rep 2020; 10: 15649.

37. Lee L, Cheung WY, Atkinson E, et al. Impact of Comorbidity on Chemotherapy Use and Outcomes in Solid Tumors: A Systematic Review. JCO 2011; 29: 106–117.

38. Ruhoy IS, Bolognese PA, Rosenblum JS, et al. Comorbidities and neurosurgical interventions in a cohort with connective tissue disorders. Front Neurol 2025; 15: 1484504.

39. Maji G, Sen S. Ranking influential nodes in complex network using edge weight degree based shell decomposition. Journal of Computational Science 2023; 74: 102179.

40. Shen Y, Wu H, Liu X, et al. Comparison of the 2021 and 2009 chronic kidney disease epidemiology collaboration creatinine equation for estimated glomerular filtration rate in a Chinese population. Clinical Biochemistry 2023; 116: 59–64.

41. Dar SH, Rahim M, Hosseini DK, et al. Impact of liver cirrhosis on ST-elevation myocardial infarction related shock and interventional management, a nationwide analysis. World J Hepatol 2022; 14: 766–777.

42. Inzucchi SE, Lipska KJ, Mayo H, et al. Metformin in Patients With Type 2 Diabetes and Kidney Disease: A Systematic Review. JAMA 2014; 312: 2668.

43. Lanas A, GarcíaLRodríguez LA, Arroyo MT, et al. Risk of upper gastrointestinal ulcer bleeding associated with selective cycloLoxygenaseL2 inhibitors, traditional nonLaspirin nonLsteroidal antiLinflammatory drugs, aspirin and combinations. Gut 2006; 55: 1731–1738.

44. Hui VW-K, Au CL, Lam ASM, et al. Drug–drug interactions between direct-acting antivirals and co-medications: a territory-wide cohort study. Hepatol Int 2022; 16: 1318–1329.

45. Hsu P-Y, Wei Y-J, Lee J-J, et al. Comedications and potential drug-drug interactions with direct-acting antivirals in hepatitis C patients on hemodialysis. Clin Mol Hepatol 2021; 27: 186–196.

46. Shu Z, Liu W, Wu H, et al. Symptom-based network classification identifies distinct clinical subgroups of liver diseases with common molecular pathways. Computer Methods and Programs in Biomedicine 2019; 174: 41–50.

47. Zhou P, Chang W, Gong D, et al. High dietary fructose promotes hepatocellular carcinoma progression by enhancing O-GlcNAcylation via microbiota-derived acetate. Cell Metabolism 2023; 35: 1961–1975.e6.

48. Zhang HE, Henderson JM, Gorrell MD. Animal models for hepatocellular carcinoma. Biochimica et Biophysica Acta (BBA) - Molecular Basis of Disease 2019; 1865: 993–1002.

49. Patmore LA, Katwaroe WK, Van Der Spek D, et al. Association Between the Presence of Metabolic Comorbidities and Liver-Related Events in Patients With Chronic Hepatitis B. Clinical Gastroenterology and Hepatology 2023; 21: 3089–3096.e1.

50. Bardhi O, Daher D, Patel M, et al. Impact of Age on Clinical Outcomes Among Patients with Hepatocellular Carcinoma: A Systematic Review and Meta-Analysis. JHEP Reports 2025; 101368.

51. Kim S, Park J, Kwon J-H, et al. The Charlson Comorbidity Index is associated with risk of 30-day mortality in patients with myocardial injury after non-cardiac surgery. Sci Rep 2021; 11: 18933.

52. Huang Y, Gou R, Diao Y, et al. Charlson comorbidity index helps predict the risk of mortality for patients with type 2 diabetic nephropathy. J Zhejiang Univ Sci B 2014; 15: 58–66.

53. Balducci L, Extermann M. Management of Cancer in the Older Person: A Practical Approach. The Oncologist 2000; 5: 224–237.

54. Sukocheva OA. Estrogen, estrogen receptors, and hepatocellular carcinoma: Are we there yet? World J Gastroenterol 2018; 24: 1–4.

55. Wei Q, Guo P, Mu K, et al. Estrogen suppresses hepatocellular carcinoma cells through ERβ-mediated upregulation of the NLRP3 inflammasome.

56. Park J-O, Nam I-C, Kim C-S, et al. Sex Differences in the Prevalence of Head and Neck Cancers: A 10-Year Follow-Up Study of 10 Million Healthy People. Cancers (Basel) 2022; 14: 2521.

57. Stroffolini T, Esvan R, Biliotti E, et al. Gender differences in chronic HBsAg carriers in Italy: Evidence for the independent role of male sex in severity of liver disease. Journal of Medical Virology 2015; 87: 1899–1903.

58. Bizzaro D, Becchetti C, Trapani S, et al. Influence of sex in alcohol-related liver disease: Pre-clinical and clinical settings. United European Gastroenterology Journal 2023; 11: 218–227.

59. Kautzky-Willer A, Harreiter J, Pacini G. Sex and Gender Differences in Risk, Pathophysiology and Complications of Type 2 Diabetes Mellitus. Endocrine Reviews 2016; 37: 278–316.

60. Wei K-R, Zheng R-S, Zhang S-W, et al. Nasopharyngeal carcinoma incidence and mortality in China, 2013. Chin J Cancer 2017; 36: 90.

61. Miller JA, Le Q-T, Pinsky BA, et al. Cost-Effectiveness of Nasopharyngeal Carcinoma Screening With Epstein-Barr Virus Polymerase Chain Reaction or Serology in High-Incidence Populations Worldwide. JNCI: Journal of the National Cancer Institute 2021; 113: 852–862.

62. Issa G, Shang Y, Strandberg R, et al. Cause-specific mortality in 13,099 patients with metabolic dysfunction-associated steatotic liver disease in Sweden. Journal of Hepatology 2025; S0168827825001564.

