## Supplementary appendix for "Impact of Comorbidity and Drug Exposure Patterns on Recurrence and Mortality Risks in Patients with Primary Hepatocellular Carcinoma: A Chinese Cohort Network Analysis Study"

[Supplementary Table 1. The](#_Toc1758303192) *[International Classification of Diseases, Clinical Modification](#_Toc1758303192)* [(ICD-9-CM and ICD-10-CM) codes for definitions of previous comorbidities and outcomes.	19](#_Toc1758303192)

### Supplementary Methods

#### Indicator for liver function

The albumin-bilirubin (ALBI) score^1^ is calculated using the following linear model:

$$ALBI= {log}_{10} bilirubin (\mu mol/L)\times0.66+albumin (g/L)\times\left( -0.085 \right)$$

#### Indicators for assessing liver fibrosis and injury

The Fibrosis-4 score assist in estimating the amount of scarring in the liver, which is a calculator developed by,^2^

$$FIB-4= \frac{Age\left( years \right)\times AST Level (U/L)}{Platelet Counts ({10}^{9}/L)\times\sqrt{ALT level (U/L)}}$$

Aspartate aminotransferase-to-platelet ratio index (APRI) serves as a measure of liver health in patients with liver disease when aspartate aminotransferase (AST) levels are above the upper limit of normal. It was calculated as following^3^:

$$APRI= \frac{AST level (U/L)}{Platelets Counts ({10}^{9}/L)}\times100$$

The serum aspartate (AST; U/L or IU/L) to alanine aminotransferase (ALT; U/L or IU/L) ratio (AST/ALT ratio) is a clinically validated biomarker for inferring the etiology of liver disease.^4^ For instance, an AST/ALT ratio > 2.0 is strongly suggestive of alcoholic liver disease (ALD), whereas a ratio < 1.0 may indicate chronic viral hepatitis or cholestatic liver diseases (e.g., primary biliary cholangitis).

The alanine aminotransferase to platelet count ratio (ALT/PLT ratio) is a noninvasive biomarker primarily used to assess the risk and severity of liver fibrosis or cirrhosis. Here, ALT activity (expressed in U/L) is normalized to platelet counts (reported as ×10⁹/L or cells/μL, with 1 ×10⁹/L equal to 1,000 cells/μL).

#### Indicator for renal function

Urea-to-creatinine ratio (UCR) can assess kidney function and fluid status by distinguishing kidney problems from other types of dehydration or kidney injury, as the formula below^5^:

$$UCR= \frac{Plasma Urea (mmol/L)}{Plasma Creatinine (\mu mol/L)}$$

1.	Johnson PJ, Berhane S, Kagebayashi C, et al. Assessment of Liver Function in Patients With Hepatocellular Carcinoma: A New Evidence-Based Approach—The ALBI Grade. *JCO* 2015; 33: 550–558.

2.	Sterling RK, Lissen E, Clumeck N, et al. Development of a simple noninvasive index to predict significant fibrosis in patients with HIV/HCV coinfection†‡. *Hepatology* 2006; 43: 1317–1325.

3.	Wai C-T, Greenson JK, Fontana RJ, et al. A Simple Noninvasive Index Can Predict Both Significant Fibrosis and Cirrhosis in Patients With Chronic Hepatitis C. *Hepatology* 2003; 38: 518–526.

4.	Amernia B, Moosavy SH, Banookh F, et al. FIB-4, APRI, and AST/ALT ratio compared to FibroScan for the assessment of hepatic fibrosis in patients with non-alcoholic fatty liver disease in Bandar Abbas, Iran. *BMC Gastroenterol* 2021; 21: 453.

5.	Van Der Slikke EC, Star BS, De Jager VD, et al. A high urea-to-creatinine ratio predicts long-term mortality independent of acute kidney injury among patients hospitalized with an infection. *Sci Rep* 2020; 10: 15649.

### Supplementary Figure 1. Prevalent comorbidity patterns in patients with liver cancer-related mortality.

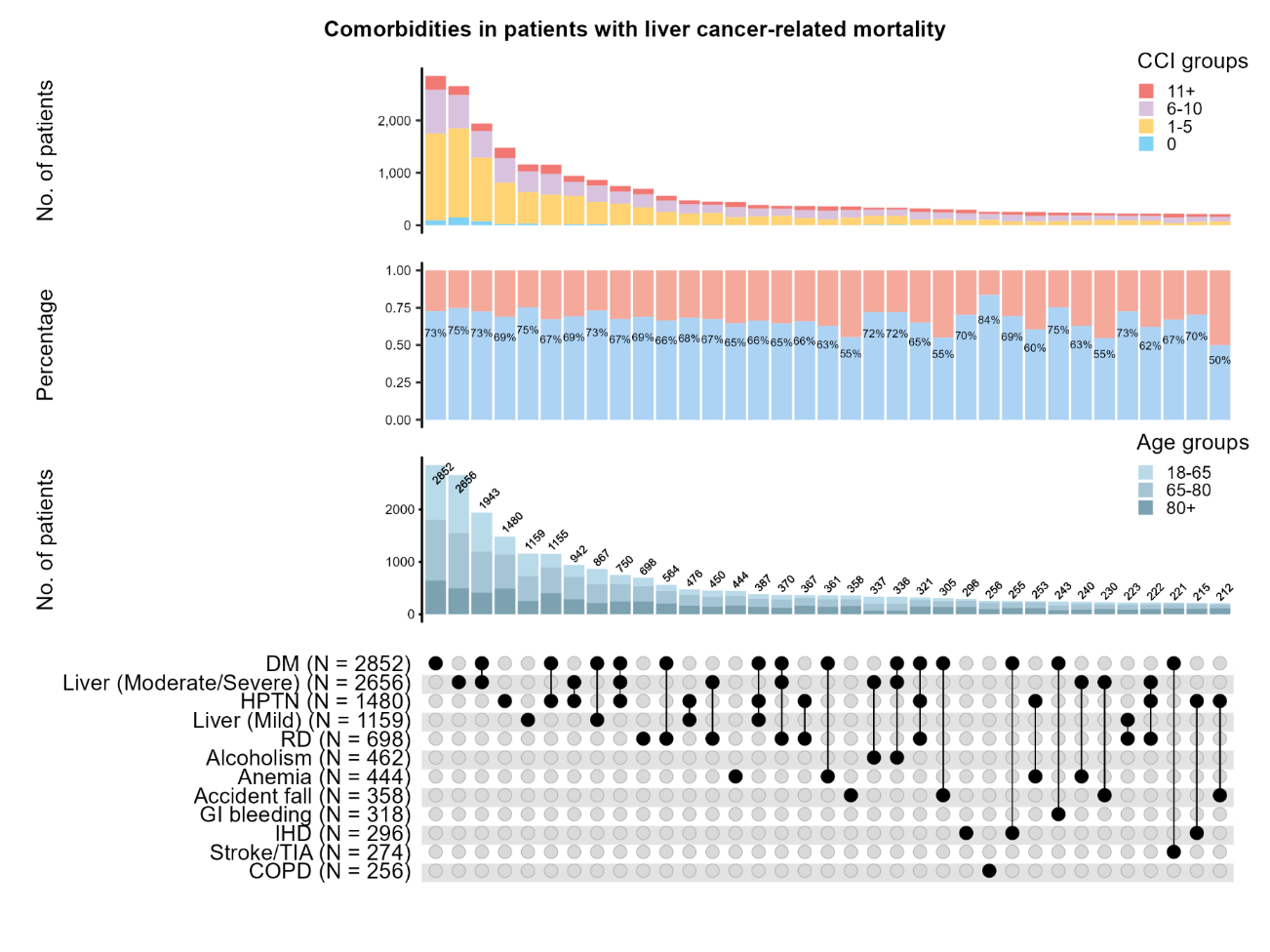

### Supplementary Figure 2. Prevalent multidrug combinations in patients with liver cancer-related mortality.

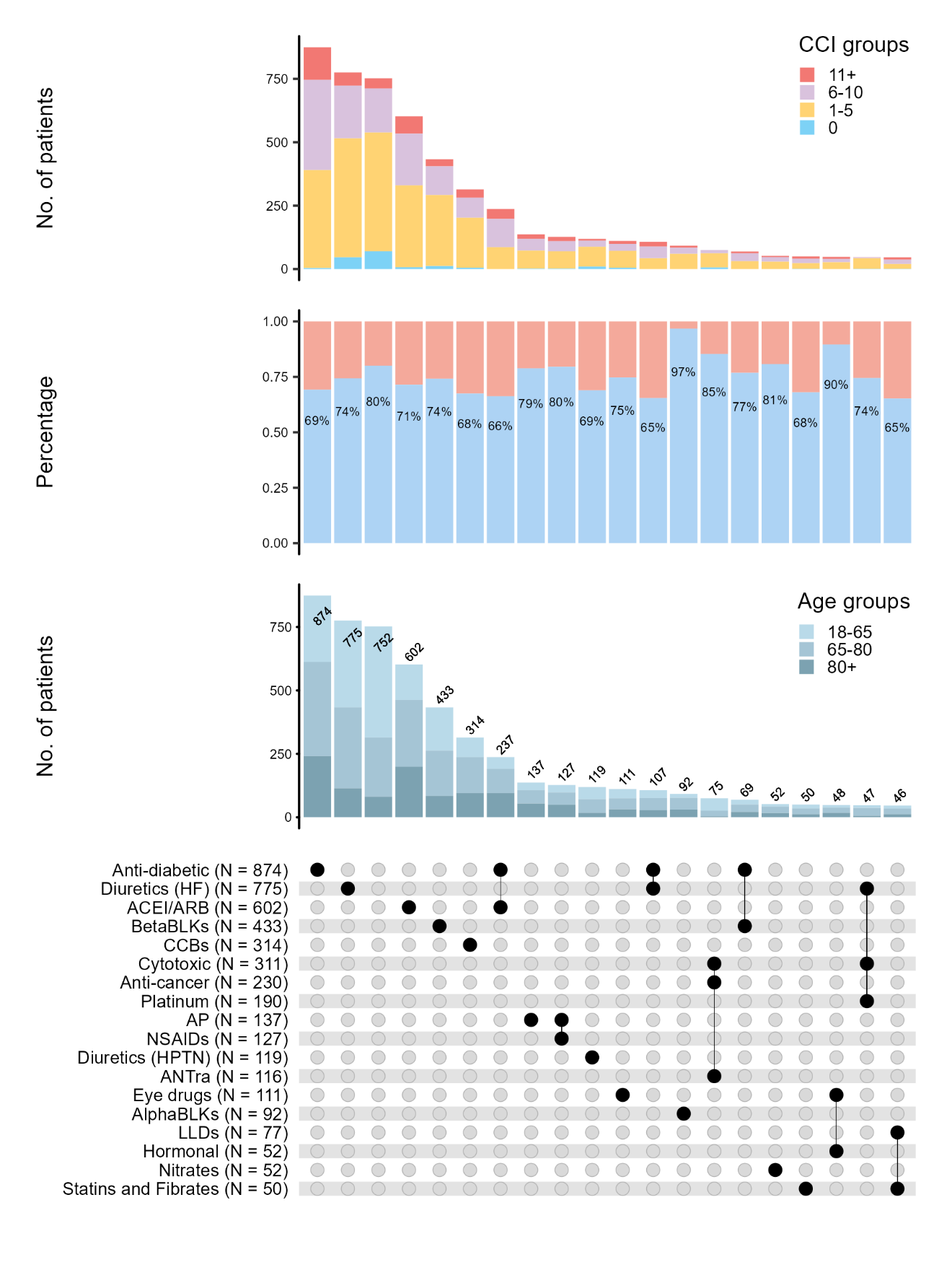

### Supplementary Figure 3. Cumulative hazard curves for primary and secondary outcomes stratified by CCI at HCC diagnosis.

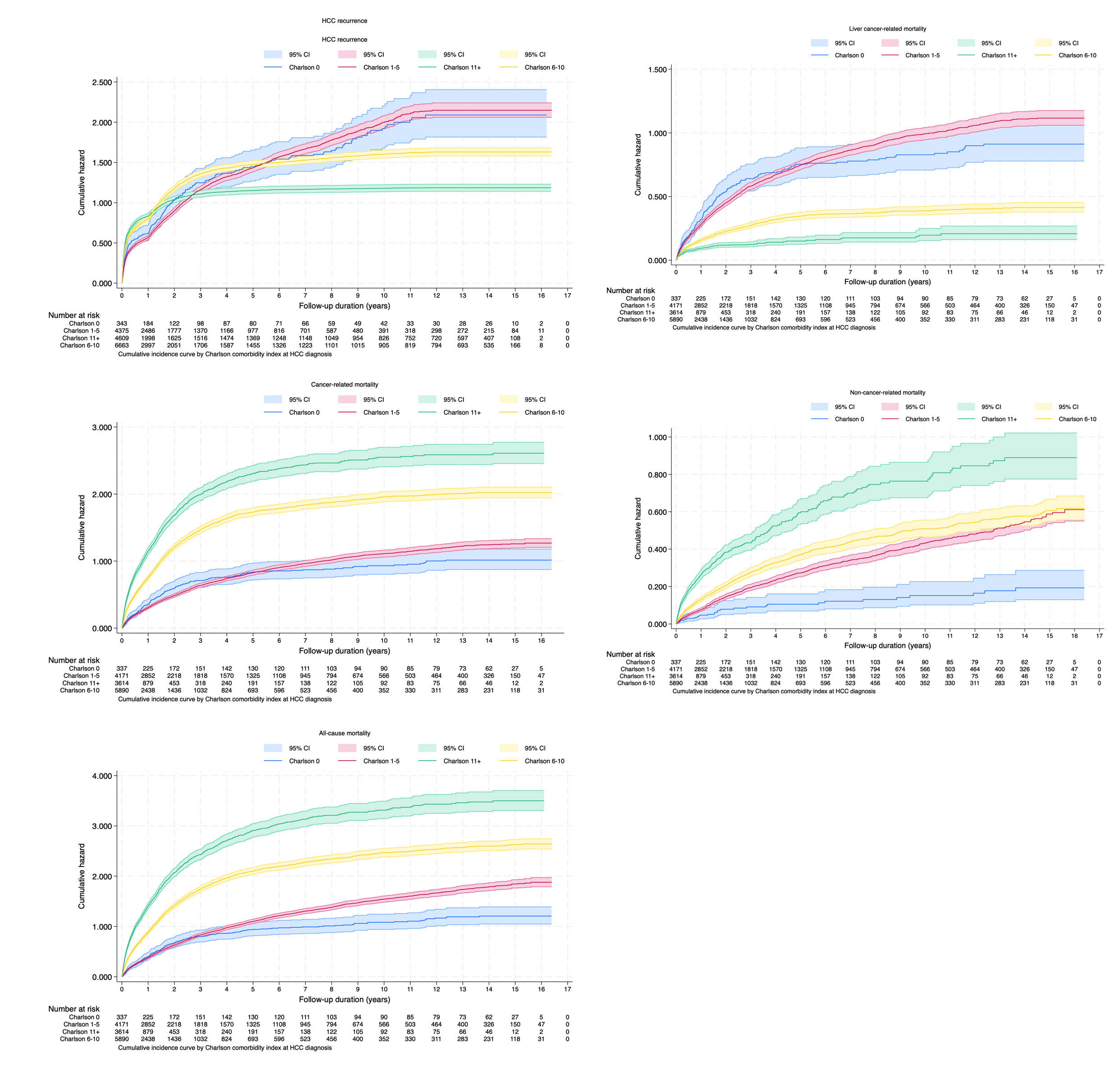

### Supplementary Figure 4. Cumulative hazard curves for primary and secondary outcomes stratified by age at HCC diagnosis.

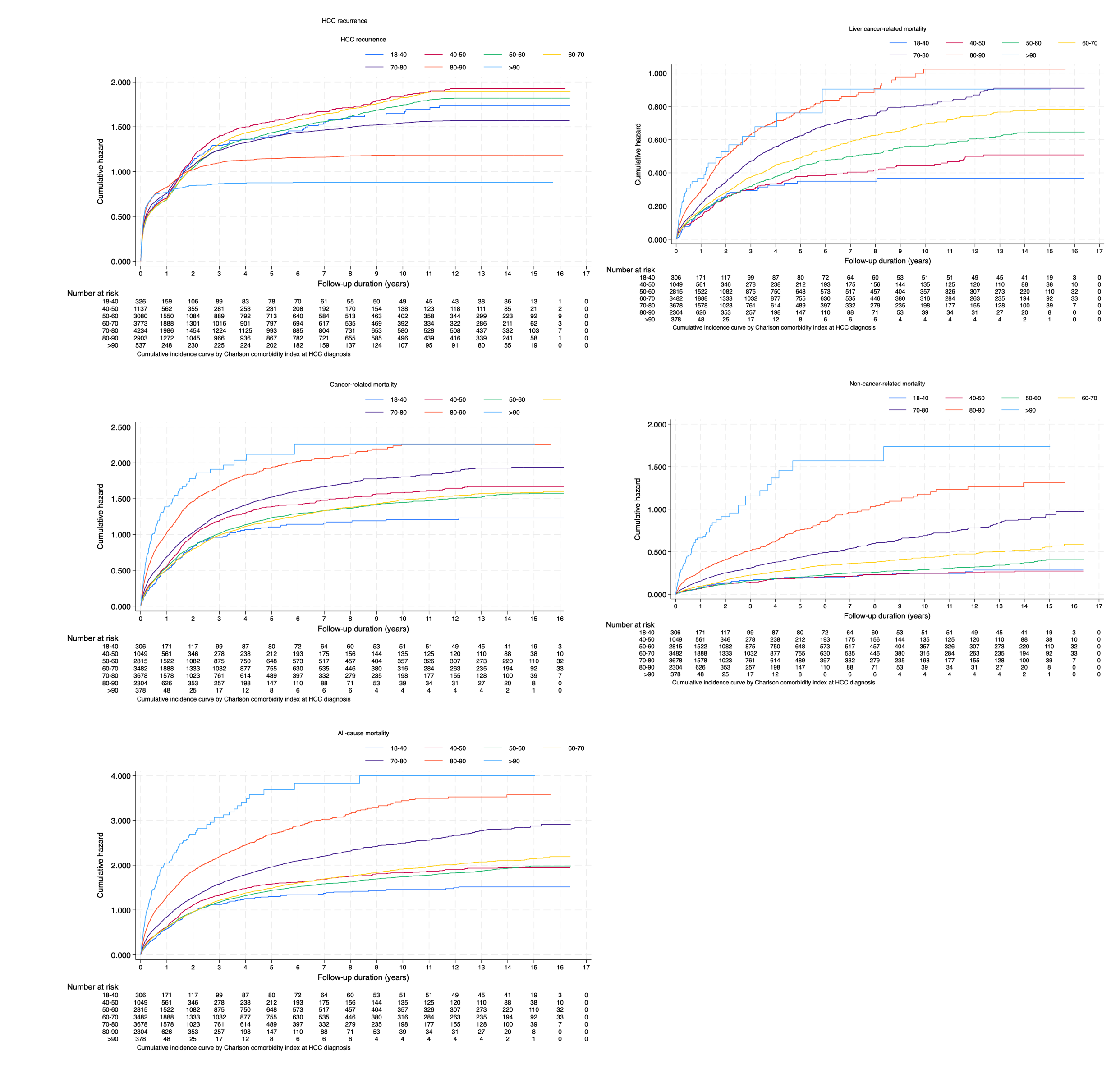

### Supplementary Figure 5. Cumulative incidence curves for primary and secondary outcomes stratified by sex at HCC diagnosis.

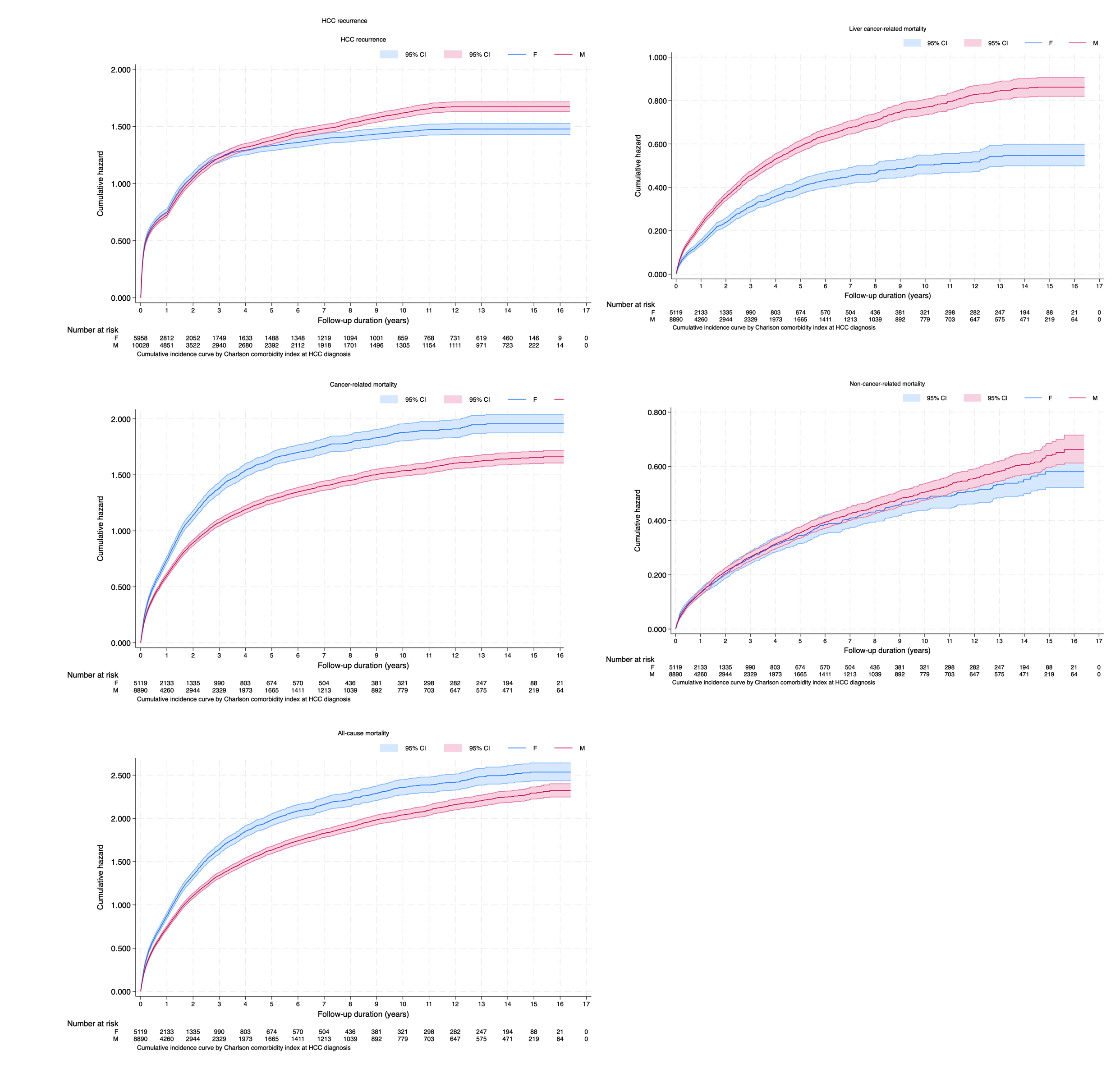

### Supplementary Figure 6. Forest plot for significant risk factors of liver-cancer related mortality using univariate Cox regression models

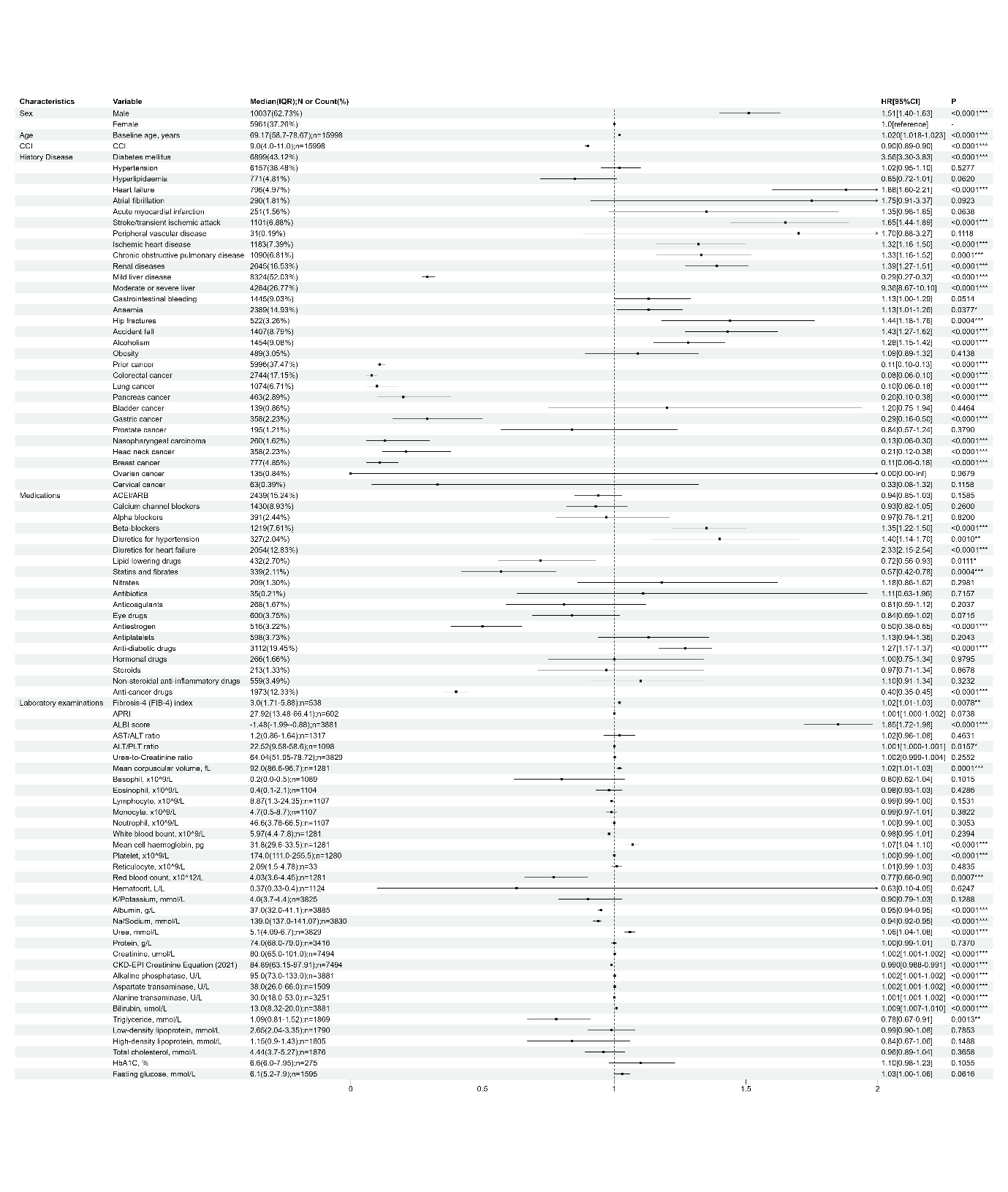

### Supplementary Figure 7. Forest plot for significant risk factors of HCC recurrence using univariate Cox regression models

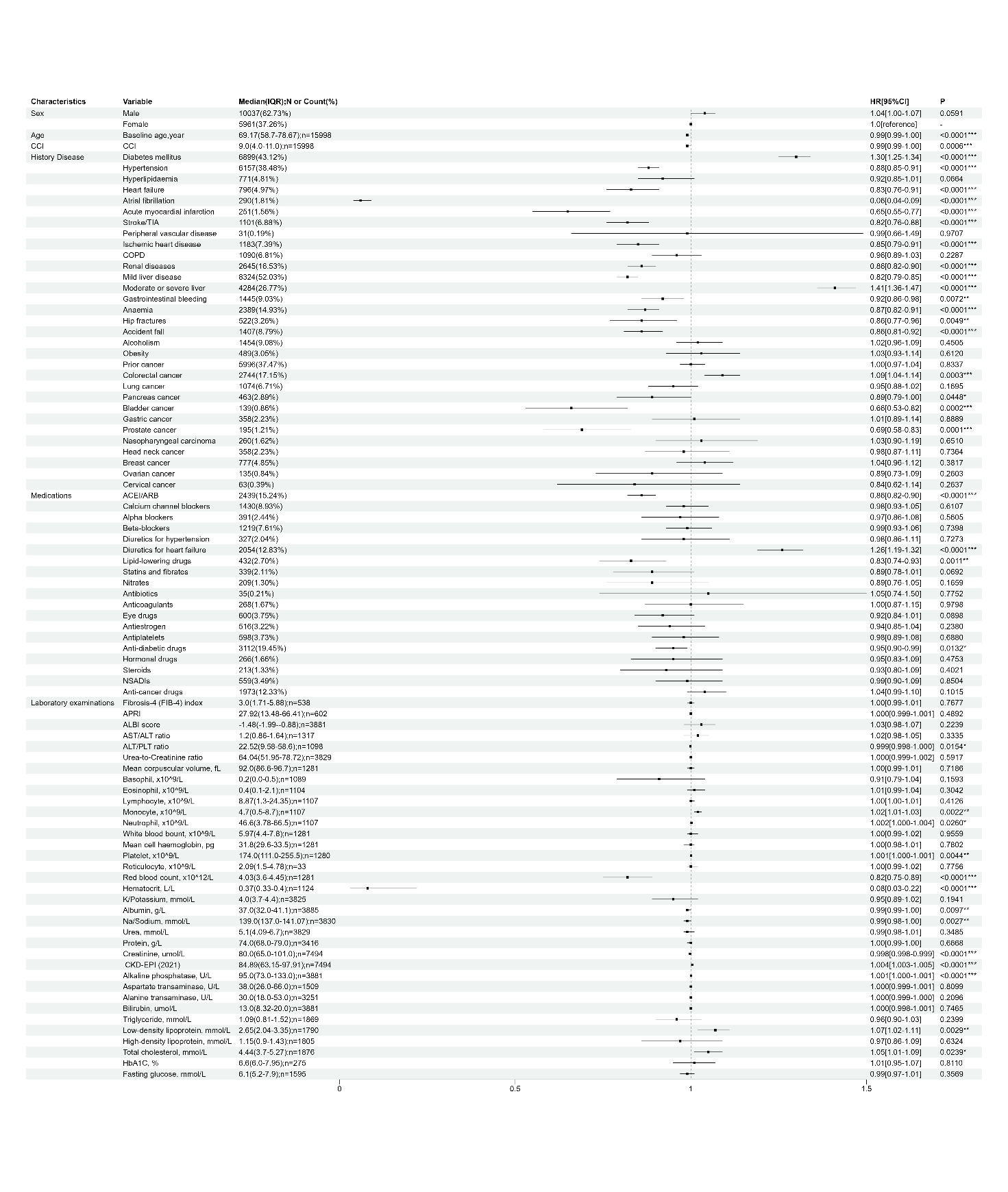

### Supplementary Figure 8. Multivariate Cox regression models for CCI and number of drug classes on primary and secondary outcomes among patients with HCC.

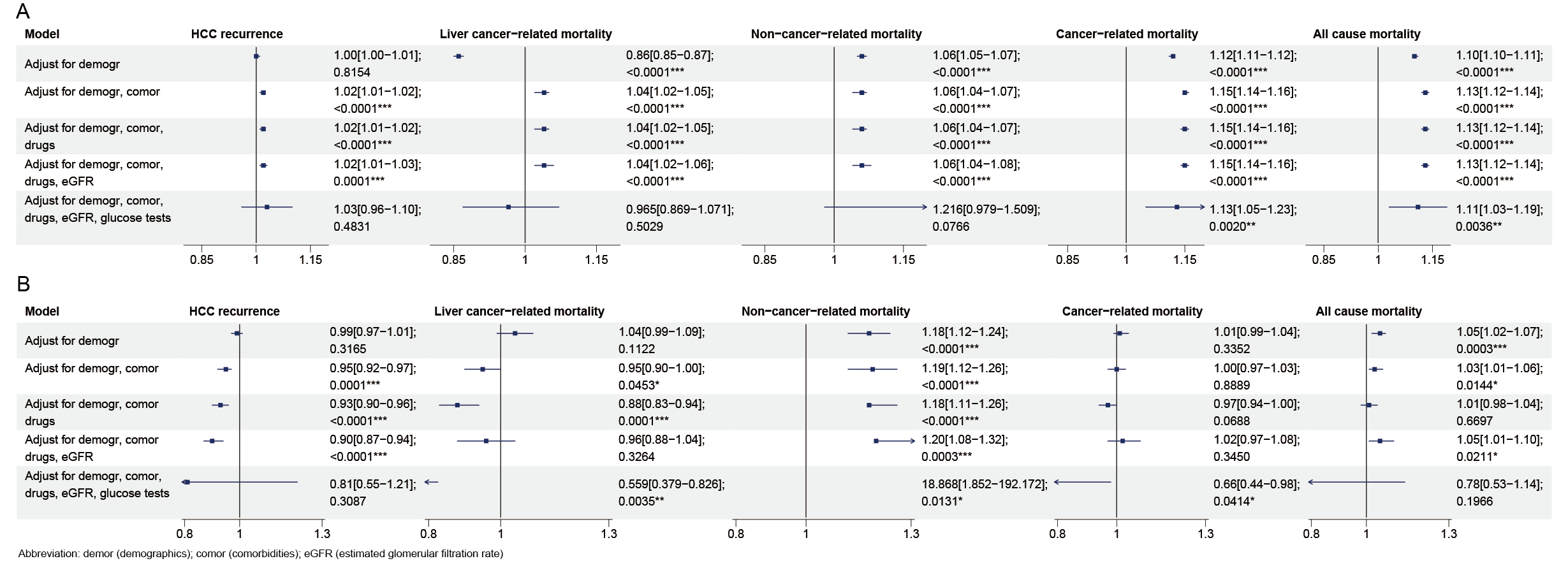

### Supplementary Figure 9. Subgroup analysis for CCI prediction strength on liver cancer-related mortality

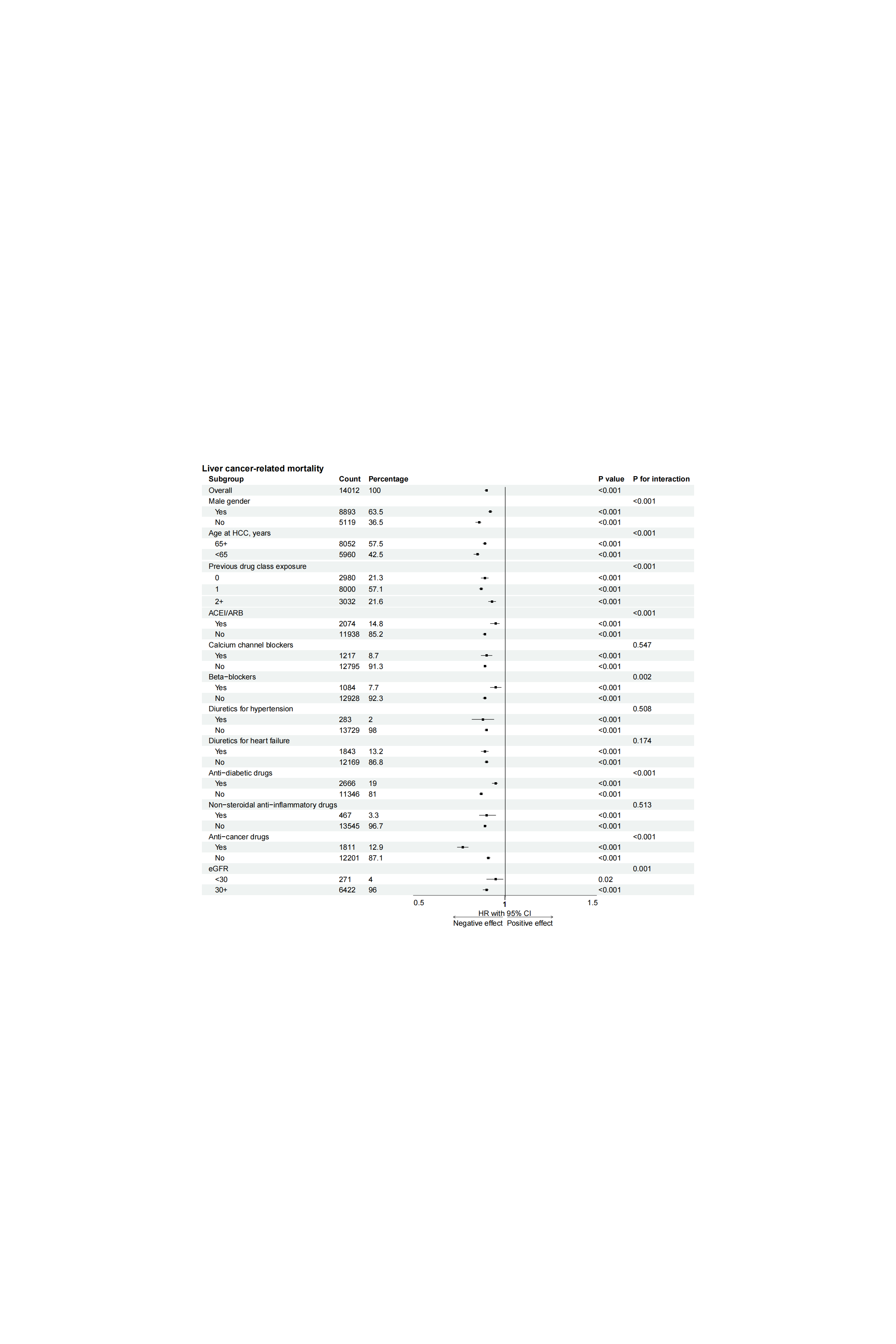

### Supplementary Figure 10. Subgroup analysis for CCI prediction strength on cancer-related mortality

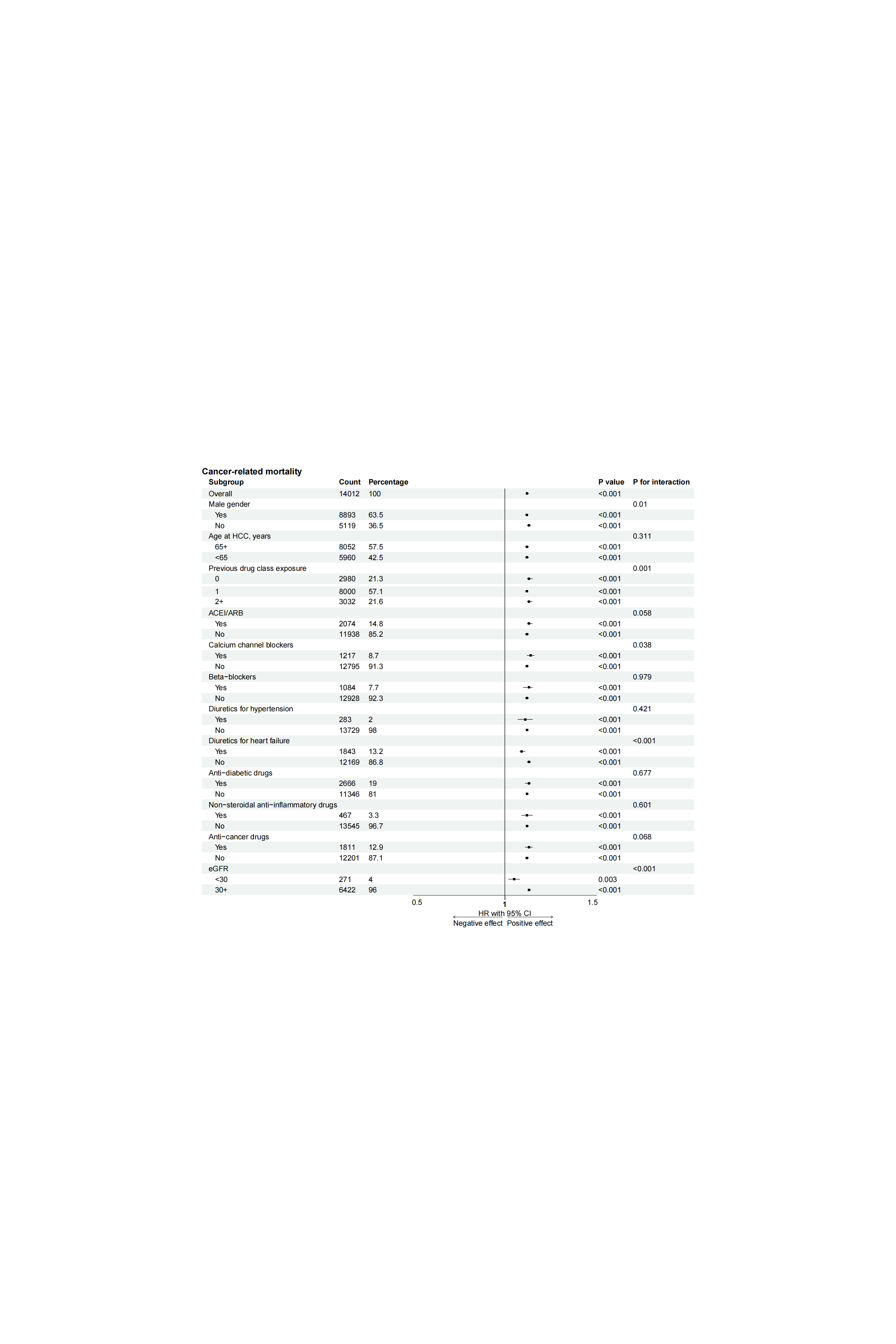

### Supplementary Figure 11. Subgroup analysis for CCI prediction strength on non-cancer-related mortality.

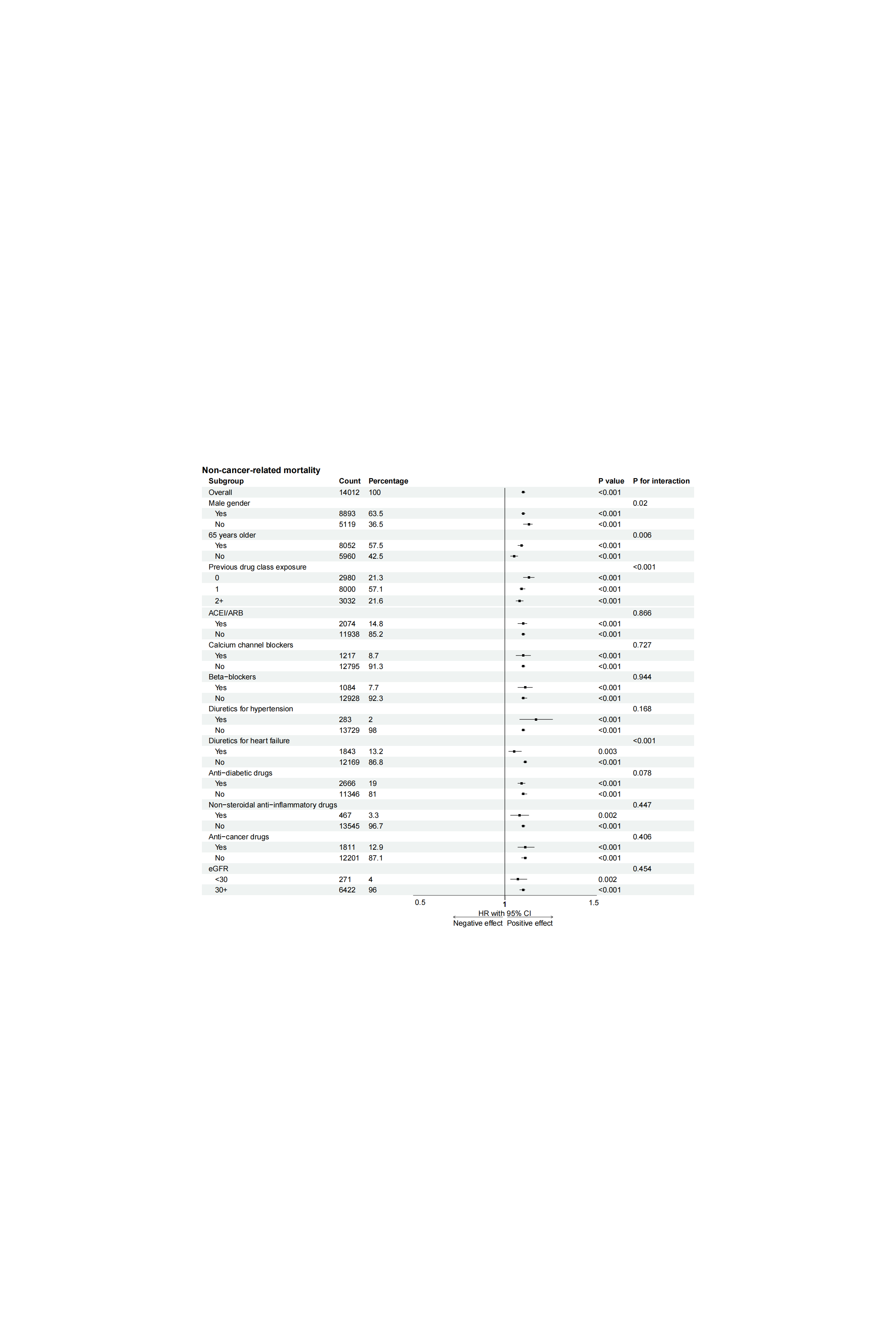

### Supplementary Figure 12. Subgroup analysis for CCI prediction strength on all-cause mortality.

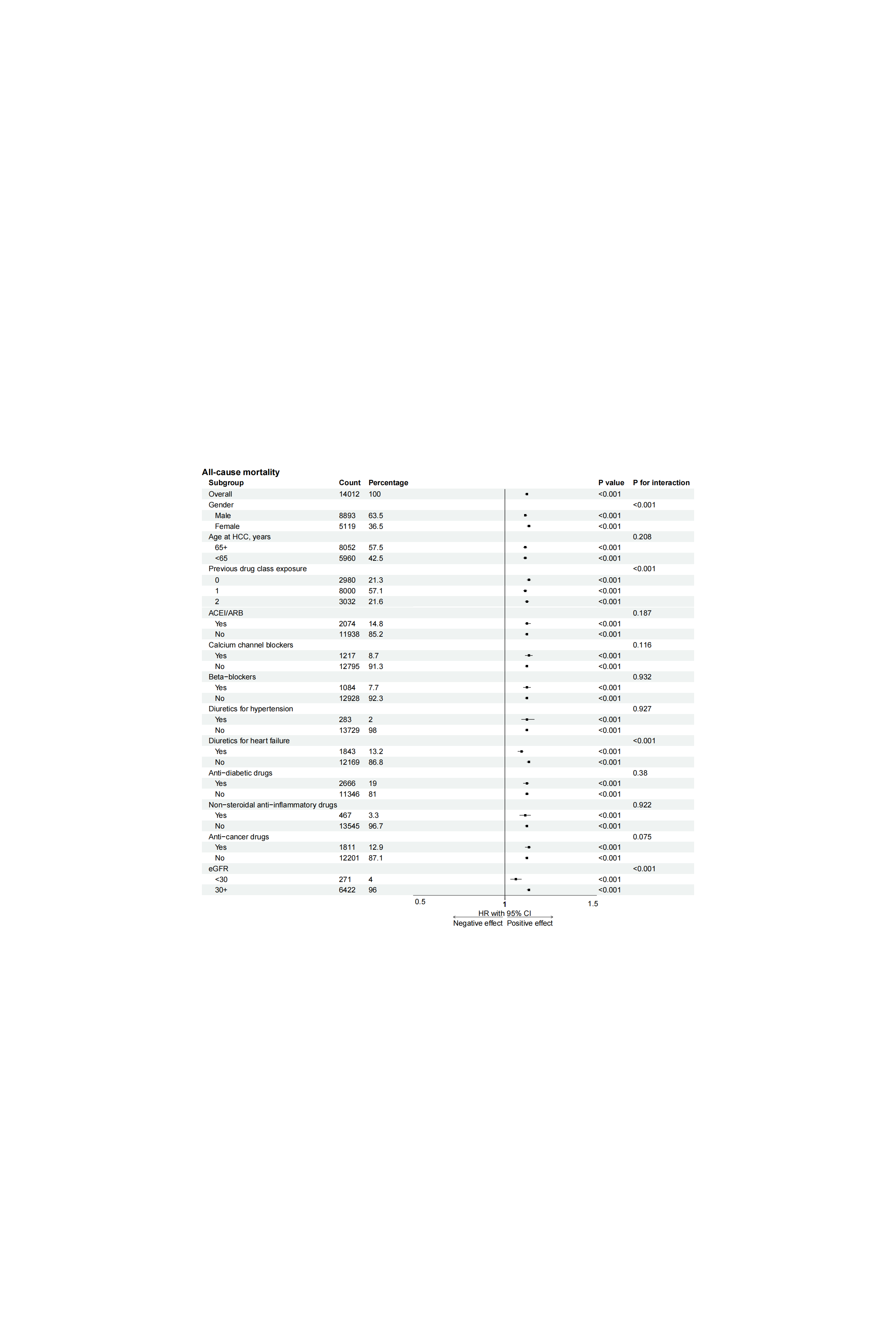

### Supplementary Figure 13. Subgroup analysis for CCI prediction strength on HCC recurrence

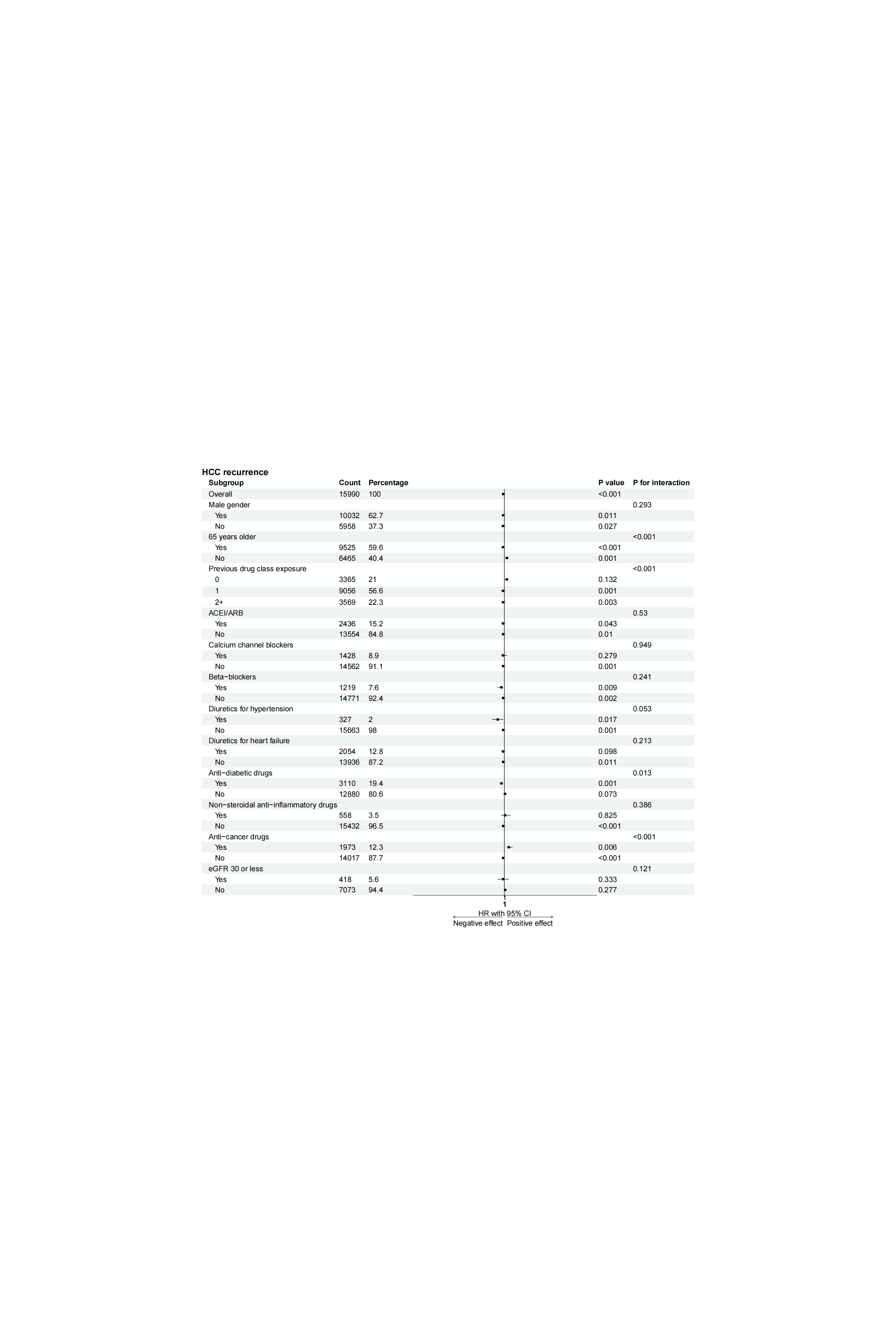

### Supplementary Table 1. The *International Classification of Diseases, Clinical Modification* (ICD-9-CM and ICD-10-CM) codes for definitions of previous comorbidities and outcomes.

**ICD-10 codes**

Cancer-related mortality (ICD-10 codes): C00-C97

Liver cancer-related mortality (ICD-10 codes): C22

**ICD-9codes**

Hepatocellular carcinoma**:** 155.0, 155.2, 197.7

Cancer: 140-239 exclude HCC

Diabetes mellitus 250 250.01 250.02 250.03 250.1 250.11 250.12 250.13 250.2 250.21 250.22 250.23 250.3 250.31 250.32 250.33 250.4 250.41 250.42 250.43 250.5 250.51 250.52 250.53 250.6 250.61 250.62 250.63 250.7 250.71 250.72 250.73 250.8 250.81 250.82 250.83 250.9 250.91 250.92 250.93

Or HbA1c test greater than or equal to 6.5%

Or fasting blood glucose tests greater than or equal to 126 mg/dl

Or on anti-diabetic drug use.

Hypertension 401 401.1 401.9 402 402.01 402.1 402.11 402.9 402.91 403 403.01 403.1 403.11 403.9 403.91 404 404.01 404.02 404.03 404.1 404.11 404.12 404.13 404.9 404.91 404.92 404.93 405 405.01 405.09 405.1 405.11 405.19 405.9 405.91 405.99 437.2 V81.1

Or blood pressure reading is equal to or greater than 130/80 mm-Hg

Or on anti-hypertensive agents use

Hyperlipidaemia 272 272 272.1 272.2 272.3

Or a lipid panel showing elevated total cholesterol (≥240 mg/dL), LDL-C (≥160 mg/dL), triglycerides (≥200 mg/dL), or low HDL-C (<40 mg/dL in men, <50 mg/dL in women).

Or on lipid-lowering drug use

Heart failure 428 428.1 428.2 428.21 428.22 428.23 428.3 428.31 428.32 428.33 428.4 428.41 428.42 428.43 428.9 398.91 402.01 402.11 402.91 404.01 404.03 404.11 404.13 404.91 404.93

Atrial fibrillation 427.31 429.4 427.32

Acute myocardial infarction 410 410.01 410.02 410.1 410.11 410.12 410.2 410.21 410.22 410.3 410.31 410.32 410.4 410.41 410.42 410.5 410.51 410.52 410.6 410.61 410.62 410.7 410.71 410.72 410.8 410.81 410.82 410.9 410.91 410.92

Stroke/transient ischemic attack 430 431 433.91 434.11 434.91 435 435.1 435.2 435.3 435.8 435.9 436 438 438 438.1 438.1 438.11 438.12 438.13 438.14 438.19 438.2 438.2 438.21 438.22 438.3 438.3 438.31 438.32 438.4 438.4 438.41 438.42 438.5 438.5 438.51 438.52 438.53 438.6 438.7 438.8 438.81 438.82 438.83 438.84 438.85 438.89 438.9

Peripheral vascular disease 250.7 443.9 443 443.1 443.2 443.21 443.22 443.23 443.24 443.29 443.8 443.81 443.82 443.89 441 443.9 785.4 V43.4

Ischemic heart disease 410.01 410.02 410.1 410.11 410.12 410.2 410.21 410.22 410.3 410.31 410.32 410.4 410.41 410.42 410.5 410.51 410.52 410.6 410.61 410.62 410.7 410.71 410.72 410.8 410.81 410.82 410.9 410.91 410.92 411 411.1 411.8 411.81 411.89 413 413.1 413.9 414 414.01 414.02 414.03 414.04 414.05 414.06 414.07 414.1 414.11 414.12 414.19 414.2 414.3 414.4 414.8 414.9 410 412

Chronic obstructive pulmonary disease 490 491 492 493 494 495 496 491.1 491.2 491.21 491.22 491.8 491.9 492.8 493.01 493.02 493.1 493.11 493.12 493.2 493.21 493.22 493.8 493.81 493.82 493.9 493.91 493.92 494.1 495.1 495.2 495.3 495.4 495.5 495.6 495.7 495.8 495.9

Renal failure 582 582 582.1 582.2 582.4 582.8 582.81 582.89 582.9 583 583 583.1 583.2 583.4 583.6 583.7 585 585.1 585.2 585.3 585.4 585.5 585.6 585.9 586 588 588 588.1 588.8 588.81 588.89 588.9 582 583 583.1 583.2 583.4 583.6 583.7 585 586 588

Mild liver disease 571 571.1 571.3 571.4 571.4 571.41 571.42 571.49 571.8 571.9 70 70.1 70.2 70.2 70.21 70.22 70.23 70.3 70.3 70.31 70.32 70.33 70.4 70.41 70.42 70.43 70.44 70.49 70.5 70.51 70.52 70.53 70.54 70.59 70.6 70.7 70.7 70.71 70.9 V02.6 V02.60 V02.61 V02.62 V02.69 291 291.1 291.2 291.3 291.4 291.5 291.8 291.81 291.82 291.89 291.9

Moderate or severe liver 456 456.1 456.2 456.2 456.21 572.2 572.3 572.4 572.8 571.2 571.5 571.6

Gastrointestinal bleeding 456 456.2 530.21 530.4 530.7 530.82 531 531.01 531.1 531.11 531.2 531.21 532 532.01 532.1 532.11 532.2 532.21 533 533.01 533.1 533.11 533.2 533.21 531.4 531.41 531.5 531.51 531.6 531.61 532.4 532.41 532.5 532.51 532.6 532.61 533.4 533.41 533.5 533.51 533.6 533.61 535.01 535.11 535.21 535.41 535.51 535.61 535.71 537.83 578 578.9 578 578.1 578.9

Anaemia 280 280.1 280.8 280.9 281 281.1 281.2 281.3 281.4 281.8 281.9 282.2 282.3 282.8 282.9 283 283.1 283.11 283.19 283.2 283.9 284 284.01 284.09 284.1 284.11 284.12 284.19 284.81 284.9 285 285.1 285.2 285.21 285.22 285.29 285.3 285.8 285.9

Hip fractures 805 805 805 805.01 805.02 805.03 805.04 805.05 805.06 805.07 805.08 805.1 805.1 805.11 805.12 805.13 805.14 805.15 805.16 805.17 805.18 805.2 805.3 805.4 805.5 805.6 805.7 805.8 805.9 812 812 812 812.01 812.02 812.03 812.09 812.1 812.1 812.11 812.12 812.13 812.19 812.2 812.2 812.21 812.3 812.3 812.31 812.4 812.4 812.41 812.42 812.43 812.44 812.49 812.5 812.5 812.51 812.52 812.53 812.54 812.59 813 813 813 813.01 813.02 813.03 813.04 813.05 813.06 813.07 813.08 813.1 813.1 813.11 813.12 813.13 813.14 813.15 813.16 813.17 813.18 813.2 813.2 813.21 813.22 813.23 813.3 813.3 813.31 813.32 813.33 813.4 813.4 813.41 813.42 813.43 813.44 813.45 813.46 813.47 813.5 813.5 813.51 813.52 813.53 813.54 813.8 813.8 813.81 813.82 813.83 813.9 813.9 813.91 813.92 813.93 814 814 814 814.01 814.02 814.03 814.04 814.05 814.06 814.07 814.08 814.09 814.1 814.1 814.11 814.12 814.13 814.14 814.15 814.16 814.17 814.18 814.19 820 820 820 820.01 820.02 820.03 820.09 820.1 820.1 820.11 820.12 820.13 820.19 820.2 820.2 820.21 820.22 820.3 820.3 820.31 820.32 820.8 820.9

Accident fall E880 E880.0 E880.1 E880.9 E881 E881.0 E881.1 E882 E883 E883.0 E883.1 E883.2 E883.9 E884 E884.0 E884.1 E884.2 E884.3 E884.4 E884.5 E884.6 E884.9 E885 E885.0 E885.1 E885.2 E885.3 E885.4 E885.9 E886 E886.0 E886.9 E887 E888 E888.0 E888.1 E888.8 E888.9

Alcoholism 291 303 571.2 305 291 291.1 291.2 291.3 291.4 291.5 291.8 291.81 291.82 291.89 291.9 303 303.01 303.02 303.03 303.9 303.91 303.92 303.93

Obesity 278.01 278

Colorectal cancer 153 153.1 153.2 153.3 153.4 153.5 153.6 153.7 153.8 153.9 154 154.1 154.2 154.3 154.8

Lung cancer 162 162.2 162.3 162.4 162.5 162.8 162.9

Pancreas cancer 157.9 157.8 157.1 157.2 157.3 157.4 157 157.9

Bladder cancer 188 188 188.1 188.2 188.3 188.4 188.5 188.6 188.7 188.8 188.9

Gastric cancer 151 151.1 151.2 151.3 151.4 151.5 151.6 151.8 151.9 230.2

Prostate cancer 185

Nasopharyngeal carcinoma 147 147.1 147.2 147.3 147.8 147.9

Head & neck cancer 147 147.1 147.2 147.3 147.8 147.9 140 140.1 140.3 140.4 140.5 140.6 140.8 140.9 141 141.1 141.2 141.3 141.4 141.5 141.6 141.8 141.9 142 142.1 142.2 142.8 142.9 143 143.1 143.8 143.9 144 144.1 144.8 144.9 145 145.1 145.2 145.3 145.4 145.5 145.6 145.8 145.9 146 146.1 146.2 146.3 146.4 146.5 146.6 146.7 146.8 146.9 147 147.1 147.2 147.3 147.8 147.9 148 148.1 148.2 148.3 148.8 148.9 149 149.1 149.8 149.9 160 160.1 160.2 160.3 160.4 160.5 160.8 160.9 161 161.1 161.2 161.3 161.8 161.9 195

Breast cancer 174 174.1 174.2 174.3 174.4 174.5 174.6 174.8 174.9 175 175.9

Ovarian cancer 183 183.2 183.3 183.4 183.5 183.8 183.9

Cervical cancer 180 180.1 180.9

### Supplementary Table 2. Common Medical Abbreviations

| **Diseases** | **Acronyms/Abbreviations** | **Disease category** |
| --- | --- | --- |
| Diabetes mellitus | DM | Metabolic diseases |
| Hypertension | HPTN | Cardiovascular diseases |
| Hyperlipidaemia | HLD | Metabolic diseases |
| Heart failure | HF | Cardiovascular diseases |
| Atrial fibrillation | AFib | Cardiovascular diseases |
| Acute myocardial infarction | AMI | Cardiovascular diseases |
| Stroke/transient ischemic attack | Stroke/TIA | Cardiovascular diseases |
| Peripheral vascular disease | PVD | Cardiovascular diseases |
| Ischemic heart disease | IHD | Cardiovascular diseases |
| Chronic obstructive pulmonary disease | COPD | Respiratory diseases |
| Renal diseases | RD | Renal diseases |
| Mild liver disease | Liver (Mild) | Digestive diseases |
| Moderate or severe liver | Liver (Moderate/Severe) | Digestive diseases |
| Gastrointestinal bleeding | GI bleeding | Digestive diseases |
| Anemia | Anemia | Hematologic diseases |
| Hip fractures | Hip fractures | Musculoskeletal disorders |
| Accident fall | Accident fall | External causes of injury |
| Alcoholism | Alcoholism | Mental and behavioral disorders |
| Obesity | Obesity | Metabolic diseases |
| Colorectal cancer | CRC | Cancer |
| Lung cancer | Lung Cancer | Cancer |
| Pancreas cancer | Pancreas Cancer | Cancer |
| Bladder cancer | Bladder Cancer | Cancer |
| Gastric cancer | Gastric Cancer | Cancer |
| Prostate cancer | Prostate cancer | Cancer |
| Nasopharyngeal carcinoma | NPC | Cancer |
| Head and neck cancer | Head neck cancer | Cancer |
| Breast cancer | Breast cancer | Cancer |
| Ovarian cancer | Ovarian cancer | Cancer |
| Cervical cancer | Cervical cancer | Cancer |
| Hepatocellular carcinoma | HCC | Cancer |

### Supplementary Table 3. Common Drugs Abbreviations

| **Drugs** | **Acronyms/ Abbreviations** | **Drug category** |
| --- | --- | --- |
| ACEI/ARB | ACEI/ARB | Cardiovascular system |
| Calcium channel blockers | CCBs | Cardiovascular system |
| Alpha blockers | AlphaBLKs | Cardiovascular system |
| Beta-blockers | BetaBLKs | Cardiovascular system |
| Diuretics for hypertension | Diuretics (HPTN) | Cardiovascular system |
| Diuretics for heart failure | Diuretics (HF) | Cardiovascular system |
| Lipid-lowering drugs | LLDs | Metabolic drugs |
| Statins and fibrates | Statins and Fibrates | Metabolic drugs |
| Nitrates | Nitrates | Cardiovascular system |
| Antibiotics | AB | Anti-infectives for systemic use |
| Anticoagulants | AC | Blood and blood forming organs |
| Eye drugs | Eye drugs | Sensory organs drugs |
| Antiestrogen | AE | Antineoplastic and immunomodulating agents |
| Antiplatelets | AP | Blood and blood forming organs |
| Anti-diabetic drugs | Anti-diabetic | Alimentary tract and metabolism |
| Hormonal drugs | Hormonal drugs | Hormonal drugs |
| Steroids | Steroids | Hormonal drugs |
| Non-steroidal anti-inflammatory drugs | NSAIDs | Musculo-skeletal system |
| Anti-cancer drugs | Anti-cancer | Antineoplastic and immunomodulating agents |
| Tyrosine kinase inhibitor | TKIs | Antineoplastic and immunomodulating agents |
| Endocrine therapy | ET | Antineoplastic and immunomodulating agents |
| Aromatase inhibitors | AIs | Antineoplastic and immunomodulating agents |
| Endocrine therapy excluding AIs | ET minus AIs | Antineoplastic and immunomodulating agents |
| Cytotoxic therapy | CT | Antineoplastic and immunomodulating agents |
| Androgen deprivation therapy | ADT | Antineoplastic and immunomodulating agents |
| Targeted therapy | TT | Antineoplastic and immunomodulating agents |
| Monoclonal antibodies | mAbs | Antineoplastic and immunomodulating agents |
| Immunomodulatory drugs | IMiDs | Antineoplastic and immunomodulating agents |
| Tamoxifen | TAM | Antineoplastic and immunomodulating agents |
| Anti-HER2 monoclonal antibody | Anti-HER2 mAb | Antineoplastic and immunomodulating agents |
| Anthracycline | ANTra | Antineoplastic and immunomodulating agents |
| Antimetabolite fluoropyrimidines | AF | Antineoplastic and immunomodulating agents |
| Microtubule inhibitor | MTI | Antineoplastic and immunomodulating agents |
| Platinum | Platinum | Antineoplastic and immunomodulating agents |
| GnRH agonist | GnRH agonist | Antineoplastic and immunomodulating agents |
| Immune checkpoint inhibitors | ICIs | Antineoplastic and immunomodulating agents |

### Supplementary Table 4. Most frequent double-comorbidity patterns in patients with HCC diagnosis.

| **Comorbidity pattern** | **All (N=15998)**  **Count (Incidence per 100,000)** | **HCC recurrence (N=12571)**  **Count (Incidence per 100,000)** | **Liver cancer-related mortality (N=3965) Count (Incidence per 100,000)** | **Cancer-related mortality (N=11615) Count (Incidence per 100,000)** | **Non-cancer-related mortality (N=2816) Count (Incidence per 100,000)** | **All-cause mortality (N=14431) Count (Incidence per 100,000)** |
| --- | --- | --- | --- | --- | --- | --- |
| Diabetes mellitus,Moderate or severe liver | 644 (4026) | 642 (5107) | 641 (16166) | 642 (5527) | 1 (36) | 643 (4456) |
| Diabetes mellitus,Mild liver disease | 401 (2507) | 329 (2617) | 237 (5977) | 296 (2548) | 59 (2095) | 355 (2460) |
| Mild liver disease,Colorectal cancer | 283 (1769) | 255 (2028) | 1 (25) | 221 (1903) | 34 (1207) | 255 (1767) |
| Moderate or severe liver,Colorectal cancer | 227 (1419) | 200 (1591) | 3 (76) | 179 (1541) | 27 (959) | 206 (1427) |
| Mild liver disease,Breast cancer | 168 (1050) | 141 (1122) | 1 (25) | 141 (1214) | 21 (746) | 162 (1123) |
| Hypertension,Mild liver disease | 143 (894) | 60 (477) | 37 (933) | 69 (594) | 33 (1172) | 102 (707) |
| Diabetes mellitus,Hypertension | 132 (825) | 116 (923) | 5 (126) | 72 (620) | 37 (1314) | 109 (755) |
| Renal diseases,Mild liver disease | 119 (744) | 89 (708) | 5 (126) | 64 (551) | 37 (1314) | 101 (700) |
| Mild liver disease,Lung cancer | 118 (738) | 92 (732) | 0 | 98 (844) | 15 (533) | 113 (783) |
| Mild liver disease,Alcoholism | 117 (731) | 98 (780) | 15 (378) | 58 (499) | 26 (923) | 84 (582) |
| Hypertension,Moderate or severe liver | 112 (700) | 110 (875) | 109 (2749) | 110 (947) | 2 (71) | 112 (776) |
| Moderate or severe liver,Lung cancer | 96 (600) | 72 (573) | 0 | 85 (732) | 10 (355) | 95 (658) |
| Mild liver disease,Anaemia | 84 (525) | 62 (493) | 6 (151) | 56 (482) | 20 (710) | 76 (527) |
| Chronic obstructive pulmonary disease,Mild liver disease | 58 (363) | 48 (382) | 3 (76) | 21 (181) | 26 (923) | 47 (326) |
| Nasopharyngeal carcinoma,Head neck cancer | 50 (313) | 40 (318) | 0 | 42 (362) | 5 (178) | 47 (326) |
| Mild liver disease,Gastrointestinal bleeding | 46 (288) | 35 (278) | 7 (177) | 22 (189) | 12 (426) | 34 (236) |
| Moderate or severe liver,Breast cancer | 44 (275) | 36 (286) | 1 (25) | 39 (336) | 4 (142) | 43 (298) |
| Renal diseases,Moderate or severe liver | 42 (263) | 40 (318) | 40 (1009) | 41 (353) | 0 | 41 (284) |
| Mild liver disease,Pancreas cancer | 42 (263) | 35 (278) | 0 | 35 (301) | 5 (178) | 40 (277) |
| Hyperlipidaemia,Mild liver disease | 36 (225) | 28 (223) | 0 | 16 (138) | 8 (284) | 24 (166) |
| Mild liver disease,Accident fall | 35 (219) | 26 (207) | 1 (25) | 10 (86) | 16 (568) | 26 (180) |
| Hypertension,Anaemia | 28 (175) | 19 (151) | 3 (76) | 18 (155) | 8 (284) | 26 (180) |
| Hypertension,Colorectal cancer | 26 (163) | 22 (175) | 0 | 21 (181) | 2 (71) | 23 (159) |
| Moderate or severe liver,Gastric cancer | 24 (150) | 23 (183) | 1 (25) | 22 (189) | 2 (71) | 24 (166) |
| Hypertension,Renal diseases | 22 (138) | 13 (103) | 2 (50) | 11 (95) | 9 (320) | 20 (139) |
| Mild liver disease,Gastric cancer | 22 (138) | 20 (159) | 0 | 18 (155) | 3 (107) | 21 (146) |
| Renal diseases,Colorectal cancer | 21 (131) | 19 (151) | 0 | 17 (146) | 1 (36) | 18 (125) |
| Hypertension,Stroke/transient ischemic attack | 20 (125) | 12 (95) | 3 (76) | 11 (95) | 6 (213) | 17 (118) |
| Moderate or severe liver,Gastrointestinal bleeding | 19 (119) | 18 (143) | 15 (378) | 15 (129) | 3 (107) | 18 (125) |
| Gastrointestinal bleeding,Colorectal cancer | 17 (106) | 14 (111) | 0 | 14 (121) | 1 (36) | 15 (104) |
| Mild liver disease,Ovarian cancer | 16 (100) | 10 (80) | 0 | 14 (121) | 1 (36) | 15 (104) |
| Anaemia,Colorectal cancer | 16 (100) | 15 (119) | 0 | 13 (112) | 3 (107) | 16 (111) |
| Ischemic heart disease,Mild liver disease | 15 (94) | 11 (88) | 1 (25) | 5 (43) | 7 (249) | 12 (83) |
| Anaemia,Breast cancer | 14 (88) | 9 (72) | 0 | 12 (103) | 1 (36) | 13 (90) |
| Renal diseases,Breast cancer | 13 (81) | 7 (56) | 0 | 12 (103) | 1 (36) | 13 (90) |
| Diabetes mellitus,Renal diseases | 12 (75) | 6 (48) | 1 (25) | 7 (60) | 5 (178) | 12 (83) |
| Hypertension,Chronic obstructive pulmonary disease | 12 (75) | 8 (64) | 1 (25) | 6 (52) | 5 (178) | 11 (76) |
| Stroke/transient ischemic attack,Mild liver disease | 12 (75) | 12 (95) | 2 (50) | 6 (52) | 5 (178) | 11 (76) |
| Hypertension,Hyperlipidaemia | 11 (69) | 6 (48) | 1 (25) | 8 (69) | 1 (36) | 9 (62) |
| Moderate or severe liver,Pancreas cancer | 11 (69) | 8 (64) | 0 | 10 (86) | 0 | 10 (69) |
| Chronic obstructive pulmonary disease,Moderate or severe liver | 10 (63) | 10 (80) | 10 (252) | 10 (86) | 0 | 10 (69) |
| Renal diseases,Anaemia | 10 (63) | 5 (40) | 0 | 10 (86) | 0 | 10 (69) |
| Mild liver disease,Head neck cancer | 10 (63) | 5 (40) | 0 | 8 (69) | 2 (71) | 10 (69) |
| Gastrointestinal bleeding,Anaemia | 10 (63) | 9 (72) | 0 | 10 (86) | 0 | 10 (69) |
| Anaemia,Ovarian cancer | 10 (63) | 10 (80) | 0 | 9 (77) | 1 (36) | 10 (69) |
| Hypertension,Accident fall | 9 (56) | 4 (32) | 1 (25) | 6 (52) | 3 (107) | 9 (62) |
| Hypertension,Breast cancer | 9 (56) | 8 (64) | 0 | 6 (52) | 1 (36) | 7 (49) |
| Stroke/transient ischemic attack,Moderate or severe liver | 9 (56) | 9 (72) | 9 (227) | 9 (77) | 0 | 9 (62) |
| Moderate or severe liver,Anaemia | 9 (56) | 9 (72) | 8 (202) | 9 (77) | 0 | 9 (62) |
| Diabetes mellitus,Colorectal cancer | 8 (50) | 7 (56) | 0 | 5 (43) | 1 (36) | 6 (42) |

### Supplementary Table 5. Top 20 most common triple-comorbidity patterns in patients with HCC diagnosis.

| **Comorbidity pattern** | **All (N=15998)**  **Count (Incidence per 100,000)** | **HCC recurrence (N=12571)**  **Count (Incidence per 100,000)** | **Liver cancer-related mortality (N=3965) Count (Incidence per 100,000)** | **Cancer-related mortality (N=11615) Count (Incidence per 100,000)** | **Non-cancer-related mortality (N=2816) Count (Incidence per 100,000)** | **All-cause mortality (N=14431) Count (Incidence per 100,000)** |
| --- | --- | --- | --- | --- | --- | --- |
| Diabetes mellitus,Hypertension,Mild liver disease | 503 (3144) | 459 (3651) | 103 (2598) | 270 (2325) | 126 (4474) | 396 (2744) |
| Diabetes mellitus,Hypertension,Moderate or severe liver | 235 (1469) | 234 (1861) | 230 (5801) | 231 (1989) | 2 (71) | 233 (1615) |
| Diabetes mellitus,Moderate or severe liver,Alcoholism | 89 (556) | 89 (708) | 89 (2245) | 89 (766) | 0 | 89 (617) |
| Diabetes mellitus,Renal diseases,Moderate or severe liver | 86 (538) | 85 (676) | 86 (2169) | 86 (740) | 0 | 86 (596) |
| Hypertension,Moderate or severe liver,Colorectal cancer | 63 (394) | 49 (390) | 1 (25) | 49 (422) | 10 (355) | 59 (409) |
| Diabetes mellitus,Mild liver disease,Colorectal cancer | 56 (350) | 48 (382) | 2 (50) | 44 (379) | 8 (284) | 52 (360) |
| Diabetes mellitus,Renal diseases,Mild liver disease | 49 (306) | 38 (302) | 26 (656) | 35 (301) | 10 (355) | 45 (312) |
| Mild liver disease,Nasopharyngeal carcinoma,Head neck cancer | 43 (269) | 37 (294) | 0 | 34 (293) | 6 (213) | 40 (277) |
| Diabetes mellitus,Mild liver disease,Anaemia | 40 (250) | 23 (183) | 23 (580) | 35 (301) | 3 (107) | 38 (263) |
| Mild liver disease,Alcoholism,Obesity | 38 (238) | 30 (239) | 0 | 17 (146) | 12 (426) | 29 (201) |
| Mild liver disease,Anaemia,Colorectal cancer | 37 (231) | 30 (239) | 0 | 28 (241) | 9 (320) | 37 (256) |
| Renal diseases,Moderate or severe liver,Colorectal cancer | 36 (225) | 30 (239) | 0 | 28 (241) | 6 (213) | 34 (236) |
| Diabetes mellitus,Mild liver disease,Alcoholism | 35 (219) | 20 (159) | 19 (479) | 25 (215) | 8 (284) | 33 (229) |
| Renal diseases,Mild liver disease,Colorectal cancer | 34 (213) | 28 (223) | 1 (25) | 31 (267) | 3 (107) | 34 (236) |
| Mild liver disease,Gastrointestinal bleeding,Colorectal cancer | 28 (175) | 26 (207) | 0 | 23 (198) | 4 (142) | 27 (187) |
| Diabetes mellitus,Moderate or severe liver,Accident fall | 25 (156) | 24 (191) | 25 (631) | 25 (215) | 0 | 25 (173) |
| Diabetes mellitus,Chronic obstructive pulmonary disease,Moderate or severe liver | 24 (150) | 24 (191) | 23 (580) | 23 (198) | 1 (36) | 24 (166) |
| Diabetes mellitus,Mild liver disease,Gastrointestinal bleeding | 24 (150) | 12 (95) | 20 (504) | 22 (189) | 1 (36) | 23 (159) |
| Hypertension,Moderate or severe liver,Lung cancer | 23 (144) | 16 (127) | 0 | 20 (172) | 2 (71) | 22 (152) |
| Mild liver disease,Alcoholism,Colorectal cancer | 22 (138) | 19 (151) | 0 | 16 (138) | 4 (142) | 20 (139) |
| Moderate or severe liver,Gastrointestinal bleeding,Colorectal cancer | 22 (138) | 19 (151) | 0 | 20 (172) | 1 (36) | 21 (146) |

### Supplementary Table 6. Top 20 most common quadruple-comorbidity patterns in patients with HCC diagnosis.

| **Comorbidity pattern** | **All (N=15998)**  **Count (Incidence per 100,000)** | **HCC recurrence (N=12571)**  **Count (Incidence per 100,000)** | **Liver cancer-related mortality (N=3965) Count (Incidence per 100,000)** | **Cancer-related mortality (N=11615) Count (Incidence per 100,000)** | **Non-cancer-related mortality (N=2816) Count (Incidence per 100,000)** | **All-cause mortality (N=14431) Count (Incidence per 100,000)** |
| --- | --- | --- | --- | --- | --- | --- |
| Diabetes mellitus,Hypertension,Mild liver disease,Colorectal cancer | 162 (1013) | 155 (1233) | 2 (50) | 119 (1025) | 28 (994) | 147 (1019) |
| Diabetes mellitus,Hypertension,Renal diseases,Mild liver disease | 108 (675) | 92 (732) | 26 (656) | 48 (413) | 48 (1705) | 96 (665) |
| Diabetes mellitus,Hypertension,Moderate or severe liver,Colorectal cancer | 67 (419) | 63 (501) | 5 (126) | 49 (422) | 10 (355) | 59 (409) |
| Diabetes mellitus,Hypertension,Mild liver disease,Anaemia | 61 (381) | 52 (414) | 13 (328) | 42 (362) | 14 (497) | 56 (388) |
| Diabetes mellitus,Hypertension,Renal diseases,Moderate or severe liver | 56 (350) | 56 (445) | 55 (1387) | 55 (474) | 1 (36) | 56 (388) |
| Diabetes mellitus,Hypertension,Mild liver disease,Lung cancer | 53 (331) | 49 (390) | 0 | 44 (379) | 7 (249) | 51 (353) |
| Diabetes mellitus,Hypertension,Mild liver disease,Alcoholism | 52 (325) | 51 (406) | 3 (76) | 18 (155) | 16 (568) | 34 (236) |
| Diabetes mellitus,Hypertension,Chronic obstructive pulmonary disease,Mild liver disease | 43 (269) | 36 (286) | 9 (227) | 22 (189) | 19 (675) | 41 (284) |
| Diabetes mellitus,Hypertension,Mild liver disease,Accident fall | 39 (244) | 36 (286) | 6 (151) | 16 (138) | 15 (533) | 31 (215) |
| Diabetes mellitus,Hypertension,Mild liver disease,Gastrointestinal bleeding | 38 (238) | 34 (270) | 9 (227) | 24 (207) | 11 (391) | 35 (243) |
| Diabetes mellitus,Hypertension,Stroke/transient ischemic attack,Mild liver disease | 35 (219) | 27 (215) | 11 (277) | 25 (215) | 8 (284) | 33 (229) |
| Diabetes mellitus,Hypertension,Hyperlipidaemia,Mild liver disease | 32 (200) | 28 (223) | 8 (202) | 20 (172) | 11 (391) | 31 (215) |
| Diabetes mellitus,Hypertension,Moderate or severe liver,Alcoholism | 32 (200) | 32 (255) | 32 (807) | 32 (276) | 0 | 32 (222) |
| Diabetes mellitus,Hypertension,Ischemic heart disease,Mild liver disease | 31 (194) | 25 (199) | 3 (76) | 13 (112) | 11 (391) | 24 (166) |
| Diabetes mellitus,Hypertension,Mild liver disease,Pancreas cancer | 30 (188) | 27 (215) | 1 (25) | 26 (224) | 2 (71) | 28 (194) |
| Diabetes mellitus,Moderate or severe liver,Alcoholism,Obesity | 25 (156) | 25 (199) | 25 (631) | 25 (215) | 0 | 25 (173) |
| Diabetes mellitus,Hypertension,Mild liver disease,Breast cancer | 24 (150) | 24 (191) | 0 | 22 (189) | 1 (36) | 23 (159) |
| Diabetes mellitus,Hypertension,Moderate or severe liver,Accident fall | 22 (138) | 22 (175) | 22 (555) | 22 (189) | 0 | 22 (152) |
| Diabetes mellitus,Hypertension,Ischemic heart disease,Moderate or severe liver | 17 (106) | 17 (135) | 17 (429) | 17 (146) | 0 | 17 (118) |
| Mild liver disease,Gastrointestinal bleeding,Anaemia,Colorectal cancer | 15 (94) | 10 (80) | 0 | 12 (103) | 0 | 12 (83) |

### Supplementary Table 7. Top 20 most common quintuple-comorbidity patterns in patients with HCC diagnosis.

| **Comorbidity pattern** | **All (N=15998)**  **Count (Incidence per 100,000)** | **HCC recurrence (N=12571)**  **Count (Incidence per 100,000)** | **Liver cancer-related mortality (N=3965) Count (Incidence per 100,000)** | **Cancer-related mortality (N=11615) Count (Incidence per 100,000)** | **Non-cancer-related mortality (N=2816) Count (Incidence per 100,000)** | **All-cause mortality (N=14431) Count (Incidence per 100,000)** |
| --- | --- | --- | --- | --- | --- | --- |
| Diabetes mellitus,Hypertension,Renal diseases,Mild liver disease,Colorectal cancer | 33 (206) | 29 (231) | 1 (25) | 20 (172) | 12 (426) | 32 (222) |
| Diabetes mellitus,Hypertension,Mild liver disease,Anaemia,Colorectal cancer | 32 (200) | 31 (247) | 0 | 22 (189) | 5 (178) | 27 (187) |
| Diabetes mellitus,Hypertension,Renal diseases,Mild liver disease,Anaemia | 25 (156) | 19 (151) | 3 (76) | 12 (103) | 11 (391) | 23 (159) |
| Diabetes mellitus,Hypertension,Mild liver disease,Alcoholism,Obesity | 25 (156) | 20 (159) | 1 (25) | 12 (103) | 8 (284) | 20 (139) |
| Diabetes mellitus,Hypertension,Mild liver disease,Hip fractures,Accident fall | 19 (119) | 17 (135) | 5 (126) | 8 (69) | 7 (249) | 15 (104) |
| Diabetes mellitus,Hypertension,Mild liver disease,Gastrointestinal bleeding,Colorectal cancer | 17 (106) | 17 (135) | 0 | 12 (103) | 2 (71) | 14 (97) |
| Diabetes mellitus,Hypertension,Stroke/transient ischemic attack,Renal diseases,Mild liver disease | 13 (81) | 12 (95) | 0 | 2 (17) | 10 (355) | 12 (83) |
| Diabetes mellitus,Hypertension,Mild liver disease,Anaemia,Alcoholism | 13 (81) | 11 (88) | 1 (25) | 6 (52) | 4 (142) | 10 (69) |
| Diabetes mellitus,Hypertension,Hyperlipidaemia,Renal diseases,Mild liver disease | 12 (75) | 10 (80) | 2 (50) | 8 (69) | 3 (107) | 11 (76) |
| Diabetes mellitus,Hypertension,Ischemic heart disease,Renal diseases,Mild liver disease | 12 (75) | 8 (64) | 3 (76) | 5 (43) | 6 (213) | 11 (76) |
| Diabetes mellitus,Hypertension,Hyperlipidaemia,Mild liver disease,Colorectal cancer | 11 (69) | 11 (88) | 0 | 8 (69) | 3 (107) | 11 (76) |
| Diabetes mellitus,Hypertension,Chronic obstructive pulmonary disease,Renal diseases,Mild liver disease | 11 (69) | 9 (72) | 1 (25) | 6 (52) | 4 (142) | 10 (69) |
| Diabetes mellitus,Hypertension,Renal diseases,Mild liver disease,Gastrointestinal bleeding | 11 (69) | 7 (56) | 4 (101) | 7 (60) | 4 (142) | 11 (76) |
| Diabetes mellitus,Hypertension,Renal diseases,Mild liver disease,Accident fall | 11 (69) | 9 (72) | 3 (76) | 8 (69) | 3 (107) | 11 (76) |
| Diabetes mellitus,Hypertension,Renal diseases,Mild liver disease,Lung cancer | 11 (69) | 9 (72) | 1 (25) | 10 (86) | 1 (36) | 11 (76) |
| Diabetes mellitus,Hypertension,Ischemic heart disease,Mild liver disease,Colorectal cancer | 10 (63) | 10 (80) | 1 (25) | 9 (77) | 1 (36) | 10 (69) |
| Diabetes mellitus,Hypertension,Chronic obstructive pulmonary disease,Mild liver disease,Colorectal cancer | 10 (63) | 10 (80) | 1 (25) | 8 (69) | 1 (36) | 9 (62) |
| Diabetes mellitus,Hypertension,Renal diseases,Mild liver disease,Alcoholism | 10 (63) | 9 (72) | 2 (50) | 4 (34) | 6 (213) | 10 (69) |
| Diabetes mellitus,Hypertension,Mild liver disease,Gastrointestinal bleeding,Anaemia | 10 (63) | 9 (72) | 2 (50) | 9 (77) | 1 (36) | 10 (69) |
| Diabetes mellitus,Hypertension,Mild liver disease,Alcoholism,Colorectal cancer | 10 (63) | 9 (72) | 1 (25) | 9 (77) | 1 (36) | 10 (69) |
| Diabetes mellitus,Hypertension,Moderate or severe liver,Anaemia,Colorectal cancer | 10 (63) | 10 (80) | 1 (25) | 8 (69) | 1 (36) | 9 (62) |

### Supplementary Table 8. Top 15 most common double (co-exposure) drug patterns in patients with HCC diagnosis.

Abbreviations: ACEI (angiotensin-converting enzyme inhibitors); ARB (Angiotensin Receptor Blockers).

| **Drug pattern** | **All (N=15998)**  **Count (Incidence per 100,000)** | **HCC recurrence (N=12571)**  **Count (Incidence per 100,000)** | **Liver cancer-related mortality (N=3965) Count (Incidence per 100,000)** | **Cancer-related mortality (N=11615) Count (Incidence per 100,000)** | **Non-cancer-related mortality (N=2816) Count (Incidence per 100,000)** | **All-cause mortality (N=14431) Count (Incidence per 100,000)** |
| --- | --- | --- | --- | --- | --- | --- |
| ACEI/ARB,Anti-diabetic drugs | 785 (4907) | 576 (4582) | 223 (5624) | 522 (4494) | 194 (6889) | 716 (4962) |
| Antiplatelets,Non-steroidal anti-inflammatory drugs | 439 (2744) | 335 (2665) | 103 (2598) | 303 (2609) | 101 (3587) | 404 (2800) |
| Antiestrogen,Anti-cancer drugs | 295 (1844) | 233 (1853) | 8 (202) | 253 (2178) | 29 (1030) | 282 (1954) |
| Diuretics for heart failure,Anti-diabetic drugs | 248 (1550) | 196 (1559) | 107 (2699) | 190 (1636) | 54 (1918) | 244 (1691) |
| Calcium channel blockers,Anti-diabetic drugs | 226 (1413) | 174 (1384) | 43 (1084) | 161 (1386) | 53 (1882) | 214 (1483) |
| Lipid-lowering drugs,Statins and fibrates | 198 (1238) | 139 (1106) | 25 (631) | 136 (1171) | 36 (1278) | 172 (1192) |
| Beta-blockers,Anti-diabetic drugs | 185 (1156) | 148 (1177) | 68 (1715) | 128 (1102) | 47 (1669) | 175 (1213) |
| Anti-diabetic drugs,Anti-cancer drugs | 129 (806) | 111 (883) | 14 (353) | 82 (706) | 31 (1101) | 113 (783) |
| Nitrates,Anti-diabetic drugs | 46 (288) | 28 (223) | 16 (404) | 35 (301) | 9 (320) | 44 (305) |
| Alpha blockers,Anti-diabetic drugs | 42 (263) | 31 (247) | 8 (202) | 34 (293) | 8 (284) | 42 (291) |
| Eye drugs,Hormonal drugs | 42 (263) | 32 (255) | 7 (177) | 36 (310) | 5 (178) | 41 (284) |
| Anticoagulants,Anti-diabetic drugs | 36 (225) | 24 (191) | 5 (126) | 22 (189) | 13 (462) | 35 (243) |
| Diuretics for hypertension,Anti-diabetic drugs | 28 (175) | 19 (151) | 10 (252) | 22 (189) | 5 (178) | 27 (187) |
| ACEI/ARB,Diuretics for hypertension | 19 (119) | 17 (135) | 6 (151) | 14 (121) | 4 (142) | 18 (125) |
| Lipid-lowering drugs,Anti-diabetic drugs | 18 (113) | 17 (135) | 4 (101) | 12 (103) | 2 (71) | 14 (97) |

### Supplementary Table 9. Top 15 most common triple (co-exposure) drug patterns in patients with HCC diagnosis.

Abbreviations: ACEI (angiotensin-converting enzyme inhibitors); ARB (Angiotensin Receptor Blockers).

| **Drug pattern** | **All (N=15998)**  **Count (Incidence per 100,000)** | **HCC recurrence (N=12571)**  **Count (Incidence per 100,000)** | **Liver cancer-related mortality (N=3965) Count (Incidence per 100,000)** | **Cancer-related mortality (N=11615) Count (Incidence per 100,000)** | **Non-cancer-related mortality (N=2816) Count (Incidence per 100,000)** | **All-cause mortality (N=14431) Count (Incidence per 100,000)** |
| --- | --- | --- | --- | --- | --- | --- |
| Eye drugs,Hormonal drugs,Steroids | 179 (1119) | 134 (1066) | 31 (782) | 119 (1025) | 47 (1669) | 166 (1150) |
| Eye drugs,Antiestrogen,Anti-cancer drugs | 163 (1019) | 122 (970) | 38 (958) | 134 (1154) | 19 (675) | 153 (1060) |
| Lipid-lowering drugs,Statins and fibrates,Anti-diabetic drugs | 112 (700) | 80 (636) | 21 (530) | 75 (646) | 30 (1065) | 105 (728) |
| Antiplatelets,Anti-diabetic drugs,Non-steroidal anti-inflammatory drugs | 102 (638) | 75 (597) | 20 (504) | 73 (628) | 26 (923) | 99 (686) |
| Antiestrogen,Anti-diabetic drugs,Anti-cancer drugs | 31 (194) | 26 (207) | 1 (25) | 28 (241) | 2 (71) | 30 (208) |
| ACEI/ARB,Lipid-lowering drugs,Anti-diabetic drugs | 29 (181) | 17 (135) | 10 (252) | 22 (189) | 6 (213) | 28 (194) |
| Eye drugs,Anti-diabetic drugs,Hormonal drugs | 11 (69) | 8 (64) | 2 (50) | 9 (77) | 2 (71) | 11 (76) |
| ACEI/ARB,Diuretics for hypertension,Anti-diabetic drugs | 8 (50) | 6 (48) | 2 (50) | 5 (43) | 1 (36) | 6 (42) |
| Lipid-lowering drugs,Antiplatelets,Non-steroidal anti-inflammatory drugs | 7 (44) | 3 (24) | 2 (50) | 5 (43) | 2 (71) | 7 (49) |
| Anticoagulants,Antiplatelets,Anti-diabetic drugs | 7 (44) | 6 (48) | 4 (101) | 7 (60) | 0 | 7 (49) |
| Beta-blockers,Lipid-lowering drugs,Anti-diabetic drugs | 3 (19) | 2 (16) | 1 (25) | 1 (9) | 1 (36) | 2 (14) |
| ACEI/ARB,Antibiotics,Anti-diabetic drugs | 2 (13) | 2 (16) | 2 (50) | 2 (17) | 0 | 2 (14) |
| Calcium channel blockers,Lipid-lowering drugs,Anti-diabetic drugs | 2 (13) | 2 (16) | 1 (25) | 1 (9) | 0 | 1 (7) |
| Diuretics for heart failure,Lipid-lowering drugs,Anti-diabetic drugs | 2 (13) | 2 (16) | 0 | 1 (9) | 1 (36) | 2 (14) |
| Lipid-lowering drugs,Nitrates,Anti-diabetic drugs | 2 (13) | 2 (16) | 0 | 2 (17) | 0 | 2 (14) |
| Antibiotics,Eye drugs,Anti-diabetic drugs | 2 (13) | 2 (16) | 1 (25) | 1 (9) | 1 (36) | 2 (14) |
| Anti-diabetic drugs,Hormonal drugs,Steroids | 2 (13) | 2 (16) | 0 | 1 (9) | 1 (36) | 2 (14) |

### Supplementary Table 10. Top 20 most common double (co-exposure) comorbidity-drug patterns in patients with HCC diagnosis.

Abbreviations: ACEI (angiotensin-converting enzyme inhibitors); ARB (Angiotensin Receptor Blockers).

| **Comorbidity-drug combination pattern** | **All (N=15998)**  **Count (Incidence per 100,000)** | **HCC recurrence (N=12571)**  **Count (Incidence per 100,000)** | **Liver cancer-related mortality (N=3965) Count (Incidence per 100,000)** | **Cancer-related mortality (N=11615) Count (Incidence per 100,000)** | **Non-cancer-related mortality (N=2816) Count (Incidence per 100,000)** | **All-cause mortality (N=14431) Count (Incidence per 100,000)** |
| --- | --- | --- | --- | --- | --- | --- |
| Mild liver disease,Diuretics for heart failure | 209 (1306) | 176 (1400) | 32 (807) | 141 (1214) | 51 (1811) | 192 (1330) |
| Moderate or severe liver,Diuretics for heart failure | 136 (850) | 135 (1074) | 132 (3329) | 134 (1154) | 2 (71) | 136 (942) |
| Mild liver disease,Beta-blockers | 127 (794) | 102 (811) | 25 (631) | 49 (422) | 39 (1385) | 88 (610) |
| Mild liver disease,Anti-cancer drugs | 116 (725) | 102 (811) | 4 (101) | 70 (603) | 27 (959) | 97 (672) |
| Hypertension,Calcium channel blockers | 102 (638) | 78 (620) | 4 (101) | 68 (585) | 18 (639) | 86 (596) |
| Hypertension,ACEI/ARB | 94 (588) | 64 (509) | 3 (76) | 37 (319) | 27 (959) | 64 (443) |
| Moderate or severe liver,Beta-blockers | 51 (319) | 46 (366) | 45 (1135) | 47 (405) | 0 | 47 (326) |
| Breast cancer,Anti-cancer drugs | 45 (281) | 42 (334) | 0 | 35 (301) | 3 (107) | 38 (263) |
| Mild liver disease,Alpha blockers | 40 (250) | 35 (278) | 2 (50) | 13 (112) | 17 (604) | 30 (208) |
| Hypertension,Diuretics for hypertension | 37 (231) | 28 (223) | 3 (76) | 23 (198) | 9 (320) | 32 (222) |
| Moderate or severe liver,Anti-cancer drugs | 36 (225) | 36 (286) | 36 (908) | 36 (310) | 0 | 36 (249) |
| Lung cancer,Anti-cancer drugs | 36 (225) | 29 (231) | 0 | 30 (258) | 4 (142) | 34 (236) |
| Colorectal cancer,Anti-cancer drugs | 35 (219) | 34 (270) | 0 | 25 (215) | 5 (178) | 30 (208) |
| Colorectal cancer,Diuretics for heart failure | 29 (181) | 29 (231) | 0 | 27 (232) | 2 (71) | 29 (201) |
| Diabetes mellitus,Anti-diabetic drugs | 24 (150) | 20 (159) | 1 (25) | 11 (95) | 5 (178) | 16 (111) |
| Mild liver disease,Anticoagulants | 23 (144) | 20 (159) | 0 | 13 (112) | 6 (213) | 19 (132) |
| Renal diseases,Diuretics for heart failure | 18 (113) | 13 (103) | 0 | 11 (95) | 4 (142) | 15 (104) |
| Anaemia,Diuretics for heart failure | 16 (100) | 9 (72) | 2 (50) | 13 (112) | 3 (107) | 16 (111) |
| Mild liver disease,Eye drugs | 14 (88) | 10 (80) | 0 | 6 (52) | 4 (142) | 10 (69) |
| Breast cancer,Diuretics for heart failure | 14 (88) | 12 (95) | 0 | 12 (103) | 1 (36) | 13 (90) |

### Supplementary Table 11. Top 20 common triple (co-exposure) comorbidity-drug patterns in HCC.

Abbreviations: ACEI (angiotensin-converting enzyme inhibitors); ARB (Angiotensin Receptor Blockers).

| **Comorbidity-drug combination pattern** | **All (N=15998)**  **Count (Incidence per 100,000)** | **HCC recurrence (N=12571)**  **Count (Incidence per 100,000)** | **Liver cancer-related mortality (N=3965) Count (Incidence per 100,000)** | **Cancer-related mortality (N=11615) Count (Incidence per 100,000)** | **Non-cancer-related mortality (N=2816) Count (Incidence per 100,000)** | **All-cause mortality (N=14431) Count (Incidence per 100,000)** |
| --- | --- | --- | --- | --- | --- | --- |
| Diabetes mellitus,Moderate or severe liver,Diuretics for heart failure | 148 (925) | 148 (1177) | 147 (3707) | 148 (1274) | 0 | 148 (1026) |
| Diabetes mellitus,Mild liver disease,Anti-diabetic drugs | 121 (756) | 95 (756) | 25 (631) | 55 (474) | 32 (1136) | 87 (603) |
| Diabetes mellitus,Moderate or severe liver,Beta-blockers | 87 (544) | 86 (684) | 87 (2194) | 87 (749) | 0 | 87 (603) |
| Diabetes mellitus,Moderate or severe liver,Anti-diabetic drugs | 81 (506) | 81 (644) | 80 (2018) | 80 (689) | 0 | 80 (554) |
| Diabetes mellitus,Mild liver disease,Diuretics for heart failure | 64 (400) | 54 (430) | 57 (1438) | 60 (517) | 4 (142) | 64 (443) |
| Mild liver disease,Colorectal cancer,Anti-cancer drugs | 61 (381) | 59 (469) | 0 | 45 (387) | 8 (284) | 53 (367) |
| Hypertension,Mild liver disease,ACEI/ARB | 59 (369) | 26 (207) | 15 (378) | 26 (224) | 13 (462) | 39 (270) |
| Mild liver disease,Breast cancer,Anti-cancer drugs | 57 (356) | 49 (390) | 0 | 47 (405) | 7 (249) | 54 (374) |
| Breast cancer,Antiestrogen,Anti-cancer drugs | 51 (319) | 44 (350) | 1 (25) | 45 (387) | 5 (178) | 50 (346) |
| Mild liver disease,Colorectal cancer,Diuretics for heart failure | 48 (300) | 43 (342) | 0 | 41 (353) | 6 (213) | 47 (326) |
| Mild liver disease,Lung cancer,Anti-cancer drugs | 47 (294) | 36 (286) | 0 | 38 (327) | 8 (284) | 46 (319) |
| Hypertension,Moderate or severe liver,Calcium channel blockers | 44 (275) | 42 (334) | 43 (1084) | 43 (370) | 1 (36) | 44 (305) |
| Moderate or severe liver,Colorectal cancer,Diuretics for heart failure | 43 (269) | 40 (318) | 0 | 39 (336) | 2 (71) | 41 (284) |
| Diabetes mellitus,Moderate or severe liver,Anti-cancer drugs | 42 (263) | 42 (334) | 42 (1059) | 42 (362) | 0 | 42 (291) |
| Hypertension,Moderate or severe liver,ACEI/ARB | 39 (244) | 39 (310) | 38 (958) | 38 (327) | 1 (36) | 39 (270) |
| Moderate or severe liver,Colorectal cancer,Anti-cancer drugs | 39 (244) | 36 (286) | 1 (25) | 32 (276) | 3 (107) | 35 (243) |
| Moderate or severe liver,Lung cancer,Anti-cancer drugs | 39 (244) | 31 (247) | 0 | 35 (301) | 4 (142) | 39 (270) |
| Hypertension,Mild liver disease,Calcium channel blockers | 38 (238) | 11 (88) | 8 (202) | 21 (181) | 7 (249) | 28 (194) |
| Diabetes mellitus,Mild liver disease,Beta-blockers | 34 (213) | 31 (247) | 30 (757) | 30 (258) | 2 (71) | 32 (222) |
| Diabetes mellitus,Hypertension,Calcium channel blockers | 30 (188) | 30 (239) | 0 | 16 (138) | 8 (284) | 24 (166) |

### Supplementary Table 12. Top 20 most common quadruple (co-exposure) comorbidity-drug patterns in patients with HCC diagnosis.

| **Comorbidity-drug combination pattern** | **All (N=15998)**  **Count (Incidence per 100,000)** | **HCC recurrence (N=12571)**  **Count (Incidence per 100,000)** | **Liver cancer-related mortality (N=3965) Count (Incidence per 100,000)** | **Cancer-related mortality (N=11615) Count (Incidence per 100,000)** | **Non-cancer-related mortality (N=2816) Count (Incidence per 100,000)** | **All-cause mortality (N=14431) Count (Incidence per 100,000)** |
| --- | --- | --- | --- | --- | --- | --- |
| Diabetes mellitus,Hypertension,Mild liver disease,Calcium channel blockers | 139 (869) | 135 (1074) | 26 (656) | 84 (723) | 30 (1065) | 114 (790) |
| Diabetes mellitus,Hypertension,Mild liver disease,ACEI/ARB | 117 (731) | 111 (883) | 16 (404) | 48 (413) | 34 (1207) | 82 (568) |
| Mild liver disease,Breast cancer,Antiestrogen,Anti-cancer drugs | 70 (438) | 57 (453) | 1 (25) | 61 (525) | 7 (249) | 68 (471) |
| Diabetes mellitus,Hypertension,Moderate or severe liver,ACEI/ARB | 63 (394) | 63 (501) | 62 (1564) | 62 (534) | 0 | 62 (430) |
| Diabetes mellitus,Hypertension,Moderate or severe liver,Calcium channel blockers | 54 (338) | 54 (430) | 53 (1337) | 53 (456) | 1 (36) | 54 (374) |
| Diabetes mellitus,Mild liver disease,Colorectal cancer,Anti-diabetic drugs | 38 (238) | 33 (263) | 0 | 29 (250) | 5 (178) | 34 (236) |
| Diabetes mellitus,Hypertension,Mild liver disease,Anti-diabetic drugs | 30 (188) | 24 (191) | 8 (202) | 18 (155) | 6 (213) | 24 (166) |
| Diabetes mellitus,Hypertension,Moderate or severe liver,Diuretics for hypertension | 30 (188) | 30 (239) | 30 (757) | 30 (258) | 0 | 30 (208) |
| Hypertension,Moderate or severe liver,Colorectal cancer,ACEI/ARB | 29 (181) | 24 (191) | 0 | 23 (198) | 4 (142) | 27 (187) |
| Diabetes mellitus,Moderate or severe liver,Alcoholism,Diuretics for heart failure | 27 (169) | 27 (215) | 27 (681) | 27 (232) | 0 | 27 (187) |
| Diabetes mellitus,Hypertension,ACEI/ARB,Anti-diabetic drugs | 26 (163) | 17 (135) | 3 (76) | 10 (86) | 11 (391) | 21 (146) |
| Diabetes mellitus,Hypertension,Mild liver disease,Diuretics for hypertension | 24 (150) | 23 (183) | 8 (202) | 16 (138) | 5 (178) | 21 (146) |
| Diabetes mellitus,Renal diseases,Moderate or severe liver,Diuretics for heart failure | 22 (138) | 22 (175) | 22 (555) | 22 (189) | 0 | 22 (152) |
| Diabetes mellitus,Renal diseases,Moderate or severe liver,Beta-blockers | 21 (131) | 20 (159) | 21 (530) | 21 (181) | 0 | 21 (146) |
| Diabetes mellitus,Hypertension,Mild liver disease,Beta-blockers | 20 (125) | 19 (151) | 4 (101) | 11 (95) | 5 (178) | 16 (111) |
| Hypertension,Moderate or severe liver,Colorectal cancer,Calcium channel blockers | 20 (125) | 15 (119) | 1 (25) | 17 (146) | 1 (36) | 18 (125) |
| Diabetes mellitus,Mild liver disease,Beta-blockers,Anti-diabetic drugs | 18 (113) | 15 (119) | 7 (177) | 12 (103) | 4 (142) | 16 (111) |
| Diabetes mellitus,Moderate or severe liver,Antiplatelets,Non-steroidal anti-inflammatory drugs | 18 (113) | 18 (143) | 18 (454) | 18 (155) | 0 | 18 (125) |
| Diabetes mellitus,Mild liver disease,Anaemia,Anti-diabetic drugs | 16 (100) | 8 (64) | 6 (151) | 13 (112) | 1 (36) | 14 (97) |
| Diabetes mellitus,Mild liver disease,Diuretics for heart failure,Anti-diabetic drugs | 16 (100) | 15 (119) | 6 (151) | 9 (77) | 6 (213) | 15 (104) |

### Supplementary Table 13. Top 20 most common quintuple (co-exposure) comorbidity-drug patterns in patients with HCC diagnosis.

| **Comorbidity-drug combination pattern** | **All (N=15998)**  **Count (Incidence per 100,000)** | **HCC recurrence (N=12571)**  **Count (Incidence per 100,000)** | **Liver cancer-related mortality (N=3965) Count (Incidence per 100,000)** | **Cancer-related mortality (N=11615) Count (Incidence per 100,000)** | **Non-cancer-related mortality (N=2816) Count (Incidence per 100,000)** | **All-cause mortality (N=14431) Count (Incidence per 100,000)** |
| --- | --- | --- | --- | --- | --- | --- |
| Diabetes mellitus,Hypertension,Mild liver disease,ACEI/ARB,Anti-diabetic drugs | 81 (506) | 65 (517) | 23 (580) | 36 (310) | 22 (781) | 58 (402) |
| Diabetes mellitus,Hypertension,Mild liver disease,Colorectal cancer,ACEI/ARB | 43 (269) | 42 (334) | 1 (25) | 34 (293) | 5 (178) | 39 (270) |
| Diabetes mellitus,Hypertension,Mild liver disease,Colorectal cancer,Calcium channel blockers | 43 (269) | 43 (342) | 1 (25) | 35 (301) | 4 (142) | 39 (270) |
| Diabetes mellitus,Hypertension,Moderate or severe liver,ACEI/ARB,Anti-diabetic drugs | 26 (163) | 25 (199) | 24 (605) | 25 (215) | 1 (36) | 26 (180) |
| Diabetes mellitus,Hypertension,Renal diseases,Mild liver disease,Calcium channel blockers | 22 (138) | 21 (167) | 2 (50) | 7 (60) | 13 (462) | 20 (139) |
| Diabetes mellitus,Hypertension,Renal diseases,Mild liver disease,ACEI/ARB | 21 (131) | 18 (143) | 4 (101) | 9 (77) | 9 (320) | 18 (125) |
| Diabetes mellitus,Hypertension,Renal diseases,Moderate or severe liver,ACEI/ARB | 15 (94) | 15 (119) | 15 (378) | 15 (129) | 0 | 15 (104) |
| Diabetes mellitus,Hypertension,Mild liver disease,Alcoholism,ACEI/ARB | 15 (94) | 15 (119) | 0 | 4 (34) | 4 (142) | 8 (55) |
| Diabetes mellitus,Hypertension,Mild liver disease,Calcium channel blockers,Anti-diabetic drugs | 15 (94) | 15 (119) | 2 (50) | 8 (69) | 6 (213) | 14 (97) |
| Chronic obstructive pulmonary disease,Mild liver disease,Eye drugs,Hormonal drugs,Steroids | 15 (94) | 12 (95) | 1 (25) | 6 (52) | 6 (213) | 12 (83) |
| Diabetes mellitus,Hypertension,Moderate or severe liver,Colorectal cancer,Calcium channel blockers | 14 (88) | 14 (111) | 0 | 10 (86) | 3 (107) | 13 (90) |
| Diabetes mellitus,Hypertension,Renal diseases,Moderate or severe liver,Calcium channel blockers | 13 (81) | 13 (103) | 13 (328) | 13 (112) | 0 | 13 (90) |
| Diabetes mellitus,Hypertension,Mild liver disease,Anaemia,ACEI/ARB | 13 (81) | 11 (88) | 2 (50) | 10 (86) | 2 (71) | 12 (83) |
| Diabetes mellitus,Hypertension,Mild liver disease,Anaemia,Calcium channel blockers | 13 (81) | 12 (95) | 2 (50) | 9 (77) | 2 (71) | 11 (76) |
| Diabetes mellitus,Hypertension,Renal diseases,Mild liver disease,Anti-diabetic drugs | 12 (75) | 9 (72) | 4 (101) | 9 (77) | 1 (36) | 10 (69) |
| Diabetes mellitus,Hypertension,Moderate or severe liver,Colorectal cancer,ACEI/ARB | 12 (75) | 12 (95) | 3 (76) | 10 (86) | 0 | 10 (69) |
| Diabetes mellitus,Hypertension,Mild liver disease,Lung cancer,ACEI/ARB | 11 (69) | 11 (88) | 0 | 10 (86) | 1 (36) | 11 (76) |
| Diabetes mellitus,Hypertension,Chronic obstructive pulmonary disease,Mild liver disease,ACEI/ARB | 10 (63) | 8 (64) | 4 (101) | 8 (69) | 2 (71) | 10 (69) |
| Diabetes mellitus,Hypertension,Mild liver disease,Alcoholism,Calcium channel blockers | 10 (63) | 9 (72) | 1 (25) | 3 (26) | 3 (107) | 6 (42) |
| Diabetes mellitus,Hypertension,Mild liver disease,Pancreas cancer,ACEI/ARB | 10 (63) | 10 (80) | 0 | 9 (77) | 0 | 9 (62) |
| Hyperlipidaemia,Mild liver disease,Colorectal cancer,Lipid-lowering drugs,Statins and fibrates | 10 (63) | 8 (64) | 0 | 6 (52) | 1 (36) | 7 (49) |
| Moderate or severe liver,Breast cancer,Eye drugs,Antiestrogen,Anti-cancer drugs | 10 (63) | 6 (48) | 1 (25) | 9 (77) | 1 (36) | 10 (69) |

### Supplementary Table 14. Descriptive characteristics stratified by recurrence and non-recurrence among patients with primary HCC

^ P value indicates statistical difference between two groups. * for p≤ 0.05, ** for p ≤ 0.01, *** for p ≤ 0.001. Abbreviations: IQR (interquartile range); CCI (Charlson Comorbidity Index); ACEI (angiotensin-converting enzyme inhibitors); ARB (Angiotensin Receptor Blockers); APRI (AST to platelet ratio index); AST (aspartate transaminase); ALT (alanine transaminase); PLT (Platelet); HbA1C (glycated haemoglobin).

| **Characteristics** | **HCC recurrence (N=12571)  Median (IQR); N or Count (%)** | **No HCC recurrence (N=3427)  Median (IQR); N or Count (%)** | **P value^** |
| --- | --- | --- | --- |
| All-cause mortality | 11453(91.10%) | 2978(86.89%) | 0.0961 |
| Within 30 days | 1665(13.24%) | 2323(67.78%) | <0.0001*** |
| Within 30-90 days | 2053(16.33%) | 147(4.28%) | <0.0001*** |
| Within 90-180 days | 1480(11.77%) | 109(3.18%) | <0.0001*** |
| Within 0.5-1 year | 1724(13.71%) | 103(3.00%) | <0.0001*** |
| Within 1-1.5 years | 1245(9.90%) | 68(1.98%) | <0.0001*** |
| Within 1.5-3 years | 1675(13.32%) | 87(2.53%) | <0.0001*** |
| Within 3-5 years | 810(6.44%) | 69(2.01%) | <0.0001*** |
| Within 5-10 years | 621(4.93%) | 52(1.51%) | <0.0001*** |
| Within 10+ years | 180(1.43%) | 20(0.58%) | 0.0001*** |
| Liver cancer-related mortality | 3391(26.97%) | 574(16.74%) | <0.0001*** |
| Within 30 days | 424(3.37%) | 467(13.62%) | <0.0001*** |
| Within 30-90 days | 515(4.09%) | 21(0.61%) | <0.0001*** |
| Within 90-180 days | 374(2.97%) | 13(0.37%) | <0.0001*** |
| Within 0.5-1 year | 490(3.89%) | 13(0.37%) | <0.0001*** |
| Within 1-1.5 years | 352(2.80%) | 21(0.61%) | <0.0001*** |
| Within 1.5-3 years | 571(4.54%) | 16(0.46%) | <0.0001*** |
| Within 3-5 years | 338(2.68%) | 10(0.29%) | <0.0001*** |
| Within 5-10 years | 262(2.08%) | 6(0.17%) | <0.0001*** |
| Within 10+ years | 65(0.51%) | 7(0.20%) | 0.0231* |
| Cancer-related mortality | 9343(74.32%) | 2272(66.29%) | 0.0002*** |
| Within 30 days | 1448(11.51%) | 1887(55.06%) | <0.0001*** |
| Within 30-90 days | 1740(13.84%) | 99(2.88%) | <0.0001*** |
| Within 90-180 days | 1229(9.77%) | 75(2.18%) | <0.0001*** |
| Within 0.5-1 year | 1445(11.49%) | 65(1.89%) | <0.0001*** |
| Within 1-1.5 years | 1036(8.24%) | 45(1.31%) | <0.0001*** |
| Within 1.5-3 years | 1343(10.68%) | 46(1.34%) | <0.0001*** |
| Within 3-5 years | 599(4.76%) | 33(0.96%) | <0.0001*** |
| Within 5-10 years | 414(3.29%) | 14(0.40%) | <0.0001*** |
| Within 10+ years | 89(0.70%) | 8(0.23%) | 0.0024** |
| Non-cancer-related mortality | 2110(16.78%) | 706(20.60%) | <0.0001*** |
| Within 30 days | 217(1.72%) | 436(12.72%) | <0.0001*** |
| Within 30-90 days | 313(2.48%) | 48(1.40%) | 0.0002*** |
| Within 90-180 days | 251(1.99%) | 34(0.99%) | 0.0001*** |
| Within 0.5-1 year | 279(2.21%) | 38(1.10%) | <0.0001*** |
| Within 1-1.5 years | 209(1.66%) | 23(0.67%) | <0.0001*** |
| Within 1.5-3 years | 332(2.64%) | 41(1.19%) | <0.0001*** |
| Within 3-5 years | 211(1.67%) | 36(1.05%) | 0.0114* |
| Within 5-10 years | 207(1.64%) | 38(1.10%) | 0.0305* |
| Within 10+ years | 91(0.72%) | 12(0.35%) | 0.0219* |
| Gender | | | |
| Male | 8016(63.76%) | 2021(58.97%) | 0.0136* |
| Female | 4555(36.23%) | 1406(41.02%) | 0.0006*** |
| Baseline age, years | 67.94(58.07-77.39);n=12571 | 73.84(61.44-82.36);n=3427 | <0.0001*** |
| 18-40 | 266(2.11%) | 60(1.75%) | 0.2123 |
| 40-50 | 961(7.64%) | 176(5.13%) | <0.0001*** |
| 50-60 | 2547(20.26%) | 534(15.58%) | <0.0001*** |
| 60-70 | 3149(25.04%) | 625(18.23%) | <0.0001*** |
| 70-80 | 3321(26.41%) | 917(26.75%) | 0.7767 |
| 80-90 | 2012(16.00%) | 893(26.05%) | <0.0001*** |
| >90 | 315(2.50%) | 222(6.47%) | <0.0001*** |
| CCI | 9.0(4.0-11.0);n=12571 | 10.0(7.0-12.0);n=3427 | <0.0001*** |
| Charlson 0 | 297(2.36%) | 46(1.34%) | 0.0004*** |
| Charlson 1-5 | 3759(29.90%) | 619(18.06%) | <0.0001*** |
| Charlson 6-10 | 5318(42.30%) | 1346(39.27%) | 0.0414* |
| Charlson 11+ | 3197(25.43%) | 1416(41.31%) | <0.0001*** |
| History Disease | | | |
| Diabetes mellitus | 5864(46.64%) | 1035(30.20%) | <0.0001*** |
| Hypertension | 4589(36.50%) | 1568(45.75%) | <0.0001*** |
| Hyperlipidaemia | 572(4.55%) | 199(5.80%) | 0.0044** |
| Heart failure | 527(4.19%) | 269(7.84%) | <0.0001*** |
| Atrial fibrillation | 26(0.20%) | 264(7.70%) | <0.0001*** |
| Acute myocardial infarction | 143(1.13%) | 108(3.15%) | <0.0001*** |
| Stroke/transient ischemic attack | 735(5.84%) | 366(10.67%) | <0.0001*** |
| Peripheral vascular disease | 23(0.18%) | 8(0.23%) | 0.7074 |
| Ischemic heart disease | 808(6.42%) | 375(10.94%) | <0.0001*** |
| Chronic obstructive pulmonary disease | 788(6.26%) | 302(8.81%) | <0.0001*** |
| Renal diseases | 1898(15.09%) | 747(21.79%) | <0.0001*** |
| Mild liver disease | 6189(49.23%) | 2135(62.29%) | <0.0001*** |
| Moderate or severe liver | 3834(30.49%) | 450(13.13%) | <0.0001*** |
| Gastrointestinal bleeding | 1042(8.28%) | 403(11.75%) | <0.0001*** |
| Anaemia | 1642(13.06%) | 747(21.79%) | <0.0001*** |
| Hip fractures | 353(2.80%) | 169(4.93%) | <0.0001*** |
| Accident fall | 970(7.71%) | 437(12.75%) | <0.0001*** |
| Alcoholism | 1148(9.13%) | 306(8.92%) | 0.7631 |
| Obesity | 377(2.99%) | 112(3.26%) | 0.465 |
| Prior cancer | 4561(36.28%) | 1435(41.87%) | <0.0001*** |
| Colorectal cancer | 2247(17.87%) | 497(14.50%) | <0.0001*** |
| Lung cancer | 745(5.92%) | 329(9.60%) | <0.0001*** |
| Pancreas cancer | 304(2.41%) | 159(4.63%) | <0.0001*** |
| Bladder cancer | 82(0.65%) | 57(1.66%) | <0.0001*** |
| Gastric cancer | 261(2.07%) | 97(2.83%) | 0.0118* |
| Prostate cancer | 121(0.96%) | 74(2.15%) | <0.0001*** |
| Nasopharyngeal carcinoma | 199(1.58%) | 61(1.77%) | 0.4723 |
| Head neck cancer | 263(2.09%) | 95(2.77%) | 0.0236* |
| Breast cancer | 609(4.84%) | 168(4.90%) | 0.9298 |
| Ovarian cancer | 96(0.76%) | 39(1.13%) | 0.0457* |
| Cervical cancer | 41(0.32%) | 22(0.64%) | 0.0143* |
| Number of prescribed drug classes | 1.0(1.0-1.0);n=12571 | 1.0(1.0-2.0);n=3427 | 0.0472* |
| Number 0 | 2595(20.64%) | 772(22.52%) | 0.0567 |
| Number 1 | 7282(57.92%) | 1777(51.85%) | 0.0008*** |
| Number 2+ | 2694(21.43%) | 878(25.62%) | <0.0001*** |
| Medications | | | |
| ACEI/ARB | 1805(14.35%) | 634(18.50%) | <0.0001*** |
| Calcium channel blockers | 1108(8.81%) | 322(9.39%) | 0.3509 |
| Alpha-blockers | 302(2.40%) | 89(2.59%) | 0.5648 |
| Beta-blockers | 986(7.84%) | 233(6.79%) | 0.0626 |
| Diuretics for hypertension | 256(2.03%) | 71(2.07%) | 0.953 |
| Diuretics for heart failure | 1724(13.71%) | 330(9.62%) | <0.0001*** |
| Lipid-lowering drugs | 299(2.37%) | 133(3.88%) | <0.0001*** |
| Statins and fibrates | 243(1.93%) | 96(2.80%) | 0.0028** |
| Nitrates | 148(1.17%) | 61(1.77%) | 0.0086** |
| Antibiotics | 31(0.24%) | 4(0.11%) | 0.2173 |
| Anticoagulants | 204(1.62%) | 64(1.86%) | 0.3695 |
| Eye drugs | 462(3.67%) | 138(4.02%) | 0.3823 |
| Antiestrogen | 404(3.21%) | 112(3.26%) | 0.9202 |
| Antiplatelets | 446(3.54%) | 152(4.43%) | 0.0225* |
| Anti-diabetic drugs | 2370(18.85%) | 742(21.65%) | 0.0030** |
| Hormonal drugs | 197(1.56%) | 69(2.01%) | 0.0884 |
| Steroids | 157(1.24%) | 56(1.63%) | 0.102 |
| Non-steroidal anti-inflammatory drugs | 419(3.33%) | 140(4.08%) | 0.0459* |
| Anti-cancer drugs | 1675(13.32%) | 298(8.69%) | <0.0001*** |
| Tyrosine kinase inhibitor | 557(4.43%) | 121(3.53%) | 0.0293* |
| Endocrine therapy | 575(4.57%) | 152(4.43%) | 0.7764 |
| Aromatase inhibitors | 351(2.79%) | 83(2.42%) | 0.2745 |
| Endocrine therapy excluding AIs | 224(1.78%) | 69(2.01%) | 0.4194 |
| Cytotoxic therapy | 798(6.34%) | 97(2.83%) | <0.0001*** |
| Androgen deprivation therapy | 78(0.62%) | 45(1.31%) | <0.0001*** |
| Targerted therapy | 22(0.17%) | 4(0.11%) | 0.6097 |
| Monoclonal antibodies | 562(4.47%) | 35(1.02%) | <0.0001*** |
| Immunomodulatory drugs | 31(0.24%) | 6(0.17%) | 0.5684 |
| Tamoxifen | 224(1.78%) | 69(2.01%) | 0.4194 |
| Anti-HER2 monoclonal antibody | 115(0.91%) | 10(0.29%) | 0.0004*** |
| Anthracycline | 351(2.79%) | 53(1.54%) | <0.0001*** |
| Antimetabolite fluoropyrimidines | 31(0.24%) | 3(0.08%) | 0.1141 |
| Microtubule inhibitor | 41(0.32%) | 15(0.43%) | 0.416 |
| Platinum | 368(2.92%) | 24(0.70%) | <0.0001*** |
| GnRH agonist | 61(0.48%) | 31(0.90%) | 0.0063** |
| Immune checkpoint inhibitors | 74(0.58%) | 7(0.20%) | 0.0077** |
| Laboratory examinations | | | |
| Fibrosis-4 (FIB-4) index | 3.06(1.74-6.19);n=516 | 1.96(1.09-2.87);n=22 | 0.004** |
| APRI | 28.33(13.92-68.42);n=577 | 19.58(10.43-39.71);n=25 | 0.0508 |
| Albumin-Bilirubin (ALBI) score | -1.48(-1.98--0.89);n=3453 | -1.55(-2.05--0.84);n=428 | 0.6647 |
| AST/ALT ratio | 1.2(0.85-1.63);n=1175 | 1.21(0.87-1.74);n=142 | 0.3918 |
| ALT/PLT ratio | 22.88(9.78-60.0);n=1051 | 12.9(7.26-27.74);n=47 | 0.0077** |
| Urea-to-Creatinine ratio | 63.83(51.89-78.18);n=3404 | 66.67(52.69-81.82);n=425 | 0.0342* |
| Mean corpuscular volume, fL | 92.05(86.5-96.7);n=1226 | 91.2(87.35-95.65);n=55 | 0.5585 |
| Basophil, x10^9/L | 0.2(0.0-0.5);n=1043 | 0.42(0.05-0.7);n=46 | 0.0367* |
| Eosinophil, x10^9/L | 0.4(0.1-2.1);n=1056 | 0.73(0.1-2.17);n=48 | 0.4991 |
| Lymphocyte, x10^9/L | 8.6(1.3-24.29);n=1058 | 16.8(1.75-24.6);n=49 | 0.2699 |
| Blast, x10^9/L | 0.0(0.0-0.0);n=123 | 0.0(0.0-0.05);n=4 | 0.4902 |
| Metamyelocyte, x10^9/L | 0.14(0.1-0.28);n=6 | - | - |
| Monocyte, x10^9/L | 4.7(0.5-8.72);n=1058 | 5.5(0.7-7.97);n=49 | 0.5357 |
| Neutrophil, x10^9/L | 46.05(3.72-66.3);n=1058 | 54.3(5.3-69.1);n=49 | 0.3048 |
| White blood bount, x10^9/L | 5.9(4.4-7.7);n=1226 | 6.3(5.19-8.18);n=55 | 0.0753 |
| Mean cell haemoglobin, pg | 31.85(29.6-33.5);n=1226 | 31.3(29.5-33.0);n=55 | 0.095 |
| Myelocyte, x10^9/L | 0.23(0.08-1.0);n=11 | - | - |
| Platelet, x10^9/L | 173.0(110.0-254.0);n=1226 | 212.5(146.5-295.0);n=54 | 0.0121* |
| Reticulocyte, x10^9/L | 2.09(1.5-4.78);n=33 | - | - |
| Red blood count, x10^12/L | 4.03(3.6-4.44);n=1226 | 4.18(3.73-4.53);n=55 | 0.3968 |
| Hematocrit, L/L | 0.37(0.33-0.4);n=1075 | 0.37(0.32-0.4);n=49 | 0.9797 |
| K/Potassium, mmol/L | 4.0(3.7-4.39);n=3400 | 4.09(3.8-4.4);n=425 | 0.1807 |
| Urate, mmol/L | 0.33(0.26-0.39);n=355 | 0.35(0.28-0.44);n=38 | 0.1194 |
| Albumin, g/L | 37.1(32.0-41.1);n=3455 | 37.0(31.7-41.0);n=430 | 0.2266 |
| Na/Sodium, mmol/L | 139.0(137.0-141.0);n=3405 | 139.2(136.0-142.0);n=425 | 0.5434 |
| Urea, mmol/L | 5.1(4.0-6.6);n=3404 | 5.6(4.2-7.1);n=425 | <0.0001*** |
| Protein, g/L | 74.0(68.3-79.0);n=3036 | 73.0(68.0-78.9);n=380 | 0.3542 |
| Creatinine, umol/L | 79.0(65.0-98.0);n=6111 | 85.5(65.85-118.5);n=1383 | <0.0001*** |
| CKD-EPI Creatinine Equation (2021) | 86.16(65.75-98.42);n=6111 | 77.12(49.07-94.74);n=1383 | <0.0001*** |
| Kidney failure (<15) | 80.0(0.63%) | 65.0(1.89%) | <0.0001*** |
| Severe dysfunction (15-30) | 152.0(1.20%) | 121.0(3.53%) | <0.0001*** |
| Moderate dysfunction (30-60) | 931.0(7.40%) | 303.0(8.84%) | 0.0111* |
| Mild renal dysfunction (60-90) | 2375.0(18.89%) | 446.0(13.01%) | <0.0001*** |
| Kidney damage with normal GFR (>90) | 2573.0(20.46%) | 448.0(13.07%) | <0.0001*** |
| Alkaline phosphatase, U/L | 95.0(73.0-134.0);n=3453 | 91.5(72.0-129.5);n=428 | 0.1821 |
| Aspartate transaminase, U/L | 39.0(27.0-67.0);n=1341 | 32.0(22.0-58.5);n=168 | 0.0026** |
| Alanine transaminase, U/L | 31.0(19.0-55.0);n=2896 | 26.0(15.0-44.0);n=355 | <0.0001*** |
| Bilirubin, umol/L | 13.0(8.6-20.0);n=3453 | 12.0(7.89-20.0);n=428 | 0.0689 |
| Triglyceride, mmol/L | 1.08(0.8-1.51);n=1486 | 1.11(0.83-1.54);n=383 | 0.2148 |
| Low-density lipoprotein, mmol/L | 2.68(2.06-3.38);n=1427 | 2.56(1.98-3.21);n=363 | 0.0357* |
| High-density lipoprotein, mmol/L | 1.17(0.92-1.43);n=1438 | 1.11(0.85-1.4);n=367 | 0.0708 |
| Total cholesterol, mmol/L | 4.45(3.73-5.3);n=1491 | 4.4(3.69-5.1);n=385 | 0.0516 |
| HbA1C, % | 6.7(6.0-8.0);n=236 | 6.6(5.8-7.6);n=39 | 0.3246 |
| Fasting glucose, mmol/L | 6.1(5.21-7.9);n=1333 | 6.05(5.2-7.7);n=262 | 0.4944 |

### Supplementary Table 15. Summary statistics of patient characteristics by baseline exposure of drug classes number.

^ P value indicates statistical difference among patients of three drug use subgroups. * for p≤ 0.05, ** for p ≤ 0.01, *** for p ≤ 0.001. Abbreviations: IQR (interquartile range); CCI (Charlson Comorbidity Index); COPD (chronic obstructive pulmonary disease); ACEI (angiotensin-converting enzyme inhibitors); ARB (Angiotensin Receptor Blockers); NSAIDs (non-steroidal anti-inflammatory drugs); APRI (AST to platelet ratio index); AST (aspartate transaminase); ALT (alanine transaminase); PLT (Platelet); HbA1C (glycated haemoglobin).

| **Characteristics** | **Drug class number 0  (N=3367) Median (IQR); N or Count (%)** | **Drug class number 1  (N=9059) Median (IQR); N or Count (%)** | **Drug class number 2+ (N=3572) Median (IQR); N or Count (%)** | **P value^** |
| --- | --- | --- | --- | --- |
| Gender | | | | |
| Male gender | 2274(67.53%) | 5721(63.15%) | 2042(57.16%) | 0.0002*** |
| Female gender | 1093(32.46%) | 3338(36.84%) | 1530(42.83%) | <0.0001*** |
| Baseline age, years | 62.26(53.93-73.73);n=3367 | 69.62(59.26-78.56);n=9059 | 73.45(63.11-81.42);n=3572 | <0.0001*** |
| 18-40 | 153(4.54%) | 142(1.56%) | 31(0.86%) | <0.0001*** |
| 40-50 | 399(11.85%) | 571(6.30%) | 167(4.67%) | <0.0001*** |
| 50-60 | 897(26.64%) | 1719(18.97%) | 465(13.01%) | <0.0001*** |
| 60-70 | 828(24.59%) | 2167(23.92%) | 779(21.80%) | 0.0259* |
| 70-80 | 652(19.36%) | 2505(27.65%) | 1081(30.26%) | <0.0001*** |
| 80-90 | 357(10.60%) | 1655(18.26%) | 893(25.00%) | <0.0001*** |
| >90 | 81(2.40%) | 300(3.31%) | 156(4.36%) | 0.0003*** |
| CCI | 8.0(3.0-10.0);n=3367 | 9.0(4.0-11.0);n=9059 | 10.0(6.0-12.0);n=3572 | <0.0001*** |
| Charlson 0 | 173(5.13%) | 157(1.73%) | 13(0.36%) | <0.0001*** |
| Charlson 1-5 | 925(27.47%) | 2691(29.70%) | 762(21.33%) | <0.0001*** |
| Charlson 6-10 | 1639(48.67%) | 3744(41.32%) | 1281(35.86%) | <0.0001*** |
| Charlson 11+ | 630(18.71%) | 2467(27.23%) | 1516(42.44%) | <0.0001*** |
| History Disease | | | | |
| Diabetes mellitus | 576(17.10%) | 3846(42.45%) | 2477(69.34%) | <0.0001*** |
| Hypertension | 151(4.48%) | 4012(44.28%) | 1994(55.82%) | <0.0001*** |
| Hyperlipidaemia | 30(0.89%) | 214(2.36%) | 527(14.75%) | <0.0001*** |
| Heart failure | 19(0.56%) | 416(4.59%) | 361(10.10%) | <0.0001*** |
| Atrial fibrillation | 44(1.30%) | 150(1.65%) | 96(2.68%) | <0.0001*** |
| Acute myocardial infarction | 12(0.35%) | 123(1.35%) | 116(3.24%) | <0.0001*** |
| Stroke/TIA | 68(2.01%) | 508(5.60%) | 525(14.69%) | <0.0001*** |
| Peripheral vascular disease | 2(0.05%) | 8(0.08%) | 21(0.58%) | <0.0001*** |
| Ischemic heart disease | 41(1.21%) | 549(6.06%) | 593(16.60%) | <0.0001*** |
| COPD | 144(4.27%) | 500(5.51%) | 446(12.48%) | <0.0001*** |
| Renal diseases | 351(10.42%) | 1473(16.26%) | 821(22.98%) | <0.0001*** |
| Mild liver disease | 1591(47.25%) | 4609(50.87%) | 2124(59.46%) | <0.0001*** |
| Moderate or severe liver | 821(24.38%) | 2539(28.02%) | 924(25.86%) | 0.2975 |
| Gastrointestinal bleeding | 265(7.87%) | 813(8.97%) | 367(10.27%) | 0.0083** |
| Anaemia | 383(11.37%) | 1290(14.23%) | 716(20.04%) | <0.0001*** |
| Hip fractures | 96(2.85%) | 266(2.93%) | 160(4.47%) | <0.0001*** |
| Accident fall | 242(7.18%) | 702(7.74%) | 463(12.96%) | <0.0001*** |
| Alcoholism | 251(7.45%) | 787(8.68%) | 416(11.64%) | <0.0001*** |
| Obesity | 86(2.55%) | 233(2.57%) | 170(4.75%) | <0.0001*** |
| Prior cancer | 1172(34.80%) | 3274(36.14%) | 1550(43.39%) | <0.0001*** |
| Colorectal cancer | 573(17.01%) | 1598(17.63%) | 573(16.04%) | 0.0967 |
| Lung cancer | 213(6.32%) | 612(6.75%) | 249(6.97%) | 0.5391 |
| Pancreas cancer | 112(3.32%) | 225(2.48%) | 126(3.52%) | 0.0152* |
| Bladder cancer | 28(0.83%) | 72(0.79%) | 39(1.09%) | 0.1306 |
| Gastric cancer | 84(2.49%) | 194(2.14%) | 80(2.23%) | 1 |
| Prostate cancer | 10(0.29%) | 101(1.11%) | 84(2.35%) | <0.0001*** |
| Nasopharyngeal carcinoma | 98(2.91%) | 124(1.36%) | 38(1.06%) | 0.0038** |
| Head neck cancer | 130(3.86%) | 162(1.78%) | 66(1.84%) | 0.0916 |
| Breast cancer | 21(0.62%) | 348(3.84%) | 408(11.42%) | <0.0001*** |
| Ovarian cancer | 25(0.74%) | 75(0.82%) | 35(0.97%) | 0.3705 |
| Cervical cancer | 25(0.74%) | 27(0.29%) | 11(0.30%) | 0.4386 |
| Number of prescribed drug classes | 0.0(0.0-0.0);n=3367 | 1.0(1.0-1.0);n=9059 | 2.0(2.0-2.0);n=3572 | <0.0001*** |
| Drug class number 0 | 3367(100.00%) | 0(0.00%) | 0(0.00%) | - |
| Drug class number 1 | 0(0.00%) | 9059(100.00%) | 0(0.00%) | - |
| Drug class number 2+ | 0(0.00%) | 0(0.00%) | 3572(100.00%) | - |
| Laboratory examinations | | | | |
| Fibrosis-4 (FIB-4) index | 2.56(1.18-3.79);n=50 | 3.07(1.71-6.35);n=401 | 3.05(1.97-6.01);n=87 | 0.4728 |
| APRI | 22.76(12.25-45.54);n=55 | 28.81(13.76-68.83);n=454 | 26.92(14.04-59.13);n=93 | 0.8613 |
| Albumin-Bilirubin (ALBI) score | -1.74(-2.12--1.23);n=518 | -1.42(-1.96--0.81);n=2671 | -1.5(-1.96--0.93);n=692 | 0.7243 |
| AST/ALT ratio | 1.1(0.75-1.47);n=188 | 1.21(0.88-1.63);n=910 | 1.17(0.87-1.76);n=219 | 0.6191 |
| ALT/PLT ratio | 21.21(8.88-43.02);n=106 | 23.25(10.04-62.65);n=806 | 16.35(8.39-47.92);n=186 | 0.0726 |
| Urea-to-Creatinine ratio | 61.33(50.19-75.42);n=492 | 64.0(52.27-78.81);n=2635 | 64.94(52.87-79.61);n=702 | 0.2163 |
| Mean corpuscular volume, fL | 92.3(86.55-96.65);n=122 | 92.2(86.7-96.8);n=952 | 90.8(85.95-95.7);n=207 | 0.0436* |
| Basophil, x10^9/L | 0.2(0.01-0.58);n=105 | 0.2(0.0-0.5);n=806 | 0.29(0.0-0.6);n=178 | 0.878 |
| Eosinophil, x10^9/L | 0.4(0.1-1.73);n=105 | 0.36(0.1-2.02);n=820 | 0.7(0.1-2.6);n=179 | 0.0573 |
| Lymphocyte, x10^9/L | 8.5(1.5-23.2);n=105 | 8.5(1.3-24.6);n=822 | 11.3(1.4-23.5);n=180 | 0.799 |
| Blast, x10^9/L | 0.0(0.0-0.06);n=9 | 0.0(0.0-0.0);n=103 | 0.0(0.0-0.05);n=15 | 0.1052 |
| Metamyelocyte, x10^9/L | - | 0.17(0.11-0.38);n=5 | 0.09(0.09-0.09);n=1 | 0.6667 |
| Monocyte, x10^9/L | 5.51(0.5-8.32);n=105 | 4.5(0.5-8.68);n=822 | 5.85(0.53-9.45);n=180 | 0.0767 |
| Neutrophil, x10^9/L | 46.6(4.0-66.3);n=105 | 45.3(3.6-66.15);n=822 | 51.5(4.65-68.25);n=180 | 0.3791 |
| White blood bount, x10^9/L | 5.88(4.72-7.47);n=122 | 5.9(4.4-7.68);n=952 | 6.2(4.5-8.28);n=207 | 0.0817 |
| Mean cell haemoglobin, pg | 31.45(29.65-33.3);n=122 | 31.85(29.65-33.6);n=952 | 31.5(29.15-33.3);n=207 | 0.1001 |
| Myelocyte, x10^9/L | - | 0.17(0.05-0.67);n=9 | 2.5(2.5-2.5);n=2 | 0.0982 |
| Platelet, x10^9/L | 193.5(151.5-261.0);n=122 | 169.0(109.5-252.0);n=951 | 183.0(110.0-268.5);n=207 | 0.3892 |
| Reticulocyte, x10^9/L | 14.29(14.29-14.29);n=2 | 1.76(1.44-5.67);n=26 | 2.37(2.26-3.11);n=5 | 0.5302 |
| Red blood count, x10^12/L | 4.15(3.7-4.6);n=122 | 4.0(3.58-4.44);n=952 | 4.06(3.62-4.42);n=207 | 0.5718 |
| Hematocrit, L/L | 0.38(0.33-0.41);n=107 | 0.37(0.33-0.4);n=829 | 0.36(0.33-0.41);n=188 | 0.7599 |
| K/Potassium, mmol/L | 4.0(3.7-4.32);n=491 | 4.0(3.7-4.32);n=2632 | 4.1(3.8-4.4);n=702 | 0.0125* |
| Urate, mmol/L | 0.3(0.23-0.36);n=38 | 0.33(0.26-0.4);n=269 | 0.34(0.27-0.42);n=86 | 0.1428 |
| Albumin, g/L | 39.0(34.8-43.0);n=518 | 37.0(32.0-41.0);n=2673 | 37.0(32.0-41.0);n=694 | 0.2285 |
| Na/Sodium, mmol/L | 140.0(138.0-142.0);n=492 | 139.0(137.0-141.0);n=2636 | 139.0(136.0-141.0);n=702 | 0.0152* |
| Urea, mmol/L | 4.6(3.78-5.9);n=492 | 5.1(4.1-6.7);n=2635 | 5.6(4.2-7.32);n=702 | <0.0001*** |
| Protein, g/L | 74.0(69.0-78.75);n=440 | 74.0(68.1-79.0);n=2362 | 73.0(68.0-77.45);n=614 | 0.0008*** |
| Creatinine, umol/L | 74.0(61.0-90.0);n=1294 | 80.0(66.0-100.0);n=4422 | 86.0(66.0-112.0);n=1778 | <0.0001*** |
| CKD-EPI Creatinine  Equation (2021) | 92.44(78.41-103.16);n=1294 | 84.45(64.07-97.31);n=4422 | 76.21(51.94-93.63);n=1778 | <0.0001*** |
| Kidney failure (<15) | 13.0(0.38%) | 60.0(0.66%) | 72.0(2.01%) | <0.0001*** |
| Severe dysfunction (15-30) | 30.0(0.89%) | 126.0(1.39%) | 117.0(3.27%) | <0.0001*** |
| Moderate dysfunction (30-60) | 111.0(3.29%) | 745.0(8.22%) | 378.0(10.58%) | <0.0001*** |
| Mild renal dysfunction (60-90) | 426.0(12.65%) | 1745.0(19.26%) | 650.0(18.19%) | 0.416 |
| Kidney damage with normal   GFR (>90) | 714.0(21.20%) | 1746.0(19.27%) | 561.0(15.70%) | <0.0001*** |
| Alkaline phosphatase, U/L | 87.0(67.0-119.0);n=518 | 97.0(74.0-137.0);n=2671 | 94.0(72.5-127.5);n=692 | 0.1312 |
| Aspartate transaminase, U/L | 34.0(26.0-58.0);n=208 | 40.0(27.0-68.0);n=1052 | 36.0(25.0-62.0);n=249 | 0.1194 |
| Alanine transaminase, U/L | 31.0(19.0-52.0);n=441 | 31.0(19.0-56.0);n=2223 | 27.0(17.0-46.0);n=587 | 0.0003*** |
| Bilirubin, umol/L | 11.0(8.0-17.0);n=518 | 13.1(8.88-21.0);n=2671 | 12.0(8.0-18.0);n=692 | 0.0185* |
| Triglyceride, mmol/L | 0.98(0.71-1.44);n=157 | 1.09(0.8-1.49);n=990 | 1.12(0.85-1.58);n=722 | 0.0072** |
| Low-density lipoprotein, mmol/L | 2.96(2.32-3.57);n=144 | 2.7(2.08-3.42);n=947 | 2.51(1.97-3.16);n=699 | <0.0001*** |
| High-density lipoprotein, mmol/L | 1.21(0.98-1.5);n=146 | 1.17(0.91-1.43);n=951 | 1.12(0.9-1.42);n=708 | 0.1536 |
| Total cholesterol, mmol/L | 4.78(3.95-5.4);n=158 | 4.53(3.76-5.31);n=994 | 4.3(3.64-5.1);n=724 | 0.0012** |
| HbA1C, % | 5.6(5.3-6.05);n=7 | 6.5(6.0-7.8);n=105 | 6.8(6.05-8.6);n=163 | 0.0445* |
| Fasting glucose, mmol/L | 5.5(5.0-6.5);n=157 | 5.8(5.12-6.9);n=910 | 7.53(5.8-9.3);n=528 | <0.0001*** |

### Supplementary Table 16. Descriptive statistics of patient baseline and clinical characteristics by mortality outcomes among patients after HCC diagnosis.

Abbreviations: IQR (interquartile range); CCI (Charlson Comorbidity Index); COPD (chronic obstructive pulmonary disease); ACEI (angiotensin-converting enzyme inhibitors); ARB (Angiotensin Receptor Blockers); NSAIDs (non-steroidal anti-inflammatory drugs); APRI (AST to platelet ratio index); AST (aspartate transaminase); ALT (alanine transaminase); PLT (Platelet); HbA1C (glycated haemoglobin).

| **Characteristics** | **All-cause mortality  (N=14431) Median (IQR); N or Count(%)** | **Cancer-related mortality (N=11615) Median (IQR); N or Count(%)** | **Non-cancer-related mortality (N=2816)  Median (IQR); N or Count(%)** | **Liver cancer-related mortality (N=3965)  Median (IQR); N or Count(%)** | **Alive (N=1567) Median (IQR); N or Count(%)** |
| --- | --- | --- | --- | --- | --- |
| Gender |  |  |  |  |  |
| Male gender | 8945(61.98%) | 7072(60.88%) | 1873(66.51%) | 2964(74.75%) | 1092(69.68%) |
| Female gender | 5486(38.01%) | 4543(39.11%) | 943(33.48%) | 1001(25.24%) | 475(30.31%) |
| Baseline age, years | 70.2(59.28-79.33);n=14431 | 69.43(58.59-78.88);n=11615 | 73.39(62.73-81.15);n=2816 | 68.83(59.42-78.15);n=3965 | 61.77(54.14-69.77);n=1567 |
| 18-40 | 256(1.77%) | 214(1.84%) | 42(1.49%) | 62(1.56%) | 70(4.46%) |
| 40-50 | 978(6.77%) | 861(7.41%) | 117(4.15%) | 227(5.72%) | 159(10.14%) |
| 50-60 | 2630(18.22%) | 2244(19.31%) | 386(13.70%) | 749(18.89%) | 451(28.78%) |
| 60-70 | 3270(22.65%) | 2638(22.71%) | 632(22.44%) | 1043(26.30%) | 504(32.16%) |
| 70-80 | 3950(27.37%) | 3108(26.75%) | 842(29.90%) | 1081(27.26%) | 288(18.37%) |
| 80-90 | 2817(19.52%) | 2179(18.76%) | 638(22.65%) | 689(17.37%) | 88(5.61%) |
| >90 | 530(3.67%) | 371(3.19%) | 159(5.64%) | 114(2.87%) | 7(0.44%) |
| CCI | 9.0(5.0-11.0);n=14431 | 9.0(6.0-11.0);n=11615 | 8.0(4.0-11.0);n=2816 | 4.0(2.0-7.0);n=3965 | 4.0(2.0-8.0);n=1567 |
| Charlson 0 | 237(1.64%) | 205(1.76%) | 32(1.13%) | 185(4.66%) | 106(6.76%) |
| Charlson 1-5 | 3526(24.43%) | 2621(22.56%) | 905(32.13%) | 2376(59.92%) | 852(54.37%) |
| Charlson 6-10 | 6180(42.82%) | 5164(44.45%) | 1016(36.07%) | 1083(27.31%) | 484(30.88%) |
| Charlson 11+ | 4488(31.09%) | 3625(31.20%) | 863(30.64%) | 321(8.09%) | 125(7.97%) |
| History Disease |  |  |  |  |  |
| Diabetes mellitus | 6444(44.65%) | 5250(45.20%) | 1194(42.40%) | 2852(71.92%) | 455(29.03%) |
| Hypertension | 5635(39.04%) | 4298(37.00%) | 1337(47.47%) | 1480(37.32%) | 522(33.31%) |
| Hyperlipidaemia | 689(4.77%) | 533(4.58%) | 156(5.53%) | 158(3.98%) | 82(5.23%) |
| Heart failure | 785(5.43%) | 566(4.87%) | 219(7.77%) | 194(4.89%) | 11(0.70%) |
| Atrial fibrillation | 289(2.00%) | 223(1.91%) | 66(2.34%) | 64(1.61%) | 1(0.06%) |
| Acute myocardial infarction | 242(1.67%) | 165(1.42%) | 77(2.73%) | 52(1.31%) | 9(0.57%) |
| Stroke/TIA | 1060(7.34%) | 769(6.62%) | 291(10.33%) | 274(6.91%) | 41(2.61%) |
| Peripheral vascular disease | 28(0.19%) | 24(0.20%) | 4(0.14%) | 11(0.27%) | 3(0.19%) |
| Ischemic heart disease | 1119(7.75%) | 804(6.92%) | 315(11.18%) | 296(7.46%) | 64(4.08%) |
| COPD | 1034(7.16%) | 761(6.55%) | 273(9.69%) | 256(6.45%) | 56(3.57%) |
| Renal diseases | 2528(17.51%) | 1924(16.56%) | 604(21.44%) | 698(17.60%) | 117(7.46%) |
| Mild liver disease | 7322(50.73%) | 5419(46.65%) | 1903(67.57%) | 1159(29.23%) | 1002(63.94%) |
| Moderate or severe liver | 4191(29.04%) | 3946(33.97%) | 245(8.70%) | 2656(66.98%) | 93(5.93%) |
| Gastrointestinal bleeding | 1358(9.41%) | 1061(9.13%) | 297(10.54%) | 318(8.02%) | 87(5.55%) |
| Anaemia | 2285(15.83%) | 1792(15.42%) | 493(17.50%) | 444(11.19%) | 104(6.63%) |
| Hip fractures | 499(3.45%) | 379(3.26%) | 120(4.26%) | 128(3.22%) | 23(1.46%) |
| Accident fall | 1330(9.21%) | 992(8.54%) | 338(12.00%) | 358(9.02%) | 77(4.91%) |
| Alcoholism | 1304(9.03%) | 1001(8.61%) | 303(10.75%) | 462(11.65%) | 150(9.57%) |
| Obesity | 447(3.09%) | 347(2.98%) | 100(3.55%) | 123(3.10%) | 42(2.68%) |
| Prior cancer | 5672(39.30%) | 4824(41.53%) | 848(30.11%) | 184(4.64%) | 324(20.67%) |
| Colorectal cancer | 2553(17.69%) | 2115(18.20%) | 438(15.55%) | 59(1.48%) | 191(12.18%) |
| Lung cancer | 1044(7.23%) | 906(7.80%) | 138(4.90%) | 13(0.32%) | 30(1.91%) |
| Pancreas cancer | 443(3.06%) | 387(3.33%) | 56(1.98%) | 12(0.30%) | 20(1.27%) |
| Bladder cancer | 135(0.93%) | 111(0.95%) | 24(0.85%) | 23(0.58%) | 4(0.25%) |
| Gastric cancer | 345(2.39%) | 294(2.53%) | 51(1.81%) | 15(0.37%) | 13(0.82%) |
| Prostate cancer | 184(1.27%) | 144(1.23%) | 40(1.42%) | 30(0.75%) | 11(0.70%) |
| Nasopharyngeal carcinoma | 245(1.69%) | 206(1.77%) | 39(1.38%) | 7(0.17%) | 15(0.95%) |
| Head neck cancer | 338(2.34%) | 280(2.41%) | 58(2.05%) | 14(0.35%) | 20(1.27%) |
| Breast cancer | 745(5.16%) | 657(5.65%) | 88(3.12%) | 17(0.42%) | 32(2.04%) |
| Ovarian cancer | 123(0.85%) | 115(0.99%) | 8(0.28%) | 1(0.02%) | 12(0.76%) |
| Cervical cancer | 60(0.41%) | 58(0.49%) | 2(0.07%) | 3(0.07%) | 3(0.19%) |
| Number of prescribed drug classes | 1.0(1.0-1.0);n=14431 | 1.0(1.0-1.0);n=11615 | 1.0(1.0-2.0);n=2816 | 1.0(1.0-1.0);n=3965 | 1.0(0.0-1.0);n=1567 |
| Drug class number 0 | 2838(19.66%) | 2397(20.63%) | 441(15.66%) | 764(19.26%) | 529(33.75%) |
| Drug class number 1 | 8270(57.30%) | 6667(57.39%) | 1603(56.92%) | 2377(59.94%) | 789(50.35%) |
| Drug class number 2+ | 3323(23.02%) | 2551(21.96%) | 772(27.41%) | 824(20.78%) | 249(15.89%) |
| Medications |  |  |  |  |  |
| ACEI/ARB | 2191(15.18%) | 1630(14.03%) | 561(19.92%) | 602(15.18%) | 248(15.82%) |
| Calcium channel blockers | 1298(8.99%) | 1026(8.83%) | 272(9.65%) | 314(7.91%) | 132(8.42%) |
| Alpha-blockers | 352(2.43%) | 267(2.29%) | 85(3.01%) | 92(2.32%) | 39(2.48%) |
| Beta-blockers | 1092(7.56%) | 841(7.24%) | 251(8.91%) | 433(10.92%) | 127(8.10%) |
| Diuretics for hypertension | 296(2.05%) | 245(2.10%) | 51(1.81%) | 119(3.00%) | 31(1.97%) |
| Diuretics for heart failure | 1984(13.74%) | 1676(14.42%) | 308(10.93%) | 775(19.54%) | 70(4.46%) |
| Lipid-lowering drugs | 382(2.64%) | 289(2.48%) | 93(3.30%) | 77(1.94%) | 50(3.19%) |
| Statins and fibrates | 299(2.07%) | 228(1.96%) | 71(2.52%) | 50(1.26%) | 40(2.55%) |
| Nitrates | 196(1.35%) | 157(1.35%) | 39(1.38%) | 52(1.31%) | 13(0.82%) |
| Antibiotics | 32(0.22%) | 24(0.20%) | 8(0.28%) | 12(0.30%) | 3(0.19%) |
| Anticoagulants | 250(1.73%) | 194(1.67%) | 56(1.98%) | 43(1.08%) | 18(1.14%) |
| Eye drugs | 559(3.87%) | 450(3.87%) | 109(3.87%) | 111(2.79%) | 41(2.61%) |
| Antiestrogen | 492(3.40%) | 435(3.74%) | 57(2.02%) | 54(1.36%) | 24(1.53%) |
| Antiplatelets | 557(3.85%) | 419(3.60%) | 138(4.90%) | 137(3.45%) | 41(2.61%) |
| Anti-diabetic drugs | 2875(19.92%) | 2180(18.76%) | 695(24.68%) | 874(22.04%) | 237(15.12%) |
| Hormonal drugs | 251(1.73%) | 190(1.63%) | 61(2.16%) | 52(1.31%) | 15(0.95%) |
| Steroids | 199(1.37%) | 145(1.24%) | 54(1.91%) | 43(1.08%) | 14(0.89%) |
| NSAIDs | 521(3.61%) | 389(3.34%) | 132(4.68%) | 127(3.20%) | 38(2.42%) |
| Anti-cancer drugs | 1827(12.66%) | 1560(13.43%) | 267(9.48%) | 230(5.80%) | 146(9.31%) |
| Tyrosine kinase inhibitor | 638(4.42%) | 521(4.48%) | 117(4.15%) | 62(1.56%) | 40(2.55%) |
| Endocrine therapy | 693(4.80%) | 604(5.20%) | 89(3.16%) | 77(1.94%) | 34(2.16%) |
| Aromatase inhibitors | 416(2.88%) | 369(3.17%) | 47(1.66%) | 10(0.25%) | 18(1.14%) |
| Endocrine therapy excluding AIs | 277(1.91%) | 235(2.02%) | 42(1.49%) | 67(1.68%) | 16(1.02%) |
| Cytotoxic therapy | 818(5.66%) | 672(5.78%) | 146(5.18%) | 311(7.84%) | 77(4.91%) |
| Androgen deprivation therapy | 105(0.72%) | 79(0.68%) | 26(0.92%) | 17(0.42%) | 18(1.14%) |
| Targerted therapy | 22(0.15%) | 17(0.14%) | 5(0.17%) | 1(0.02%) | 4(0.25%) |
| Monoclonal antibodies | 560(3.88%) | 497(4.27%) | 63(2.23%) | 16(0.40%) | 37(2.36%) |
| Immunomodulatory drugs | 36(0.24%) | 31(0.26%) | 5(0.17%) | 19(0.47%) | 1(0.06%) |
| Tamoxifen | 277(1.91%) | 235(2.02%) | 42(1.49%) | 67(1.68%) | 16(1.02%) |
| Anti-HER2 monoclonal antibody | 112(0.77%) | 96(0.82%) | 16(0.56%) | 0(0.00%) | 13(0.82%) |
| Anthracycline | 372(2.57%) | 323(2.78%) | 49(1.74%) | 116(2.92%) | 32(2.04%) |
| Antimetabolite fluoropyrimidines | 26(0.18%) | 19(0.16%) | 7(0.24%) | 3(0.07%) | 8(0.51%) |
| Microtubule inhibitor | 54(0.37%) | 47(0.40%) | 7(0.24%) | 2(0.05%) | 2(0.12%) |
| Platinum | 357(2.47%) | 276(2.37%) | 81(2.87%) | 190(4.79%) | 35(2.23%) |
| GnRH agonist | 75(0.51%) | 54(0.46%) | 21(0.74%) | 13(0.32%) | 17(1.08%) |
| Immune checkpoint inhibitors | 74(0.51%) | 57(0.49%) | 17(0.60%) | 34(0.85%) | 7(0.44%) |
| Laboratory examinations |  |  |  |  |  |
| Fibrosis-4 (FIB-4) index | 3.06(1.76-5.95);n=492 | 2.92(1.7-5.73);n=391 | 3.34(2.21-6.47);n=101 | 4.41(2.51-8.08);n=201 | 2.19(1.48-5.19);n=46 |
| APRI | 28.36(13.48-66.14);n=552 | 26.85(13.02-64.6);n=439 | 32.38(17.73-74.77);n=113 | 46.14(26.3-89.43);n=226 | 21.97(13.56-67.95);n=50 |
| Albumin-Bilirubin (ALBI) score | -1.46(-1.96--0.86);n=3545 | -1.46(-1.97--0.87);n=2877 | -1.42(-1.92--0.79);n=668 | -1.14(-1.57--0.53);n=1142 | -1.7(-2.17--1.19);n=336 |
| AST/ALT ratio | 1.21(0.87-1.67);n=1182 | 1.22(0.88-1.68);n=950 | 1.19(0.83-1.64);n=232 | 1.18(0.85-1.54);n=415 | 1.05(0.76-1.36);n=135 |
| ALT/PLT ratio | 22.54(9.36-57.65);n=1009 | 22.03(8.93-55.21);n=820 | 24.71(10.59-64.37);n=189 | 48.27(25.94-100.0);n=378 | 21.66(12.42-69.23);n=89 |
| Urea-to-Creatinine ratio | 64.15(52.04-78.82);n=3521 | 64.18(52.23-78.86);n=2860 | 63.95(51.38-78.72);n=661 | 63.59(52.72-77.89);n=1102 | 61.38(50.7-76.88);n=308 |
| Mean corpuscular volume, fL | 91.95(86.6-96.6);n=1178 | 91.8(86.4-96.5);n=957 | 92.3(88.0-97.1);n=221 | 93.6(89.8-97.8);n=437 | 92.4(86.3-97.15);n=103 |
| Basophil, x10^9/L | 0.2(0.0-0.5);n=1000 | 0.2(0.0-0.5);n=818 | 0.2(0.0-0.5);n=182 | 0.1(0.0-0.5);n=352 | 0.1(0.0-0.5);n=89 |
| Eosinophil, x10^9/L | 0.4(0.1-2.08);n=1014 | 0.4(0.1-2.0);n=829 | 0.6(0.12-2.52);n=185 | 0.2(0.1-1.8);n=358 | 0.2(0.1-2.01);n=90 |
| Lymphocyte, x10^9/L | 9.2(1.3-24.4);n=1017 | 9.44(1.32-24.6);n=832 | 9.1(1.2-22.9);n=185 | 2.2(1.1-21.7);n=359 | 3.78(1.25-22.8);n=90 |
| Blast, x10^9/L | 0.0(0.0-0.0);n=119 | 0.0(0.0-0.0);n=106 | 0.0(0.0-0.0);n=13 | 0.0(0.0-0.0);n=53 | 0.0(0.0-0.05);n=8 |
| Metamyelocyte, x10^9/L | 0.14(0.1-0.28);n=6 | 0.14(0.1-0.28);n=6 | - | 0.08(0.08-0.08);n=2 | - |
| Monocyte, x10^9/L | 4.9(0.5-8.71);n=1017 | 4.82(0.5-8.73);n=832 | 5.0(0.5-8.7);n=185 | 0.9(0.4-8.2);n=359 | 2.2(0.4-8.25);n=90 |
| Neutrophil, x10^9/L | 47.4(4.0-66.8);n=1017 | 48.25(4.0-66.85);n=832 | 47.2(3.84-66.2);n=185 | 8.7(3.1-62.15);n=359 | 19.9(2.78-60.9);n=90 |
| White blood bount, x10^9/L | 6.0(4.49-7.8);n=1178 | 5.98(4.5-7.9);n=957 | 6.0(4.21-7.5);n=221 | 5.5(4.29-7.6);n=437 | 5.73(4.3-7.32);n=103 |
| Mean cell haemoglobin, pg | 31.8(29.6-33.5);n=1178 | 31.7(29.4-33.4);n=957 | 32.2(29.8-33.6);n=221 | 32.8(30.9-33.9);n=437 | 32.1(29.8-33.5);n=103 |
| Myelocyte, x10^9/L | 0.2(0.08-0.84);n=10 | 0.23(0.1-1.0);n=9 | 0.04(0.04-0.04);n=1 | 1.0(1.0-1.0);n=1 | 2.0(2.0-2.0);n=1 |
| Platelet, x10^9/L | 175.0(112.0-257.0);n=1178 | 180.0(115.0-263.0);n=957 | 163.0(104.0-226.0);n=221 | 123.0(83.0-174.0);n=437 | 164.5(106.0-243.0);n=102 |
| Reticulocyte, x10^9/L | 2.1(1.52-5.67);n=31 | 1.93(1.52-3.46);n=24 | 17.04(1.55-36.58);n=7 | 2.46(1.37-7.02);n=12 | 1.07(1.07-1.07);n=2 |
| Red blood count, x10^12/L | 4.02(3.58-4.44);n=1178 | 4.02(3.59-4.45);n=957 | 4.0(3.55-4.4);n=221 | 4.01(3.57-4.48);n=437 | 4.16(3.77-4.62);n=103 |
| Hematocrit, L/L | 0.37(0.33-0.4);n=1031 | 0.37(0.33-0.4);n=835 | 0.36(0.33-0.4);n=196 | 0.38(0.34-0.41);n=391 | 0.38(0.35-0.41);n=93 |
| K/Potassium, mmol/L | 4.0(3.7-4.4);n=3517 | 4.0(3.7-4.36);n=2857 | 4.04(3.7-4.4);n=660 | 4.0(3.7-4.3);n=1099 | 4.0(3.7-4.38);n=308 |
| Urate, mmol/L | 0.33(0.26-0.4);n=353 | 0.33(0.26-0.39);n=278 | 0.34(0.26-0.42);n=75 | 0.33(0.26-0.39);n=154 | 0.32(0.25-0.37);n=40 |
| Albumin, g/L | 37.0(32.0-41.0);n=3548 | 37.0(32.0-41.0);n=2880 | 37.0(31.38-41.0);n=668 | 36.0(31.0-39.42);n=1142 | 40.0(35.64-43.28);n=337 |
| Na/Sodium, mmol/L | 139.0(136.9-141.0);n=3522 | 139.0(136.69-141.0);n=2861 | 139.0(137.0-141.0);n=661 | 139.0(136.0-141.0);n=1102 | 140.0(138.0-142.0);n=308 |
| Urea, mmol/L | 5.1(4.09-6.7);n=3521 | 5.1(4.0-6.7);n=2860 | 5.4(4.3-7.1);n=661 | 5.3(4.2-6.8);n=1102 | 4.88(4.0-6.1);n=308 |
| Protein, g/L | 74.0(68.0-79.0);n=3118 | 73.7(68.0-78.7);n=2516 | 74.9(68.0-79.0);n=602 | 74.0(69.0-79.0);n=1046 | 74.0(70.1-79.0);n=298 |
| Creatinine, umol/L | 80.6(65.0-102.75);n=6811 | 79.0(64.0-100.0);n=5421 | 85.0(69.0-111.0);n=1390 | 84.0(70.0-104.0);n=1955 | 77.0(64.0-91.0);n=683 |
| CKD-EPI Creatinine  Equation (2021) | 83.85(61.68-97.36);n=6811 | 84.95(63.35-98.23);n=5421 | 77.86(54.91-93.54);n=1390 | 82.71(60.61-96.11);n=1955 | 92.13(78.97-100.6);n=683 |
| Kidney failure (<15) | 144.0(0.99%) | 86.0(0.74%) | 58.0(2.05%) | 31.0(0.78%) | 1.0(0.06%) |
| Severe dysfunction (15-30) | 270.0(1.87%) | 206.0(1.77%) | 64.0(2.27%) | 96.0(2.42%) | 3.0(0.19%) |
| Moderate dysfunction (30-60) | 1185.0(8.21%) | 894.0(7.69%) | 291.0(10.33%) | 350.0(8.82%) | 49.0(3.12%) |
| Mild renal dysfunction (60-90) | 2569.0(17.80%) | 2038.0(17.54%) | 531.0(18.85%) | 759.0(19.14%) | 252.0(16.08%) |
| Kidney damage with normal   GFR (>90) | 2643.0(18.31%) | 2197.0(18.91%) | 446.0(15.83%) | 719.0(18.13%) | 378.0(24.12%) |
| Alkaline phosphatase, U/L | 96.0(74.0-135.0);n=3545 | 97.0(74.0-137.0);n=2877 | 95.0(74.0-131.0);n=668 | 104.0(79.0-141.0);n=1142 | 82.0(67.0-109.5);n=336 |
| Aspartate transaminase, U/L | 38.0(26.0-67.0);n=1359 | 38.0(26.0-66.0);n=1084 | 42.0(26.0-68.5);n=275 | 53.0(35.5-86.0);n=476 | 37.0(27.0-54.5);n=150 |
| Alanine transaminase, U/L | 30.0(18.0-53.0);n=2958 | 29.0(18.0-52.0);n=2411 | 31.0(20.0-56.5);n=547 | 45.0(29.0-83.0);n=943 | 34.0(22.0-58.0);n=293 |
| Bilirubin, umol/L | 13.0(8.13-20.0);n=3545 | 13.0(8.0-20.0);n=2877 | 13.0(9.0-20.0);n=668 | 18.0(12.7-27.0);n=1142 | 12.9(9.0-18.6);n=336 |
| Triglyceride, mmol/L | 1.09(0.81-1.53);n=1712 | 1.09(0.82-1.5);n=1338 | 1.1(0.8-1.58);n=374 | 0.99(0.78-1.35);n=497 | 1.06(0.78-1.47);n=157 |
| Low-density lipoprotein, mmol/L | 2.65(2.02-3.34);n=1635 | 2.65(2.02-3.35);n=1273 | 2.61(2.03-3.32);n=362 | 2.6(1.95-3.21);n=475 | 2.79(2.25-3.42);n=155 |
| High-density lipoprotein, mmol/L | 1.14(0.9-1.43);n=1651 | 1.14(0.9-1.43);n=1284 | 1.15(0.9-1.41);n=367 | 1.16(0.91-1.47);n=477 | 1.25(0.98-1.49);n=154 |
| Total cholesterol, mmol/L | 4.42(3.7-5.24);n=1719 | 4.44(3.7-5.23);n=1342 | 4.4(3.7-5.26);n=377 | 4.4(3.6-5.13);n=500 | 4.59(3.98-5.42);n=157 |
| HbA1C, % | 6.7(6.0-8.05);n=259 | 6.7(6.0-8.25);n=200 | 6.6(6.2-7.25);n=59 | 6.8(6.0-7.95);n=95 | 6.2(5.9-7.05);n=16 |
| Fasting glucose, mmol/L | 6.1(5.2-7.9);n=1501 | 6.1(5.2-7.9);n=1170 | 6.3(5.32-7.9);n=331 | 6.2(5.2-8.2);n=473 | 6.0(5.16-7.6);n=94 |

### Supplementary Table 17. Descriptive statistics of patient baseline characteristics and subsequent outcomes by different age at primary HCC diagnosis.

Abbreviations: IQR (interquartile range); HCC (Hepatocellular carcinoma); CCI (Charlson Comorbidity Index); COPD (chronic obstructive pulmonary disease); TIA (transient ischemic attack); NPD (nasopharyngeal carcinoma); ACEI (angiotensin-converting enzyme inhibitors); ARB (Angiotensin Receptor Blockers); AST (aspartate transaminase); APRI (AST to platelet ratio index); ALT (alanine transaminase); HbA1C (glycated haemoglobin).

| **Characteristics** | **All (N=15998) Median (IQR);N or Count(%)** | **18-40 (N=326) Median (IQR);N or Count(%)** | **40-50 (N=1137) Median (IQR);N or Count(%)** | **50-60 (N=3081) Median (IQR);N or Count(%)** | **60-70 (N=3774) Median (IQR);N or Count(%)** | **70-80 (N=4238) Median (IQR);N or Count(%)** | **80-90 (N=2905) Median (IQR);N or Count(%)** | **>90 (N=537) Median (IQR);N or Count(%)** |
| --- | --- | --- | --- | --- | --- | --- | --- | --- |
| ***Outcomes*** | | | | | | | | |
| HCC recurrence | 12571(78.57%) | 266(81.59%) | 961(84.52%) | 2547(82.66%) | 3149(83.43%) | 3321(78.36%) | 2012(69.25%) | 315(58.65%) |
| <1 year | 8333(52.08%) | 167(51.22%) | 575(50.57%) | 1531(49.69%) | 1886(49.97%) | 2252(53.13%) | 1633(56.21%) | 289(53.81%) |
| 1-2 years | 2090(13.06%) | 53(16.25%) | 207(18.20%) | 466(15.12%) | 587(15.55%) | 532(12.55%) | 227(7.81%) | 18(3.35%) |
| 2-4 years | 1261(7.88%) | 23(7.05%) | 102(8.97%) | 292(9.47%) | 400(10.59%) | 329(7.76%) | 109(3.75%) | 6(1.11%) |
| >=4 years | 887(5.54%) | 23(7.05%) | 77(6.77%) | 258(8.37%) | 276(7.31%) | 208(4.90%) | 43(1.48%) | 2(0.37%) |
| All-cause mortality | 14431(90.20%) | 256(78.52%) | 978(86.01%) | 2630(85.36%) | 3270(86.64%) | 3950(93.20%) | 2817(96.97%) | 530(98.69%) |
| Within 30 days | 3988(24.92%) | 46(14.11%) | 205(18.02%) | 568(18.43%) | 647(17.14%) | 1115(26.30%) | 1120(38.55%) | 287(53.44%) |
| Within 30-90 days | 2200(13.75%) | 38(11.65%) | 138(12.13%) | 337(10.93%) | 432(11.44%) | 608(14.34%) | 535(18.41%) | 112(20.85%) |
| Within 90-180 days | 1589(9.93%) | 27(8.28%) | 95(8.35%) | 288(9.34%) | 374(9.90%) | 420(9.91%) | 334(11.49%) | 51(9.49%) |
| Within 0.5-1 year | 1827(11.42%) | 44(13.49%) | 138(12.13%) | 366(11.87%) | 433(11.47%) | 517(12.19%) | 290(9.98%) | 39(7.26%) |
| Within 1-1.5 years | 1313(8.20%) | 30(9.20%) | 132(11.60%) | 289(9.38%) | 330(8.74%) | 345(8.14%) | 170(5.85%) | 17(3.16%) |
| Within 1.5-3 years | 1762(11.01%) | 42(12.88%) | 151(13.28%) | 358(11.61%) | 526(13.93%) | 472(11.13%) | 199(6.85%) | 14(2.60%) |
| Within 3-5 years | 879(5.49%) | 16(4.90%) | 62(5.45%) | 206(6.68%) | 244(6.46%) | 241(5.68%) | 102(3.51%) | 8(1.48%) |
| Within 5-10 years | 673(4.20%) | 10(3.06%) | 43(3.78%) | 153(4.96%) | 225(5.96%) | 177(4.17%) | 63(2.16%) | 2(0.37%) |
| Within 10+ years | 200(1.25%) | 3(0.92%) | 14(1.23%) | 65(2.10%) | 59(1.56%) | 55(1.29%) | 4(0.13%) | 0(0.00%) |
| Cancer-related mortality | 11615(72.60%) | 214(65.64%) | 861(75.72%) | 2244(72.83%) | 2638(69.89%) | 3108(73.33%) | 2179(75.00%) | 371(69.08%) |
| Within 30 days | 3335(20.84%) | 37(11.34%) | 182(16.00%) | 503(16.32%) | 557(14.75%) | 934(22.03%) | 914(31.46%) | 208(38.73%) |
| Within 30-90 days | 1839(11.49%) | 35(10.73%) | 124(10.90%) | 303(9.83%) | 372(9.85%) | 503(11.86%) | 419(14.42%) | 83(15.45%) |
| Within 90-180 days | 1304(8.15%) | 22(6.74%) | 81(7.12%) | 253(8.21%) | 307(8.13%) | 340(8.02%) | 267(9.19%) | 34(6.33%) |
| Within 0.5-1 year | 1510(9.43%) | 39(11.96%) | 128(11.25%) | 322(10.45%) | 362(9.59%) | 409(9.65%) | 225(7.74%) | 25(4.65%) |
| Within 1-1.5 years | 1081(6.75%) | 24(7.36%) | 115(10.11%) | 254(8.24%) | 277(7.33%) | 270(6.37%) | 131(4.50%) | 10(1.86%) |
| Within 1.5-3 years | 1389(8.68%) | 36(11.04%) | 137(12.04%) | 293(9.50%) | 401(10.62%) | 373(8.80%) | 142(4.88%) | 7(1.30%) |
| Within 3-5 years | 632(3.95%) | 13(3.98%) | 50(4.39%) | 172(5.58%) | 178(4.71%) | 162(3.82%) | 54(1.85%) | 3(0.55%) |
| Within 5-10 years | 428(2.67%) | 7(2.14%) | 33(2.90%) | 108(3.50%) | 156(4.13%) | 96(2.26%) | 27(0.92%) | 1(0.18%) |
| Within 10+ years | 97(0.60%) | 1(0.30%) | 11(0.96%) | 36(1.16%) | 28(0.74%) | 21(0.49%) | 0(0.00%) | 0(0.00%) |
| Non-cancer-related mortality | 2816(17.60%) | 42(12.88%) | 117(10.29%) | 386(12.52%) | 632(16.74%) | 842(19.86%) | 638(21.96%) | 159(29.60%) |
| Within 30 days | 653(4.08%) | 9(2.76%) | 23(2.02%) | 65(2.10%) | 90(2.38%) | 181(4.27%) | 206(7.09%) | 79(14.71%) |
| Within 30-90 days | 361(2.25%) | 3(0.92%) | 14(1.23%) | 34(1.10%) | 60(1.58%) | 105(2.47%) | 116(3.99%) | 29(5.40%) |
| Within 90-180 days | 285(1.78%) | 5(1.53%) | 14(1.23%) | 35(1.13%) | 67(1.77%) | 80(1.88%) | 67(2.30%) | 17(3.16%) |
| Within 0.5-1 year | 317(1.98%) | 5(1.53%) | 10(0.87%) | 44(1.42%) | 71(1.88%) | 108(2.54%) | 65(2.23%) | 14(2.60%) |
| Within 1-1.5 years | 232(1.45%) | 6(1.84%) | 17(1.49%) | 35(1.13%) | 53(1.40%) | 75(1.76%) | 39(1.34%) | 7(1.30%) |
| Within 1.5-3 years | 373(2.33%) | 6(1.84%) | 14(1.23%) | 65(2.10%) | 125(3.31%) | 99(2.33%) | 57(1.96%) | 7(1.30%) |
| Within 3-5 years | 247(1.54%) | 3(0.92%) | 12(1.05%) | 34(1.10%) | 66(1.74%) | 79(1.86%) | 48(1.65%) | 5(0.93%) |
| Within 5-10 years | 245(1.53%) | 3(0.92%) | 10(0.87%) | 45(1.46%) | 69(1.82%) | 81(1.91%) | 36(1.23%) | 1(0.18%) |
| Within 10+ years | 103(0.64%) | 2(0.61%) | 3(0.26%) | 29(0.94%) | 31(0.82%) | 34(0.80%) | 4(0.13%) | 0(0.00%) |
| Liver cancer-related mortality | 3965(24.78%) | 62(19.01%) | 227(19.96%) | 749(24.31%) | 1043(27.63%) | 1081(25.50%) | 689(23.71%) | 114(21.22%) |
| Within 30 days | 891(5.56%) | 5(1.53%) | 42(3.69%) | 119(3.86%) | 170(4.50%) | 246(5.80%) | 247(8.50%) | 62(11.54%) |
| Within 30-90 days | 536(3.35%) | 12(3.68%) | 36(3.16%) | 104(3.37%) | 123(3.25%) | 125(2.94%) | 109(3.75%) | 27(5.02%) |
| Within 90-180 days | 387(2.41%) | 5(1.53%) | 15(1.31%) | 69(2.23%) | 110(2.91%) | 104(2.45%) | 74(2.54%) | 10(1.86%) |
| Within 0.5-1 year | 503(3.14%) | 16(4.90%) | 30(2.63%) | 103(3.34%) | 127(3.36%) | 152(3.58%) | 71(2.44%) | 4(0.74%) |
| Within 1-1.5 years | 373(2.33%) | 7(2.14%) | 32(2.81%) | 68(2.20%) | 104(2.75%) | 94(2.21%) | 64(2.20%) | 4(0.74%) |
| Within 1.5-3 years | 587(3.66%) | 11(3.37%) | 34(2.99%) | 107(3.47%) | 174(4.61%) | 182(4.29%) | 75(2.58%) | 4(0.74%) |
| Within 3-5 years | 348(2.17%) | 5(1.53%) | 19(1.67%) | 90(2.92%) | 105(2.78%) | 98(2.31%) | 29(0.99%) | 2(0.37%) |
| Within 5-10 years | 268(1.67%) | 1(0.30%) | 11(0.96%) | 64(2.07%) | 107(2.83%) | 64(1.51%) | 20(0.68%) | 1(0.18%) |
| Within 10+ years | 72(0.45%) | 0(0.00%) | 8(0.70%) | 25(0.81%) | 23(0.60%) | 16(0.37%) | 0(0.00%) | 0(0.00%) |
| ***Patient characteristics*** | | | | | | | | |
| Gender | | | | | | | | |
| Male gender | 10037(62.73%) | 170(52.14%) | 630(55.40%) | 1946(63.16%) | 2672(70.80%) | 2794(65.92%) | 1615(55.59%) | 210(39.10%) |
| Female gender | 5961(37.26%) | 156(47.85%) | 507(44.59%) | 1135(36.83%) | 1102(29.19%) | 1444(34.07%) | 1290(44.40%) | 327(60.89%) |
| Baseline age, years | 69.17(58.7-78.67);n=15998 | 35.8(31.87-38.29);n=326 | 46.59(44.15-48.37);n=1137 | 55.7(53.05-58.04);n=3081 | 64.85(62.38-67.52);n=3774 | 75.04(72.53-77.47);n=4238 | 83.81(81.8-86.36);n=2905 | 92.52(91.05-94.57);n=537 |
| 18-40 | 326(2.03%) | 326(99.99%) | 0(0.00%) | 0(0.00%) | 0(0.00%) | 0(0.00%) | 0(0.00%) | 0(0.00%) |
| 40-50 | 1137(7.10%) | 0(0.00%) | 1137(100.00%) | 0(0.00%) | 0(0.00%) | 0(0.00%) | 0(0.00%) | 0(0.00%) |
| 50-60 | 3081(19.25%) | 0(0.00%) | 0(0.00%) | 3081(100.00%) | 0(0.00%) | 0(0.00%) | 0(0.00%) | 0(0.00%) |
| 60-70 | 3774(23.59%) | 0(0.00%) | 0(0.00%) | 0(0.00%) | 3774(100.00%) | 0(0.00%) | 0(0.00%) | 0(0.00%) |
| 70-80 | 4238(26.49%) | 0(0.00%) | 0(0.00%) | 0(0.00%) | 0(0.00%) | 4238(100.00%) | 0(0.00%) | 0(0.00%) |
| 80-90 | 2905(18.15%) | 0(0.00%) | 0(0.00%) | 0(0.00%) | 0(0.00%) | 0(0.00%) | 2905(100.00%) | 0(0.00%) |
| >90 | 537(3.35%) | 0(0.00%) | 0(0.00%) | 0(0.00%) | 0(0.00%) | 0(0.00%) | 0(0.00%) | 537(99.99%) |
| CCI | 9.0(4.0-11.0);n=15998 | 6.0(0.0-8.0);n=326 | 8.0(1.0-8.0);n=1137 | 7.0(2.0-9.0);n=3081 | 8.0(3.0-10.0);n=3774 | 10.0(5.0-11.0);n=4238 | 12.0(8.0-13.0);n=2905 | 12.0(10.0-13.0);n=537 |
| Charlson 0 | 343(2.14%) | 97(29.75%) | 246(21.63%) | 0(0.00%) | 0(0.00%) | 0(0.00%) | 0(0.00%) | 0(0.00%) |
| Charlson 1-5 | 4378(27.36%) | 17(5.21%) | 99(8.70%) | 1152(37.39%) | 1378(36.51%) | 1203(28.38%) | 476(16.38%) | 53(9.86%) |
| Charlson 6-10 | 6664(41.65%) | 212(65.03%) | 784(68.95%) | 1849(60.01%) | 1960(51.93%) | 1039(24.51%) | 680(23.40%) | 140(26.07%) |
| Charlson 11+ | 4613(28.83%) | 0(0.00%) | 8(0.70%) | 80(2.59%) | 436(11.55%) | 1996(47.09%) | 1749(60.20%) | 344(64.05%) |
| History Disease | | | | | | | | |
| Diabetes mellitus | 6899(43.12%) | 46(14.11%) | 245(21.54%) | 1003(32.55%) | 1635(43.32%) | 2094(49.41%) | 1583(54.49%) | 293(54.56%) |
| Hypertension | 6157(38.48%) | 19(5.82%) | 126(11.08%) | 666(21.61%) | 1276(33.81%) | 1958(46.20%) | 1767(60.82%) | 345(64.24%) |
| Hyperlipidaemia | 771(4.81%) | 3(0.92%) | 31(2.72%) | 133(4.31%) | 162(4.29%) | 250(5.89%) | 169(5.81%) | 23(4.28%) |
| Heart failure | 796(4.97%) | 1(0.30%) | 8(0.70%) | 32(1.03%) | 81(2.14%) | 222(5.23%) | 349(12.01%) | 103(19.18%) |
| Atrial fibrillation | 290(1.81%) | 3(0.92%) | 15(1.31%) | 35(1.13%) | 49(1.29%) | 83(1.95%) | 83(2.85%) | 22(4.09%) |
| Acute myocardial infarction | 251(1.56%) | 1(0.30%) | 4(0.35%) | 12(0.38%) | 34(0.90%) | 82(1.93%) | 95(3.27%) | 23(4.28%) |
| Stroke/TIA | 1101(6.88%) | 1(0.30%) | 5(0.43%) | 53(1.72%) | 164(4.34%) | 361(8.51%) | 427(14.69%) | 90(16.75%) |
| Peripheral vascular disease | 31(0.19%) | 0(0.00%) | 1(0.08%) | 2(0.06%) | 7(0.18%) | 10(0.23%) | 10(0.34%) | 1(0.18%) |
| Ischemic heart disease | 1183(7.39%) | 2(0.61%) | 10(0.87%) | 52(1.68%) | 202(5.35%) | 403(9.50%) | 413(14.21%) | 101(18.80%) |
| COPD | 1090(6.81%) | 2(0.61%) | 13(1.14%) | 83(2.69%) | 171(4.53%) | 383(9.03%) | 363(12.49%) | 75(13.96%) |
| Renal diseases | 2645(16.53%) | 35(10.73%) | 106(9.32%) | 327(10.61%) | 493(13.06%) | 741(17.48%) | 779(26.81%) | 164(30.54%) |
| Mild liver disease | 8324(52.03%) | 123(37.73%) | 500(43.97%) | 1487(48.26%) | 1912(50.66%) | 2238(52.80%) | 1709(58.82%) | 355(66.10%) |
| Moderate or severe liver | 4284(26.77%) | 46(14.11%) | 178(15.65%) | 592(19.21%) | 1233(32.67%) | 1301(30.69%) | 826(28.43%) | 108(20.11%) |
| Gastrointestinal bleeding | 1445(9.03%) | 10(3.06%) | 48(4.22%) | 182(5.90%) | 285(7.55%) | 418(9.86%) | 396(13.63%) | 106(19.73%) |
| Anaemia | 2389(14.93%) | 30(9.20%) | 115(10.11%) | 307(9.96%) | 425(11.26%) | 654(15.43%) | 699(24.06%) | 159(29.60%) |
| Hip fractures | 522(3.26%) | 4(1.22%) | 8(0.70%) | 31(1.00%) | 61(1.61%) | 129(3.04%) | 204(7.02%) | 85(15.82%) |
| Accident fall | 1407(8.79%) | 10(3.06%) | 23(2.02%) | 129(4.18%) | 221(5.85%) | 337(7.95%) | 506(17.41%) | 181(33.70%) |
| Alcoholism | 1454(9.08%) | 21(6.44%) | 75(6.59%) | 245(7.95%) | 356(9.43%) | 407(9.60%) | 297(10.22%) | 53(9.86%) |
| Obesity | 489(3.05%) | 7(2.14%) | 26(2.28%) | 87(2.82%) | 90(2.38%) | 138(3.25%) | 117(4.02%) | 24(4.46%) |
| Prior cancer | 5996(37.47%) | 115(35.27%) | 451(39.66%) | 1130(36.67%) | 1325(35.10%) | 1572(37.09%) | 1209(41.61%) | 194(36.12%) |
| Colorectal cancer | 2744(17.15%) | 32(9.81%) | 114(10.02%) | 397(12.88%) | 628(16.64%) | 824(19.44%) | 637(21.92%) | 112(20.85%) |
| Lung cancer | 1074(6.71%) | 6(1.84%) | 73(6.42%) | 173(5.61%) | 242(6.41%) | 311(7.33%) | 238(8.19%) | 31(5.77%) |
| Pancreas cancer | 463(2.89%) | 1(0.30%) | 24(2.11%) | 77(2.49%) | 125(3.31%) | 121(2.85%) | 101(3.47%) | 14(2.60%) |
| Bladder cancer | 139(0.86%) | 0(0.00%) | 2(0.17%) | 8(0.25%) | 21(0.55%) | 51(1.20%) | 46(1.58%) | 11(2.04%) |
| Gastric cancer | 358(2.23%) | 4(1.22%) | 14(1.23%) | 52(1.68%) | 78(2.06%) | 109(2.57%) | 92(3.16%) | 9(1.67%) |
| Prostate cancer | 195(1.21%) | 0(0.00%) | 0(0.00%) | 1(0.03%) | 33(0.87%) | 76(1.79%) | 71(2.44%) | 14(2.60%) |
| NPC | 260(1.62%) | 19(5.82%) | 52(4.57%) | 96(3.11%) | 59(1.56%) | 19(0.44%) | 15(0.51%) | 0(0.00%) |
| Head neck cancer | 358(2.23%) | 20(6.13%) | 60(5.27%) | 116(3.76%) | 82(2.17%) | 45(1.06%) | 35(1.20%) | 0(0.00%) |
| Breast cancer | 777(4.85%) | 39(11.96%) | 138(12.13%) | 266(8.63%) | 148(3.92%) | 104(2.45%) | 72(2.47%) | 10(1.86%) |
| Ovarian cancer | 135(0.84%) | 5(1.53%) | 29(2.55%) | 57(1.85%) | 22(0.58%) | 12(0.28%) | 8(0.27%) | 2(0.37%) |
| Cervical cancer | 63(0.39%) | 8(2.45%) | 7(0.61%) | 13(0.42%) | 12(0.31%) | 14(0.33%) | 7(0.24%) | 2(0.37%) |
| Number of prescribed drug classes | 1.0(1.0-1.0);n=15998 | 1.0(0.0-1.0);n=326 | 1.0(0.0-1.0);n=1137 | 1.0(0.0-1.0);n=3081 | 1.0(1.0-1.0);n=3774 | 1.0(1.0-2.0);n=4238 | 1.0(1.0-2.0);n=2905 | 1.0(1.0-2.0);n=537 |
| Drug class number 0 | 3367(21.04%) | 153(46.93%) | 399(35.09%) | 897(29.11%) | 828(21.93%) | 652(15.38%) | 357(12.28%) | 81(15.08%) |
| Drug class number 1 | 9059(56.62%) | 142(43.55%) | 571(50.21%) | 1719(55.79%) | 2167(57.41%) | 2505(59.10%) | 1655(56.97%) | 300(55.86%) |
| Drug class number 2+ | 3572(22.32%) | 31(9.50%) | 167(14.68%) | 465(15.09%) | 779(20.64%) | 1081(25.50%) | 893(30.74%) | 156(29.05%) |
| Medications | | | | | | | | |
| ACEI/ARB | 2439(15.24%) | 7(2.14%) | 49(4.30%) | 274(8.89%) | 511(13.54%) | 785(18.52%) | 681(23.44%) | 132(24.58%) |
| Calcium channel blockers | 1430(8.93%) | 7(2.14%) | 38(3.34%) | 164(5.32%) | 322(8.53%) | 424(10.00%) | 394(13.56%) | 81(15.08%) |
| Alpha-blockers | 391(2.44%) | 0(0.00%) | 1(0.08%) | 30(0.97%) | 72(1.90%) | 144(3.39%) | 127(4.37%) | 17(3.16%) |
| Beta-blockers | 1219(7.61%) | 6(1.84%) | 54(4.74%) | 242(7.85%) | 294(7.79%) | 366(8.63%) | 219(7.53%) | 38(7.07%) |
| Diuretics for hypertension | 327(2.04%) | 5(1.53%) | 19(1.67%) | 53(1.72%) | 71(1.88%) | 105(2.47%) | 60(2.06%) | 14(2.60%) |
| Diuretics for heart failure | 2054(12.83%) | 42(12.88%) | 159(13.98%) | 468(15.18%) | 501(13.27%) | 549(12.95%) | 278(9.56%) | 57(10.61%) |
| Lipid-lowering drugs | 432(2.70%) | 0(0.00%) | 12(1.05%) | 45(1.46%) | 94(2.49%) | 145(3.42%) | 120(4.13%) | 16(2.97%) |
| Statins and fibrates | 339(2.11%) | 0(0.00%) | 12(1.05%) | 41(1.33%) | 77(2.04%) | 115(2.71%) | 86(2.96%) | 8(1.48%) |
| Nitrates | 209(1.30%) | 0(0.00%) | 4(0.35%) | 17(0.55%) | 42(1.11%) | 65(1.53%) | 70(2.40%) | 11(2.04%) |
| Antibiotics | 35(0.21%) | 4(1.22%) | 3(0.26%) | 8(0.25%) | 11(0.29%) | 6(0.14%) | 3(0.10%) | 0(0.00%) |
| Anticoagulants | 268(1.67%) | 8(2.45%) | 26(2.28%) | 47(1.52%) | 63(1.66%) | 75(1.76%) | 45(1.54%) | 4(0.74%) |
| Eye drugs | 600(3.75%) | 9(2.76%) | 60(5.27%) | 109(3.53%) | 141(3.73%) | 148(3.49%) | 114(3.92%) | 19(3.53%) |
| Antiestrogen | 516(3.22%) | 23(7.05%) | 102(8.97%) | 159(5.16%) | 91(2.41%) | 76(1.79%) | 53(1.82%) | 12(2.23%) |
| Antiplatelets | 598(3.73%) | 1(0.30%) | 17(1.49%) | 42(1.36%) | 123(3.25%) | 187(4.41%) | 188(6.47%) | 40(7.44%) |
| Anti-diabetic drugs | 3112(19.45%) | 8(2.45%) | 72(6.33%) | 406(13.17%) | 747(19.79%) | 988(23.31%) | 766(26.36%) | 125(23.27%) |
| Hormonal drugs | 266(1.66%) | 1(0.30%) | 5(0.43%) | 20(0.64%) | 52(1.37%) | 95(2.24%) | 81(2.78%) | 12(2.23%) |
| Steroids | 213(1.33%) | 0(0.00%) | 3(0.26%) | 18(0.58%) | 40(1.05%) | 73(1.72%) | 67(2.30%) | 12(2.23%) |
| NSAIDs | 559(3.49%) | 1(0.30%) | 17(1.49%) | 40(1.29%) | 115(3.04%) | 174(4.10%) | 173(5.95%) | 39(7.26%) |
| Anti-cancer drugs | 1973(12.33%) | 87(26.68%) | 295(25.94%) | 557(18.07%) | 511(13.54%) | 371(8.75%) | 136(4.68%) | 16(2.97%) |
| Tyrosine kinase inhibitor | 678(4.23%) | 14(4.29%) | 79(6.94%) | 153(4.96%) | 186(4.92%) | 172(4.05%) | 67(2.30%) | 7(1.30%) |
| Endocrine therapy | 727(4.54%) | 27(8.28%) | 129(11.34%) | 214(6.94%) | 129(3.41%) | 130(3.06%) | 81(2.78%) | 17(3.16%) |
| Aromatase inhibitors | 434(2.71%) | 18(5.52%) | 84(7.38%) | 147(4.77%) | 71(1.88%) | 64(1.51%) | 42(1.44%) | 8(1.48%) |
| Endocrine therapy excluding AIs | 293(1.83%) | 9(2.76%) | 45(3.95%) | 67(2.17%) | 58(1.53%) | 66(1.55%) | 39(1.34%) | 9(1.67%) |
| Cytotoxic therapy | 895(5.59%) | 40(12.26%) | 106(9.32%) | 263(8.53%) | 257(6.80%) | 192(4.53%) | 35(1.20%) | 2(0.37%) |
| Androgen deprivation therapy | 123(0.76%) | 7(2.14%) | 3(0.26%) | 6(0.19%) | 18(0.47%) | 51(1.20%) | 35(1.20%) | 3(0.55%) |
| Targerted therapy | 26(0.16%) | 2(0.61%) | 4(0.35%) | 10(0.32%) | 5(0.13%) | 5(0.11%) | 0(0.00%) | 0(0.00%) |
| Monoclonal antibodies | 597(3.73%) | 19(5.82%) | 70(6.15%) | 172(5.58%) | 201(5.32%) | 117(2.76%) | 17(0.58%) | 1(0.18%) |
| Immunomodulatory drugs | 37(0.23%) | 0(0.00%) | 7(0.61%) | 7(0.22%) | 7(0.18%) | 12(0.28%) | 4(0.13%) | 0(0.00%) |
| Tamoxifen | 293(1.83%) | 9(2.76%) | 45(3.95%) | 67(2.17%) | 58(1.53%) | 66(1.55%) | 39(1.34%) | 9(1.67%) |
| Anti-HER2 monoclonal antibody | 125(0.78%) | 9(2.76%) | 23(2.02%) | 45(1.46%) | 36(0.95%) | 12(0.28%) | 0(0.00%) | 0(0.00%) |
| Anthracycline | 404(2.52%) | 24(7.36%) | 58(5.10%) | 142(4.60%) | 106(2.80%) | 63(1.48%) | 10(0.34%) | 1(0.18%) |
| Antimetabolite fluoropyrimidines | 34(0.21%) | 1(0.30%) | 2(0.17%) | 12(0.38%) | 11(0.29%) | 7(0.16%) | 1(0.03%) | 0(0.00%) |
| Microtubule inhibitor | 56(0.35%) | 1(0.30%) | 9(0.79%) | 18(0.58%) | 15(0.39%) | 12(0.28%) | 1(0.03%) | 0(0.00%) |
| Platinum | 392(2.45%) | 14(4.29%) | 37(3.25%) | 88(2.85%) | 123(3.25%) | 107(2.52%) | 22(0.75%) | 1(0.18%) |
| GnRH agonist | 92(0.57%) | 7(2.14%) | 3(0.26%) | 6(0.19%) | 12(0.31%) | 39(0.92%) | 23(0.79%) | 2(0.37%) |
| Immune checkpoint inhibitors | 81(0.50%) | 6(1.84%) | 11(0.96%) | 15(0.48%) | 31(0.82%) | 13(0.30%) | 5(0.17%) | 0(0.00%) |
| Laboratory examinations | | | | | | | | |
| Fibrosis-4 (FIB-4) index | 3.0(1.71-5.88);n=538 | 1.07(0.82-3.84);n=16 | 1.79(0.93-3.08);n=52 | 2.62(1.51-4.92);n=142 | 3.36(2.17-6.89);n=146 | 3.44(2.13-6.4);n=138 | 4.12(2.29-7.44);n=40 | 9.03(4.45-14.25);n=4 |
| APRI | 27.92(13.48-66.41);n=602 | 14.87(11.69-47.08);n=19 | 20.44(10.31-37.61);n=61 | 28.72(13.47-64.83);n=157 | 34.65(17.82-79.33);n=155 | 28.26(15.34-69.23);n=153 | 24.14(11.08-53.17);n=50 | 29.51(23.47-101.9);n=7 |
| Albumin-Bilirubin (ALBI) score | -1.48(-1.99--0.88);n=3881 | -1.86(-2.34--1.27);n=92 | -1.83(-2.26--1.28);n=366 | -1.61(-2.15--1.01);n=923 | -1.37(-1.9--0.85);n=961 | -1.38(-1.87--0.81);n=1037 | -1.36(-1.72--0.68);n=444 | -1.21(-1.61--0.55);n=58 |
| AST/ALT ratio | 1.2(0.86-1.64);n=1317 | 1.43(0.88-1.95);n=40 | 1.1(0.81-1.59);n=141 | 1.09(0.79-1.46);n=324 | 1.21(0.85-1.59);n=333 | 1.22(0.9-1.65);n=351 | 1.45(1.03-2.01);n=116 | 1.83(1.05-2.35);n=12 |
| ALT/PLT ratio | 22.52(9.58-58.6);n=1098 | 24.37(9.88-51.83);n=24 | 15.9(7.9-38.55);n=100 | 23.13(10.45-55.36);n=277 | 33.33(12.34-80.37);n=315 | 19.81(8.49-53.28);n=288 | 13.39(5.7-28.62);n=84 | 19.07(7.54-30.01);n=10 |
| Urea-to-Creatinine ratio | 64.04(51.95-78.72);n=3829 | 54.76(46.81-72.83);n=85 | 61.89(48.66-75.67);n=352 | 63.24(50.68-78.23);n=895 | 62.35(51.35-75.95);n=948 | 64.12(53.53-80.0);n=1040 | 70.46(56.94-83.89);n=452 | 76.07(62.17-98.53);n=57 |
| Mean corpuscular volume, fL | 92.0(86.6-96.7);n=1281 | 91.05(85.4-94.3);n=30 | 90.7(84.0-95.55);n=124 | 91.9(86.1-96.45);n=331 | 92.6(88.0-97.2);n=352 | 93.0(87.95-97.05);n=326 | 90.3(84.05-95.0);n=104 | 89.45(88.2-95.3);n=14 |
| Basophil, x10^9/L | 0.2(0.0-0.5);n=1089 | 0.1(0.0-0.6);n=27 | 0.25(0.0-0.5);n=112 | 0.2(0.01-0.6);n=280 | 0.18(0.0-0.5);n=308 | 0.2(0.0-0.6);n=278 | 0.2(0.0-0.5);n=72 | 0.15(0.0-0.3);n=12 |
| Eosinophil, x10^9/L | 0.4(0.1-2.1);n=1104 | 0.25(0.08-1.34);n=28 | 0.32(0.1-2.27);n=112 | 0.4(0.1-1.85);n=284 | 0.3(0.1-2.1);n=312 | 0.6(0.1-2.1);n=281 | 0.5(0.1-2.2);n=75 | 0.5(0.25-2.5);n=12 |
| Lymphocyte, x10^9/L | 8.87(1.3-24.35);n=1107 | 8.25(1.3-17.8);n=28 | 10.7(1.3-26.5);n=112 | 9.79(1.4-25.3);n=284 | 6.23(1.2-25.4);n=312 | 11.5(1.3-23.85);n=282 | 9.55(1.4-19.65);n=76 | 7.4(1.76-15.0);n=13 |
| Blast, x10^9/L | 0.0(0.0-0.0);n=127 | 0.0(0.0-0.0);n=2 | 0.0(0.0-0.0);n=13 | 0.0(0.0-0.0);n=33 | 0.0(0.0-0.0);n=37 | 0.0(0.0-0.0);n=33 | 0.0(0.0-0.0);n=9 | - |
| Metamyelocyte, x10^9/L | 0.14(0.1-0.28);n=6 | 0.67(0.67-0.67);n=1 | - | 0.06(0.06-0.06);n=2 | 0.11(0.11-0.11);n=1 | - | 0.28(0.28-0.28);n=2 | - |
| Monocyte, x10^9/L | 4.7(0.5-8.7);n=1107 | 2.0(0.35-7.77);n=28 | 5.5(0.5-9.27);n=112 | 4.86(0.4-8.6);n=284 | 3.23(0.48-8.5);n=312 | 5.3(0.5-8.9);n=282 | 4.8(0.6-8.37);n=76 | 5.9(0.77-9.0);n=13 |
| Neutrophil, x10^9/L | 46.6(3.78-66.5);n=1107 | 53.35(3.12-69.65);n=28 | 50.65(4.3-64.95);n=112 | 44.5(3.2-65.95);n=284 | 42.9(3.39-61.85);n=312 | 51.09(4.4-69.15);n=282 | 54.25(4.7-69.65);n=76 | 58.2(5.3-73.9);n=13 |
| White blood bount, x10^9/L | 5.97(4.4-7.8);n=1281 | 5.9(3.78-7.57);n=30 | 5.9(4.1-8.16);n=124 | 5.8(4.4-7.65);n=331 | 5.9(4.4-7.7);n=352 | 5.93(4.53-7.64);n=326 | 6.7(5.15-8.22);n=104 | 7.0(6.0-8.45);n=14 |
| Mean cell haemoglobin, pg | 31.8(29.6-33.5);n=1281 | 31.8(29.3-33.3);n=30 | 30.95(28.85-33.0);n=124 | 31.7(29.25-33.5);n=331 | 32.2(30.0-33.8);n=352 | 32.15(30.2-33.55);n=326 | 31.05(28.75-33.35);n=104 | 30.55(29.5-32.9);n=14 |
| Myelocyte, x10^9/L | 0.23(0.08-1.0);n=11 | 0.67(0.38-1.34);n=3 | - | 1.0(0.52-2.5);n=3 | 0.05(0.04-0.52);n=3 | - | 0.2(0.2-0.2);n=2 | - |
| Platelet, x10^9/L | 174.0(111.0-255.5);n=1280 | 193.0(117.0-302.5);n=30 | 220.5(154.0-275.5);n=124 | 178.0(118.0-267.5);n=330 | 152.5(96.0-233.5);n=352 | 170.5(111.0-239.5);n=326 | 198.5(132.5-283.0);n=104 | 222.5(161.0-283.5);n=14 |
| Reticulocyte, x10^9/L | 2.09(1.5-4.78);n=33 | 4.78(4.78-4.78);n=1 | 25.96(25.96-25.96);n=2 | 26.47(26.47-26.47);n=2 | 1.5(1.24-2.37);n=13 | 1.93(1.66-2.86);n=8 | 3.11(1.82-15.62);n=7 | - |
| Red blood count, x10^12/L | 4.03(3.6-4.45);n=1281 | 4.17(3.82-4.52);n=30 | 4.09(3.67-4.65);n=124 | 4.1(3.68-4.61);n=331 | 4.05(3.68-4.43);n=352 | 3.96(3.57-4.36);n=326 | 3.85(3.34-4.25);n=104 | 3.78(3.46-4.14);n=14 |
| Hematocrit, L/L | 0.37(0.33-0.4);n=1124 | 0.36(0.33-0.41);n=23 | 0.36(0.33-0.4);n=103 | 0.37(0.33-0.41);n=281 | 0.38(0.34-0.4);n=317 | 0.36(0.33-0.4);n=297 | 0.34(0.31-0.38);n=91 | 0.33(0.27-0.36);n=12 |
| K/Potassium, mmol/L | 4.0(3.7-4.4);n=3825 | 4.0(3.7-4.3);n=85 | 4.0(3.7-4.3);n=352 | 4.0(3.76-4.31);n=894 | 4.0(3.7-4.3);n=947 | 4.1(3.8-4.4);n=1039 | 4.1(3.7-4.4);n=451 | 4.0(3.6-4.4);n=57 |
| Urate, mmol/L | 0.33(0.26-0.4);n=393 | 0.32(0.28-0.33);n=6 | 0.27(0.24-0.32);n=31 | 0.32(0.26-0.38);n=112 | 0.33(0.26-0.38);n=108 | 0.35(0.26-0.43);n=97 | 0.36(0.28-0.45);n=35 | 0.52(0.36-0.82);n=4 |
| Albumin, g/L | 37.0(32.0-41.1);n=3885 | 40.0(35.0-44.0);n=92 | 40.0(35.25-43.0);n=366 | 38.7(34.0-42.7);n=925 | 37.0(32.0-41.0);n=962 | 36.72(31.1-40.42);n=1038 | 34.1(29.05-39.0);n=444 | 32.65(26.5-37.0);n=58 |
| Na/Sodium, mmol/L | 139.0(137.0-141.07);n=3830 | 140.0(138.0-142.0);n=85 | 140.0(138.0-142.0);n=352 | 139.56(137.0-141.51);n=896 | 139.0(137.0-141.0);n=948 | 139.0(136.0-141.04);n=1040 | 139.0(136.0-141.0);n=452 | 139.0(135.0-142.0);n=57 |
| Urea, mmol/L | 5.1(4.09-6.7);n=3829 | 3.87(3.0-4.88);n=85 | 4.3(3.4-5.18);n=352 | 4.7(3.9-5.9);n=895 | 5.0(4.08-6.5);n=948 | 5.6(4.4-7.1);n=1040 | 6.4(4.8-8.2);n=452 | 7.2(5.3-9.5);n=57 |
| Protein, g/L | 74.0(68.0-79.0);n=3416 | 77.0(70.0-80.4);n=78 | 75.0(70.0-79.9);n=325 | 75.0(70.0-80.0);n=809 | 74.0(69.0-79.0);n=843 | 73.8(68.0-78.1);n=914 | 70.0(65.4-76.2);n=394 | 69.5(59.1-74.0);n=53 |
| Creatinine, umol/L | 80.0(65.0-101.0);n=7494 | 66.0(54.0-78.0);n=145 | 69.0(58.0-83.0);n=510 | 74.0(62.0-89.0);n=1459 | 80.0(65.0-96.0);n=1809 | 85.0(69.0-105.0);n=2012 | 92.0(71.0-120.0);n=1332 | 96.0(69.0-124.55);n=227 |
| CKD-EPI Creatinine  Equation (2021) | 84.89(63.15-97.91);n=7494 | 116.18(104.53-121.94);n=145 | 107.48(95.24-112.18);n=510 | 99.57(83.22-105.4);n=1459 | 88.7(70.96-98.21);n=1809 | 76.8(59.48-90.45);n=2012 | 63.7(45.63-83.8);n=1332 | 54.87(39.25-78.65);n=227 |
| Kidney failure (<15) | 145.0(0.90%) | 1.0(0.30%) | 8.0(0.70%) | 20.0(0.64%) | 28.0(0.74%) | 35.0(0.82%) | 47.0(1.61%) | 6.0(1.11%) |
| Severe dysfunction (15-30) | 273.0(1.70%) | 1.0(0.30%) | 6.0(0.52%) | 20.0(0.64%) | 42.0(1.11%) | 76.0(1.79%) | 103.0(3.54%) | 25.0(4.65%) |
| Moderate dysfunction (30-60) | 1234.0(7.71%) | 0.0(0.00%) | 10.0(0.87%) | 74.0(2.40%) | 197.0(5.21%) | 401.0(9.46%) | 454.0(15.62%) | 98.0(18.24%) |
| Mild renal dysfunction (60-90) | 2821.0(17.63%) | 21.0(6.44%) | 77.0(6.77%) | 395.0(12.82%) | 668.0(17.70%) | 967.0(22.81%) | 603.0(20.75%) | 90.0(16.75%) |
| Kidney damage with normal   GFR (>90) | 3021.0(18.88%) | 122.0(37.42%) | 409.0(35.97%) | 950.0(30.83%) | 874.0(23.15%) | 533.0(12.57%) | 125.0(4.30%) | 8.0(1.48%) |
| Alkaline phosphatase, U/L | 95.0(73.0-133.0);n=3881 | 80.0(58.5-124.0);n=92 | 89.0(69.0-127.0);n=366 | 97.0(73.0-137.5);n=923 | 96.0(74.0-134.0);n=961 | 95.0(73.0-131.0);n=1037 | 96.5(75.0-132.0);n=444 | 89.5(76.0-135.0);n=58 |
| Aspartate transaminase, U/L | 38.0(26.0-66.0);n=1509 | 39.5(27.0-75.0);n=44 | 35.0(27.0-57.0);n=155 | 41.0(28.0-71.0);n=369 | 40.0(27.0-65.0);n=361 | 41.0(26.0-72.0);n=396 | 30.0(21.0-53.0);n=165 | 35.0(22.0-90.0);n=19 |
| Alanine transaminase, U/L | 30.0(18.0-53.0);n=3251 | 25.5(16.5-43.5);n=78 | 32.0(20.0-51.0);n=309 | 34.0(22.0-62.0);n=764 | 33.0(21.0-61.0);n=835 | 29.0(17.0-51.0);n=889 | 19.0(13.0-33.0);n=337 | 19.0(12.0-30.0);n=39 |
| Bilirubin, umol/L | 13.0(8.32-20.0);n=3881 | 11.0(8.0-16.6);n=92 | 11.0(6.91-16.6);n=366 | 12.1(8.0-19.74);n=923 | 14.5(9.5-22.0);n=961 | 13.0(9.0-20.8);n=1037 | 11.95(8.0-18.0);n=444 | 13.0(7.75-20.05);n=58 |
| Triglyceride, mmol/L | 1.09(0.81-1.52);n=1869 | 0.97(0.78-1.35);n=11 | 1.16(0.78-1.63);n=88 | 1.1(0.78-1.69);n=334 | 1.04(0.82-1.5);n=451 | 1.1(0.83-1.51);n=620 | 1.1(0.81-1.43);n=331 | 1.06(0.69-1.63);n=34 |
| Low-density lipoprotein, mmol/L | 2.65(2.04-3.35);n=1790 | 2.66(2.28-3.62);n=10 | 2.75(2.18-3.27);n=83 | 2.76(2.19-3.59);n=318 | 2.61(2.05-3.33);n=431 | 2.66(2.02-3.33);n=596 | 2.5(1.98-3.2);n=319 | 2.84(2.09-3.24);n=33 |
| High-density lipoprotein, mmol/L | 1.15(0.9-1.43);n=1805 | 1.06(0.77-1.42);n=10 | 1.15(0.9-1.4);n=87 | 1.16(0.9-1.45);n=321 | 1.14(0.91-1.41);n=433 | 1.19(0.94-1.44);n=600 | 1.1(0.86-1.43);n=321 | 1.22(0.93-1.43);n=33 |
| Total cholesterol, mmol/L | 4.44(3.7-5.27);n=1876 | 4.69(3.72-5.82);n=12 | 4.56(3.85-5.52);n=88 | 4.68(3.98-5.5);n=337 | 4.37(3.7-5.2);n=453 | 4.45(3.7-5.26);n=620 | 4.3(3.55-5.1);n=332 | 4.34(3.8-5.35);n=34 |
| HbA1C, % | 6.6(6.0-7.95);n=275 | 14.3(14.3-14.3);n=1 | 6.25(5.8-6.75);n=10 | 6.7(6.0-8.7);n=49 | 6.65(6.0-7.8);n=74 | 7.0(6.15-8.65);n=88 | 6.4(6.0-7.2);n=45 | 5.9(4.75-6.25);n=8 |
| Fasting glucose, mmol/L | 6.1(5.2-7.9);n=1595 | 5.3(4.7-6.9);n=25 | 5.6(5.0-6.61);n=100 | 6.18(5.22-7.9);n=274 | 6.3(5.4-8.3);n=359 | 6.2(5.3-7.8);n=474 | 6.1(5.18-7.82);n=312 | 5.8(5.15-7.66);n=51 |

### Supplementary Table 18. Univariable Cox regression models to identify significant risk predictors of secondary outcomes

* for p≤ 0.05, ** for p ≤ 0.01, *** for p ≤ 0.001. Abbreviations: HR (hazard ratio); CCI (Charlson Comorbidity Index); COPD (chronic obstructive pulmonary disease); ACEI (angiotensin-converting enzyme inhibitors); ARB (Angiotensin Receptor Blockers); NSAIDs (non-steroidal anti-inflammatory drugs); AST (aspartate transaminase); APRI (AST to platelet ratio index); ALT (alanine transaminase); HbA1C (glycated haemoglobin).

| **Characteristics** | **All-cause mortality  HR [95% CI]; P value** | **Cancer-related mortality  HR [95% CI]; P value** | **Non-cancer-related mortality  HR [95% CI]; P value** |
| --- | --- | --- | --- |
| Gender | | | |
| Male | 0.84[0.81-0.88];<0.0001*** | 0.81[0.77-0.84];<0.0001*** | 1.03[0.94-1.12];0.5458 |
| Female | 1.18[1.14-1.23];<0.0001*** | 1.24[1.19-1.29];<0.0001*** | 0.97[0.90-1.06];0.5458 |
| Baseline age, years | 1.021[1.020-1.023];<0.0001*** | 1.02[1.01-1.02];<0.0001*** | 1.05[1.04-1.05];<0.0001*** |
| 18-40 | 0.64[0.56-0.72];<0.0001*** | 0.68[0.59-0.78];<0.0001*** | 0.48[0.35-0.66];<0.0001*** |
| 40-50 | 0.85[0.79-0.91];<0.0001*** | 0.94[0.88-1.01];0.1110 | 0.49[0.40-0.59];<0.0001*** |
| 50-60 | 0.76[0.73-0.80];<0.0001*** | 0.83[0.79-0.87];<0.0001*** | 0.51[0.46-0.58];<0.0001*** |
| 60-70 | 0.79[0.76-0.82];<0.0001*** | 0.79[0.76-0.83];<0.0001*** | 0.77[0.70-0.84];<0.0001*** |
| 70-80 | 1.16[1.11-1.20];<0.0001*** | 1.11[1.06-1.16];<0.0001*** | 1.36[1.24-1.48];<0.0001*** |
| 80-90 | 1.76[1.68-1.85];<0.0001*** | 1.62[1.54-1.71];<0.0001*** | 2.43[2.20-2.67];<0.0001*** |
| >90 | 2.58[2.32-2.86];<0.0001*** | 2.07[1.82-2.35];<0.0001*** | 4.96[4.13-5.95];<0.0001*** |
| CCI | 1.12[1.11-1.12];<0.0001*** | 1.12[1.12-1.13];<0.0001*** | 1.10[1.09-1.11];<0.0001*** |
| Charlson 0 | 0.48[0.42-0.55];<0.0001*** | 0.53[0.46-0.61];<0.0001*** | 0.29[0.21-0.42];<0.0001*** |
| Charlson 1-5 | 0.49[0.47-0.51];<0.0001*** | 0.45[0.43-0.47];<0.0001*** | 0.67[0.62-0.73];<0.0001*** |
| Charlson 6-10 | 1.26[1.21-1.30];<0.0001*** | 1.33[1.28-1.39];<0.0001*** | 0.99[0.91-1.07];0.7598 |
| Charlson 11+ | 2.05[1.97-2.14];<0.0001*** | 2.02[1.94-2.12];<0.0001*** | 2.19[2.00-2.39];<0.0001*** |
| History Disease | | | |
| Diabetes mellitus | 1.17[1.13-1.21];<0.0001*** | 1.19[1.14-1.23];<0.0001*** | 1.09[1.01-1.18];0.0287* |
| Hypertension | 1.10[1.06-1.14];<0.0001*** | 1.00[0.96-1.04];0.9332 | 1.60[1.47-1.73];<0.0001*** |
| Hyperlipidaemia | 1.02[0.94-1.11];0.6050 | 0.99[0.90-1.09];0.8954 | 1.13[0.95-1.35];0.1616 |
| Heart failure | 1.92[1.76-2.08];<0.0001*** | 1.65[1.50-1.82];<0.0001*** | 3.14[2.69-3.68];<0.0001*** |
| Atrial fibrillation | 2.54[1.91-3.39];<0.0001*** | 2.13[1.50-3.01];<0.0001*** | 4.30[2.59-7.15];<0.0001*** |
| Acute myocardial infarction | 1.63[1.40-1.89];<0.0001*** | 1.38[1.15-1.66];0.0005*** | 2.67[2.04-3.51];<0.0001*** |
| Stroke/TIA | 1.66[1.55-1.78];<0.0001*** | 1.47[1.35-1.60];<0.0001*** | 2.50[2.18-2.87];<0.0001*** |
| Peripheral vascular disease | 1.27[0.85-1.90];0.2389 | 1.31[0.84-2.03];0.2291 | 1.11[0.42-2.97];0.8279 |
| Ischemic heart disease | 1.35[1.26-1.44];<0.0001*** | 1.19[1.09-1.29];<0.0001*** | 2.05[1.80-2.34];<0.0001*** |
| COPD | 1.43[1.33-1.53];<0.0001*** | 1.27[1.17-1.38];<0.0001*** | 2.12[1.85-2.43];<0.0001*** |
| Renal diseases | 1.34[1.28-1.41];<0.0001*** | 1.23[1.16-1.30];<0.0001*** | 1.86[1.69-2.05];<0.0001*** |
| Mild liver disease | 0.88[0.85-0.91];<0.0001*** | 0.74[0.71-0.77];<0.0001*** | 1.87[1.72-2.03];<0.0001*** |
| Moderate or severe liver | 1.35[1.30-1.40];<0.0001*** | 1.71[1.65-1.79];<0.0001*** | 0.29[0.25-0.33];<0.0001*** |
| Gastrointestinal bleeding | 1.33[1.25-1.42];<0.0001*** | 1.27[1.18-1.36];<0.0001*** | 1.60[1.40-1.83];<0.0001*** |
| Anaemia | 1.68[1.60-1.77];<0.0001*** | 1.61[1.52-1.70];<0.0001*** | 2.03[1.82-2.26];<0.0001*** |
| Hip fractures | 1.57[1.42-1.74];<0.0001*** | 1.45[1.29-1.63];<0.0001*** | 2.10[1.72-2.58];<0.0001*** |
| Accident fall | 1.44[1.35-1.54];<0.0001*** | 1.30[1.21-1.40];<0.0001*** | 2.06[1.82-2.34];<0.0001*** |
| Alcoholism | 0.99[0.93-1.05];0.7095 | 0.94[0.87-1.01];0.0689 | 1.20[1.06-1.36];0.0051** |
| Obesity | 1.13[1.02-1.25];0.0206* | 1.09[0.97-1.23];0.1464 | 1.29[1.04-1.60];0.0231* |
| Prior cancer | 1.47[1.41-1.52];<0.0001*** | 1.58[1.51-1.64];<0.0001*** | 1.07[0.98-1.17];0.1313 |
| Colorectal cancer | 1.08[1.04-1.13];0.0005*** | 1.11[1.06-1.17];<0.0001*** | 0.97[0.87-1.08];0.5410 |
| Lung cancer | 1.94[1.81-2.09];<0.0001*** | 2.04[1.89-2.21];<0.0001*** | 1.50[1.24-1.81];<0.0001*** |
| Pancreas cancer | 1.98[1.77-2.21];<0.0001*** | 2.15[1.91-2.42];<0.0001*** | 1.26[0.92-1.73];0.1484 |
| Bladder cancer | 1.78[1.45-2.18];<0.0001*** | 1.79[1.43-2.24];<0.0001*** | 1.75[1.08-2.81];0.0221* |
| Gastric cancer | 1.75[1.55-1.98];<0.0001*** | 1.81[1.59-2.06];<0.0001*** | 1.50[1.11-2.04];0.0080** |
| Prostate cancer | 1.26[1.07-1.49];0.0059** | 1.19[0.98-1.44];0.0742 | 1.58[1.12-2.23];0.0089** |
| Nasopharyngeal carcinoma | 1.27[1.11-1.46];0.0006*** | 1.33[1.15-1.55];0.0002*** | 1.02[0.72-1.45];0.9108 |
| Head neck cancer | 1.37[1.21-1.54];<0.0001*** | 1.40[1.23-1.60];<0.0001*** | 1.22[0.92-1.63];0.1689 |
| Breast cancer | 1.36[1.26-1.47];<0.0001*** | 1.48[1.36-1.62];<0.0001*** | 0.84[0.67-1.06];0.1365 |
| Ovarian cancer | 1.18[0.97-1.43];0.1022 | 1.36[1.11-1.67];0.0029** | 0.42[0.20-0.88];0.0222* |
| Cervical cancer | 1.84[1.36-2.50];0.0001*** | 2.17[1.59-2.98];<0.0001*** | 0.46[0.12-1.85];0.2757 |
| Number of prescribed drug classes | 1.12[1.10-1.15];<0.0001*** | 1.07[1.05-1.10];<0.0001*** | 1.32[1.26-1.39];<0.0001*** |
| Drug class number 0 | 0.84[0.81-0.88];<0.0001*** | 0.90[0.86-0.95];0.0001*** | 0.63[0.56-0.70];<0.0001*** |
| Drug class number 1 | 1.01[0.98-1.05];0.4584 | 1.01[0.97-1.06];0.4889 | 1.01[0.93-1.10];0.7812 |
| Drug class number 2+ | 1.16[1.11-1.21];<0.0001*** | 1.09[1.03-1.14];0.0008*** | 1.49[1.36-1.63];<0.0001*** |
| Medications | | | |
| ACEI/ARB | 0.95[0.90-0.99];0.0272* | 0.86[0.81-0.91];<0.0001*** | 1.31[1.19-1.45];<0.0001*** |
| Calcium channel blockers | 1.05[0.99-1.12];0.1100 | 1.03[0.96-1.10];0.4651 | 1.16[1.01-1.32];0.0348* |
| Alpha-blockers | 1.03[0.92-1.15];0.6445 | 0.98[0.86-1.12];0.7460 | 1.22[0.97-1.54];0.0953 |
| Beta-blockers | 0.89[0.83-0.95];0.0007*** | 0.84[0.78-0.90];<0.0001*** | 1.11[0.97-1.27];0.1137 |
| Diuretics for hypertension | 0.99[0.87-1.12];0.8607 | 1.01[0.88-1.16];0.8731 | 0.90[0.67-1.20];0.4776 |
| Diuretics for heart failure | 1.47[1.39-1.54];<0.0001*** | 1.54[1.46-1.63];<0.0001*** | 1.18[1.04-1.33];0.0120* |
| Lipid-lowering drugs | 1.01[0.90-1.14];0.8075 | 0.97[0.85-1.10];0.6110 | 1.20[0.95-1.52];0.1230 |
| Statins and fibrates | 1.00[0.88-1.14];0.9548 | 0.95[0.82-1.10];0.5279 | 1.20[0.92-1.55];0.1726 |
| Nitrates | 1.31[1.12-1.53];0.0008*** | 1.31[1.10-1.56];0.0025** | 1.31[0.91-1.88];0.1422 |
| Antibiotics | 0.83[0.58-1.18];0.2953 | 0.77[0.51-1.16];0.2091 | 1.06[0.53-2.13];0.8603 |
| Anticoagulants | 1.22[1.06-1.40];0.0044** | 1.17[1.00-1.37];0.0474* | 1.42[1.06-1.89];0.0168* |
| Eye drugs | 1.09[0.99-1.19];0.0751 | 1.07[0.97-1.19];0.1950 | 1.15[0.94-1.41];0.1632 |
| Antiestrogen | 1.15[1.05-1.27];0.0042** | 1.27[1.14-1.41];<0.0001*** | 0.68[0.51-0.91];0.0084** |
| Antiplatelets | 1.27[1.16-1.40];<0.0001*** | 1.18[1.06-1.32];0.0022** | 1.64[1.36-1.98];<0.0001*** |
| Anti-diabetic drugs | 1.09[1.05-1.14];0.0001*** | 1.01[0.96-1.06];0.7118 | 1.46[1.33-1.60];<0.0001*** |
| Hormonal drugs | 1.27[1.11-1.45];0.0006*** | 1.17[1.00-1.37];0.0508 | 1.68[1.28-2.19];0.0002*** |
| Steroids | 1.20[1.03-1.40];0.0203* | 1.07[0.89-1.28];0.4890 | 1.75[1.31-2.33];0.0002*** |
| NSAIDs | 1.27[1.15-1.40];<0.0001*** | 1.17[1.05-1.31];0.0051** | 1.68[1.39-2.03];<0.0001*** |
| Anti-cancer drugs | 0.90[0.86-0.95];0.0001*** | 0.96[0.91-1.02];0.1869 | 0.66[0.58-0.75];<0.0001*** |
| Tyrosine kinase inhibitor | 1.03[0.95-1.12];0.4370 | 1.04[0.95-1.15];0.3694 | 0.99[0.82-1.21];0.9507 |
| Endocrine therapy | 1.18[1.09-1.29];0.0001*** | 1.29[1.18-1.41];<0.0001*** | 0.76[0.60-0.95];0.0182* |
| Aromatase inhibitors | 1.20[1.08-1.33];0.0008*** | 1.33[1.19-1.49];<0.0001*** | 0.63[0.45-0.87];0.0058** |
| Endocrine therapy excluding AIs | 1.15[1.01-1.31];0.0318* | 1.20[1.04-1.38];0.0105* | 0.95[0.69-1.30];0.7329 |
| Cytotoxic therapy | 0.79[0.73-0.85];<0.0001*** | 0.80[0.74-0.87];<0.0001*** | 0.73[0.62-0.86];0.0003*** |
| Androgen deprivation therapy | 0.75[0.60-0.93];0.0083** | 0.70[0.54-0.89];0.0045** | 0.95[0.63-1.45];0.8152 |
| Targeted therapy | 0.77[0.51-1.17];0.2275 | 0.74[0.46-1.20];0.2224 | 0.89[0.37-2.15];0.8009 |
| Monoclonal antibodies | 0.86[0.79-0.94];0.0006*** | 0.95[0.86-1.04];0.2534 | 0.50[0.39-0.65];<0.0001*** |
| Immunomodulatory drugs | 1.07[0.75-1.53];0.7071 | 1.10[0.74-1.63];0.6342 | 0.95[0.39-2.28];0.9039 |
| Tamoxifen | 1.15[1.01-1.31];0.0318* | 1.20[1.04-1.38];0.0105* | 0.95[0.69-1.30];0.7329 |
| Anti-HER2 monoclonal antibody | 0.92[0.76-1.12];0.4041 | 0.99[0.80-1.21];0.8976 | 0.66[0.40-1.10];0.1082 |
| Anthracycline | 0.87[0.78-0.96];0.0090** | 0.93[0.83-1.05];0.2413 | 0.60[0.45-0.80];0.0005*** |
| Antimetabolite fluoropyrimidines | 0.48[0.33-0.71];0.0002*** | 0.44[0.28-0.69];0.0004*** | 0.63[0.30-1.33];0.2258 |
| Microtubule inhibitor | 1.56[1.15-2.10];0.0038** | 1.60[1.15-2.22];0.0050** | 1.37[0.65-2.88];0.4036 |
| Platinum | 0.72[0.64-0.80];<0.0001*** | 0.69[0.61-0.78];<0.0001*** | 0.82[0.65-1.03];0.0814 |
| GnRH agonist | 0.63[0.49-0.81];0.0004*** | 0.56[0.41-0.76];0.0002*** | 0.90[0.57-1.43];0.6591 |
| Immune checkpoint inhibitors | 0.77[0.61-0.97];0.0263* | 0.73[0.56-0.95];0.0213* | 0.92[0.57-1.47];0.7161 |
| Laboratory examinations | | | |
| Fibrosis-4 (FIB-4) index | 0.99[0.98-1.01];0.3954 | 0.99[0.98-1.01];0.3008 | 1.00[0.97-1.03];0.8718 |
| APRI | 0.999[0.998-1.000];0.1404 | 0.999[0.998-1.000];0.0958 | 1.000[0.998-1.002];0.9797 |
| Albumin-Bilirubin (ALBI) score | 1.16[1.11-1.21];<0.0001*** | 1.13[1.08-1.19];<0.0001*** | 1.27[1.15-1.40];<0.0001*** |
| AST/ALT ratio | 1.05[1.02-1.08];0.0018** | 1.05[1.02-1.08];0.0024** | 1.03[0.96-1.11];0.3683 |
| ALT/PLT ratio | 0.999[0.998-0.999];0.0006*** | 0.998[0.998-0.999];0.0006*** | 0.999[0.998-1.001];0.4384 |
| Urea-to-Creatinine ratio | 1.002[1.001-1.004];0.0048** | 1.002[1.001-1.004];0.0027** | 1.000[0.997-1.004];0.7928 |
| Mean corpuscular volume, fL | 1.00[0.99-1.00];0.3367 | 1.00[0.99-1.00];0.1796 | 1.00[0.99-1.02];0.5726 |
| Basophil, x10^9/L | 1.09[0.95-1.25];0.2168 | 1.08[0.93-1.26];0.2944 | 1.11[0.81-1.52];0.5030 |
| Eosinophil, x10^9/L | 1.03[1.00-1.05];0.0641 | 1.02[0.99-1.05];0.2267 | 1.05[0.99-1.12];0.0761 |
| Lymphocyte, x10^9/L | 1.00[1.00-1.01];0.0280* | 1.01[1.00-1.01];0.0292* | 1.00[0.99-1.01];0.5959 |
| Blast, x10^9/L | 0.02[0.00-4.67];0.1600 | 0.03[0.00-9.39];0.2300 | 0.00[0.00-17840.58];0.4343 |
| Metamyelocyte, x10^9/L | 1.21[0.03-47.32];0.9188 | 1.21[0.03-47.32];0.9188 | - |
| Monocyte, x10^9/L | 1.02[1.01-1.03];0.0003*** | 1.02[1.01-1.04];0.0004*** | 1.01[0.99-1.04];0.3139 |
| Neutrophil, x10^9/L | 1.00[1.00-1.01];0.0001*** | 1.00[1.00-1.01];0.0005*** | 1.00[1.00-1.01];0.0732 |
| White blood bount, x10^9/L | 1.01[1.00-1.02];0.1464 | 1.01[1.00-1.02];0.1135 | 1.00[0.96-1.04];0.9661 |
| Mean cell haemoglobin, pg | 0.99[0.97-1.00];0.1414 | 0.98[0.97-1.00];0.0656 | 1.01[0.97-1.05];0.6677 |
| Myelocyte, x10^9/L | 0.93[0.50-1.71];0.8117 | 0.99[0.54-1.80];0.9622 | 0.00[0.00-Inf];0.9942 |
| Platelet, x10^9/L | 1.001[1.001-1.002];<0.0001*** | 1.002[1.001-1.002];<0.0001*** | 0.999[0.998-1.001];0.2576 |
| Reticulocyte, x10^9/L | 1.00[0.99-1.02];0.7449 | 0.99[0.97-1.02];0.6507 | 1.02[1.00-1.05];0.1111 |
| Red blood count, x10^12/L | 0.79[0.72-0.86];<0.0001*** | 0.81[0.73-0.90];<0.0001*** | 0.70[0.57-0.87];0.0010** |
| Hematocrit, L/L | 0.03[0.01-0.10];<0.0001*** | 0.03[0.01-0.12];<0.0001*** | 0.03[0.00-0.38];0.0068** |
| K/Potassium, mmol/L | 0.99[0.92-1.07];0.7904 | 0.98[0.91-1.07];0.6851 | 1.02[0.86-1.21];0.8209 |
| Urate, mmol/L | 1.17[0.45-3.04];0.7402 | 0.75[0.25-2.25];0.6079 | 4.96[0.78-31.51];0.0894 |
| Albumin, g/L | 0.97[0.97-0.98];<0.0001*** | 0.97[0.97-0.98];<0.0001*** | 0.96[0.95-0.97];<0.0001*** |
| Na/Sodium, mmol/L | 0.96[0.95-0.97];<0.0001*** | 0.96[0.95-0.97];<0.0001*** | 0.97[0.95-0.99];0.0055** |
| Urea, mmol/L | 1.03[1.02-1.04];<0.0001*** | 1.02[1.00-1.03];0.0164* | 1.08[1.06-1.11];<0.0001*** |
| Protein, g/L | 0.99[0.99-1.00];0.0004*** | 0.99[0.99-1.00];0.0007*** | 0.99[0.98-1.00];0.2702 |
| Creatinine, umol/L | 1.001[1.001-1.002];<0.0001*** | 1.001[1.000-1.001];0.0115* | 1.003[1.002-1.003];<0.0001*** |
| CKD-EPI Creatinine  Equation (2021) | 0.992[0.991-0.993];<0.0001*** | 0.99[0.99-1.00];<0.0001*** | 0.982[0.980-0.984];<0.0001*** |
| Kidney failure (<15) | 2.21[1.80-2.70];<0.0001*** | 1.63[1.26-2.12];0.0002*** | 4.70[3.40-6.49];<0.0001*** |
| Severe dysfunction (15-30) | 2.12[1.82-2.47];<0.0001*** | 1.86[1.56-2.23];<0.0001*** | 3.18[2.39-4.22];<0.0001*** |
| Moderate dysfunction (30-60) | 1.37[1.28-1.47];<0.0001*** | 1.25[1.16-1.36];<0.0001*** | 1.86[1.62-2.13];<0.0001*** |
| Mild renal dysfunction (60-90) | 0.98[0.93-1.04];0.5312 | 0.98[0.92-1.03];0.4066 | 1.01[0.91-1.14];0.8047 |
| Kidney damage with normal   GFR (>90) | 0.79[0.75-0.83];<0.0001*** | 0.86[0.81-0.91];<0.0001*** | 0.56[0.50-0.63];<0.0001*** |
| Alkaline phosphatase, U/L | 1.002[1.001-1.002];<0.0001*** | 1.002[1.001-1.002];<0.0001*** | 1.001[1.001-1.002];0.0009*** |
| Aspartate transaminase, U/L | 1.000[0.999-1.001];0.9049 | 1.000[0.999-1.001];0.8185 | 0.999[0.998-1.001];0.4654 |
| Alanine transaminase, U/L | 0.999[0.998-0.999];<0.0001*** | 0.999[0.998-0.999];<0.0001*** | 0.999[0.997-1.000];0.0319* |
| Bilirubin, umol/L | 1.003[1.001-1.005];0.0004*** | 1.003[1.001-1.005];0.0013** | 1.00[1.00-1.01];0.1493 |
| Triglyceride, mmol/L | 1.01[0.95-1.08];0.7911 | 1.01[0.93-1.08];0.8773 | 1.02[0.89-1.17];0.7859 |
| Low-density lipoprotein, mmol/L | 1.00[0.96-1.05];0.8922 | 1.02[0.96-1.07];0.5503 | 0.96[0.86-1.06];0.4040 |
| High-density lipoprotein, mmol/L | 0.74[0.65-0.84];<0.0001*** | 0.77[0.67-0.89];0.0005*** | 0.65[0.50-0.86];0.0021** |
| Total cholesterol, mmol/L | 0.98[0.94-1.02];0.3130 | 0.99[0.94-1.04];0.7184 | 0.93[0.85-1.02];0.1414 |
| HbA1C, % | 1.06[0.99-1.14];0.0975 | 1.08[0.99-1.17];0.0688 | 1.01[0.85-1.19];0.9426 |
| Fasting glucose, mmol/L | 1.00[0.98-1.01];0.5892 | 0.99[0.97-1.01];0.5558 | 1.00[0.96-1.04];0.9756 |

### Supplementary Table 19. Descriptive statistics of patient’s outcomes and characteristics by prior cancer v.s. no prior cancer upon diagnosis of HCC.

^ P value indicates statistical difference among patients of two subgroups. * for p≤ 0.05, ** for p ≤ 0.01, *** for p ≤ 0.001. Abbreviations: HR (hazard ratio); HCC (Hepatocellular carcinoma); CCI (Charlson Comorbidity Index); COPD (chronic obstructive pulmonary disease); ACEI (angiotensin-converting enzyme inhibitors); ARB (Angiotensin Receptor Blockers); NSAIDs (Non-steroidal anti-inflammatory drugs); AST (aspartate transaminase); APRI (AST to platelet ratio index); ALT (alanine transaminase); HbA1C (glycated haemoglobin).

| **Characteristics** | **Prior cancer (N=5996)  Median (IQR); N or Count (%)** | **No Prior cancer (N=10020)  Median (IQR); N or Count (%)** | **P value^** |
| --- | --- | --- | --- |
| ***Outcomes*** | | | |
| HCC recurrence | 4561(76.06%) | 8010(80.08%) | 0.0385* |
| Time from HCC to HCC recurrence, years | 0.54(0.06-4.59);n=5996 | 0.89(0.07-4.8);n=10002 | <0.0001*** |
| <1 year | 3264(54.43%) | 5069(50.67%) | 0.0104* |
| 1-2 years | 794(13.24%) | 1296(12.95%) | 0.6676 |
| 2-4 years | 364(6.07%) | 897(8.96%) | <0.0001*** |
| >=4 years | 139(2.31%) | 748(7.47%) | <0.0001*** |
| All-cause mortality | 5672(94.59%) | 8759(87.57%) | 0.0011** |
| Time to mortality, years | 0.34(0.05-1.36);n=5996 | 0.73(0.11-3.28);n=10002 | <0.0001*** |
| Within 30 days | 1805(30.10%) | 2183(21.82%) | <0.0001*** |
| Within 30-90 days | 893(14.89%) | 1307(13.06%) | 0.0051** |
| Within 90-180 days | 633(10.55%) | 956(9.55%) | 0.0685 |
| Within 0.5-1 year | 768(12.80%) | 1059(10.58%) | 0.0002*** |
| Within 1-1.5 years | 554(9.23%) | 759(7.58%) | 0.0008*** |
| Within 1.5-3 years | 648(10.80%) | 1114(11.13%) | 0.5802 |
| Within 3-5 years | 227(3.78%) | 652(6.51%) | <0.0001*** |
| Within 5-10 years | 116(1.93%) | 557(5.56%) | <0.0001*** |
| Within 10+ years | 28(0.46%) | 172(1.71%) | <0.0001*** |
| Cancer-related mortality | 4824(80.45%) | 6791(67.89%) | <0.0001*** |
| Time to cancer mortality, years | 0.34(0.05-1.36);n=5996 | 0.73(0.11-3.28);n=10002 | <0.0001*** |
| Within 30 days | 1564(26.08%) | 1771(17.70%) | <0.0001*** |
| Within 30-90 days | 770(12.84%) | 1069(10.68%) | 0.0003*** |
| Within 90-180 days | 547(9.12%) | 757(7.56%) | 0.0015** |
| Within 0.5-1 year | 652(10.87%) | 858(8.57%) | <0.0001*** |
| Within 1-1.5 years | 478(7.97%) | 603(6.02%) | <0.0001*** |
| Within 1.5-3 years | 551(9.18%) | 838(8.37%) | 0.1127 |
| Within 3-5 years | 178(2.96%) | 454(4.53%) | <0.0001*** |
| Within 5-10 years | 76(1.26%) | 352(3.51%) | <0.0001*** |
| Within 10+ years | 8(0.13%) | 89(0.88%) | <0.0001*** |
| Non-cancer-related mortality | 848(14.14%) | 1968(19.67%) | <0.0001*** |
| Time to non-cancer mortality, years | 0.34(0.05-1.36);n=5996 | 0.73(0.11-3.28);n=10002 | <0.0001*** |
| Within 30 days | 241(4.01%) | 412(4.11%) | 0.7985 |
| Within 30-90 days | 123(2.05%) | 238(2.37%) | 0.2047 |
| Within 90-180 days | 86(1.43%) | 199(1.98%) | 0.0137* |
| Within 0.5-1 year | 116(1.93%) | 201(2.00%) | 0.7915 |
| Within 1-1.5 years | 76(1.26%) | 156(1.55%) | 0.1594 |
| Within 1.5-3 years | 97(1.61%) | 276(2.75%) | <0.0001*** |
| Within 3-5 years | 49(0.81%) | 198(1.97%) | <0.0001*** |
| Within 5-10 years | 40(0.66%) | 205(2.04%) | <0.0001*** |
| Within 10+ years | 20(0.33%) | 83(0.82%) | 0.0002*** |
| Liver cancer-related mortality | 184(3.06%) | 3781(37.80%) | <0.0001*** |
| Time to liver cancer mortality, years | 0.34(0.05-1.36);n=5996 | 0.73(0.11-3.28);n=10002 | <0.0001*** |
| Within 30 days | 50(0.83%) | 841(8.40%) | <0.0001*** |
| Within 30-90 days | 33(0.55%) | 503(5.02%) | <0.0001*** |
| Within 90-180 days | 16(0.26%) | 371(3.70%) | <0.0001*** |
| Within 0.5-1 year | 17(0.28%) | 486(4.85%) | <0.0001*** |
| Within 1-1.5 years | 25(0.41%) | 348(3.47%) | <0.0001*** |
| Within 1.5-3 years | 18(0.30%) | 569(5.68%) | <0.0001*** |
| Within 3-5 years | 14(0.23%) | 334(3.33%) | <0.0001*** |
| Within 5-10 years | 10(0.16%) | 258(2.57%) | <0.0001*** |
| Within 10+ years | 1(0.01%) | 71(0.70%) | <0.0001*** |
| ***Patient characteristics*** | | | |
| Gender |  |  |  |
| Male gender | 3215(53.61%) | 6822(68.20%) | <0.0001*** |
| Female gender | 2781(46.38%) | 3180(31.79%) | <0.0001*** |
| Baseline age, years | 69.84(58.67-79.46);n=5996 | 68.69(58.74-78.18);n=10002 | 0.0019** |
| 18-40 | 115(1.91%) | 211(2.10%) | 0.4496 |
| 40-50 | 451(7.52%) | 686(6.85%) | 0.1503 |
| 50-60 | 1130(18.84%) | 1951(19.50%) | 0.4101 |
| 60-70 | 1325(22.09%) | 2449(24.48%) | 0.0070** |
| 70-80 | 1572(26.21%) | 2666(26.65%) | 0.6565 |
| 80-90 | 1209(20.16%) | 1696(16.95%) | <0.0001*** |
| >90 | 194(3.23%) | 343(3.42%) | 0.5539 |
| CCI | 10.0(9.0-12.0);n=5996 | 6.0(3.0-10.0);n=10002 | <0.0001*** |
| Charlson 0 | 1(0.01%) | 342(3.41%) | <0.0001*** |
| Charlson 1-5 | 121(2.01%) | 4257(42.56%) | <0.0001*** |
| Charlson 6-10 | 2896(48.29%) | 3768(37.67%) | <0.0001*** |
| Charlson 11+ | 2978(49.66%) | 1635(16.34%) | <0.0001*** |
| History Disease |  |  |  |
| Diabetes mellitus | 2147(35.80%) | 4752(47.51%) | <0.0001*** |
| Hypertension | 2371(39.54%) | 3786(37.85%) | 0.1614 |
| Hyperlipidaemia | 301(5.02%) | 470(4.69%) | 0.4032 |
| Heart failure | 325(5.42%) | 471(4.70%) | 0.062 |
| Atrial fibrillation | 147(2.45%) | 143(1.42%) | <0.0001*** |
| Acute myocardial infarction | 126(2.10%) | 125(1.24%) | <0.0001*** |
| Stroke/TIA | 448(7.47%) | 653(6.52%) | 0.0362* |
| Peripheral vascular disease | 11(0.18%) | 20(0.19%) | 0.9652 |
| Ischemic heart disease | 495(8.25%) | 688(6.87%) | 0.0031** |
| COPD | 436(7.27%) | 654(6.53%) | 0.1032 |
| Renal diseases | 1066(17.77%) | 1579(15.78%) | 0.0059** |
| Mild liver disease | 3378(56.33%) | 4946(49.45%) | <0.0001*** |
| Moderate or severe liver | 1702(28.38%) | 2582(25.81%) | 0.0074** |
| Gastrointestinal bleeding | 701(11.69%) | 744(7.43%) | <0.0001*** |
| Anaemia | 1161(19.36%) | 1228(12.27%) | <0.0001*** |
| Hip fractures | 213(3.55%) | 309(3.08%) | 0.1343 |
| Accident fall | 565(9.42%) | 842(8.41%) | 0.0503 |
| Alcoholism | 414(6.90%) | 1040(10.39%) | <0.0001*** |
| Obesity | 173(2.88%) | 316(3.15%) | 0.3689 |
| Prior cancer | 5996(100.00%) | 0(0.00%) | - |
| Colorectal cancer | 2744(45.76%) | 0(0.00%) | - |
| Lung cancer | 1074(17.91%) | 0(0.00%) | - |
| Pancreas cancer | 463(7.72%) | 0(0.00%) | - |
| Bladder cancer | 139(2.31%) | 0(0.00%) | - |
| Gastric cancer | 358(5.97%) | 0(0.00%) | - |
| Prostate cancer | 195(3.25%) | 0(0.00%) | - |
| Nasopharyngeal carcinoma | 260(4.33%) | 0(0.00%) | - |
| Head neck cancer | 358(5.97%) | 0(0.00%) | - |
| Breast cancer | 777(12.95%) | 0(0.00%) | - |
| Ovarian cancer | 135(2.25%) | 0(0.00%) | - |
| Cervical cancer | 63(1.05%) | 0(0.00%) | - |
| Number of prescribed drug classes | 1.0(1.0-2.0);n=5996 | 1.0(1.0-1.0);n=10002 | <0.0001*** |
| Drug class number 0 | 1172(19.54%) | 2195(21.94%) | 0.0038** |
| Drug class number 1 | 3274(54.60%) | 5785(57.83%) | 0.0361* |
| Drug class number 2+ | 1550(25.85%) | 2022(20.21%) | <0.0001*** |
| Medications |  |  |  |
| ACEI/ARB | 948(15.81%) | 1491(14.90%) | 0.195 |
| Calcium channel blockers | 524(8.73%) | 906(9.05%) | 0.5501 |
| Alpha-blockers | 146(2.43%) | 245(2.44%) | 0.9973 |
| Beta-blockers | 318(5.30%) | 901(9.00%) | <0.0001*** |
| Diuretics for hypertension | 74(1.23%) | 253(2.52%) | <0.0001*** |
| Diuretics for heart failure | 493(8.22%) | 1561(15.60%) | <0.0001*** |
| Lipid-lowering drugs | 184(3.06%) | 248(2.47%) | 0.0345* |
| Statins and fibrates | 154(2.56%) | 185(1.84%) | 0.0034** |
| Nitrates | 65(1.08%) | 144(1.43%) | 0.0685 |
| Antibiotics | 7(0.11%) | 28(0.27%) | 0.05 |
| Anticoagulants | 116(1.93%) | 152(1.51%) | 0.0598 |
| Eye drugs | 292(4.86%) | 308(3.07%) | <0.0001*** |
| Antiestrogen | 370(6.17%) | 146(1.45%) | <0.0001*** |
| Antiplatelets | 225(3.75%) | 373(3.72%) | 0.9767 |
| Anti-diabetic drugs | 1205(20.09%) | 1907(19.06%) | 0.198 |
| Hormonal drugs | 115(1.91%) | 151(1.50%) | 0.0632 |
| Steroids | 89(1.48%) | 124(1.23%) | 0.2234 |
| NSAIDs | 207(3.45%) | 352(3.51%) | 0.864 |
| Anti-cancer drugs | 1220(20.34%) | 753(7.52%) | <0.0001*** |
| Tyrosine kinase inhibitor | 376(6.27%) | 302(3.01%) | <0.0001*** |
| Endocrine therapy | 501(8.35%) | 226(2.25%) | <0.0001*** |
| Aromatase inhibitors | 351(5.85%) | 83(0.82%) | <0.0001*** |
| Endocrine therapy excluding AIs | 150(2.50%) | 143(1.42%) | <0.0001*** |
| Cytotoxic therapy | 283(4.71%) | 612(6.11%) | 0.0005*** |
| Androgen deprivation therapy | 96(1.60%) | 27(0.26%) | <0.0001*** |
| Targerted therapy | 19(0.31%) | 7(0.06%) | 0.0004*** |
| Monoclonal antibodies | 445(7.42%) | 152(1.51%) | <0.0001*** |
| Immunomodulatory drugs | 3(0.05%) | 34(0.33%) | 0.0004*** |
| Tamoxifen | 150(2.50%) | 143(1.42%) | <0.0001*** |
| Anti-HER2 monoclonal antibody | 99(1.65%) | 26(0.25%) | <0.0001*** |
| Anthracycline | 154(2.56%) | 250(2.49%) | 0.8336 |
| Antimetabolite fluoropyrimidines | 27(0.45%) | 7(0.06%) | <0.0001*** |
| Microtubule inhibitor | 37(0.61%) | 19(0.18%) | <0.0001*** |
| Platinum | 57(0.95%) | 335(3.34%) | <0.0001*** |
| GnRH agonist | 68(1.13%) | 24(0.23%) | <0.0001*** |
| Immune checkpoint inhibitors | 18(0.30%) | 63(0.62%) | 0.0066** |
| Laboratory examinations |  |  |  |
| Fibrosis-4 (FIB-4) index | 1.75(1.02-2.94);n=164 | 3.9(2.27-7.71);n=374 | <0.0001*** |
| APRI | 12.9(7.97-23.06);n=191 | 40.0(22.0-85.06);n=411 | <0.0001*** |
| Albumin-Bilirubin (ALBI) score | -1.74(-2.16--1.19);n=1196 | -1.37(-1.88--0.76);n=2685 | <0.0001*** |
| AST/ALT ratio | 1.23(0.87-1.82);n=354 | 1.19(0.85-1.59);n=963 | 0.0245* |
| ALT/PLT ratio | 9.63(5.63-16.76);n=395 | 39.71(16.93-85.16);n=703 | <0.0001*** |
| Urea-to-Creatinine ratio | 65.15(51.68-81.28);n=1199 | 63.74(52.08-77.91);n=2630 | 0.2831 |
| Mean corpuscular volume, fL | 89.7(83.8-94.6);n=476 | 93.0(88.9-97.5);n=805 | <0.0001*** |
| Basophil, x10^9/L | 0.33(0.05-0.61);n=435 | 0.05(0.0-0.5);n=654 | <0.0001*** |
| Eosinophil, x10^9/L | 1.02(0.11-2.48);n=436 | 0.22(0.1-1.7);n=668 | <0.0001*** |
| Lymphocyte, x10^9/L | 16.6(1.92-28.0);n=437 | 2.1(1.1-21.25);n=670 | <0.0001*** |
| Blast, x10^9/L | 0.0(0.0-0.0);n=45 | 0.0(0.0-0.0);n=82 | 0.3034 |
| Metamyelocyte, x10^9/L | 0.17(0.13-0.28);n=3 | 0.11(0.08-0.39);n=3 | 1 |
| Monocyte, x10^9/L | 6.85(0.7-9.4);n=437 | 0.95(0.4-8.0);n=670 | <0.0001*** |
| Neutrophil, x10^9/L | 56.5(6.26-69.2);n=437 | 8.8(3.2-63.2);n=670 | <0.0001*** |
| White blood bount, x10^9/L | 6.2(4.72-7.76);n=476 | 5.8(4.31-7.8);n=805 | 0.0111* |
| Mean cell haemoglobin, pg | 30.7(28.15-32.8);n=476 | 32.4(30.4-33.8);n=805 | <0.0001*** |
| Myelocyte, x10^9/L | 0.23(0.14-1.5);n=7 | 0.36(0.04-0.84);n=4 | 0.3436 |
| Platelet, x10^9/L | 243.5(181.5-307.5);n=476 | 137.0(90.0-203.0);n=804 | <0.0001*** |
| Reticulocyte, x10^9/L | 1.74(1.58-2.68);n=11 | 2.24(1.37-12.26);n=22 | 0.6331 |
| Red blood count, x10^12/L | 4.01(3.62-4.41);n=476 | 4.04(3.59-4.49);n=805 | 0.5619 |
| Hematocrit, L/L | 0.35(0.32-0.39);n=401 | 0.38(0.34-0.41);n=723 | <0.0001*** |
| K/Potassium, mmol/L | 4.0(3.7-4.4);n=1198 | 4.0(3.7-4.38);n=2627 | 0.4086 |
| Urate, mmol/L | 0.3(0.24-0.36);n=74 | 0.34(0.27-0.41);n=319 | 0.0107* |
| Albumin, g/L | 38.0(33.0-42.0);n=1198 | 37.0(32.0-41.0);n=2687 | <0.0001*** |
| Na/Sodium, mmol/L | 139.0(137.0-141.7);n=1199 | 139.0(137.0-141.0);n=2631 | 0.4116 |
| Urea, mmol/L | 4.9(3.88-6.4);n=1199 | 5.2(4.2-6.8);n=2630 | <0.0001*** |
| Protein, g/L | 73.0(67.8-78.0);n=1037 | 74.0(69.0-79.0);n=2379 | <0.0001*** |
| Creatinine, umol/L | 76.0(61.0-98.0);n=2947 | 82.0(68.0-103.0);n=4547 | <0.0001*** |
| CKD-EPI Creatinine  Equation (2021) | 86.02(64.26-99.82);n=2947 | 83.95(62.59-97.06);n=4547 | <0.0001*** |
| Kidney failure (<15) | 61.0(1.01%) | 84.0(0.83%) | 0.2937 |
| Severe dysfunction (15-30) | 106.0(1.76%) | 167.0(1.66%) | 0.6942 |
| Moderate dysfunction (30-60) | 446.0(7.43%) | 788.0(7.87%) | 0.3655 |
| Mild renal dysfunction (60-90) | 1089.0(18.16%) | 1732.0(17.31%) | 0.265 |
| Kidney damage with normal   GFR (>90) | 1245.0(20.76%) | 1776.0(17.75%) | 0.0001*** |
| Alkaline phosphatase, U/L | 91.0(70.0-131.0);n=1196 | 97.0(74.0-134.0);n=2685 | 0.0056** |
| Aspartate transaminase, U/L | 29.0(21.0-42.0);n=412 | 44.0(29.0-74.0);n=1097 | <0.0001*** |
| Alanine transaminase, U/L | 22.0(14.0-34.0);n=1011 | 36.0(22.0-63.0);n=2240 | <0.0001*** |
| Bilirubin, umol/L | 10.0(7.0-14.0);n=1196 | 14.9(10.0-22.4);n=2685 | <0.0001*** |
| Triglyceride, mmol/L | 1.18(0.88-1.68);n=585 | 1.04(0.79-1.47);n=1284 | <0.0001*** |
| Low-density lipoprotein, mmol/L | 2.59(2.02-3.3);n=559 | 2.67(2.06-3.36);n=1231 | 0.6002 |
| High-density lipoprotein, mmol/L | 1.15(0.93-1.43);n=564 | 1.16(0.9-1.43);n=1241 | 0.3995 |
| Total cholesterol, mmol/L | 4.5(3.77-5.3);n=587 | 4.41(3.7-5.25);n=1289 | 0.2821 |
| HbA1C, % | 6.5(6.0-8.8);n=65 | 6.7(6.0-7.8);n=210 | 0.6348 |
| Fasting glucose, mmol/L | 6.28(5.28-8.02);n=398 | 6.1(5.2-7.8);n=1197 | 0.3423 |

### Supplementary Table 20. Landmark sensitivity analysis were conducted impact of CCI, the number of drug classes, calculated biomarkers, liver tests, and lipid and glucose tests on subsequent outcomes after initial HCC diagnosis, after excluding mortality within 30, 60, 90, 180 days, and 1 and 2 years were conducted for sensitivity analysis

* for p≤ 0.05, ** for p ≤ 0.01, *** for p ≤ 0.001. Abbreviations: HR (hazard ratio); CCI (Charlson Comorbidity Index); HCC (Hepatocellular carcinoma)

| ***A. Landmark sensitivity analysis for CCI impact on subsequent outcomes after initial diagnosis*** | | | | | | | | | | | | | | | | |
| --- | --- | --- | --- | --- | --- | --- | --- | --- | --- | --- | --- | --- | --- | --- | --- | --- |
| **Patients** | | | **Comorbidity** | **HCC recurrence  HR [95% CI]; P value** | | **Liver cancer-related.mortality  HR [95% CI];P value** | | | **Cancer-related mortality HR [95% CI];P value** | | | **Non-cancer-related mortality  HR [95% CI];P value** | | | **All-cause mortality  HR [95% CI];P value** | |
| Exclude mortality within 2*365 days | | | CCI | 1.01[1.00-1.02];0.0198* | | 0.87[0.85-0.89];<0.0001*** | | | 1.05[1.04-1.06];<0.0001*** | | | 1.05[1.03-1.07];<0.0001*** | | | 1.05[1.04-1.06];<0.0001*** | |
|  |  |  | Charlson 0 | 0.80[0.68-0.95];0.0094** | | 0.79[0.59-1.06];0.1125 | | | 0.49[0.37-0.64];<0.0001*** | | | 0.25[0.14-0.43];<0.0001*** | | | 0.41[0.32-0.52];<0.0001*** | |
|  |  |  | Charlson 1-5 | 1.01[0.94-1.07];0.8578 | | 3.20[2.75-3.72];<0.0001*** | | | 0.81[0.74-0.89];<0.0001*** | | | 0.98[0.85-1.13];0.7991 | | | 0.86[0.80-0.93];0.0001*** | |
|  |  |  | Charlson 6-10 | 1.02[0.95-1.09];0.5784 | | 0.38[0.32-0.45];<0.0001*** | | | 1.24[1.13-1.36];<0.0001*** | | | 1.09[0.94-1.27];0.2289 | | | 1.19[1.10-1.29];<0.0001*** | |
|  |  |  | Charlson 11+ | 1.04[0.93-1.16];0.4767 | | 0.16[0.10-0.27];<0.0001*** | | | 1.41[1.23-1.61];<0.0001*** | | | 1.41[1.14-1.75];0.0018** | | | 1.41[1.25-1.58];<0.0001*** | |
| Exclude mortality within 365 days | | | CCI | 1.03[1.02-1.03];<0.0001*** | | 0.87[0.85-0.88];<0.0001*** | | | 1.08[1.08-1.09];<0.0001*** | | | 1.06[1.04-1.07];<0.0001*** | | | 1.08[1.07-1.09];<0.0001*** | |
|  |  |  | Charlson 0 | 0.79[0.69-0.91];0.0013** | | 1.08[0.88-1.34];0.4544 | | | 0.56[0.46-0.69];<0.0001*** | | | 0.28[0.17-0.43];<0.0001*** | | | 0.48[0.40-0.58];<0.0001*** | |
|  |  |  | Charlson 1-5 | 0.87[0.82-0.91];<0.0001*** | | 2.87[2.56-3.21];<0.0001*** | | | 0.60[0.56-0.64];<0.0001*** | | | 0.94[0.84-1.05];0.2506 | | | 0.67[0.63-0.71];<0.0001*** | |
|  |  |  | Charlson 6-10 | 1.13[1.07-1.19];<0.0001*** | | 0.42[0.37-0.48];<0.0001*** | | | 1.48[1.39-1.58];<0.0001*** | | | 1.05[0.93-1.18];0.4274 | | | 1.37[1.29-1.45];<0.0001*** | |
|  |  |  | Charlson 11+ | 1.16[1.08-1.26];0.0001*** | | 0.18[0.13-0.25];<0.0001*** | | | 1.62[1.48-1.77];<0.0001*** | | | 1.54[1.31-1.81];<0.0001*** | | | 1.60[1.48-1.73];<0.0001*** | |
| Exclude mortality within 180 days | | | CCI | 1.04[1.03-1.04];<0.0001*** | | 0.87[0.86-0.88];<0.0001*** | | | 1.10[1.09-1.10];<0.0001*** | | | 1.07[1.06-1.08];<0.0001*** | | | 1.09[1.08-1.10];<0.0001*** | |
|  |  |  | Charlson 0 | 0.80[0.70-0.91];0.0007*** | | 1.24[1.04-1.49];0.0189* | | | 0.59[0.50-0.70];<0.0001*** | | | 0.29[0.19-0.43];<0.0001*** | | | 0.52[0.44-0.60];<0.0001*** | |
|  |  |  | Charlson 1-5 | 0.79[0.76-0.83];<0.0001*** | | 2.68[2.44-2.95];<0.0001*** | | | 0.54[0.51-0.57];<0.0001*** | | | 0.84[0.76-0.93];0.0008*** | | | 0.60[0.57-0.63];<0.0001*** | |
|  |  |  | Charlson 6-10 | 1.18[1.13-1.24];<0.0001*** | | 0.46[0.41-0.51];<0.0001*** | | | 1.48[1.40-1.56];<0.0001*** | | | 1.07[0.96-1.19];0.2137 | | | 1.38[1.31-1.45];<0.0001*** | |
|  |  |  | Charlson 11+ | 1.21[1.14-1.29];<0.0001*** | | 0.20[0.15-0.26];<0.0001*** | | | 1.74[1.62-1.87];<0.0001*** | | | 1.71[1.49-1.95];<0.0001*** | | | 1.73[1.63-1.84];<0.0001*** | |
| Exclude mortality within 120 days | | | CCI | 1.04[1.04-1.05];<0.0001*** | | 0.87[0.86-0.88];<0.0001*** | | | 1.10[1.10-1.11];<0.0001*** | | | 1.07[1.06-1.09];<0.0001*** | | | 1.10[1.09-1.10];<0.0001*** | |
|  |  |  | Charlson 0 | 0.78[0.69-0.89];0.0002*** | | 1.22[1.03-1.46];0.0252* | | | 0.57[0.48-0.67];<0.0001*** | | | 0.29[0.20-0.43];<0.0001*** | | | 0.50[0.43-0.58];<0.0001*** | |
|  |  |  | Charlson 1-5 | 0.77[0.74-0.80];<0.0001*** | | 2.61[2.39-2.86];<0.0001*** | | | 0.52[0.49-0.55];<0.0001*** | | | 0.80[0.73-0.88];<0.0001*** | | | 0.57[0.55-0.60];<0.0001*** | |
|  |  |  | Charlson 6-10 | 1.18[1.13-1.23];<0.0001*** | | 0.48[0.44-0.53];<0.0001*** | | | 1.47[1.40-1.55];<0.0001*** | | | 1.06[0.96-1.17];0.2760 | | | 1.37[1.31-1.43];<0.0001*** | |
|  |  |  | Charlson 11+ | 1.26[1.19-1.34];<0.0001*** | | 0.22[0.18-0.28];<0.0001*** | | | 1.81[1.70-1.93];<0.0001*** | | | 1.82[1.61-2.06];<0.0001*** | | | 1.81[1.71-1.92];<0.0001*** | |
| Exclude mortality within 90 days | | | CCI | 1.04[1.04-1.05];<0.0001*** | | 0.88[0.87-0.89];<0.0001*** | | | 1.11[1.10-1.11];<0.0001*** | | | 1.08[1.07-1.09];<0.0001*** | | | 1.10[1.09-1.11];<0.0001*** | |
|  |  |  | Charlson 0 | 0.78[0.69-0.88];0.0001*** | | 1.23[1.04-1.46];0.0171* | | | 0.57[0.48-0.67];<0.0001*** | | | 0.29[0.20-0.43];<0.0001*** | | | 0.50[0.43-0.58];<0.0001*** | |
|  |  |  | Charlson 1-5 | 0.76[0.72-0.79];<0.0001*** | | 2.56[2.35-2.79];<0.0001*** | | | 0.50[0.48-0.53];<0.0001*** | | | 0.78[0.71-0.85];<0.0001*** | | | 0.56[0.53-0.58];<0.0001*** | |
|  |  |  | Charlson 6-10 | 1.17[1.12-1.22];<0.0001*** | | 0.49[0.45-0.54];<0.0001*** | | | 1.44[1.37-1.51];<0.0001*** | | | 1.08[0.98-1.19];0.1230 | | | 1.35[1.29-1.41];<0.0001*** | |
|  |  |  | Charlson 11+ | 1.29[1.22-1.36];<0.0001*** | | 0.26[0.21-0.32];<0.0001*** | | | 1.88[1.77-1.99];<0.0001*** | | | 1.82[1.62-2.05];<0.0001*** | | | 1.87[1.77-1.97];<0.0001*** | |
| Exclude mortality within 60 days | | | CCI | 1.05[1.04-1.05];<0.0001*** | | 0.88[0.87-0.89];<0.0001*** | | | 1.11[1.10-1.12];<0.0001*** | | | 1.08[1.07-1.09];<0.0001*** | | | 1.10[1.10-1.11];<0.0001*** | |
|  |  |  | Charlson 0 | 0.78[0.69-0.88];0.0001*** | | 1.29[1.10-1.52];0.0019** | | | 0.58[0.50-0.68];<0.0001*** | | | 0.29[0.20-0.42];<0.0001*** | | | 0.51[0.44-0.59];<0.0001*** | |
|  |  |  | Charlson 1-5 | 0.74[0.71-0.77];<0.0001*** | | 2.46[2.27-2.66];<0.0001*** | | | 0.49[0.47-0.52];<0.0001*** | | | 0.76[0.69-0.83];<0.0001*** | | | 0.54[0.52-0.57];<0.0001*** | |
|  |  |  | Charlson 6-10 | 1.17[1.13-1.22];<0.0001*** | | 0.52[0.47-0.56];<0.0001*** | | | 1.41[1.35-1.48];<0.0001*** | | | 1.06[0.97-1.16];0.2075 | | | 1.33[1.28-1.39];<0.0001*** | |
|  |  |  | Charlson 11+ | 1.31[1.24-1.37];<0.0001*** | | 0.27[0.22-0.33];<0.0001*** | | | 1.91[1.80-2.02];<0.0001*** | | | 1.87[1.68-2.09];<0.0001*** | | | 1.90[1.81-2.00];<0.0001*** | |
| Exclude mortality within 30 days | | | CCI | 1.05[1.04-1.05];<0.0001*** | | 0.88[0.88-0.89];<0.0001*** | | | 1.11[1.11-1.12];<0.0001*** | | | 1.09[1.08-1.10];<0.0001*** | | | 1.11[1.10-1.12];<0.0001*** | |
|  |  |  | Charlson 0 | 0.77[0.68-0.87];<0.0001*** | | 1.30[1.11-1.52];0.0012** | | | 0.57[0.49-0.66];<0.0001*** | | | 0.29[0.20-0.42];<0.0001*** | | | 0.50[0.44-0.57];<0.0001*** | |
|  |  |  | Charlson 1-5 | 0.72[0.69-0.75];<0.0001*** | | 2.36[2.19-2.55];<0.0001*** | | | 0.47[0.45-0.50];<0.0001*** | | | 0.73[0.67-0.79];<0.0001*** | | | 0.52[0.50-0.54];<0.0001*** | |
|  |  |  | Charlson 6-10 | 1.17[1.13-1.22];<0.0001*** | | 0.55[0.51-0.60];<0.0001*** | | | 1.39[1.33-1.46];<0.0001*** | | | 1.01[0.93-1.11];0.7882 | | | 1.31[1.26-1.36];<0.0001*** | |
|  |  |  | Charlson 11+ | 1.32[1.27-1.39];<0.0001*** | | 0.30[0.25-0.35];<0.0001*** | | | 1.94[1.85-2.04];<0.0001*** | | | 2.05[1.85-2.26];<0.0001*** | | | 1.96[1.88-2.05];<0.0001*** | |
| B. ***Landmark sensitivity analysis for drug classes impact on subsequent outcomes after HCC diagnosis*** | | | | | | | | | | | | | | | | |
| Patients | | Drug classes | | HCC recurrence HR [95% CI]; P value | | | Liver cancer-related.mortality HR [95% CI];P value | | | Cancer-related mortality HR [95% CI];P value | | | Non-cancer-related mortality HR [95% CI];P value | | All-cause mortality HR [95% CI]; P value | |
| Exclude mortality within 2*365 days | | Number of prescribed drug classes | | 1.09[1.04-1.14];0.0002*** | | | 1.20[1.11-1.30];<0.0001*** | | | 1.21[1.14-1.28];<0.0001*** | | | 1.53[1.41-1.67];<0.0001*** | | 1.30[1.24-1.36];<0.0001*** | |
|  |  | N=0 | | 0.85[0.78-0.92];<0.0001*** | | | 0.62[0.53-0.73];<0.0001*** | | | 0.64[0.57-0.71];<0.0001*** | | | 0.42[0.34-0.52];<0.0001*** | | 0.57[0.51-0.63];<0.0001*** | |
|  |  | N=1 | | 1.07[1.00-1.14];0.0500 | | | 1.34[1.18-1.52];<0.0001*** | | | 1.28[1.16-1.40];<0.0001*** | | | 1.19[1.03-1.37];0.0170* | | 1.25[1.16-1.35];<0.0001*** | |
|  |  | N=2+ | | 1.09[1.00-1.18];0.0475* | | | 1.08[0.92-1.26];0.3582 | | | 1.13[1.01-1.26];0.0367* | | | 1.73[1.48-2.03];<0.0001*** | | 1.29[1.18-1.41];<0.0001*** | |
| Exclude mortality within 365 days | | Number of prescribed drug classes | | 1.07[1.04-1.11];0.0001*** | | | 1.18[1.11-1.26];<0.0001*** | | | 1.15[1.10-1.20];<0.0001*** | | | 1.44[1.34-1.55];<0.0001*** | | 1.22[1.17-1.26];<0.0001*** | |
|  |  | N=0 | | 0.86[0.80-0.91];<0.0001*** | | | 0.63[0.56-0.72];<0.0001*** | | | 0.71[0.65-0.77];<0.0001*** | | | 0.46[0.39-0.54];<0.0001*** | | 0.64[0.59-0.69];<0.0001*** | |
|  |  | N=1 | | 1.08[1.02-1.14];0.0048** | | | 1.32[1.20-1.46];<0.0001*** | | | 1.21[1.13-1.29];<0.0001*** | | | 1.22[1.09-1.37];0.0006*** | | 1.21[1.14-1.28];<0.0001*** | |
|  |  | N=2+ | | 1.05[0.98-1.12];0.1546 | | | 1.06[0.93-1.19];0.3856 | | | 1.08[1.00-1.17];0.0623 | | | 1.51[1.33-1.72];<0.0001*** | | 1.18[1.10-1.26];<0.0001*** | |
| Exclude mortality within 180 days | | Number of prescribed drug classes | | 1.04[1.01-1.08];0.0077** | | | 1.14[1.08-1.21];<0.0001*** | | | 1.10[1.06-1.14];<0.0001*** | | | 1.37[1.28-1.46];<0.0001*** | | 1.16[1.12-1.20];<0.0001*** | |
|  |  | N=0 | | 0.89[0.84-0.94];0.0001*** | | | 0.69[0.62-0.77];<0.0001*** | | | 0.77[0.72-0.83];<0.0001*** | | | 0.52[0.45-0.60];<0.0001*** | | 0.71[0.67-0.76];<0.0001*** | |
|  |  | N=1 | | 1.09[1.04-1.14];0.0006*** | | | 1.28[1.17-1.40];<0.0001*** | | | 1.18[1.11-1.24];<0.0001*** | | | 1.18[1.06-1.31];0.0018** | | 1.18[1.12-1.24];<0.0001*** | |
|  |  | N=2+ | | 0.99[0.94-1.05];0.8526 | | | 1.01[0.91-1.13];0.8275 | | | 1.02[0.96-1.10];0.4976 | | | 1.43[1.27-1.61];<0.0001*** | | 1.11[1.05-1.18];0.0006*** | |
| Exclude mortality within 120 days | | Number of prescribed drug classes | | 1.03[1.00-1.07];0.0229* | | | 1.12[1.06-1.18];<0.0001*** | | | 1.08[1.05-1.12];<0.0001*** | | | 1.36[1.28-1.44];<0.0001*** | | 1.14[1.11-1.18];<0.0001*** | |
|  |  | N=0 | | 0.91[0.86-0.96];0.0003*** | | | 0.72[0.65-0.80];<0.0001*** | | | 0.81[0.76-0.86];<0.0001*** | | | 0.55[0.48-0.62];<0.0001*** | | 0.75[0.70-0.79];<0.0001*** | |
|  |  | N=1 | | 1.08[1.03-1.13];0.0006*** | | | 1.25[1.15-1.35];<0.0001*** | | | 1.14[1.08-1.20];<0.0001*** | | | 1.13[1.03-1.25];0.0120* | | 1.14[1.09-1.19];<0.0001*** | |
|  |  | N=2+ | | 0.99[0.93-1.04];0.5919 | | | 1.01[0.91-1.11];0.9088 | | | 1.02[0.96-1.09];0.5466 | | | 1.45[1.30-1.62];<0.0001*** | | 1.11[1.05-1.17];0.0003*** | |
| Exclude mortality within 90 days | | Number of prescribed drug classes | | 1.03[1.00-1.06];0.0288* | | | 1.12[1.06-1.18];<0.0001*** | | | 1.08[1.05-1.12];<0.0001*** | | | 1.35[1.27-1.43];<0.0001*** | | 1.14[1.10-1.17];<0.0001*** | |
|  |  | N=0 | | 0.92[0.87-0.97];0.0015** | | | 0.72[0.66-0.80];<0.0001*** | | | 0.83[0.78-0.88];<0.0001*** | | | 0.57[0.50-0.65];<0.0001*** | | 0.76[0.72-0.81];<0.0001*** | |
|  |  | N=1 | | 1.06[1.02-1.11];0.0043** | | | 1.24[1.15-1.35];<0.0001*** | | | 1.11[1.06-1.17];<0.0001*** | | | 1.11[1.01-1.22];0.0297* | | 1.11[1.07-1.16];<0.0001*** | |
|  |  | N=2+ | | 0.99[0.94-1.05];0.8010 | | | 1.01[0.91-1.11];0.9139 | | | 1.04[0.98-1.10];0.2380 | | | 1.44[1.29-1.60];<0.0001*** | | 1.12[1.06-1.18];<0.0001*** | |
| Exclude mortality within 60 days | | Number of prescribed drug classes | | 1.03[1.00-1.06];0.0331* | | | 1.12[1.06-1.17];<0.0001*** | | | 1.08[1.05-1.12];<0.0001*** | | | 1.33[1.25-1.40];<0.0001*** | | 1.13[1.10-1.16];<0.0001*** | |
|  |  | N=0 | | 0.93[0.89-0.98];0.0064** | | | 0.74[0.67-0.81];<0.0001*** | | | 0.84[0.79-0.89];<0.0001*** | | | 0.59[0.53-0.67];<0.0001*** | | 0.78[0.74-0.82];<0.0001*** | |
|  |  | N=1 | | 1.04[1.00-1.09];0.0438* | | | 1.21[1.12-1.31];<0.0001*** | | | 1.09[1.04-1.14];0.0004*** | | | 1.09[0.99-1.19];0.0739 | | 1.09[1.04-1.14];0.0001*** | |
|  |  | N=2+ | | 1.01[0.96-1.06];0.7587 | | | 1.03[0.93-1.13];0.5800 | | | 1.06[1.00-1.12];0.0620 | | | 1.43[1.29-1.59];<0.0001*** | | 1.13[1.08-1.19];<0.0001*** | |
| Exclude mortality within 30 days | | Number of prescribed drug classes | | 1.02[0.99-1.05];0.1258 | | | 1.11[1.05-1.16];<0.0001*** | | | 1.08[1.05-1.11];<0.0001*** | | | 1.33[1.26-1.40];<0.0001*** | | 1.13[1.10-1.16];<0.0001*** | |
|  |  | N=0 | | 0.96[0.91-1.00];0.0571 | | | 0.77[0.70-0.84];<0.0001*** | | | 0.86[0.82-0.91];<0.0001*** | | | 0.60[0.54-0.68];<0.0001*** | | 0.80[0.77-0.85];<0.0001*** | |
|  |  | N=1 | | 1.03[0.99-1.07];0.1663 | | | 1.17[1.09-1.26];<0.0001*** | | | 1.06[1.02-1.11];0.0059** | | | 1.06[0.97-1.15];0.2113 | | 1.06[1.02-1.10];0.0025** | |
|  |  | N=2+ | | 1.01[0.96-1.05];0.8167 | | | 1.04[0.95-1.13];0.4408 | | | 1.06[1.01-1.12];0.0307* | | | 1.46[1.33-1.61];<0.0001*** | | 1.14[1.09-1.19];<0.0001*** | |
| **C. Landmark sensitivity analysis for calculated** **biomarkers on subsequent outcomes after HCC diagnosis** | | | | | | | | | | | | | | | | |
| Patients | | Calculated biomarkers | | HCC recurrence HR [95% CI]; P value | | | Liver cancer-related mortality HR [95% CI];P value | | | Cancer-related mortality HR [95% CI];P value | | | Non-cancer-related mortality HR [95% CI];P value | | All-cause mortality HR [95% CI];P value | |
| Exclude mortality within 2*365 days | | Fibrosis-4 (FIB-4) index | | 1.00[0.99-1.02];0.6104 | | | 1.02[1.00-1.04];0.0190* | | | 1.01[0.99-1.03];0.3109 | | | 0.99[0.94-1.04];0.6439 | | 1.01[0.99-1.02];0.4825 | |
|  |  | APRI | | 1.000[0.999-1.001];0.7367 | | | 1.001[1.000-1.003];0.0457* | | | 1.001[0.999-1.002];0.3279 | | | 1.000[0.997-1.003];0.9136 | | 1.001[0.999-1.002];0.3562 | |
|  |  | ALBI score | | 1.11[1.03-1.19];0.0090** | | | 1.70[1.52-1.90];<0.0001*** | | | 1.20[1.10-1.32];0.0001*** | | | 1.22[1.04-1.42];0.0141* | | 1.21[1.11-1.31];<0.0001*** | |
|  |  | AST/ALT ratio | | 0.99[0.94-1.05];0.8216 | | | 1.01[0.93-1.10];0.7479 | | | 1.00[0.93-1.08];0.9400 | | | 0.97[0.82-1.14];0.6843 | | 1.00[0.93-1.06];0.8922 | |
|  |  | ALT/PLT ratio | | 1.000[0.999-1.001];0.7663 | | | 1.001[1.000-1.002];0.0111* | | | 1.000[1.000-1.001];0.2861 | | | 1.000[0.998-1.002];0.7629 | | 1.000[1.000-1.001];0.4289 | |
|  |  | Urea-to-Creatinine ratio | | 0.999[0.997-1.002];0.5678 | | | 1.00[0.99-1.00];0.1491 | | | 0.999[0.996-1.002];0.5034 | | | 1.00[0.99-1.00];0.3297 | | 0.998[0.996-1.001];0.2821 | |
|  |  | CKD-EPI (2021) | | 0.998[0.996-1.000];0.1092 | | | 0.99[0.99-1.00];0.0054** | | | 1.00[0.99-1.00];0.0602 | | | 0.98[0.97-0.98];<0.0001*** | | 0.992[0.989-0.994];<0.0001*** | |
| Exclude mortality within 365 days | | Fibrosis-4 (FIB-4) index | | 1.00[0.99-1.01];0.8044 | | | 1.02[1.00-1.03];0.0090** | | | 1.00[0.98-1.02];0.8620 | | | 1.01[0.98-1.04];0.5006 | | 1.00[0.99-1.02];0.8781 | |
|  |  | APRI | | 1.000[0.999-1.001];0.4938 | | | 1.001[1.000-1.002];0.1064 | | | 1.000[0.998-1.001];0.4172 | | | 1.000[0.999-1.002];0.5911 | | 1.000[0.999-1.001];0.6380 | |
|  |  | ALBI score | | 1.11[1.05-1.18];0.0002*** | | | 1.80[1.65-1.97];<0.0001*** | | | 1.13[1.06-1.21];0.0003*** | | | 1.31[1.16-1.48];<0.0001*** | | 1.17[1.10-1.24];<0.0001*** | |
|  |  | AST/ALT ratio | | 1.01[0.97-1.05];0.6227 | | | 1.02[0.95-1.09];0.6163 | | | 1.03[0.98-1.07];0.2715 | | | 1.00[0.90-1.11];0.9670 | | 1.02[0.98-1.06];0.3239 | |
|  |  | ALT/PLT ratio | | 0.999[0.999-1.000];0.0496* | | | 1.001[1.000-1.001];0.0418* | | | 0.999[0.998-1.000];0.0520 | | | 1.000[0.998-1.001];0.7835 | | 0.999[0.999-1.000];0.0634 | |
|  |  | Urea-to-Creatinine ratio | | 0.999[0.997-1.001];0.5205 | | | 0.999[0.996-1.002];0.5697 | | | 0.999[0.997-1.002];0.5643 | | | 1.00[0.99-1.00];0.4480 | | 0.999[0.997-1.001];0.3866 | |
|  |  | CKD-EPI (2021) | | 0.999[0.997-1.001];0.2551 | | | 0.99[0.99-1.00];0.0002*** | | | 0.999[0.997-1.001];0.3072 | | | 0.981[0.978-0.984];<0.0001*** | | 0.99[0.99-1.00];<0.0001*** | |
| Exclude mortality within 180 days | | Fibrosis-4 (FIB-4) index | | 1.00[0.99-1.01];0.8510 | | | 1.02[1.01-1.03];0.0071** | | | 0.99[0.98-1.01];0.4122 | | | 1.01[0.98-1.03];0.7105 | | 1.00[0.98-1.01];0.5729 | |
|  |  | APRI | | 1.000[0.999-1.000];0.2851 | | | 1.001[1.000-1.002];0.0676 | | | 0.999[0.998-1.000];0.1791 | | | 1.000[0.998-1.002];0.8584 | | 0.999[0.999-1.000];0.2641 | |
|  |  | ALBI score | | 1.10[1.04-1.15];0.0003*** | | | 1.84[1.71-2.00];<0.0001*** | | | 1.13[1.07-1.20];<0.0001*** | | | 1.31[1.17-1.46];<0.0001*** | | 1.17[1.11-1.23];<0.0001*** | |
|  |  | AST/ALT ratio | | 1.02[0.99-1.06];0.1515 | | | 1.01[0.95-1.08];0.6577 | | | 1.04[1.00-1.08];0.0487* | | | 1.01[0.93-1.11];0.7416 | | 1.03[1.00-1.07];0.0574 | |
|  |  | ALT/PLT ratio | | 0.999[0.999-1.000];0.0179* | | | 1.001[1.000-1.001];0.0165* | | | 0.999[0.998-1.000];0.0071** | | | 1.000[0.998-1.001];0.5687 | | 0.999[0.998-1.000];0.0077** | |
|  |  | Urea-to-Creatinine ratio | | 1.001[0.999-1.003];0.2425 | | | 1.000[0.997-1.003];0.9642 | | | 1.001[0.999-1.003];0.4716 | | | 0.999[0.995-1.003];0.7935 | | 1.000[0.999-1.002];0.6015 | |
|  |  | CKD-EPI (2021) | | 0.999[0.998-1.001];0.2760 | | | 0.99[0.99-1.00];<0.0001*** | | | 0.999[0.997-1.000];0.1125 | | | 0.98[0.98-0.99];<0.0001*** | | 0.99[0.99-1.00];<0.0001*** | |
| Exclude mortality within 120 days | | Fibrosis-4 (FIB-4) index | | 1.00[0.99-1.01];0.7961 | | | 1.02[1.00-1.03];0.0087** | | | 0.99[0.98-1.01];0.3281 | | | 1.00[0.98-1.03];0.7238 | | 0.99[0.98-1.01];0.4766 | |
|  |  | APRI | | 0.999[0.999-1.000];0.2416 | | | 1.001[1.000-1.002];0.0776 | | | 0.999[0.998-1.000];0.1286 | | | 1.000[0.998-1.002];0.8394 | | 0.999[0.999-1.000];0.2067 | |
|  |  | ALBI score | | 1.10[1.05-1.15];0.0001*** | | | 1.83[1.70-1.98];<0.0001*** | | | 1.13[1.07-1.19];<0.0001*** | | | 1.31[1.18-1.46];<0.0001*** | | 1.16[1.11-1.22];<0.0001*** | |
|  |  | AST/ALT ratio | | 1.03[0.99-1.06];0.1090 | | | 1.01[0.95-1.08];0.6632 | | | 1.04[1.00-1.08];0.0281* | | | 1.01[0.92-1.11];0.8154 | | 1.04[1.00-1.07];0.0397* | |
|  |  | ALT/PLT ratio | | 0.999[0.998-1.000];0.0134* | | | 1.001[1.000-1.001];0.0155* | | | 0.999[0.998-1.000];0.0045** | | | 1.000[0.998-1.001];0.5774 | | 0.999[0.998-1.000];0.0053** | |
|  |  | Urea-to-Creatinine ratio | | 1.001[0.999-1.003];0.1972 | | | 1.000[0.997-1.003];0.8202 | | | 1.001[0.999-1.003];0.2721 | | | 0.999[0.995-1.003];0.6027 | | 1.001[0.999-1.002];0.4576 | |
|  |  | CKD-EPI (2021) | | 0.999[0.998-1.000];0.1436 | | | 0.99[0.99-1.00];<0.0001*** | | | 0.998[0.996-1.000];0.0177* | | | 0.98[0.98-0.99];<0.0001*** | | 0.99[0.99-1.00];<0.0001*** | |
| Exclude mortality within 90 days | | Fibrosis-4 (FIB-4) index | | 1.00[0.99-1.01];0.7849 | | | 1.02[1.01-1.03];0.0077** | | | 0.99[0.98-1.01];0.3222 | | | 1.00[0.98-1.03];0.7238 | | 0.99[0.98-1.01];0.4694 | |
|  |  | APRI | | 0.999[0.999-1.000];0.2255 | | | 1.001[1.000-1.002];0.0743 | | | 0.999[0.998-1.000];0.1168 | | | 1.000[0.998-1.002];0.8394 | | 0.999[0.998-1.000];0.1911 | |
|  |  | ALBI score | | 1.10[1.05-1.15];0.0001*** | | | 1.83[1.70-1.97];<0.0001*** | | | 1.13[1.07-1.19];<0.0001*** | | | 1.32[1.19-1.47];<0.0001*** | | 1.17[1.11-1.22];<0.0001*** | |
|  |  | AST/ALT ratio | | 1.03[0.99-1.06];0.1032 | | | 1.01[0.95-1.08];0.6988 | | | 1.04[1.00-1.08];0.0301* | | | 1.01[0.93-1.11];0.7978 | | 1.04[1.00-1.07];0.0408* | |
|  |  | ALT/PLT ratio | | 0.999[0.998-1.000];0.0122* | | | 1.001[1.000-1.001];0.0102* | | | 0.999[0.998-1.000];0.0040** | | | 1.000[0.998-1.001];0.5980 | | 0.999[0.998-1.000];0.0049** | |
|  |  | Urea-to-Creatinine ratio | | 1.001[0.999-1.002];0.4153 | | | 1.000[0.997-1.003];0.8286 | | | 1.001[0.999-1.003];0.2285 | | | 1.00[0.99-1.00];0.4830 | | 1.001[0.999-1.002];0.4482 | |
|  |  | CKD-EPI (2021) | | 0.999[0.997-1.000];0.0423* | | | 0.99[0.99-1.00];<0.0001*** | | | 0.998[0.996-0.999];0.0025** | | | 0.98[0.98-0.99];<0.0001*** | | 0.99[0.99-1.00];<0.0001*** | |
| Exclude mortality within 60 days | | Fibrosis-4 (FIB-4) index | | 1.00[0.99-1.01];0.8320 | | | 1.02[1.01-1.03];0.0078** | | | 0.99[0.97-1.01];0.2979 | | | 1.00[0.98-1.03];0.7238 | | 0.99[0.98-1.01];0.4410 | |
|  |  | APRI | | 1.000[0.999-1.000];0.2495 | | | 1.001[1.000-1.002];0.0738 | | | 0.999[0.998-1.000];0.1085 | | | 1.000[0.998-1.002];0.8394 | | 0.999[0.998-1.000];0.1796 | |
|  |  | ALBI score | | 1.09[1.04-1.14];0.0002*** | | | 1.83[1.70-1.97];<0.0001*** | | | 1.13[1.07-1.19];<0.0001*** | | | 1.31[1.18-1.46];<0.0001*** | | 1.17[1.11-1.22];<0.0001*** | |
|  |  | AST/ALT ratio | | 1.03[1.00-1.06];0.0821 | | | 1.02[0.96-1.08];0.5864 | | | 1.04[1.00-1.08];0.0269* | | | 1.01[0.93-1.11];0.7947 | | 1.04[1.00-1.07];0.0366* | |
|  |  | ALT/PLT ratio | | 0.999[0.998-1.000];0.0093** | | | 1.001[1.000-1.001];0.0116* | | | 0.999[0.998-1.000];0.0023** | | | 1.000[0.998-1.001];0.5811 | | 0.999[0.998-1.000];0.0030** | |
|  |  | Urea-to-Creatinine ratio | | 1.001[0.999-1.003];0.1843 | | | 1.001[0.998-1.003];0.7207 | | | 1.001[1.000-1.003];0.1120 | | | 0.999[0.995-1.003];0.6829 | | 1.001[0.999-1.003];0.2152 | |
|  |  | CKD-EPI (2021) | | 0.998[0.997-1.000];0.0091** | | | 0.992[0.990-0.994];<0.0001*** | | | 0.997[0.996-0.999];0.0002*** | | | 0.98[0.98-0.99];<0.0001*** | | 0.99[0.99-1.00];<0.0001*** | |
| Exclude mortality within 30 days | | Fibrosis-4 (FIB-4) index | | 1.00[0.99-1.01];0.8128 | | | 1.02[1.01-1.03];0.0078** | | | 0.99[0.97-1.01];0.2566 | | | 1.00[0.97-1.03];0.8718 | | 0.99[0.98-1.01];0.3486 | |
|  |  | APRI | | 1.000[0.999-1.000];0.2538 | | | 1.001[1.000-1.002];0.0738 | | | 0.999[0.998-1.000];0.0907 | | | 1.000[0.998-1.002];0.9797 | | 0.999[0.998-1.000];0.1347 | |
|  |  | ALBI score | | 1.09[1.04-1.14];0.0003*** | | | 1.81[1.69-1.95];<0.0001*** | | | 1.12[1.07-1.18];<0.0001*** | | | 1.28[1.16-1.42];<0.0001*** | | 1.15[1.10-1.21];<0.0001*** | |
|  |  | AST/ALT ratio | | 1.03[1.00-1.06];0.0786 | | | 1.02[0.95-1.08];0.6276 | | | 1.04[1.00-1.08];0.0263* | | | 1.03[0.96-1.11];0.4304 | | 1.04[1.01-1.07];0.0194* | |
|  |  | ALT/PLT ratio | | 0.999[0.998-1.000];0.0077** | | | 1.001[1.000-1.001];0.0139* | | | 0.999[0.998-0.999];0.0014** | | | 0.999[0.998-1.001];0.4816 | | 0.999[0.998-1.000];0.0015** | |
|  |  | Urea-to-Creatinine ratio | | 1.001[1.000-1.003];0.1085 | | | 1.000[0.998-1.003];0.7483 | | | 1.002[1.000-1.003];0.0493* | | | 0.999[0.995-1.003];0.6326 | | 1.001[1.000-1.003];0.1204 | |
|  |  | CKD-EPI (2021) | | 0.998[0.997-0.999];0.0012** | | | 0.992[0.990-0.994];<0.0001*** | | | 0.997[0.995-0.998];<0.0001*** | | | 0.982[0.980-0.984];<0.0001*** | | 0.993[0.992-0.994];<0.0001*** | |
| ***D. Landmark sensitivity analysis for liver tests impact on subsequent outcomes after HCC diagnosis*** | | | | | | | | | | | | | | | | |
| **Patients** | | **Liver tests** | | **HCC recurrence HR [95% CI];P value** | | | **Liver cancer-related.mortality HR [95% CI];P value** | | | **Cancer-related mortality HR [95% CI];P value** | | | **Non-cancer-related mortality HR [95% CI];P value** | | **All-cause mortality**  **HR [95% CI];P value** | |
| Exclude mortality within 2*365 days | | Alkaline phosphatase, U/L | | 1.002[1.001-1.003];<0.0001*** | | | 1.002[1.001-1.003];0.0040** | | | 1.001[1.000-1.002];0.0057** | | | 1.000[0.998-1.002];0.8583 | | 1.001[1.000-1.002];0.0136* | |
|  |  | Aspartate transaminase, U/L | | 1.000[0.999-1.001];0.6844 | | | 1.002[1.001-1.003];0.0024** | | | 1.001[1.000-1.002];0.0644 | | | 1.000[0.998-1.002];0.8100 | | 1.001[1.000-1.002];0.0837 | |
|  |  | Alanine transaminase, U/L | | 1.000[1.000-1.001];0.4979 | | | 1.001[1.001-1.002];0.0002*** | | | 1.000[1.000-1.001];0.1610 | | | 1.000[0.999-1.001];0.9784 | | 1.000[1.000-1.001];0.2215 | |
|  |  | Bilirubin, umol/L | | 1.00[1.00-1.01];0.2320 | | | 1.02[1.01-1.02];<0.0001*** | | | 1.00[1.00-1.01];0.2555 | | | 1.00[0.99-1.01];0.6567 | | 1.00[1.00-1.01];0.2282 | |
| Exclude mortality within 365 days | | Alkaline phosphatase, U/L | | 1.001[1.001-1.002];<0.0001*** | | | 1.002[1.001-1.003];<0.0001*** | | | 1.002[1.001-1.002];<0.0001*** | | | 1.001[1.000-1.002];0.0199* | | 1.002[1.001-1.002];<0.0001*** | |
|  |  | Aspartate transaminase, U/L | | 1.000[0.999-1.000];0.3084 | | | 1.001[1.001-1.002];0.0012** | | | 1.000[0.999-1.001];0.4949 | | | 1.000[0.998-1.002];0.9626 | | 1.000[0.999-1.001];0.5337 | |
|  |  | Alanine transaminase, U/L | | 1.000[0.999-1.000];0.3078 | | | 1.001[1.001-1.002];<0.0001*** | | | 1.000[0.999-1.000];0.2162 | | | 1.000[0.999-1.001];0.6869 | | 1.000[0.999-1.000];0.2001 | |
|  |  | Bilirubin, umol/L | | 1.001[0.998-1.004];0.3488 | | | 1.01[1.01-1.02];<0.0001*** | | | 1.00[1.00-1.01];0.3144 | | | 1.01[1.00-1.01];0.0537 | | 1.00[1.00-1.01];0.0757 | |
| Exclude mortality within 180 days | | Alkaline phosphatase, U/L | | 1.002[1.001-1.002];<0.0001*** | | | 1.002[1.002-1.003];<0.0001*** | | | 1.002[1.001-1.002];<0.0001*** | | | 1.001[1.000-1.002];0.0025** | | 1.002[1.001-1.002];<0.0001*** | |
|  |  | Aspartate transaminase, U/L | | 1.000[0.999-1.000];0.4790 | | | 1.002[1.001-1.002];<0.0001*** | | | 1.000[1.000-1.001];0.3006 | | | 1.000[0.998-1.001];0.6610 | | 1.000[1.000-1.001];0.4739 | |
|  |  | Alanine transaminase, U/L | | 0.999[0.999-1.000];0.0700 | | | 1.001[1.001-1.002];<0.0001*** | | | 0.999[0.999-1.000];0.0178* | | | 0.999[0.998-1.001];0.3710 | | 0.999[0.999-1.000];0.0118* | |
|  |  | Bilirubin, umol/L | | 1.001[0.998-1.003];0.4972 | | | 1.012[1.010-1.014];<0.0001*** | | | 1.002[0.999-1.005];0.2228 | | | 1.01[1.00-1.01];0.0368* | | 1.00[1.00-1.01];0.0443* | |
| Exclude mortality within 120 days | | Alkaline phosphatase, U/L | | 1.001[1.001-1.002];<0.0001*** | | | 1.002[1.001-1.003];<0.0001*** | | | 1.002[1.001-1.002];<0.0001*** | | | 1.002[1.001-1.002];0.0001*** | | 1.002[1.001-1.002];<0.0001*** | |
|  |  | Aspartate transaminase, U/L | | 1.000[0.999-1.000];0.3794 | | | 1.002[1.001-1.002];<0.0001*** | | | 1.000[1.000-1.001];0.4305 | | | 1.000[0.998-1.001];0.6289 | | 1.000[0.999-1.001];0.6317 | |
|  |  | Alanine transaminase, U/L | | 0.999[0.999-1.000];0.0201* | | | 1.001[1.001-1.002];<0.0001*** | | | 0.999[0.998-1.000];0.0046** | | | 0.999[0.998-1.000];0.1879 | | 0.999[0.998-1.000];0.0018** | |
|  |  | Bilirubin, umol/L | | 1.000[0.998-1.003];0.8490 | | | 1.011[1.009-1.013];<0.0001*** | | | 1.001[0.998-1.004];0.6156 | | | 1.00[1.00-1.01];0.0891 | | 1.002[0.999-1.004];0.2310 | |
| Exclude mortality within 90 days | | Alkaline phosphatase, U/L | | 1.001[1.001-1.001];<0.0001*** | | | 1.002[1.002-1.003];<0.0001*** | | | 1.002[1.002-1.002];<0.0001*** | | | 1.001[1.001-1.002];0.0002*** | | 1.002[1.002-1.002];<0.0001*** | |
|  |  | Aspartate transaminase, U/L | | 1.000[0.999-1.000];0.4275 | | | 1.002[1.001-1.002];<0.0001*** | | | 1.000[0.999-1.001];0.5219 | | | 1.000[0.998-1.001];0.6073 | | 1.000[0.999-1.001];0.7354 | |
|  |  | Alanine transaminase, U/L | | 0.999[0.999-1.000];0.0259* | | | 1.001[1.001-1.002];<0.0001*** | | | 0.999[0.999-1.000];0.0082** | | | 0.999[0.998-1.000];0.1492 | | 0.999[0.999-1.000];0.0026** | |
|  |  | Bilirubin, umol/L | | 1.001[0.999-1.003];0.4267 | | | 1.010[1.008-1.012];<0.0001*** | | | 1.001[0.999-1.004];0.2741 | | | 1.00[1.00-1.01];0.0433* | | 1.002[1.000-1.005];0.0629 | |
| Exclude mortality within 60 days | | Alkaline phosphatase, U/L | | 1.001[1.001-1.001];<0.0001*** | | | 1.002[1.001-1.002];<0.0001*** | | | 1.002[1.002-1.002];<0.0001*** | | | 1.001[1.001-1.002];0.0004*** | | 1.002[1.002-1.002];<0.0001*** | |
|  |  | Aspartate transaminase, U/L | | 1.000[0.999-1.000];0.4066 | | | 1.002[1.001-1.002];<0.0001*** | | | 1.000[0.999-1.001];0.5743 | | | 0.999[0.998-1.001];0.5294 | | 1.000[0.999-1.001];0.8278 | |
|  |  | Alanine transaminase, U/L | | 0.999[0.999-1.000];0.0106* | | | 1.001[1.001-1.002];<0.0001*** | | | 0.999[0.998-1.000];0.0023** | | | 0.999[0.998-1.000];0.1271 | | 0.999[0.998-1.000];0.0006*** | |
|  |  | Bilirubin, umol/L | | 1.001[0.999-1.003];0.4016 | | | 1.009[1.008-1.011];<0.0001*** | | | 1.002[0.999-1.004];0.2036 | | | 1.00[1.00-1.01];0.0949 | | 1.002[1.000-1.004];0.0625 | |
| Exclude mortality within 30 days | | Alkaline phosphatase, U/L | | 1.001[1.001-1.001];<0.0001*** | | | 1.002[1.001-1.002];<0.0001*** | | | 1.002[1.001-1.002];<0.0001*** | | | 1.001[1.000-1.002];0.0016** | | 1.002[1.001-1.002];<0.0001*** | |
|  |  | Aspartate transaminase, U/L | | 1.000[0.999-1.000];0.3500 | | | 1.002[1.001-1.002];<0.0001*** | | | 1.000[0.999-1.001];0.8827 | | | 0.999[0.998-1.001];0.4614 | | 1.000[0.999-1.001];0.8419 | |
|  |  | Alanine transaminase, U/L | | 0.999[0.999-1.000];0.0029** | | | 1.001[1.001-1.002];<0.0001*** | | | 0.999[0.998-0.999];0.0002*** | | | 0.999[0.997-1.000];0.0595 | | 0.999[0.998-0.999];<0.0001*** | |
|  |  | Bilirubin, umol/L | | 1.001[0.999-1.003];0.3659 | | | 1.009[1.007-1.010];<0.0001*** | | | 1.001[0.998-1.003];0.4949 | | | 1.00[1.00-1.01];0.1430 | | 1.001[0.999-1.004];0.2162 | |
| ***E. Landmark sensitivity analysis for lipid and glucose impact on subsequent outcomes after HCC diagnosis*** | | | | | | | | | | | | | | | | |
| Patients | Lipid and glucose tests | | | | HCC recurrence  HR [95% CI];P value | | | Liver cancer-related mortality HR [95% CI]; P value | | | Cancer-related mortality HR [95% CI];P value | | | Non-cancer-related mortality HR [95% CI]; P value | | All-cause mortality HR [95% CI];P value |
| Exclude mortality within 2*365 days | Triglyceride, mmol/L | | | | 0.95[0.84-1.08];0.4656 | | | 0.82[0.64-1.06];0.1234 | | | 0.95[0.80-1.13];0.5856 | | | 1.09[0.89-1.34];0.3896 | | 1.00[0.88-1.15];0.9494 |
|  | Low-density lipoprotein, mmol/L | | | | 1.03[0.93-1.13];0.6153 | | | 0.80[0.67-0.96];0.0153* | | | 0.87[0.76-1.00];0.0467* | | | 1.03[0.86-1.23];0.7494 | | 0.93[0.83-1.03];0.1618 |
|  | High-density lipoprotein, mmol/L | | | | 1.25[0.99-1.58];0.0612 | | | 1.12[0.76-1.67];0.5568 | | | 1.02[0.74-1.41];0.8929 | | | 0.97[0.64-1.49];0.9052 | | 1.00[0.78-1.30];0.9713 |
|  | Total cholesterol, mmol/L | | | | 1.05[0.97-1.14];0.2224 | | | 0.86[0.74-0.99];0.0414* | | | 0.91[0.81-1.03];0.1263 | | | 1.06[0.92-1.24];0.4154 | | 0.97[0.88-1.06];0.4593 |
|  | HbA1C, % | | | | 0.94[0.80-1.09];0.4055 | | | 1.02[0.82-1.27];0.8601 | | | 1.07[0.89-1.29];0.4442 | | | 1.14[0.91-1.43];0.2650 | | 1.10[0.95-1.27];0.1963 |
|  | Fasting glucose, mmol/L | | | | 1.02[0.99-1.06];0.1571 | | | 1.08[1.03-1.14];0.0016** | | | 1.07[1.02-1.11];0.0024** | | | 1.03[0.97-1.10];0.3196 | | 1.06[1.02-1.09];0.0022** |
| Exclude mortality within 365 days | Triglyceride, mmol/L | | | | 0.98[0.89-1.08];0.6251 | | | 0.78[0.62-0.96];0.0218* | | | 1.06[0.95-1.19];0.3045 | | | 1.03[0.86-1.24];0.7270 | | 1.05[0.96-1.16];0.2934 |
|  | Low-density lipoprotein, mmol/L | | | | 1.02[0.94-1.10];0.6900 | | | 0.79[0.68-0.92];0.0021** | | | 0.91[0.82-1.01];0.0907 | | | 0.97[0.84-1.13];0.7255 | | 0.93[0.86-1.02];0.1096 |
|  | High-density lipoprotein, mmol/L | | | | 1.16[0.96-1.40];0.1140 | | | 1.13[0.82-1.55];0.4660 | | | 0.94[0.74-1.20];0.6217 | | | 1.00[0.70-1.43];0.9910 | | 0.96[0.79-1.17];0.6862 |
|  | Total cholesterol, mmol/L | | | | 1.03[0.96-1.10];0.3815 | | | 0.84[0.74-0.95];0.0058** | | | 0.95[0.87-1.03];0.2240 | | | 1.00[0.88-1.14];0.9797 | | 0.96[0.90-1.04];0.3056 |
|  | HbA1C, % | | | | 0.96[0.85-1.08];0.5006 | | | 1.04[0.88-1.23];0.6552 | | | 1.02[0.90-1.17];0.7355 | | | 1.16[0.96-1.41];0.1235 | | 1.06[0.95-1.19];0.2677 |
|  | Fasting glucose, mmol/L | | | | 1.00[0.98-1.03];0.7192 | | | 1.06[1.02-1.10];0.0032** | | | 1.03[1.00-1.06];0.0332* | | | 1.00[0.95-1.06];0.9987 | | 1.02[1.00-1.05];0.0671 |
| Exclude mortality within 180 days | Triglyceride, mmol/L | | | | 0.94[0.87-1.02];0.1581 | | | 0.71[0.58-0.88];0.0013** | | | 1.03[0.93-1.14];0.6139 | | | 1.03[0.87-1.22];0.7229 | | 1.03[0.94-1.12];0.5384 |
|  | Low-density lipoprotein, mmol/L | | | | 1.03[0.96-1.11];0.4095 | | | 0.85[0.75-0.97];0.0156* | | | 0.96[0.89-1.05];0.4114 | | | 0.95[0.82-1.09];0.4426 | | 0.96[0.89-1.03];0.2703 |
|  | High-density lipoprotein, mmol/L | | | | 1.01[0.86-1.19];0.8908 | | | 1.04[0.78-1.39];0.7926 | | | 0.90[0.73-1.10];0.2989 | | | 0.85[0.61-1.18];0.3354 | | 0.88[0.74-1.05];0.1644 |
|  | Total cholesterol, mmol/L | | | | 1.03[0.97-1.09];0.4147 | | | 0.86[0.77-0.96];0.0080** | | | 0.97[0.90-1.04];0.4086 | | | 0.95[0.84-1.07];0.3731 | | 0.96[0.91-1.03];0.2417 |
|  | HbA1C, % | | | | 0.93[0.83-1.03];0.1626 | | | 1.05[0.91-1.22];0.4896 | | | 1.00[0.89-1.12];0.9499 | | | 1.15[0.95-1.38];0.1410 | | 1.03[0.94-1.14];0.5004 |
|  | Fasting glucose, mmol/L | | | | 1.00[0.97-1.02];0.7907 | | | 1.05[1.02-1.08];0.0043** | | | 1.02[0.99-1.04];0.2035 | | | 1.01[0.97-1.06];0.6041 | | 1.02[0.99-1.04];0.1724 |
| Exclude mortality within 120 days | Triglyceride, mmol/L | | | | 0.93[0.86-1.01];0.0878 | | | 0.70[0.57-0.85];0.0004*** | | | 1.00[0.91-1.11];0.9355 | | | 0.99[0.84-1.16];0.8640 | | 1.00[0.92-1.09];0.9849 |
|  | Low-density lipoprotein, mmol/L | | | | 1.06[0.99-1.13];0.1028 | | | 0.88[0.78-0.99];0.0366* | | | 0.98[0.91-1.06];0.7003 | | | 0.98[0.86-1.11];0.7063 | | 0.98[0.92-1.05];0.5999 |
|  | High-density lipoprotein, mmol/L | | | | 0.96[0.82-1.12];0.6197 | | | 0.98[0.74-1.29];0.8695 | | | 0.86[0.71-1.04];0.1258 | | | 0.80[0.58-1.09];0.1535 | | 0.84[0.72-0.99];0.0402* |
|  | Total cholesterol, mmol/L | | | | 1.04[0.98-1.10];0.2205 | | | 0.88[0.79-0.97];0.0117* | | | 0.98[0.91-1.05];0.5146 | | | 0.95[0.85-1.07];0.3999 | | 0.97[0.92-1.03];0.3201 |
|  | HbA1C, % | | | | 0.95[0.86-1.05];0.3589 | | | 1.10[0.96-1.27];0.1600 | | | 1.05[0.94-1.17];0.3995 | | | 1.07[0.90-1.28];0.4409 | | 1.05[0.96-1.16];0.2616 |
|  | Fasting glucose, mmol/L | | | | 1.00[0.98-1.02];0.9236 | | | 1.05[1.02-1.08];0.0033** | | | 1.02[0.99-1.04];0.1372 | | | 1.01[0.97-1.06];0.6148 | | 1.02[1.00-1.04];0.1218 |
| Exclude mortality within 90 days | Triglyceride, mmol/L | | | | 0.95[0.88-1.02];0.1764 | | | 0.72[0.60-0.87];0.0007*** | | | 1.01[0.93-1.11];0.7539 | | | 1.00[0.86-1.17];0.9671 | | 1.01[0.94-1.09];0.7706 |
|  | Low-density lipoprotein, mmol/L | | | | 1.07[1.01-1.14];0.0287* | | | 0.91[0.81-1.02];0.0894 | | | 1.01[0.94-1.09];0.7244 | | | 0.99[0.87-1.12];0.8182 | | 1.01[0.94-1.07];0.8518 |
|  | High-density lipoprotein, mmol/L | | | | 0.93[0.80-1.09];0.3790 | | | 0.98[0.75-1.28];0.8812 | | | 0.85[0.70-1.01];0.0718 | | | 0.77[0.56-1.04];0.0912 | | 0.82[0.70-0.96];0.0158* |
|  | Total cholesterol, mmol/L | | | | 1.04[0.99-1.10];0.1105 | | | 0.90[0.81-0.99];0.0286* | | | 1.00[0.93-1.06];0.8771 | | | 0.96[0.86-1.07];0.4655 | | 0.99[0.93-1.04];0.6145 |
|  | HbA1C, % | | | | 0.94[0.86-1.04];0.2329 | | | 1.07[0.93-1.22];0.3319 | | | 1.03[0.92-1.14];0.6220 | | | 1.06[0.89-1.26];0.5403 | | 1.03[0.95-1.13];0.4600 |
|  | Fasting glucose, mmol/L | | | | 1.00[0.98-1.02];0.9658 | | | 1.04[1.01-1.07];0.0105* | | | 1.01[0.99-1.03];0.3743 | | | 1.01[0.97-1.05];0.7047 | | 1.01[0.99-1.03];0.3358 |
| Exclude mortality within 60 days | Triglyceride, mmol/L | | | | 0.98[0.91-1.05];0.5111 | | | 0.73[0.61-0.87];0.0005*** | | | 1.02[0.94-1.11];0.6257 | | | 1.02[0.87-1.18];0.8410 | | 1.02[0.95-1.10];0.6009 |
|  | Low-density lipoprotein, mmol/L | | | | 1.09[1.03-1.15];0.0042** | | | 0.89[0.80-0.99];0.0389* | | | 1.02[0.96-1.09];0.5447 | | | 0.99[0.88-1.11];0.8262 | | 1.01[0.96-1.07];0.6761 |
|  | High-density lipoprotein, mmol/L | | | | 0.91[0.79-1.05];0.1858 | | | 0.94[0.73-1.22];0.6561 | | | 0.84[0.71-1.00];0.0499* | | | 0.71[0.52-0.96];0.0241* | | 0.81[0.70-0.94];0.0047** |
|  | Total cholesterol, mmol/L | | | | 1.06[1.01-1.11];0.0240* | | | 0.89[0.81-0.97];0.0118* | | | 1.00[0.95-1.06];0.8757 | | | 0.95[0.86-1.06];0.3791 | | 0.99[0.94-1.04];0.7646 |
|  | HbA1C, % | | | | 0.99[0.91-1.08];0.8634 | | | 1.14[1.01-1.29];0.0320* | | | 1.08[0.99-1.19];0.0878 | | | 1.05[0.88-1.25];0.5988 | | 1.08[0.99-1.17];0.0798 |
|  | Fasting glucose, mmol/L | | | | 1.00[0.98-1.02];0.9319 | | | 1.04[1.00-1.07];0.0248* | | | 1.01[0.99-1.03];0.4975 | | | 1.01[0.96-1.05];0.8151 | | 1.01[0.99-1.03];0.4784 |
| Exclude mortality within 30 days | Triglyceride, mmol/L | | | | 0.97[0.90-1.04];0.3299 | | | 0.75[0.64-0.89];0.0008*** | | | 1.01[0.93-1.09];0.8732 | | | 1.01[0.88-1.17];0.8761 | | 1.01[0.94-1.08];0.8298 |
|  | Low-density lipoprotein, mmol/L | | | | 1.07[1.02-1.13];0.0081** | | | 0.93[0.84-1.03];0.1568 | | | 1.02[0.96-1.09];0.4410 | | | 0.96[0.86-1.08];0.5107 | | 1.01[0.96-1.07];0.7167 |
|  | High-density lipoprotein, mmol/L | | | | 0.91[0.79-1.03];0.1444 | | | 0.85[0.66-1.09];0.1897 | | | 0.79[0.67-0.93];0.0035** | | | 0.71[0.53-0.95];0.0212* | | 0.77[0.67-0.89];0.0002*** |
|  | Total cholesterol, mmol/L | | | | 1.05[1.00-1.10];0.0394* | | | 0.91[0.84-1.00];0.0385* | | | 1.00[0.95-1.05];0.9024 | | | 0.95[0.86-1.05];0.3213 | | 0.99[0.94-1.03];0.5603 |
|  | HbA1C, % | | | | 1.00[0.92-1.08];0.9688 | | | 1.14[1.01-1.28];0.0324* | | | 1.07[0.98-1.17];0.1170 | | | 1.04[0.88-1.24];0.6225 | | 1.07[0.99-1.15];0.1049 |
|  | Fasting glucose, mmol/L | | | | 0.99[0.97-1.01];0.3347 | | | 1.03[1.00-1.06];0.0550 | | | 1.00[0.98-1.02];0.9191 | | | 1.00[0.96-1.04];0.8895 | | 1.00[0.98-1.02];0.9793 |
